# Subjective and Objective Cognitive Functioning in Chronic Pain: Distinct Associations with Multidimensional Symptom Burden and Resting-State EEG

**DOI:** 10.64898/2026.03.19.26348793

**Authors:** Paul Theo Zebhauser, Felix S. Bott, Enayatullah Baki, Elisabeth S. May, Markus Ploner

**Affiliations:** Center for Interdisciplinary Pain Medicine, TUM School of Medicine and Health, Technical University of Munich, Munich, Germany; Department of Neurology, TUM School of Medicine and Health, Technical University of Munich, Munich, Germany; TUM-Neuroimaging Center, TUM School of Medicine and Health, Technical University of Munich, Munich, Germany

## Abstract

Cognitive dysfunction is increasingly recognized as an important feature of chronic pain (CP). However, subjective cognitive complaints and objectively measured cognitive performance frequently diverge. Whether and how these two aspects of cognitive functioning differentially relate to the broad symptomatology and brain function in CP remains unclear. Here, 114 individuals with CP completed patient-reported outcome measures on cognitive functioning and multidimensional CP symptoms, as well as a visuospatial working memory task, and resting-state EEG. Bayesian correlations, network analyses, and Bayesian regression models examined how subjective and objective cognitive functioning relate to multidimensional CP symptoms and EEG activity/connectivity, while controlling for age and sex. Additional models tested whether EEG associations were independent of broader symptom burden. Results indicated that subjective and objective cognitive functioning were uncorrelated. Subjective cognitive functioning was strongly associated with psychosocial symptoms, whereas objective cognitive functioning was largely independent of broader symptom burden. EEG revealed associations between subjective cognitive functioning and bilateral frontotemporal beta connectivity; however, these relationships were substantially attenuated after accounting for broader CP symptom burden. Objective cognitive functioning showed no robust associations with EEG. These findings indicate a dissociation between subjective cognitive complaints and objective cognitive performance in CP. Subjective cognitive complaints were primarily associated with psychosocial symptom burden and beta-band hypoconnectivity. In contrast, objective cognitive performance was unrelated to the broader symptomatology of CP and EEG measures. This dissociation may inform more targeted interventions, optimize the allocation of cognitive assessment resources, and ultimately improve long-term functional outcomes in CP.

## 1. Introduction

Chronic pain (CP) is a multidimensional condition that includes a variety of symptoms beyond the experience of persistent pain. Along with affective and physical symptoms [8], cognitive dysfunction is increasingly recognized as an important feature of CP. People with CP frequently report subjective cognitive complaints, such as difficulties with concentration and memory [39; 42]. Objective cognitive testing has demonstrated reduced performance, particularly in working memory and other domains of executive functioning [5; 16; 23]. Importantly, cognitive dysfunction may not merely accompany CP but may also influence its clinical course. Cognitive deficits have been linked to the development of CP [2], and cognitive training has been associated with improved treatment outcomes in people with CP [18]. Furthermore, CP may contribute to cognitive decline [19; 25]. Together, these findings point to a close interplay between CP and cognitive dysfunction [47].

However, subjective cognitive complaints (assessed using patient-reported outcome measures) and objectively measured cognitive performance can diverge in CP [3; 39; 44], mirroring findings in depression [35] and other neuropsychiatric conditions [41]. This dissociation suggests that subjective and objective assessments capture distinct aspects of cognitive dysfunction. Prior work suggests that subjective cognitive complaints in CP relate to depressive symptoms and affective distress [27; 31; 43] and may contribute to disability more than objective deficits [39]. However, it remains unclear whether and how subjective and objective cognitive measures relate differentially to the broad spectrum of CP symptoms. Previous data-driven approaches have shown that the spectrum of CP symptoms can be meaningfully described along two dimensions: affective and physical burden [46]. These dimensions provide a useful framework for examining how subjective and objective cognitive functioning relate to the symptom profile of CP.

Divergence between subjective and objective cognitive functioning may reflect distinct individual profiles with different needs. Improved understanding of these profiles could inform targeted interventions, optimize resource allocation, and enhance the prediction of long-term functional outcomes and disease trajectories in CP. For example, individuals with prominent subjective complaints may benefit more from interventions targeting affective burden, whereas those with objective cognitive deficits may require neuropsychological rehabilitation. More broadly, such differentiation could inform patient stratification, prioritize in-depth cognitive assessment, and guide personalized treatment strategies.

While CP is increasingly conceptualized as a disorder of brain function [24], the neural correlates of cognitive functioning in CP, including the divergence between subjective and objective functioning, are largely unexplored. Functional imaging studies have reported altered brain activity patterns associated with working memory and executive deficits in fibromyalgia [14; 26]. However, evidence in broader CP populations and on the differential neural correlates of subjective versus objective cognitive functioning is lacking. Mapping these relationships could deepen the mechanistic understanding of cognitive dysfunction in CP and, ultimately, improve clinical care.

Here, we aimed to (1) characterize the relationship between subjective and objective cognitive functioning in CP and their associations with a broad spectrum of psychological, physical, and functional symptoms and (2) examine their relationships with resting-state EEG measures of neural activity and connectivity (Fig. 1).

**Figure 1:**
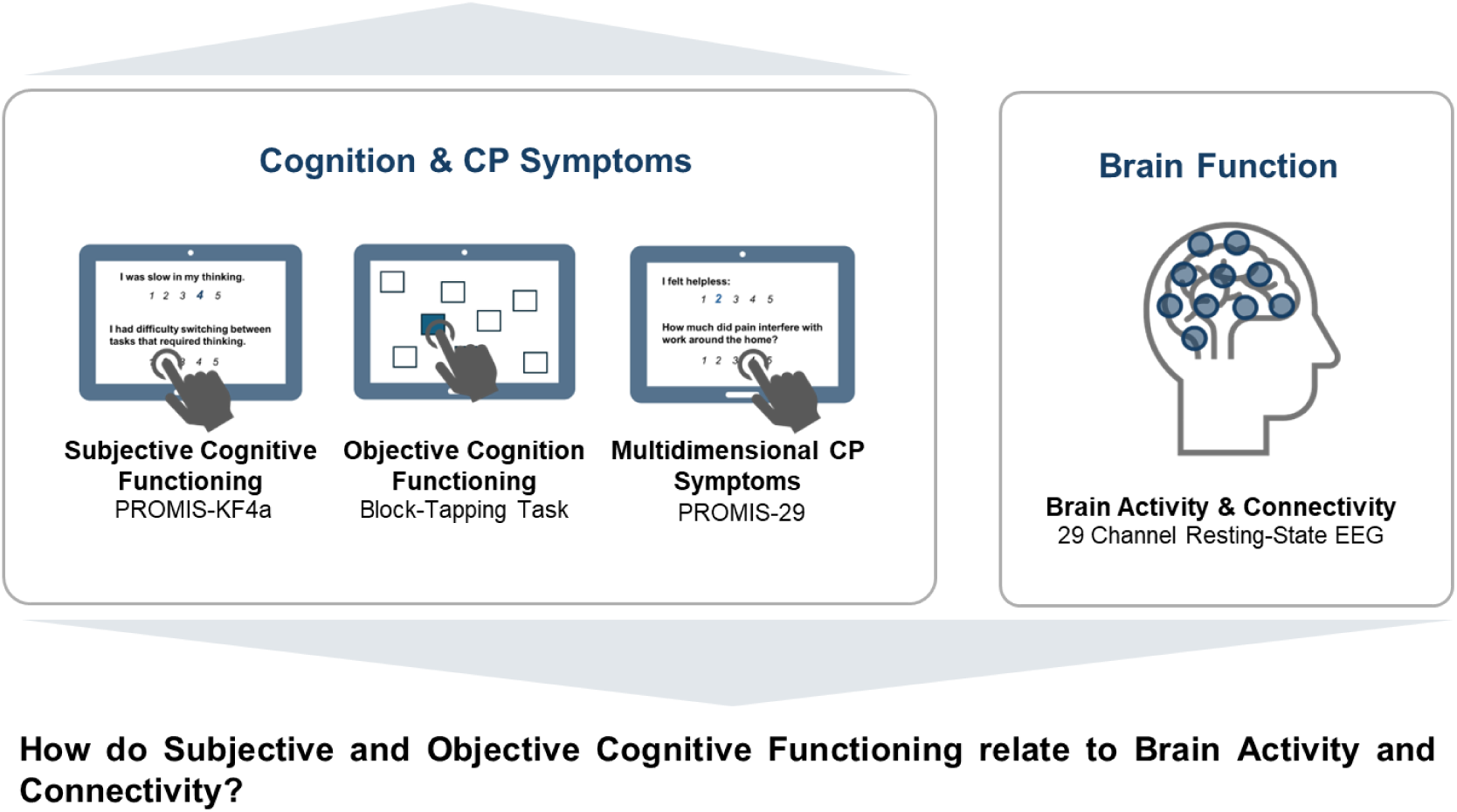
Overview of study design and main research questions. CP = Chronic Pain.

## 2. Methods

### 2.1. Preregistration & Ethics

The study protocol and analysis plan were preregistered (https://osf.io/3vpbz/overview). Data and analysis code will be made publicly available upon publication of this manuscript (https://osf.io/3vpbz/files/osfstorage). The study protocol was approved by the Ethics Committee of the School of Medicine and Health of the Technical University of Munich. The study was conducted in accordance with the Declaration of Helsinki.

### 2.2. Participants

Adults (≥18 years) with chronic pain of mixed etiologies (≥3 months duration) were recruited via convenience sampling from the outpatient clinic of the Center for Interdisciplinary Pain Medicine at TUM University Hospital Rechts der Isar, Munich, Germany. Exclusion criteria were severe concomitant neurological (e.g., multiple sclerosis) or psychiatric (e.g., post-traumatic stress disorder, schizophrenia) disorders, a primary headache disorder, or reported regular or recent intake of benzodiazepines. Other centrally acting medications were not considered exclusion criteria, as a prior study provided evidence against effects on EEG metrics [45].

### 2.3. Patient-Reported Outcomes and Working Memory Task

To assess subjective cognitive functioning, we administered the PROMIS Cognitive Function 4a, an internationally available, validated patient-reported outcome measure [20]. This questionnaire comprises 4 questions, each scored 1-5 on a Likert scale, yielding a summary score of 4-20; higher scores indicate fewer symptoms.

For objective cognitive functioning, we used a customized tablet-based visual working memory task. Working memory, often impaired in CP [5], is a core executive control mechanism underlying higher-order cognition such as learning, decision-making, and problem-solving [1]. Given its close relationship with multiple cognitive domains, working memory capacity has been proposed as a key indicator of global cognition [9]. In this task, participants observed a sequence of spatial locations and were required to reproduce it in reverse order (two sequences at each stimulus length of 2–7 items; see Fig. S1 for a screenshot of stimulus presentation). The task engages visuospatial working memory by requiring the temporary maintenance and active manipulation of spatial sequences. Performance yields a total score ranging from 0 to 12, with one point awarded for each correctly reproduced sequence.

To assess the broader symptomatology of CP, we administered the PROMIS-29v2.1 [15], which captures depression, anxiety, fatigue, pain interference, pain intensity, physical functioning, sleep disturbance, and social participation across 29 items rated on 5-point Likert scales. To reduce questionnaire data dimensionality and assess overall symptom burden for subsequent analyses, we performed principal component analysis (PCA) on the PROMIS-29 scales. Scores for physical functioning and social participation were inverted prior to analysis so that higher values consistently reflected greater symptom burden. We identified two latent symptom dimensions, affective and physical burden, mirroring previous findings in a larger CP sample [46] (see Table S2 for detailed PCA specifications and results, including component loadings and explained variance of components). Individual component scores for affective and physical burden (first two components) were extracted and used in subsequent analyses examining associations with cognitive measures, demographic variables, and EEG features.

All patient-reported outcome measures and the working memory task were fully computerized, including the instructions.

### 2.4. EEG Preprocessing

Participants were seated comfortably in a quiet, low-noise environment and instructed to remain relaxed but awake. Resting-state EEG was recorded for 5 minutes each under eyes-open and eyes-closed conditions. Only eyes-closed data were used for the current analyses. Signals were acquired using a 29-channel dry-electrode system (CGX Quick-32r, San Diego, CA) with wireless amplification, covering standard 10–20 positions plus Fpz, AF7/8, FC5/6, CP5/6, PO7/8, and Oz. Data were referenced and grounded at A1 (left earlobe), sampled at 500 Hz, and recorded with impedances below 2500 kΩ, consistent with higher impedance levels typically observed in dry EEG systems compared to wet systems [22]. EEG data were preprocessed and analyzed in MATLAB using the DISCOVER-EEG pipeline [13] with EEGLab [10] and FieldTrip [29] toolboxes. Preprocessing (default settings) included line noise removal, bad channel rejection, re-referencing, independent component analysis, bad component removal, bad channel interpolation, and bad segment removal.

### 2.5. EEG Analysis

We analyzed a comprehensive set of activity and connectivity features commonly used in resting-state EEG research, both globally and regionally. Distributional properties were assessed with the Kolmogorov–Smirnov test, and variables with p < 0.05 were evaluated for skewness. As all non-normally distributed variables (mainly power metrics) were right-skewed and non-negative, they were log-transformed. Global power and peak alpha frequency (PAF) were calculated in sensor space, while all connectivity and region-specific measures were derived in source space.

Source reconstruction was conducted with an array-gain linear constrained minimum variance (LCMV) beamformer [13]. Data were then projected to the source space defined by the 100-parcel Schaefer atlas (7-network version) [34]. Lead fields were computed using a volume conduction model based on the MNI template (standard_bem.mat) in FieldTrip. Spatial filters were derived from the covariance matrices of band-pass filtered data and the lead fields, with 5% regularization to address rank deficiencies. Functional connectivity was then estimated between all reconstructed virtual time series for each frequency band. Frequency bands were defined according to the *COBIDAS MEEG* recommendations [30]: theta (4 to < 8 Hz), alpha (8 to < 13 Hz), beta (13 to 30 Hz), and gamma (> 30 to 80 Hz).

Global peak alpha frequency (PAF) was calculated as the center of gravity of the alpha spectrum, weighting each frequency by its power and averaging across all electrodes in sensor space. Global power was computed by averaging absolute power values across predefined frequency bands and channels. Global connectivity for each frequency band was quantified as the amplitude envelope correlation (AEC) averaged across all pairwise source-level connections (upper triangle of the connectivity matrix), a standard approach in EEG research [6; 7; 17; 38]. Regional EEG features were calculated in the above-specified source space, using the 100-parcel Schaefer atlas [34]. Spatially resolved analogues of the global EEG features were used for region-specific analyses. Regional power was calculated from the source absolute power for each parcel, as provided by DISCOVER-EEG [13]. Regional peak alpha frequency (PAF) was calculated as the center of gravity of the alpha spectrum for each parcel in source space. Regional connectivity was quantified as the mean amplitude envelope correlation (AEC) between each parcel and all other parcels, commonly referred to as node strength [33].

### 2.6. Statistical Analysis

For a sample of *n* = 114, a power analysis for a multiple regression model with four predictors (subjective/objective cognitive functioning, age, and sex), assuming a significance level (α) of 0.05 and statistical power (1 – β) of 0.80, indicates that this sample size is sufficient to detect a small-to-medium effect size (Cohen’s *f²* = 0.11). Therefore, the study was adequately powered to conduct the planned main analysis.

Differences in objective and subjective cognitive functioning with sex were examined using Bayesian t-tests and the *BayesFactor* package in R *4.4.2 [37]* (default JZS prior: Cauchy 0, 0.707). Associations with age and other symptoms of CP (PROMIS-29 scores, and PCA-derived affective and physical burden) were computed using Bayesian correlations and the *BayesFactor* package in R (default Jeffrey’s adjusted beta prior); effect sizes were estimated using Pearson’s correlation coefficients [28]. Bayes Factors (BF_10_) for those and all other models were interpreted as very strong, strong, moderate, or weak evidence (>30, 10-30, 3–10, 1-3 regarding the alternative hypothesis; <1/30, 1/30-1/10, 1/10-1/3, 1/3–1 regarding the null hypothesis; [40]). To further assess associations between CP symptoms and cognitive functioning, we estimated a network of the respective variables, including PROMIS-29 profiles, subjective and objective cognitive functioning scores, using EBIC Graphical Lasso (EBIC-glasso) [21] with R packages *qgraph* and *bootnet*. This approach computes regularized partial correlations, capturing direct associations between variables. It imposes sparsity by shrinking small partial correlations toward zero, and EBIC selects the optimal regularization, yielding a robust network of the strongest direct relationships for visualization and descriptive interpretation [12]. To examine the relationship between cognitive functioning and EEG metrics, Bayesian multiple regression models were estimated using the *BayesFactor* package in R, with subjective and objective cognitive functioning as predictors and the respective EEG feature of interest as the dependent variable. Age and sex were included as covariates in the null model to account for potential confounding effects on EEG activity and their association with cognitive functioning; sex was dummy-coded. In additional models, we aimed to assess whether potential associations were independent of broader CP symptom burden. To that end, we added affective and physical burden component scores (see above for a description of the PCA) into the null model to control for their respective symptom severity. Predictor importance was assessed via Bayesian model averaging, yielding posterior coefficient estimates and Bayes factors for inclusion, reflecting the strength of evidence for each predictor. Default JZS priors were used for regression coefficients (r-scale = 0.354), with a binomial model prior (α = β = 1). As Bayesian inference provides probabilistic estimates for each effect, frequentist-style corrections for multiple tests were not applied [4; 36].

## 3. Results

### 3.1. Participant characteristics

Demographic and clinical characteristics of the participants are shown in Table 1. The most common CP diagnoses were back pain, other musculoskeletal pain, and neuropathic pain, reflecting a heterogeneous mixed-etiology sample. The mean pain intensity over the last 7 days was 5.4 (0-10 numeric rating scale), and the mean age was 56.9 years. Women comprised 57% of the sample.

**Table 1.** Demographic and Clinical Characteristics.

|  |  |
| --- | --- |
| n | 114 |
| Female (%) | 57% |
| Back Pain (%) | 39.5% |
| Chronic Pelvic Pain (%) | 7% |
| Chronic Widespread Pain (%) | 2.6% |
| Facial Pain (%) | 5.3% |
| (other) Musculoskeletal Pain (%) | 18.4% |
| Neuropathic Pain (%) | 27.2% |
|  | *Mean (SD)* |
| Subjective Cognitive Function (4-20)* | 15.1 (3.7) |
| Blocktapping backwards [Working Memory] (0-12) | 5.7 (3.0) |
| Physical Function (4-20) | 9.1 (4.4) |
| Anxiety (4-20) | 8.2 (3.1) |
| Depression (4-20) | 8.6 (3.3) |
| Fatigue (4-20) | 11.3 (4.1) |
| Sleep Interference (4-20) | 11.4 (4.0) |
| Social Participation (4-20) | 11.5 (4.5) |
| Pain Interference (4-20) | 12.7 (4.2) |
| Pain Intensity (0-10) | 5.4 (2.2) |
| Age | 56.9 (17.1) |

*\*higher values indicate better functioning*

### 3.2. Associations of Cognitive Functioning with Other Symptoms and Demographics

Bayesian correlation analyses (Fig. 2A) provided moderate evidence against an association between subjective and objective cognitive functioning (r = 0.02, BF_10_ = 0.22).

**Figure 2:**
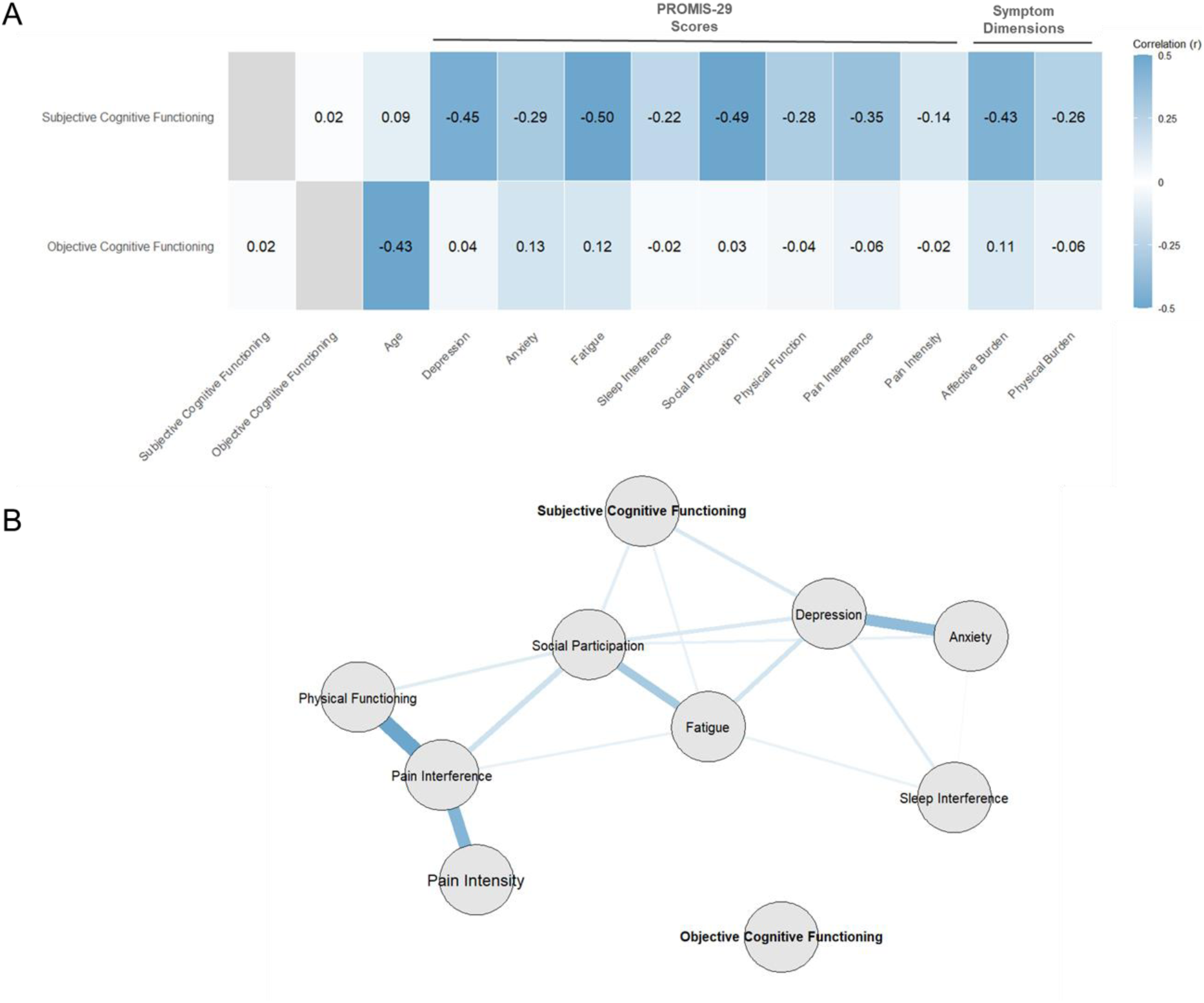
Clinical Associations of Subjective and Objective Cognitive Functioning. A) Correlations of subjective and objective cognitive functioning with other symptoms and PCA-derived symptom dimensions affective and physical burden. Darker shades of blue indicate larger correlation coefficients. Note that higher values of subjective cognitive functioning indicate better functioning. B) Network visualization (EBIC-glasso) displays regularized partial correlations between PROMIS-29 domains and subjective/objective cognitive functioning, reflecting direct associations while shrinking small edges toward zero. Edge weights represent the absolute value of partial correlations, with larger weights indicating stronger associations.

Subjective cognitive functioning showed very strong evidence for negative associations with depression (r = −0.45, BF_10_ > 10^3^), fatigue (r = −0.50, BF_10_ > 10^3^), social participation (r = −0.49, BF_10_ > 10^3^), and pain interference (r = −0.35, BF_10_ > 10^2^). This pattern was supported by strong evidence for an association with the PCA-derived affective burden dimension of CP symptoms (r = −0.43, BF_10_ > 10^3^). Associations with the remaining CP symptoms and PCA-derived physical burden were also observed, but they were weaker (all r’s < 0.3, all BF_10_’s between 3 and 30, see Table S3 for a full overview). As an exception, evidence against an association was found only for pain intensity (r = −0.14, BF_10_ = 0.61).

In contrast, objective cognitive functioning showed very strong evidence for a negative association with age (r = −0.43, BF_10_ > 10^3^), but evidence against associations with clinical variables (all r’s < 0.15, all BF_10_ < 1).

Network visualization (EBIC-glasso) corroborated these findings, indicating direct associations between subjective cognitive functioning and depression, fatigue, and social participation (see Fig. 2B), but no relations for objective cognitive functioning.

Bayesian t-Tests provided evidence against sex differences in both subjective and objective cognitive functioning (BF_10_ = 0.29 and 0.56, respectively).

Overall, these findings indicate that subjective cognitive complaints were closely embedded within the broader symptom profile of CP, with the strongest links to psychosocial symptom domains. In contrast, objective cognitive performance was largely independent of clinical symptom burden.

### 3.3. Associations of Cognitive Functioning with EEG

First, we analyzed associations between cognitive functioning and *global* EEG features. Bayesian regression analyses with EEG as the dependent variable and subjective and objective cognitive functioning as predictors (controlling for age and sex in the null model) provided overall evidence against associations between cognition and EEG features (global power, connectivity, PAF; see Table S4 for a summary of all models’ statistics).

We then analyzed associations between subjective and objective cognitive functioning and *regional* EEG features. Bayesian regression models revealed associations between regional beta connectivity of bilateral frontotemporal areas and subjective cognitive functioning (Fig. 3A). Strength of evidence varied across parcels, with weak evidence in 22 parcels, (r^2^ = 0.01 to 0.03, BF_10_ = 1.0 to 2.9), moderate evidence in 13 parcels (r^2^ = 0.03 to 0.04, BF_10_ = 3.1 to 6.8), strong evidence in 2 parcels (r^2^ = 0.05, BF_10_ = 10.0 and 11.3) and very strong evidence in 1 parcel (r^2^ = 0.09, BF_10_ = 81.7). No consistent associations were observed between subjective or objective cognitive functioning and other regional activity or connectivity measures (see S4 for a summary of all models’ statistics). In summary, regional analyses indicated an association between subjective cognitive complaints and frontotemporal beta hypoconnectivity.

**Figure 3:**
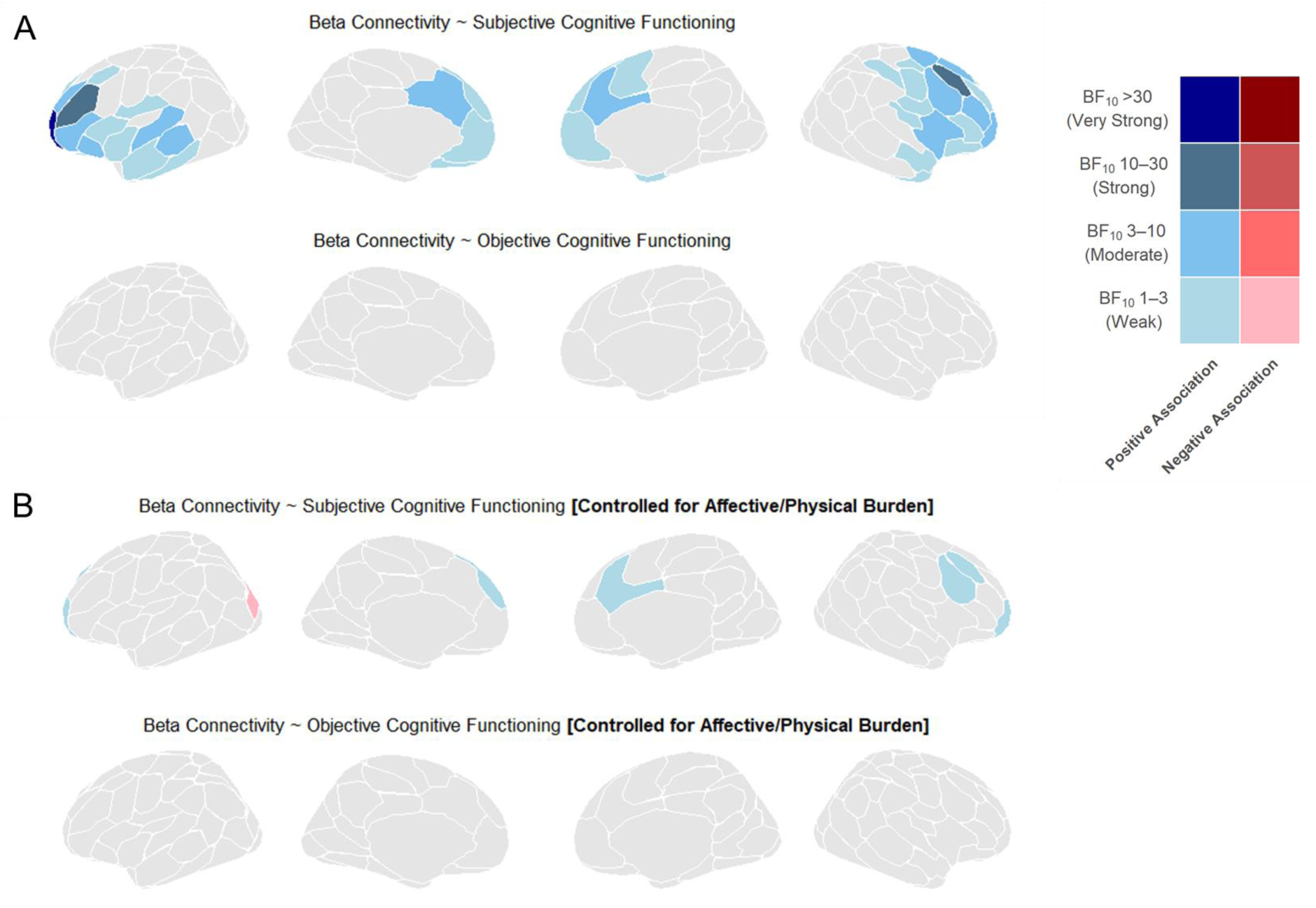
Regression analyses of Subjective and Objective Cognitive Functioning on regional beta connectivity. Parcellations of the Schaefer 100 parcel atlas are plotted with BFs and direction of effects (positive or negative association). Connectivity was calculated as the node strength of single parcels. Results for theta, alpha, and gamma connectivity/activity are not shown, as no consistent associations were found. A) Results of regression models including Age and Sex as covariates (null model). B) Results of regression models additionally including affective and physical burden as covariates (null model). BF = Bayes Factor.

In additional analyses, affective and physical burden were added to the null model to control for overall symptom burden when investigating associations between cognitive functioning and EEG. This was motivated by evidence for associations of subjective cognitive functioning with the broader symptomatology of CP (see Fig. 2A). After including these variables, the association between beta connectivity and subjective cognitive functioning was strongly attenuated (Fig. 3B.). Only weak evidence remained for positive associations in 6 parcels and for a negative association in 1 parcel (r^2^ = 0.01 to 0.02, BF_10_ = 1.0 to 2.1). Across all remaining EEG features, no consistent, interpretable associations with subjective or objective cognitive functioning emerged (see Table S5 for a summary of all models’ statistics).

Overall, we found an association between subjective cognitive complaints and beta hypoconnectivity, which was substantially attenuated after accounting for broader affective and physical symptom burden.

## 4. Discussion

### 4.1. Summary of Findings

In the present study, subjective cognitive complaints and objective working memory performance were dissociated at both behavioral and neural levels. Subjective cognitive complaints were embedded within the broader symptomatology of CP and inversely related to beta-band connectivity. In contrast, objective cognitive performance was largely independent of overall symptom burden and resting-state EEG measures. Importantly, associations between subjective cognitive functioning and beta connectivity were substantially attenuated after accounting for broader symptom burden, suggesting that these neural correlates may reflect shared variance with overall symptom load rather than isolated cognitive impairment.

### 4.2. Interpretation of Findings

Our findings extend previous reports of divergence between subjective complaints and objective performance in CP [3; 39; 44]. Prior research has linked patient-reported cognitive difficulties primarily to depressive and affective symptoms [27; 31; 43], with stronger links to disability than objective cognitive deficits [39]. The present results provide convergent behavioral and neural evidence for this distinction: subjective cognitive complaints appeared to reflect the multidimensional symptom architecture of CP, rather than serving as a direct proxy of neurocognitive abilities. From a clinical perspective, subjective complaints may reflect the experience of increased cognitive load in the context of persistent physical and affective distress.

At the neural level, associations between subjective cognitive complaints and beta hypoconnectivity are broadly consistent with prior work implicating beta oscillations in cognitive control and sustained attention [11]. However, the substantial attenuation of these effects after accounting for broader symptom burden suggests that altered beta connectivity may reflect a neural correlate of overall, predominantly affective distress [46], or a closely related increase in perceived cognitive load. In contrast, objective working memory performance showed no EEG associations. This absence of associations suggests that resting-state measures may be less sensitive to inter-individual variability in working memory performance in CP. Task-based EEG or fMRI paradigms, which more directly probe fronto-parietal control networks, may be better suited to detect such associations [32]. Taken together, these findings suggest that EEG resting-state measures in CP may primarily capture brain states associated with general symptom burden rather than specific cognitive processes.

### 4.3. Clinical Implications

Our findings have several implications for clinical assessment and research. The dissociation between subjective and objective cognitive functioning underscores the importance of assessing both domains independently. Patients may report significant cognitive difficulties despite normal objective performance, while others with measurable deficits may not perceive impairment.

Therefore, subjective cognitive complaints may reflect overall psychosocial symptom burden, whereas objective cognitive functioning may index neurocognitive capacity that is relatively independent of CP symptom architecture. This distinction may help refine patient stratification strategies and guide personalized treatment approaches. In clinical practice, patient-reported cognitive complaints may represent a particularly relevant target for routine assessment and, in some clinical scenarios, may more directly capture the lived experience of people with CP than extensive neuropsychological testing. Finally, the strong embedding of subjective cognitive functioning within psychosocial, and especially affective, symptoms suggests that interventions targeting mood and fatigue may alleviate cognitive complaints.

### 4.4. Strengths and limitations

This study has several strengths. Its multimodal design integrated patient-reported outcome measures, objective cognitive testing, and EEG measures within a single framework. The simultaneous examination of subjective and objective cognitive functioning within a broad symptom framework allowed for a comprehensive assessment that extends beyond single-domain approaches. Furthermore, the use of a Bayesian analytic framework enabled quantification of evidence both for and against specific associations, reducing reliance on dichotomous significance testing. In addition, the inclusion of standardized PROMIS-29 domains provided a comprehensive, transdiagnostic assessment of symptom burden, enhancing the clinical interpretability and comparability of results across conditions and populations.

Several limitations warrant consideration. Objective cognitive functioning was assessed using a single visuospatial working memory task. Although working memory represents a central cognitive domain in CP [5], different patterns may emerge across other executive, attentional, or memory functions. The sample size was modest, and replication in larger cohorts is needed. Moreover, the cross-sectional design precludes conclusions about causality or temporal relationships between cognitive functioning, symptom burden, and neural measures. Finally, the findings reflect the specific measures and symptom profiles of this mixed-etiology sample and should be interpreted within this context.

### 4.5. Conclusion and Outlook

Together, our findings provide convergent behavioral and neural evidence that subjective and objective cognitive functioning in CP represent dissociable constructs. Subjective cognitive complaints were closely linked to psychosocial symptom burden and associated with alterations in beta-band connectivity, whereas objective working memory performance was largely independent of both symptom burden and resting-state EEG measures.

These findings highlight the importance of distinguishing between subjective and objective aspects of cognition in both research and clinical practice. Future longitudinal studies may determine whether these dimensions differentially predict disease trajectories and treatment response in CP. Such insights could inform more targeted and individualized intervention strategies and ultimately improve functional outcomes.

## Author Contributions

*CRediT author statement*. P.T.Z.: Conceptualization, Methodology, Software, Formal analysis, Investigation, Data Curation, Writing - Original Draft, Writing - Review & Editing, Visualization, Funding acquisition. F.B.: Conceptualization, Methodology, Software, Data Curation, Writing - Review & Editing. E.B.: Investigation, Data Curation, Writing - Review & Editing. E.M.: Writing - Review & Editing. M.P.: Conceptualization, Methodology, Resources, Writing - Original Draft, Writing - Review & Editing, Supervision, Project administration, Funding acquisition.

## Data Availability

Data and analysis code will be made publicly available upon publication of this manuscript (OSF registries).

## Acknowledgements

The authors used Grammarly and ChatGPT to improve language and readability while preparing this manuscript. The authors edited the content as needed and take full responsibility for the content of the work.

