## Supplemental Material S1-S5 for "Subjective and Objective Cognitive Functioning in Chronic Pain: Distinct Associations with Multidimensional Symptom Burden and Resting-State EEG"

**Supplemental Digital Content**

**Figure S1**: Screenshot of stimulus presentation (working memory task)

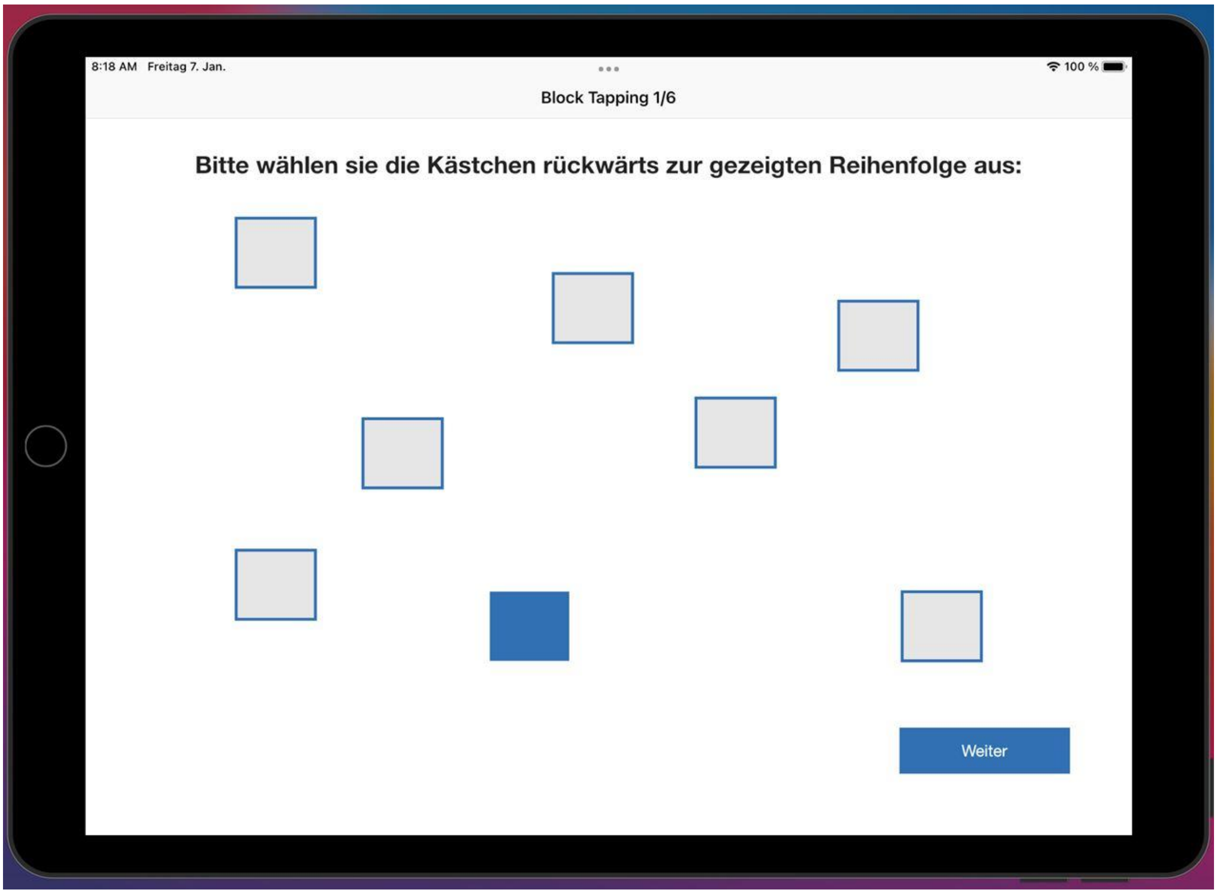

**Table S2:** PCA Results of Scores on PROMIS-29 Questionnaire Data: Loadings (RC1=Physical Burden, RC2=Affective Burden) and explained variance.

| | RC1 | RC2 |
|---|---|---|
| Pain_Intensity | 0.783 | 0.045 |
| Physical_Function | 0.820 | 0.187 |
| Anxiety | -0.010 | 0.870 |
| Depression | 0.291 | 0.840 |
| Fatigue | 0.480 | 0.679 |
| Sleep_Interference | 0.147 | 0.591 |
| Social_Participation | 0.610 | 0.586 |
| Pain_Interference | 0.876 | 0.323 |
| Proportion Var | 0.345 | 0.345 |
| Cumulative Var | 0.345 | 0.690 |

**Table S3** Correlations of Subjective and Objective Cognitive Functioning with PROMIS-Scores, PCA-derived Symptom Dimensions, and Age

| Cognition Measure | Outcome | BF10 | r |
|---|---|---|---|
| Subjective Cognition | Age | 0.3412652 | 0.09257907 |
| Subjective Cognition | Depression | 40,619.9250907 | -0.45183917 |
| Subjective Cognition | Anxiety | 26.9145063 | -0.29448684 |
| Subjective Cognition | Fatigue | 1,249,591.8898027 | -0.50381828 |
| Subjective Cognition | Sleep Interference | 3.0566100 | -0.22045071 |
| Subjective Cognition | Social Participation | 642,901.4119422 | -0.49441460 |
| Subjective Cognition | Physical Functioning | 15.6672223 | -0.27818039 |
| Subjective Cognition | Pain Interference | 202.8220962 | -0.34749543 |
| Subjective Cognition | Pain Intensity | 0.6094601 | -0.13899302 |
| Subjective Cognition | Affective Burden | 12,159.6041245 | -0.43115946 |

| Cognition Measure | Outcome | BF10 | r |
|---|---|---|---|
| Subjective Cognition | Physical Burden | 8.1172493 | -0.25672487 |
| Subjective Cognition | Objective Cognition | 0.2205310 | 0.02002715 |
| Objective Cognition | Age | 8,868.9678674 | -0.42550269 |
| Objective Cognition | Depression | 0.2341172 | 0.03903424 |
| Objective Cognition | Anxiety | 0.5405659 | 0.13078737 |
| Objective Cognition | Fatigue | 0.4489886 | 0.11691895 |
| Objective Cognition | Sleep Interference | 0.2223414 | -0.02355174 |
| Objective Cognition | Social Participation | 0.2232784 | 0.02517299 |
| Objective Cognition | Physical Functioning | 0.2312805 | -0.03598429 |
| Objective Cognition | Pain Interference | 0.2579707 | -0.05781754 |
| Objective Cognition | Pain Intensity | 0.2204720 | -0.01990135 |

| Cognition Measure | Outcome | BF10 | r |
|---|---|---|---|
| Objective Cognition | Affective Burden | 0.3969528 | 0.10669448 |
| Objective Cognition | Physical Burden | 0.2675605 | -0.06345378 |
| Objective Cognition | Subjective Cognition | 0.2205310 | 0.02002715 |

**Table S4** Full Summary of Regression Models of EEG features (DV) and Subjective/Cognitive Functioning (Predictors). ROI numbering refers to the labeling of the 100 parcel 7 network version of the Schaefer-Yeo atlases (2018)

| DV | Predictor | Sum_PostModProb_Incl | Sum_PostModProb_Excl | Sum_PriorModProb_Incl | BF_Incl | Frequentist_p_value | postMean | postStd |
|---|---|---|---|---|---|---|---|---|
| AEC_average_alpha | Blocktapping | 0.2017476 | 0.79825237 | 0.5 | 0.2527366 | 0.7382010857 | 0.00577013583 | 0.02434126 |
| AEC_average_alpha | Sum_PC | 0.2239857 | 0.77601431 | 0.5 | 0.2886360 | 0.4352680899 | -0.01810748607 | 0.02514299 |
| AEC_average_beta | Blocktapping | 0.2780224 | 0.72197761 | 0.5 | 0.3850845 | 0.9503944380 | 0.00009321378 | 0.02261741 |
| AEC_average_beta | Sum_PC | 0.5588210 | 0.44117903 | 0.5 | 1.2666535 | 0.0440663429 | 0.10658542920 | 0.05820945 |
| AEC_average_gamma | Blocktapping | 0.2158385 | 0.78416148 | 0.5 | 0.2752475 | 0.5365340881 | -0.01644964058 | 0.02617064 |
| AEC_average_gamma | Sum_PC | 0.2081083 | 0.79189174 | 0.5 | 0.2627989 | 0.6067608562 | -0.01185823571 | 0.02229726 |
| AEC_average_theta | Blocktapping | 0.2511745 | 0.74882545 | 0.5 | 0.3354247 | 0.3643867715 | -0.02338457493 | 0.02960942 |
| AEC_average_theta | Sum_PC | 0.2475759 | 0.75242411 | 0.5 | 0.3290377 | 0.3584867668 | 0.02171386472 | 0.02613093 |
| alpha_ROI1_AECnodestrength | Blocktapping | 0.2536149 | 0.74638508 | 0.5 | 0.3397910 | 0.5477108075 | 0.01315677945 | 0.02702415 |
| alpha_ROI1_AECnodestrength | Sum_PC | 0.3534651 | 0.64653487 | 0.5 | 0.5467070 | 0.1534122503 | -0.04770215488 | 0.03851737 |
| alpha_ROI10_AECnodestrength | Blocktapping | 0.2478507 | 0.75214933 | 0.5 | 0.3295232 | 0.3639275247 | 0.02467298596 | 0.03051395 |
| alpha_ROI10_AECnodestrength | Sum_PC | 0.2042337 | 0.79576626 | 0.5 | 0.2566504 | 0.7117601882 | -0.00631623505 | 0.02074831 |
| alpha_ROI100_AECnodestrength | Blocktapping | 0.2259735 | 0.77402647 | 0.5 | 0.2919455 | 0.6523197961 | 0.00923769720 | 0.02480563 |
| alpha_ROI100_AECnodestrength | Sum_PC | 0.2827338 | 0.71726618 | 0.5 | 0.3941826 | 0.2747291595 | -0.03083630973 | 0.03024698 |
| alpha_ROI11_AECnodestrength | Blocktapping | 0.1926063 | 0.80739371 | 0.5 | 0.2385531 | 0.6963729711 | 0.00746332526 | 0.02436648 |
| alpha_ROI11_AECnodestrength | Sum_PC | 0.1941682 | 0.80583179 | 0.5 | 0.2409538 | 0.6624782364 | -0.00907446462 | 0.02226734 |
| alpha_ROI12_AECnodestrength | Blocktapping | 0.2948993 | 0.70510075 | 0.5 | 0.4182370 | 0.2283824754 | 0.04028165380 | 0.03589525 |
| alpha_ROI12_AECnodestrength | Sum_PC | 0.2213461 | 0.77865394 | 0.5 | 0.2842676 | 0.6791708830 | -0.00739521829 | 0.02149404 |
| alpha_ROI13_AECnodestrength | Blocktapping | 0.2843461 | 0.71565391 | 0.5 | 0.3973235 | 0.2569948926 | 0.03344740438 | 0.03308227 |
| alpha_ROI13_AECnodestrength | Sum_PC | 0.2523461 | 0.74765394 | 0.5 | 0.3375172 | 0.4086707767 | -0.01921382289 | 0.02622461 |
| alpha_ROI14_AECnodestrength | Blocktapping | 0.1534042 | 0.84659582 | 0.5 | 0.1812012 | 0.6658978548 | 0.00639583456 | 0.01649783 |
| alpha_ROI14_AECnodestrength | Sum_PC | 0.3941739 | 0.60582611 | 0.5 | 0.6506387 | 0.5703235056 | -0.02360834992 | 0.04449558 |
| alpha_ROI15_AECnodestrength | Blocktapping | 0.1897581 | 0.81024190 | 0.5 | 0.2341993 | 0.8982037699 | 0.00293660062 | 0.02295411 |
| alpha_ROI15_AECnodestrength | Sum_PC | 0.2139293 | 0.78607068 | 0.5 | 0.2721502 | 0.4877127082 | -0.01464990081 | 0.02425831 |
| alpha_ROI16_AECnodestrength | Blocktapping | 0.3716923 | 0.62830769 | 0.5 | 0.5915769 | 0.4109561153 | 0.02086148644 | 0.02839010 |

| DV | Predictor | Sum_PostModProb_Incl | Sum_PostModProb_Excl | Sum_PriorModProb_Incl | BF_Incl | Frequentist_p_value | postMean | postStd |
|---|---|---|---|---|---|---|---|---|
| alpha_ROI16_AECnodestrength | Sum_PC | 0.6877181 | 0.31228188 | 0.5 | 2.2022352 | 0.0233150930 | -0.13895757917 | 0.06550250 |
| alpha_ROI17_AECnodestrength | Blocktapping | 0.2119505 | 0.78804955 | 0.5 | 0.2689557 | 0.5603473507 | 0.01439938795 | 0.02644361 |
| alpha_ROI17_AECnodestrength | Sum_PC | 0.1994739 | 0.80052606 | 0.5 | 0.2491786 | 0.8148162930 | -0.00384470977 | 0.02141559 |
| alpha_ROI18_AECnodestrength | Blocktapping | 0.2905128 | 0.70948716 | 0.5 | 0.4094688 | 0.2441504135 | 0.03651869982 | 0.03377759 |
| alpha_ROI18_AECnodestrength | Sum_PC | 0.2412105 | 0.75878947 | 0.5 | 0.3178886 | 0.4839280850 | -0.01368029842 | 0.02430665 |
| alpha_ROI19_AECnodestrength | Blocktapping | 0.1819942 | 0.81800584 | 0.5 | 0.2224852 | 0.8381072670 | 0.00337095504 | 0.02268583 |
| alpha_ROI19_AECnodestrength | Sum_PC | 0.1926457 | 0.80735434 | 0.5 | 0.2386135 | 0.5928444851 | -0.01008200434 | 0.02215543 |
| alpha_ROI2_AECnodestrength | Blocktapping | 0.2473622 | 0.75263778 | 0.5 | 0.3286604 | 0.8601559260 | 0.00266094262 | 0.02305194 |
| alpha_ROI2_AECnodestrength | Sum_PC | 0.4273405 | 0.57265951 | 0.5 | 0.7462384 | 0.0911909980 | -0.07175420582 | 0.04545982 |
| alpha_ROI20_AECnodestrength | Blocktapping | 0.2872702 | 0.71272978 | 0.5 | 0.4030563 | 0.3037139178 | 0.02818482359 | 0.03245336 |
| alpha_ROI20_AECnodestrength | Sum_PC | 0.2989769 | 0.70102306 | 0.5 | 0.4264866 | 0.2529083498 | -0.03323463422 | 0.03011763 |
| alpha_ROI21_AECnodestrength | Blocktapping | 0.3666821 | 0.63331792 | 0.5 | 0.5789858 | 0.1545654678 | 0.05281906210 | 0.04057521 |
| alpha_ROI21_AECnodestrength | Sum_PC | 0.2954585 | 0.70454146 | 0.5 | 0.4193629 | 0.3275939925 | -0.02400951437 | 0.02669989 |
| alpha_ROI22_AECnodestrength | Blocktapping | 0.2433768 | 0.75662324 | 0.5 | 0.3216618 | 0.3669408377 | 0.02324605004 | 0.02990751 |
| alpha_ROI22_AECnodestrength | Sum_PC | 0.2173281 | 0.78267190 | 0.5 | 0.2776746 | 0.5850680141 | -0.01169299766 | 0.02341870 |
| alpha_ROI23_AECnodestrength | Blocktapping | 0.1833826 | 0.81661739 | 0.5 | 0.2245637 | 0.9488410982 | 0.00251664251 | 0.02226547 |
| alpha_ROI23_AECnodestrength | Sum_PC | 0.1934099 | 0.80659007 | 0.5 | 0.2397871 | 0.7038331417 | -0.00848624592 | 0.02151528 |
| alpha_ROI24_AECnodestrength | Blocktapping | 0.2265582 | 0.77344177 | 0.5 | 0.2929221 | 0.4189113746 | 0.02085374774 | 0.02788742 |
| alpha_ROI24_AECnodestrength | Sum_PC | 0.2048306 | 0.79516941 | 0.5 | 0.2575936 | 0.6272152935 | -0.00999933515 | 0.02202460 |
| alpha_ROI25_AECnodestrength | Blocktapping | 0.2102533 | 0.78974671 | 0.5 | 0.2662288 | 0.5247590158 | 0.01494919003 | 0.02596845 |
| alpha_ROI25_AECnodestrength | Sum_PC | 0.1943933 | 0.80560666 | 0.5 | 0.2413006 | 0.9138471450 | -0.00156973483 | 0.02170700 |
| alpha_ROI26_AECnodestrength | Blocktapping | 0.1923632 | 0.80763681 | 0.5 | 0.2381803 | 0.6850774091 | 0.00922620462 | 0.02380497 |
| alpha_ROI26_AECnodestrength | Sum_PC | 0.1878857 | 0.81211435 | 0.5 | 0.2313537 | 0.8670339212 | -0.00372093459 | 0.02119596 |
| alpha_ROI27_AECnodestrength | Blocktapping | 0.1868541 | 0.81314595 | 0.5 | 0.2297915 | 0.8988365129 | 0.00354034479 | 0.02304212 |
| alpha_ROI27_AECnodestrength | Sum_PC | 0.1975878 | 0.80241216 | 0.5 | 0.2462423 | 0.6052011273 | 0.01263630428 | 0.02201877 |
| alpha_ROI28_AECnodestrength | Blocktapping | 0.2146804 | 0.78531957 | 0.5 | 0.2733670 | 0.5098733877 | -0.01699416942 | 0.02668006 |
| alpha_ROI28_AECnodestrength | Sum_PC | 0.1939131 | 0.80608689 | 0.5 | 0.2405610 | 0.9074850217 | -0.00303224559 | 0.02116100 |
| alpha_ROI29_AECnodestrength | Blocktapping | 0.2278829 | 0.77211711 | 0.5 | 0.2951403 | 0.5408753862 | 0.01371660663 | 0.02627346 |
| alpha_ROI29_AECnodestrength | Sum_PC | 0.2678047 | 0.73219527 | 0.5 | 0.3657559 | 0.2988649031 | -0.02809334314 | 0.02946752 |

| DV | Predictor | Sum_PostModProb_Incl | Sum_PostModProb_Excl | Sum_PriorModProb_Incl | BF_Incl | Frequentist_p_value | postMean | postStd |
|---|---|---|---|---|---|---|---|---|
| alpha_ROI3_AECnodestrength | Blocktapping | 0.2004820 | 0.79951796 | 0.5 | 0.2507536 | 0.9068241793 | 0.00152054994 | 0.02248299 |
| alpha_ROI3_AECnodestrength | Sum_PC | 0.2583327 | 0.74166735 | 0.5 | 0.3483134 | 0.2956906204 | -0.02871849071 | 0.02903066 |
| alpha_ROI30_AECnodestrength | Blocktapping | 0.3234123 | 0.67658772 | 0.5 | 0.4780050 | 0.2542443681 | -0.03683257774 | 0.03519649 |
| alpha_ROI30_AECnodestrength | Sum_PC | 0.3283113 | 0.67168874 | 0.5 | 0.4887848 | 0.2511876391 | -0.03438800193 | 0.03159322 |
| alpha_ROI31_AECnodestrength | Blocktapping | 0.1912374 | 0.80876262 | 0.5 | 0.2364567 | 0.7090815828 | 0.00892938323 | 0.02448227 |
| alpha_ROI31_AECnodestrength | Sum_PC | 0.1831526 | 0.81684738 | 0.5 | 0.2242189 | 0.9904313718 | -0.00019387804 | 0.02085662 |
| alpha_ROI32_AECnodestrength | Blocktapping | 0.2091396 | 0.79086041 | 0.5 | 0.2644456 | 0.5498381303 | 0.01408166234 | 0.02674010 |
| alpha_ROI32_AECnodestrength | Sum_PC | 0.1925855 | 0.80741449 | 0.5 | 0.2385212 | 0.9914846502 | 0.00120162110 | 0.02061964 |
| alpha_ROI33_AECnodestrength | Blocktapping | 0.2653314 | 0.73466862 | 0.5 | 0.3611579 | 0.4594672690 | 0.01525877736 | 0.02654306 |
| alpha_ROI33_AECnodestrength | Sum_PC | 0.3696217 | 0.63037826 | 0.5 | 0.5863491 | 0.1457071412 | -0.05076405249 | 0.03969533 |
| alpha_ROI34_AECnodestrength | Blocktapping | 0.2527066 | 0.74729335 | 0.5 | 0.3381626 | 0.4651437736 | 0.01756527756 | 0.02676827 |
| alpha_ROI34_AECnodestrength | Sum_PC | 0.3181071 | 0.68189285 | 0.5 | 0.4665061 | 0.1926154533 | -0.03951595190 | 0.03332329 |
| alpha_ROI35_AECnodestrength | Blocktapping | 0.2206060 | 0.77939399 | 0.5 | 0.2830481 | 0.4652068057 | 0.01845030495 | 0.02733025 |
| alpha_ROI35_AECnodestrength | Sum_PC | 0.1946391 | 0.80536088 | 0.5 | 0.2416794 | 0.8085042276 | -0.00427134234 | 0.02034709 |
| alpha_ROI36_AECnodestrength | Blocktapping | 0.1958813 | 0.80411873 | 0.5 | 0.2435974 | 0.8633353322 | 0.00310125144 | 0.02344074 |
| alpha_ROI36_AECnodestrength | Sum_PC | 0.2149055 | 0.78509449 | 0.5 | 0.2737320 | 0.4945957729 | -0.01448206208 | 0.02412284 |
| alpha_ROI37_AECnodestrength | Blocktapping | 0.2513589 | 0.74864109 | 0.5 | 0.3357536 | 0.3865510217 | 0.02308170141 | 0.02808509 |
| alpha_ROI37_AECnodestrength | Sum_PC | 0.2624954 | 0.73750462 | 0.5 | 0.3559237 | 0.3329464999 | -0.02478695667 | 0.02792354 |
| alpha_ROI38_AECnodestrength | Blocktapping | 0.1977680 | 0.80223201 | 0.5 | 0.2465222 | 0.6411838115 | 0.01032738937 | 0.02557786 |
| alpha_ROI38_AECnodestrength | Sum_PC | 0.1853670 | 0.81463303 | 0.5 | 0.2275466 | 0.8122858519 | 0.00421711401 | 0.02083309 |
| alpha_ROI39_AECnodestrength | Blocktapping | 0.2897342 | 0.71026582 | 0.5 | 0.4079236 | 0.2812398023 | 0.03164112585 | 0.03142107 |
| alpha_ROI39_AECnodestrength | Sum_PC | 0.2902539 | 0.70974609 | 0.5 | 0.4089546 | 0.2795591178 | -0.02796574259 | 0.02952340 |
| alpha_ROI4_AECnodestrength | Blocktapping | 0.2098889 | 0.79011113 | 0.5 | 0.2656448 | 0.9043570169 | -0.00239133548 | 0.02433199 |
| alpha_ROI4_AECnodestrength | Sum_PC | 0.2537146 | 0.74628537 | 0.5 | 0.3399700 | 0.3115328974 | -0.02567963781 | 0.02772932 |
| alpha_ROI40_AECnodestrength | Blocktapping | 0.3693130 | 0.63068696 | 0.5 | 0.5855727 | 0.4475633288 | 0.01752075093 | 0.02743083 |
| alpha_ROI40_AECnodestrength | Sum_PC | 0.7105438 | 0.28945617 | 0.5 | 2.4547545 | 0.0199669090 | -0.14419466287 | 0.06516272 |
| alpha_ROI41_AECnodestrength | Blocktapping | 0.2712386 | 0.72876139 | 0.5 | 0.3721913 | 0.5671542497 | 0.01395174132 | 0.02712502 |
| alpha_ROI41_AECnodestrength | Sum_PC | 0.3965597 | 0.60344035 | 0.5 | 0.6571646 | 0.1227137487 | -0.05791208515 | 0.04176676 |
| alpha_ROI42_AECnodestrength | Blocktapping | 0.1991111 | 0.80088888 | 0.5 | 0.2486127 | 0.6327750699 | 0.01077836729 | 0.02506387 |

| DV | Predictor | Sum_PostModProb_Incl | Sum_PostModProb_Excl | Sum_PriorModProb_Incl | BF_Incl | Frequentist_p_value | postMean | postStd |
|---|---|---|---|---|---|---|---|---|
| alpha_ROI42_AEC nodestrength | Sum_PC | 0.1932796 | 0.80672036 | 0.5 | 0.2395869 | 0.6821940891 | -0.00755906447 | 0.02125009 |
| alpha_ROI43_AEC nodestrength | Blocktapping | 0.1947291 | 0.80527089 | 0.5 | 0.2418181 | 0.6368452076 | 0.01085283406 | 0.02360258 |
| alpha_ROI43_AEC nodestrength | Sum_PC | 0.1882684 | 0.81173156 | 0.5 | 0.2319344 | 0.7565556725 | 0.00663254658 | 0.02136340 |
| alpha_ROI44_AEC nodestrength | Blocktapping | 0.2144236 | 0.78557636 | 0.5 | 0.2729507 | 0.4964592903 | -0.01806671426 | 0.02615127 |
| alpha_ROI44_AEC nodestrength | Sum_PC | 0.1937012 | 0.80629882 | 0.5 | 0.2402350 | 0.8704339035 | -0.00367691179 | 0.02098985 |
| alpha_ROI45_AEC nodestrength | Blocktapping | 0.1826778 | 0.81732223 | 0.5 | 0.2235076 | 0.7218017440 | -0.00727513405 | 0.02226090 |
| alpha_ROI45_AEC nodestrength | Sum_PC | 0.1786395 | 0.82136052 | 0.5 | 0.2174922 | 0.9459820199 | -0.00097935313 | 0.02009366 |
| alpha_ROI46_AEC nodestrength | Blocktapping | 0.2163027 | 0.78369726 | 0.5 | 0.2760029 | 0.5222211968 | -0.01372130811 | 0.02623647 |
| alpha_ROI46_AEC nodestrength | Sum_PC | 0.2121412 | 0.78785877 | 0.5 | 0.2692630 | 0.6114271166 | 0.01073950063 | 0.02371383 |
| alpha_ROI47_AEC nodestrength | Blocktapping | 0.1873202 | 0.81267980 | 0.5 | 0.2304969 | 0.8949182820 | 0.00114873348 | 0.02319266 |
| alpha_ROI47_AEC nodestrength | Sum_PC | 0.1904476 | 0.80955236 | 0.5 | 0.2352506 | 0.7703653115 | -0.00503749072 | 0.02169707 |
| alpha_ROI48_AEC nodestrength | Blocktapping | 0.1894749 | 0.81052511 | 0.5 | 0.2337681 | 0.9073695410 | 0.00287276345 | 0.02320332 |
| alpha_ROI48_AEC nodestrength | Sum_PC | 0.2061192 | 0.79388085 | 0.5 | 0.2596349 | 0.5659324650 | -0.01171884240 | 0.02295021 |
| alpha_ROI49_AEC nodestrength | Blocktapping | 0.2097225 | 0.79027746 | 0.5 | 0.2653784 | 0.7562883651 | 0.00551432069 | 0.02320837 |
| alpha_ROI49_AEC nodestrength | Sum_PC | 0.2695142 | 0.73048585 | 0.5 | 0.3689519 | 0.2665085989 | -0.03121574159 | 0.03012023 |
| alpha_ROI5_AECn odestrength | Blocktapping | 0.2038925 | 0.79610754 | 0.5 | 0.2561117 | 0.8985916441 | -0.00276210259 | 0.02243937 |
| alpha_ROI5_AECn odestrength | Sum_PC | 0.2591966 | 0.74080340 | 0.5 | 0.3498858 | 0.3046315619 | -0.02847501149 | 0.02949713 |
| alpha_ROI50_AEC nodestrength | Blocktapping | 0.2635665 | 0.73643352 | 0.5 | 0.3578958 | 0.4859444314 | 0.01716443692 | 0.02656404 |
| alpha_ROI50_AEC nodestrength | Sum_PC | 0.3540969 | 0.64590314 | 0.5 | 0.5482198 | 0.1468905579 | -0.04967065016 | 0.03823650 |
| alpha_ROI51_AEC nodestrength | Blocktapping | 0.2450632 | 0.75493685 | 0.5 | 0.3246141 | 0.4011266191 | 0.02099964050 | 0.02827923 |
| alpha_ROI51_AEC nodestrength | Sum_PC | 0.2462994 | 0.75370059 | 0.5 | 0.3267868 | 0.3839435344 | -0.01972444469 | 0.02671501 |
| alpha_ROI52_AEC nodestrength | Blocktapping | 0.2246792 | 0.77532080 | 0.5 | 0.2897887 | 0.5070763074 | 0.01609412409 | 0.02646925 |
| alpha_ROI52_AEC nodestrength | Sum_PC | 0.2237948 | 0.77620524 | 0.5 | 0.2883191 | 0.4952509360 | -0.01435398368 | 0.02467750 |
| alpha_ROI53_AEC nodestrength | Blocktapping | 0.1942882 | 0.80571178 | 0.5 | 0.2411386 | 0.7951525679 | 0.00547852963 | 0.02293976 |
| alpha_ROI53_AEC nodestrength | Sum_PC | 0.2021437 | 0.79785629 | 0.5 | 0.2533585 | 0.5644445998 | -0.01255248328 | 0.02280782 |
| alpha_ROI54_AEC nodestrength | Blocktapping | 0.2076897 | 0.79231031 | 0.5 | 0.2621317 | 0.6596610798 | 0.00931247749 | 0.02451362 |
| alpha_ROI54_AEC nodestrength | Sum_PC | 0.2205098 | 0.77949025 | 0.5 | 0.2828897 | 0.4656190197 | -0.01570512906 | 0.02458530 |
| alpha_ROI55_AEC nodestrength | Blocktapping | 0.1975008 | 0.80249917 | 0.5 | 0.2461072 | 0.8822341172 | -0.00252024364 | 0.02425500 |
| alpha_ROI55_AEC nodestrength | Sum_PC | 0.2212371 | 0.77876285 | 0.5 | 0.2840880 | 0.4725866103 | -0.01639719725 | 0.02526560 |

| DV | Predictor | Sum_PostModProb_Incl | Sum_PostModProb_Excl | Sum_PriorModProb_Incl | BF_Incl | Frequentist_p_value | postMean | postStd |
|---|---|---|---|---|---|---|---|---|
| alpha_ROI56_AECnodestrength | Blocktapping | 0.2111640 | 0.78883600 | 0.5 | 0.2676906 | 0.5320816075 | 0.01370030436 | 0.02615873 |
| alpha_ROI56_AECnodestrength | Sum_PC | 0.2030193 | 0.79698066 | 0.5 | 0.2547356 | 0.6092334040 | -0.01095706089 | 0.02254661 |
| alpha_ROI57_AECnodestrength | Blocktapping | 0.1934233 | 0.80657672 | 0.5 | 0.2398077 | 0.9610986453 | -0.00087332321 | 0.02266435 |
| alpha_ROI57_AECnodestrength | Sum_PC | 0.2149698 | 0.78503020 | 0.5 | 0.2738363 | 0.4936551765 | -0.01565283074 | 0.02482272 |
| alpha_ROI58_AECnodestrength | Blocktapping | 0.1956483 | 0.80435171 | 0.5 | 0.2432372 | 0.7549632179 | -0.00601993335 | 0.02377981 |
| alpha_ROI58_AECnodestrength | Sum_PC | 0.2081003 | 0.79189966 | 0.5 | 0.2627862 | 0.6440400631 | -0.00951495066 | 0.02290988 |
| alpha_ROI59_AECnodestrength | Blocktapping | 0.2035052 | 0.79649478 | 0.5 | 0.2555010 | 0.6268938927 | 0.01188473768 | 0.02463692 |
| alpha_ROI59_AECnodestrength | Sum_PC | 0.1969715 | 0.80302848 | 0.5 | 0.2452858 | 0.6996563014 | -0.00657166957 | 0.02187627 |
| alpha_ROI6_AECnodestrength | Blocktapping | 0.2179161 | 0.78208394 | 0.5 | 0.2786351 | 0.8049440716 | -0.00664628570 | 0.02323914 |
| alpha_ROI6_AECnodestrength | Sum_PC | 0.2892474 | 0.71075258 | 0.5 | 0.4069594 | 0.2415635123 | -0.03624139391 | 0.03110287 |
| alpha_ROI60_AECnodestrength | Blocktapping | 0.2059276 | 0.79407243 | 0.5 | 0.2593310 | 0.6151225903 | 0.01068344459 | 0.02508055 |
| alpha_ROI60_AECnodestrength | Sum_PC | 0.2079023 | 0.79209772 | 0.5 | 0.2624705 | 0.6011723576 | -0.01045404735 | 0.02320694 |
| alpha_ROI61_AECnodestrength | Blocktapping | 0.1882003 | 0.81179966 | 0.5 | 0.2318310 | 0.9526872955 | -0.00055893341 | 0.02400252 |
| alpha_ROI61_AECnodestrength | Sum_PC | 0.1872771 | 0.81272290 | 0.5 | 0.2304317 | 0.8212184820 | -0.00453163708 | 0.02112493 |
| alpha_ROI62_AECnodestrength | Blocktapping | 0.1824415 | 0.81755847 | 0.5 | 0.2231541 | 0.7671235231 | 0.00662638110 | 0.02312159 |
| alpha_ROI62_AECnodestrength | Sum_PC | 0.1829699 | 0.81703010 | 0.5 | 0.2239451 | 0.7729053827 | -0.00480407770 | 0.02080831 |
| alpha_ROI63_AECnodestrength | Blocktapping | 0.1890687 | 0.81093128 | 0.5 | 0.2331501 | 0.9675060931 | 0.00003480673 | 0.02380728 |
| alpha_ROI63_AECnodestrength | Sum_PC | 0.1977375 | 0.80226246 | 0.5 | 0.2464749 | 0.5968518396 | -0.01127394084 | 0.02244725 |
| alpha_ROI64_AECnodestrength | Blocktapping | 0.2063812 | 0.79361876 | 0.5 | 0.2600509 | 0.6929649164 | -0.00945693461 | 0.02358125 |
| alpha_ROI64_AECnodestrength | Sum_PC | 0.2399500 | 0.76005002 | 0.5 | 0.3157029 | 0.3848119156 | -0.02104543420 | 0.02647602 |
| alpha_ROI65_AECnodestrength | Blocktapping | 0.2198483 | 0.78015166 | 0.5 | 0.2818021 | 0.8723889146 | 0.00271252444 | 0.02316787 |
| alpha_ROI65_AECnodestrength | Sum_PC | 0.3230471 | 0.67695291 | 0.5 | 0.4772076 | 0.1788721489 | -0.04484909455 | 0.03563499 |
| alpha_ROI66_AECnodestrength | Blocktapping | 0.1894380 | 0.81056197 | 0.5 | 0.2337120 | 0.9914649051 | -0.00014952428 | 0.02221147 |
| alpha_ROI66_AECnodestrength | Sum_PC | 0.2032327 | 0.79676729 | 0.5 | 0.2550716 | 0.5775779649 | -0.01251501057 | 0.02319432 |
| alpha_ROI67_AECnodestrength | Blocktapping | 0.2521690 | 0.74783102 | 0.5 | 0.3372005 | 0.4836948725 | 0.01597685278 | 0.02645485 |
| alpha_ROI67_AECnodestrength | Sum_PC | 0.3218913 | 0.67810872 | 0.5 | 0.4746898 | 0.1998992948 | -0.03964831710 | 0.03493781 |
| alpha_ROI68_AECnodestrength | Blocktapping | 0.1989555 | 0.80104452 | 0.5 | 0.2483701 | 0.9417777275 | 0.00186247811 | 0.02401870 |
| alpha_ROI68_AECnodestrength | Sum_PC | 0.2274437 | 0.77255629 | 0.5 | 0.2944041 | 0.4400543157 | -0.01742342255 | 0.02593035 |
| alpha_ROI69_AECnodestrength | Blocktapping | 0.2576953 | 0.74230467 | 0.5 | 0.3471557 | 0.4501684278 | 0.01836883652 | 0.02789457 |

| DV | Predictor | Sum_PostModProb_Incl | Sum_PostModProb_Excl | Sum_PriorModProb_Incl | BF_Incl | Frequentist_p_value | postMean | postStd |
|---|---|---|---|---|---|---|---|---|
| alpha_ROI69_AECnodestrength | Sum_PC | 0.3249929 | 0.67500713 | 0.5 | 0.4814658 | 0.1879221744 | -0.04088224721 | 0.03398705 |
| alpha_ROI7_AECnodestrength | Blocktapping | 0.2925794 | 0.70742061 | 0.5 | 0.4135862 | 0.4566596852 | 0.01822105453 | 0.02766213 |
| alpha_ROI7_AECnodestrength | Sum_PC | 0.4318719 | 0.56812811 | 0.5 | 0.7601664 | 0.0955454192 | -0.06731400154 | 0.04480832 |
| alpha_ROI70_AECnodestrength | Blocktapping | 0.2458800 | 0.75411995 | 0.5 | 0.3260490 | 0.3940937509 | 0.02301391265 | 0.02927259 |
| alpha_ROI70_AECnodestrength | Sum_PC | 0.2326625 | 0.76733749 | 0.5 | 0.3032075 | 0.4715860944 | -0.01587674452 | 0.02464543 |
| alpha_ROI71_AECnodestrength | Blocktapping | 0.2320447 | 0.76795534 | 0.5 | 0.3021591 | 0.4478278790 | 0.01881808595 | 0.02804563 |
| alpha_ROI71_AECnodestrength | Sum_PC | 0.2350416 | 0.76495837 | 0.5 | 0.3072607 | 0.4151292667 | -0.01898933481 | 0.02533761 |
| alpha_ROI72_AECnodestrength | Blocktapping | 0.1888340 | 0.81116604 | 0.5 | 0.2327932 | 0.8582040199 | 0.00320289207 | 0.02300495 |
| alpha_ROI72_AECnodestrength | Sum_PC | 0.2015535 | 0.79844652 | 0.5 | 0.2524320 | 0.5805851357 | -0.01224995765 | 0.02276234 |
| alpha_ROI73_AECnodestrength | Blocktapping | 0.1859324 | 0.81406762 | 0.5 | 0.2283992 | 0.8010472392 | -0.00649645290 | 0.02313097 |
| alpha_ROI73_AECnodestrength | Sum_PC | 0.1882036 | 0.81179641 | 0.5 | 0.2318359 | 0.7348003540 | -0.00638139165 | 0.02135825 |
| alpha_ROI74_AECnodestrength | Blocktapping | 0.2650688 | 0.73493120 | 0.5 | 0.3606716 | 0.3409821952 | 0.02447001032 | 0.03034852 |
| alpha_ROI74_AECnodestrength | Sum_PC | 0.2789820 | 0.72101800 | 0.5 | 0.3869279 | 0.2885748300 | -0.02867138940 | 0.02928753 |
| alpha_ROI75_AECnodestrength | Blocktapping | 0.1903697 | 0.80963026 | 0.5 | 0.2351317 | 0.8612426344 | 0.00240509500 | 0.02287996 |
| alpha_ROI75_AECnodestrength | Sum_PC | 0.2032873 | 0.79671270 | 0.5 | 0.2551576 | 0.6113909814 | -0.01103112687 | 0.02385590 |
| alpha_ROI76_AECnodestrength | Blocktapping | 0.1880469 | 0.81195306 | 0.5 | 0.2315983 | 0.8131506127 | -0.00604390575 | 0.02346850 |
| alpha_ROI76_AECnodestrength | Sum_PC | 0.2023624 | 0.79763757 | 0.5 | 0.2537022 | 0.5873351999 | -0.01299012103 | 0.02267837 |
| alpha_ROI77_AECnodestrength | Blocktapping | 0.1978815 | 0.80211849 | 0.5 | 0.2466986 | 0.8551572247 | 0.00373864047 | 0.02314310 |
| alpha_ROI77_AECnodestrength | Sum_PC | 0.2432810 | 0.75671898 | 0.5 | 0.3214945 | 0.3443082349 | -0.02420375500 | 0.02736067 |
| alpha_ROI78_AECnodestrength | Blocktapping | 0.2283859 | 0.77161408 | 0.5 | 0.2959846 | 0.4956244190 | -0.01684715087 | 0.02664724 |
| alpha_ROI78_AECnodestrength | Sum_PC | 0.2312670 | 0.76873299 | 0.5 | 0.3008418 | 0.4621754932 | -0.01848397966 | 0.02563453 |
| alpha_ROI79_AECnodestrength | Blocktapping | 0.2053942 | 0.79460585 | 0.5 | 0.2584856 | 0.5624836866 | -0.01248832785 | 0.02607737 |
| alpha_ROI79_AECnodestrength | Sum_PC | 0.1897355 | 0.81026450 | 0.5 | 0.2341649 | 0.9294877289 | 0.00066902835 | 0.02119870 |
| alpha_ROI8_AECnodestrength | Blocktapping | 0.2229819 | 0.77701812 | 0.5 | 0.2869713 | 0.8183554117 | 0.00537905390 | 0.02329774 |
| alpha_ROI8_AECnodestrength | Sum_PC | 0.3304786 | 0.66952138 | 0.5 | 0.4936043 | 0.1627922991 | -0.04547139114 | 0.03564304 |
| alpha_ROI80_AECnodestrength | Blocktapping | 0.1950432 | 0.80495678 | 0.5 | 0.2423027 | 0.9399130776 | -0.00239853659 | 0.02316405 |
| alpha_ROI80_AECnodestrength | Sum_PC | 0.2199757 | 0.78002430 | 0.5 | 0.2820113 | 0.4865758349 | -0.01554976785 | 0.02535600 |
| alpha_ROI81_AECnodestrength | Blocktapping | 0.1886957 | 0.81130428 | 0.5 | 0.2325832 | 0.8821970401 | 0.00276237003 | 0.02320435 |
| alpha_ROI81_AECnodestrength | Sum_PC | 0.2016136 | 0.79838641 | 0.5 | 0.2525263 | 0.6270407946 | -0.00995807922 | 0.02286475 |

| DV | Predictor | Sum_PostModProb_Incl | Sum_PostModProb_Excl | Sum_PriorModProb_Incl | BF_Incl | Frequentist_p_value | postMean | postStd |
|---|---|---|---|---|---|---|---|---|
| alpha_ROI82_AECnodestrength | Blocktapping | 0.2387485 | 0.76125150 | 0.5 | 0.3136263 | 0.4864477200 | 0.01658889303 | 0.02679462 |
| alpha_ROI82_AECnodestrength | Sum_PC | 0.2775483 | 0.72245172 | 0.5 | 0.3841755 | 0.2688686783 | -0.02786069420 | 0.02983996 |
| alpha_ROI83_AECnodestrength | Blocktapping | 0.2082285 | 0.79177149 | 0.5 | 0.2629907 | 0.8309220924 | 0.00622143686 | 0.02518210 |
| alpha_ROI83_AECnodestrength | Sum_PC | 0.2254163 | 0.77458369 | 0.5 | 0.2910161 | 0.4367125907 | 0.01780439317 | 0.02550474 |
| alpha_ROI84_AECnodestrength | Blocktapping | 0.1643358 | 0.83566418 | 0.5 | 0.1966529 | 0.9739852615 | -0.00010290292 | 0.02089889 |
| alpha_ROI84_AECnodestrength | Sum_PC | 0.1692048 | 0.83079520 | 0.5 | 0.2036661 | 0.9464650729 | -0.00153053576 | 0.01998742 |
| alpha_ROI85_AECnodestrength | Blocktapping | 0.2120146 | 0.78798540 | 0.5 | 0.2690591 | 0.5084368781 | -0.01701274795 | 0.02545079 |
| alpha_ROI85_AECnodestrength | Sum_PC | 0.1975075 | 0.80249253 | 0.5 | 0.2461175 | 0.7204642071 | 0.00660347794 | 0.02262779 |
| alpha_ROI86_AECnodestrength | Blocktapping | 0.1918587 | 0.80814128 | 0.5 | 0.2374074 | 0.8057878789 | 0.00522070734 | 0.02391813 |
| alpha_ROI86_AECnodestrength | Sum_PC | 0.1989280 | 0.80107203 | 0.5 | 0.2483272 | 0.6309073366 | -0.00975592122 | 0.02276462 |
| alpha_ROI87_AECnodestrength | Blocktapping | 0.2194030 | 0.78059700 | 0.5 | 0.2810708 | 0.5081934604 | 0.01446091368 | 0.02634722 |
| alpha_ROI87_AECnodestrength | Sum_PC | 0.2290290 | 0.77097099 | 0.5 | 0.2970657 | 0.4293012351 | -0.01806753421 | 0.02507479 |
| alpha_ROI88_AECnodestrength | Blocktapping | 0.1851695 | 0.81483049 | 0.5 | 0.2272491 | 0.9376464581 | -0.00147146956 | 0.02329185 |
| alpha_ROI88_AECnodestrength | Sum_PC | 0.1869452 | 0.81305478 | 0.5 | 0.2299294 | 0.8997803935 | 0.00174164190 | 0.02122744 |
| alpha_ROI89_AECnodestrength | Blocktapping | 0.1898612 | 0.81013883 | 0.5 | 0.2343563 | 0.7417874220 | 0.00679573681 | 0.02271438 |
| alpha_ROI89_AECnodestrength | Sum_PC | 0.2184542 | 0.78154577 | 0.5 | 0.2795156 | 0.4077988260 | -0.01777480324 | 0.02399382 |
| alpha_ROI9_AECnodestrength | Blocktapping | 0.1966941 | 0.80330591 | 0.5 | 0.2448558 | 0.8391655745 | -0.00534298332 | 0.02327878 |
| alpha_ROI9_AECnodestrength | Sum_PC | 0.2331426 | 0.76685739 | 0.5 | 0.3040234 | 0.4052287673 | -0.02155063871 | 0.02639828 |
| alpha_ROI90_AECnodestrength | Blocktapping | 0.2174180 | 0.78258196 | 0.5 | 0.2778214 | 0.6350749424 | 0.00873413025 | 0.02317186 |
| alpha_ROI90_AECnodestrength | Sum_PC | 0.2919562 | 0.70804384 | 0.5 | 0.4123419 | 0.2591460485 | -0.03415451876 | 0.03299371 |
| alpha_ROI91_AECnodestrength | Blocktapping | 0.1974906 | 0.80250945 | 0.5 | 0.2460912 | 0.9973350341 | -0.00012920838 | 0.02296676 |
| alpha_ROI91_AECnodestrength | Sum_PC | 0.2397377 | 0.76026229 | 0.5 | 0.3153355 | 0.3769753950 | -0.02179019503 | 0.02694223 |
| alpha_ROI92_AECnodestrength | Blocktapping | 0.2106340 | 0.78936602 | 0.5 | 0.2668394 | 0.5198052064 | -0.01810357225 | 0.02641985 |
| alpha_ROI92_AECnodestrength | Sum_PC | 0.1922965 | 0.80770350 | 0.5 | 0.2380781 | 0.8733748115 | -0.00240440591 | 0.02100134 |
| alpha_ROI93_AECnodestrength | Blocktapping | 0.2071847 | 0.79281531 | 0.5 | 0.2613278 | 0.6659988592 | 0.00965127433 | 0.02493540 |
| alpha_ROI93_AECnodestrength | Sum_PC | 0.2355475 | 0.76445251 | 0.5 | 0.3081257 | 0.3931010227 | -0.02099846363 | 0.02590901 |
| alpha_ROI94_AECnodestrength | Blocktapping | 0.1943540 | 0.80564595 | 0.5 | 0.2412400 | 0.7307212995 | -0.00694625562 | 0.02413604 |
| alpha_ROI94_AECnodestrength | Sum_PC | 0.1907005 | 0.80929951 | 0.5 | 0.2356365 | 0.8231314525 | -0.00474861610 | 0.02171774 |
| alpha_ROI95_AECnodestrength | Blocktapping | 0.2024485 | 0.79755152 | 0.5 | 0.2538375 | 0.7437229174 | -0.00822521227 | 0.02409791 |

| DV | Predictor | Sum_PostModProb_Incl | Sum_PostModProb_Excl | Sum_PriorModProb_Incl | BF_Incl | Frequentist_p_value | postMean | postStd |
|---|---|---|---|---|---|---|---|---|
| alpha_ROI95_AEC nodestrength | Sum_PC | 0.2230077 | 0.77699234 | 0.5 | 0.2870140 | 0.4638074073 | -0.01739957898 | 0.02460517 |
| alpha_ROI96_AEC nodestrength | Blocktapping | 0.2143831 | 0.78561685 | 0.5 | 0.2728851 | 0.4977621538 | -0.01655434785 | 0.02687826 |
| alpha_ROI96_AEC nodestrength | Sum_PC | 0.2037254 | 0.79627462 | 0.5 | 0.2558481 | 0.8728246318 | 0.00202909186 | 0.02229211 |
| alpha_ROI97_AEC nodestrength | Blocktapping | 0.1835815 | 0.81641850 | 0.5 | 0.2248620 | 0.9115613824 | -0.00109349591 | 0.02327025 |
| alpha_ROI97_AEC nodestrength | Sum_PC | 0.1881541 | 0.81184593 | 0.5 | 0.2317608 | 0.7442477442 | 0.00643245856 | 0.02102550 |
| alpha_ROI98_AEC nodestrength | Blocktapping | 0.1899748 | 0.81002525 | 0.5 | 0.2345294 | 0.7985519731 | -0.00416169498 | 0.02427948 |
| alpha_ROI98_AEC nodestrength | Sum_PC | 0.1883337 | 0.81166626 | 0.5 | 0.2320335 | 0.8603234253 | 0.00334212754 | 0.02178135 |
| alpha_ROI99_AEC nodestrength | Blocktapping | 0.2018595 | 0.79814046 | 0.5 | 0.2529123 | 0.6784183860 | 0.00868756699 | 0.02404851 |
| alpha_ROI99_AEC nodestrength | Sum_PC | 0.2176129 | 0.78238709 | 0.5 | 0.2781397 | 0.4632737308 | -0.01538678691 | 0.02446143 |
| AlphaPow1 | Blocktapping | 0.3163764 | 0.68362358 | 0.5 | 0.4627933 | 0.1860018899 | 0.04398797076 | 0.03768841 |
| AlphaPow1 | Sum_PC | 0.2374630 | 0.76253704 | 0.5 | 0.3114117 | 0.5837925964 | -0.01119911989 | 0.02264123 |
| AlphaPow10 | Blocktapping | 0.1836058 | 0.81639416 | 0.5 | 0.2248985 | 0.7873563229 | 0.00483381064 | 0.02232784 |
| AlphaPow10 | Sum_PC | 0.1802777 | 0.81972234 | 0.5 | 0.2199253 | 0.9885775787 | -0.00038195537 | 0.01974006 |
| AlphaPow100 | Blocktapping | 0.3799127 | 0.62008727 | 0.5 | 0.6126762 | 0.1446485081 | 0.05381297327 | 0.04302934 |
| AlphaPow100 | Sum_PC | 0.3158770 | 0.68412302 | 0.5 | 0.4617254 | 0.3036304902 | -0.02612048387 | 0.02933142 |
| AlphaPow11 | Blocktapping | 0.2317973 | 0.76820271 | 0.5 | 0.3017397 | 0.3637542358 | 0.02446506911 | 0.02853218 |
| AlphaPow11 | Sum_PC | 0.1920822 | 0.80791777 | 0.5 | 0.2377497 | 0.8663414506 | 0.00381866728 | 0.02037627 |
| AlphaPow12 | Blocktapping | 0.2252486 | 0.77475144 | 0.5 | 0.2907365 | 0.4916102959 | 0.01728567141 | 0.02700997 |
| AlphaPow12 | Sum_PC | 0.2096549 | 0.79034509 | 0.5 | 0.2652701 | 0.7366642573 | 0.00699208541 | 0.02161934 |
| AlphaPow13 | Blocktapping | 0.2918379 | 0.70816214 | 0.5 | 0.4121060 | 0.2019835774 | 0.04167809902 | 0.03634425 |
| AlphaPow13 | Sum_PC | 0.2038662 | 0.79613384 | 0.5 | 0.2560702 | 0.9882275939 | -0.00027937145 | 0.01954175 |
| AlphaPow14 | Blocktapping | 0.3512877 | 0.64871229 | 0.5 | 0.5415154 | 0.1534269056 | 0.05535543488 | 0.04159394 |
| AlphaPow14 | Sum_PC | 0.2388239 | 0.76117615 | 0.5 | 0.3137564 | 0.6372811982 | 0.01016717019 | 0.02215845 |
| AlphaPow15 | Blocktapping | 0.4331821 | 0.56681791 | 0.5 | 0.7642350 | 0.0977373651 | 0.07753460118 | 0.05098828 |
| AlphaPow15 | Sum_PC | 0.2509179 | 0.74908209 | 0.5 | 0.3349672 | 0.8442398737 | 0.00544570068 | 0.02092422 |
| AlphaPow16 | Blocktapping | 0.2355357 | 0.76446428 | 0.5 | 0.3081056 | 0.3920361533 | 0.02134636698 | 0.02828856 |
| AlphaPow16 | Sum_PC | 0.2137208 | 0.78627917 | 0.5 | 0.2718129 | 0.5693947340 | -0.01156188529 | 0.02223148 |
| AlphaPow17 | Blocktapping | 0.2282159 | 0.77178415 | 0.5 | 0.2956991 | 0.3906976136 | 0.02134497269 | 0.02747534 |
| AlphaPow17 | Sum_PC | 0.1915987 | 0.80840135 | 0.5 | 0.2370093 | 0.9543669452 | 0.00107953443 | 0.02070915 |
| AlphaPow18 | Blocktapping | 0.4895467 | 0.51045329 | 0.5 | 0.9590431 | 0.0683588088 | 0.09594664646 | 0.05457059 |
| AlphaPow18 | Sum_PC | 0.2749416 | 0.72505842 | 0.5 | 0.3791992 | 0.8750387968 | -0.00237997581 | 0.02212318 |
| AlphaPow19 | Blocktapping | 0.2981854 | 0.70181464 | 0.5 | 0.4248777 | 0.2106818304 | 0.04386133244 | 0.03672534 |
| AlphaPow19 | Sum_PC | 0.2172386 | 0.78276136 | 0.5 | 0.2775286 | 0.7211224208 | 0.00728150359 | 0.02112796 |
| AlphaPow2 | Blocktapping | 0.2672587 | 0.73274126 | 0.5 | 0.3647382 | 0.2864828666 | 0.03059091070 | 0.03188387 |
| AlphaPow2 | Sum_PC | 0.2228679 | 0.77713215 | 0.5 | 0.2867824 | 0.5945625038 | -0.01160427093 | 0.02331230 |
| AlphaPow20 | Blocktapping | 0.4743625 | 0.52563747 | 0.5 | 0.9024519 | 0.0706170741 | 0.08970371573 | 0.05431713 |
| AlphaPow20 | Sum_PC | 0.2754778 | 0.72452220 | 0.5 | 0.3802200 | 0.6236243958 | -0.00901971212 | 0.02145428 |

| DV | Predictor | Sum_PostModProb_Incl | Sum_PostModProb_Excl | Sum_PriorModProb_Incl | BF_Incl | Frequentist_p_value | postMean | postStd |
|---|---|---|---|---|---|---|---|---|
| AlphaPow21 | Blocktapping | 0.4915418 | 0.50845819 | 0.5 | 0.9667300 | 0.0680223622 | 0.09560626757 | 0.05594991 |
| AlphaPow21 | Sum_PC | 0.2640440 | 0.73595597 | 0.5 | 0.3587769 | 0.8955485199 | 0.00347898884 | 0.02152332 |
| AlphaPow22 | Blocktapping | 0.2868393 | 0.71316066 | 0.5 | 0.4022086 | 0.2435485851 | 0.03706022234 | 0.03435014 |
| AlphaPow22 | Sum_PC | 0.2292703 | 0.77072973 | 0.5 | 0.2974717 | 0.5322239311 | 0.01394463008 | 0.02255193 |
| AlphaPow23 | Blocktapping | 0.3759823 | 0.62401768 | 0.5 | 0.6025187 | 0.1369633061 | 0.06055469915 | 0.04456187 |
| AlphaPow23 | Sum_PC | 0.2594487 | 0.74055131 | 0.5 | 0.3503453 | 0.5447441340 | 0.01487743616 | 0.02377781 |
| AlphaPow24 | Blocktapping | 0.2087562 | 0.79124384 | 0.5 | 0.2638329 | 0.4740015057 | 0.01734229656 | 0.02641043 |
| AlphaPow24 | Sum_PC | 0.1843528 | 0.81564722 | 0.5 | 0.2260202 | 0.9898740397 | 0.00053478483 | 0.02022165 |
| AlphaPow25 | Blocktapping | 0.2197061 | 0.78029393 | 0.5 | 0.2815683 | 0.4193855948 | 0.02112582594 | 0.02653460 |
| AlphaPow25 | Sum_PC | 0.1909188 | 0.80908117 | 0.5 | 0.2359699 | 0.7272650580 | 0.00807632377 | 0.02016804 |
| AlphaPow26 | Blocktapping | 0.2157690 | 0.78423097 | 0.5 | 0.2751345 | 0.4485268829 | 0.01870036138 | 0.02592795 |
| AlphaPow26 | Sum_PC | 0.1984406 | 0.80155939 | 0.5 | 0.2475682 | 0.6778749358 | 0.00833896809 | 0.02100306 |
| AlphaPow27 | Blocktapping | 0.1716951 | 0.82830488 | 0.5 | 0.2072849 | 0.7614877412 | 0.00513956779 | 0.02136569 |
| AlphaPow27 | Sum_PC | 0.1742603 | 0.82573966 | 0.5 | 0.2110355 | 0.8699141024 | -0.00241872198 | 0.01949158 |
| AlphaPow28 | Blocktapping | 0.2188994 | 0.78110057 | 0.5 | 0.2802449 | 0.4090562730 | 0.02109459762 | 0.02783136 |
| AlphaPow28 | Sum_PC | 0.1858841 | 0.81411588 | 0.5 | 0.2283264 | 0.8526337645 | 0.00479324384 | 0.02047125 |
| AlphaPow29 | Blocktapping | 0.3860095 | 0.61399052 | 0.5 | 0.6286896 | 0.1252481933 | 0.06338935797 | 0.04409124 |
| AlphaPow29 | Sum_PC | 0.2381684 | 0.76183164 | 0.5 | 0.3126260 | 0.8806855888 | -0.00197507820 | 0.02084285 |
| AlphaPow3 | Blocktapping | 0.2669747 | 0.73302528 | 0.5 | 0.3642094 | 0.2735453561 | 0.03198568816 | 0.03220612 |
| AlphaPow3 | Sum_PC | 0.2126168 | 0.78738318 | 0.5 | 0.2700297 | 0.6576460982 | -0.00863754134 | 0.02204316 |
| AlphaPow30 | Blocktapping | 0.3384286 | 0.66157141 | 0.5 | 0.5115526 | 0.1681781164 | 0.05120790715 | 0.04156024 |
| AlphaPow30 | Sum_PC | 0.2283526 | 0.77164744 | 0.5 | 0.2959286 | 0.7405276428 | 0.00645297617 | 0.02157888 |
| AlphaPow31 | Blocktapping | 0.1902869 | 0.80971309 | 0.5 | 0.2350053 | 0.5542881839 | 0.01244036829 | 0.02419847 |
| AlphaPow31 | Sum_PC | 0.1751514 | 0.82484859 | 0.5 | 0.2123437 | 0.9487371469 | -0.00110007015 | 0.01923675 |
| AlphaPow32 | Blocktapping | 0.2451156 | 0.75488442 | 0.5 | 0.3247061 | 0.3191081290 | 0.02655943837 | 0.02906695 |
| AlphaPow32 | Sum_PC | 0.1940879 | 0.80591206 | 0.5 | 0.2408302 | 0.9901982935 | -0.00003273273 | 0.01986660 |
| AlphaPow33 | Blocktapping | 0.2295215 | 0.77047854 | 0.5 | 0.2978947 | 0.3830803565 | 0.02033459990 | 0.02739041 |
| AlphaPow33 | Sum_PC | 0.2005195 | 0.79948049 | 0.5 | 0.2508123 | 0.6591091867 | -0.00871557867 | 0.02047381 |
| AlphaPow34 | Blocktapping | 0.4219505 | 0.57804950 | 0.5 | 0.7299557 | 0.1015748992 | 0.07541945814 | 0.04888025 |
| AlphaPow34 | Sum_PC | 0.2522506 | 0.74774939 | 0.5 | 0.3373465 | 0.7759057356 | 0.00646229012 | 0.02233184 |
| AlphaPow35 | Blocktapping | 0.1826885 | 0.81731153 | 0.5 | 0.2235237 | 0.7121904238 | 0.00854877487 | 0.02251311 |
| AlphaPow35 | Sum_PC | 0.1920300 | 0.80796998 | 0.5 | 0.2376697 | 0.5879117422 | 0.01035013307 | 0.02191098 |
| AlphaPow36 | Blocktapping | 0.3605501 | 0.63944993 | 0.5 | 0.5638441 | 0.1455411631 | 0.05428816017 | 0.04186863 |
| AlphaPow36 | Sum_PC | 0.2594786 | 0.74052143 | 0.5 | 0.3503998 | 0.5262301724 | -0.01355484916 | 0.02409783 |
| AlphaPow37 | Blocktapping | 0.3862936 | 0.61370639 | 0.5 | 0.6294437 | 0.1195410299 | 0.06552581023 | 0.04679761 |
| AlphaPow37 | Sum_PC | 0.2359311 | 0.76406894 | 0.5 | 0.3087824 | 0.7956790318 | -0.00482410077 | 0.02122754 |
| AlphaPow38 | Blocktapping | 0.1988364 | 0.80116363 | 0.5 | 0.2481845 | 0.5639497909 | 0.01240932953 | 0.02427257 |
| AlphaPow38 | Sum_PC | 0.1903581 | 0.80964186 | 0.5 | 0.2351140 | 0.6800190248 | 0.00772892496 | 0.02044843 |
| AlphaPow39 | Blocktapping | 0.2113653 | 0.78863466 | 0.5 | 0.2680143 | 0.4825544261 | 0.01464946732 | 0.02477790 |
| AlphaPow39 | Sum_PC | 0.1902782 | 0.80972180 | 0.5 | 0.2349921 | 0.9616944358 | -0.00119522369 | 0.01953917 |
| AlphaPow4 | Blocktapping | 0.3371371 | 0.66286287 | 0.5 | 0.5086077 | 0.1848047868 | 0.04637180892 | 0.03821547 |

| DV | Predictor | Sum_PostModProb_Incl | Sum_PostModProb_Excl | Sum_PriorModProb_Incl | BF_Incl | Frequentist_p_value | postMean | postStd |
|---|---|---|---|---|---|---|---|---|
| AlphaPow4 | Sum_PC | 0.2593245 | 0.74067546 | 0.5 | 0.3501190 | 0.4511742938 | -0.01591335718 | 0.02516373 |
| AlphaPow40 | Blocktapping | 0.2226766 | 0.77732341 | 0.5 | 0.2864658 | 0.4268626812 | 0.01955435795 | 0.02694968 |
| AlphaPow40 | Sum_PC | 0.2074450 | 0.79255503 | 0.5 | 0.2617420 | 0.7442741074 | -0.00668978736 | 0.02239444 |
| AlphaPow41 | Blocktapping | 0.3390362 | 0.66096375 | 0.5 | 0.5129423 | 0.1626045259 | 0.05331667057 | 0.04163689 |
| AlphaPow41 | Sum_PC | 0.2264756 | 0.77352437 | 0.5 | 0.2927841 | 0.8556834297 | 0.00353241153 | 0.02116080 |
| AlphaPow42 | Blocktapping | 0.1997698 | 0.80023021 | 0.5 | 0.2496404 | 0.5058457347 | 0.01514437997 | 0.02396715 |
| AlphaPow42 | Sum_PC | 0.1804166 | 0.81958345 | 0.5 | 0.2201320 | 0.8891325177 | 0.00256561621 | 0.01888208 |
| AlphaPow43 | Blocktapping | 0.1855969 | 0.81440312 | 0.5 | 0.2278931 | 0.5902861853 | 0.01152098955 | 0.02270181 |
| AlphaPow43 | Sum_PC | 0.1747875 | 0.82521249 | 0.5 | 0.2118091 | 0.8710206761 | 0.00329066956 | 0.01886512 |
| AlphaPow44 | Blocktapping | 0.1816461 | 0.81835393 | 0.5 | 0.2219652 | 0.6447267883 | 0.00840331480 | 0.02236816 |
| AlphaPow44 | Sum_PC | 0.1848232 | 0.81517679 | 0.5 | 0.2267278 | 0.6044470373 | -0.00913832004 | 0.02147549 |
| AlphaPow45 | Blocktapping | 0.1507042 | 0.84929580 | 0.5 | 0.1774461 | 0.9533982745 | -0.00203067514 | 0.01850984 |
| AlphaPow45 | Sum_PC | 0.1528526 | 0.84714745 | 0.5 | 0.1804321 | 0.9334377174 | -0.00149804382 | 0.01742098 |
| AlphaPow46 | Blocktapping | 0.1697542 | 0.83024580 | 0.5 | 0.2044626 | 0.7881572143 | 0.00521858951 | 0.02094639 |
| AlphaPow46 | Sum_PC | 0.1700781 | 0.82992191 | 0.5 | 0.2049326 | 0.7639709945 | -0.00565926858 | 0.01954637 |
| AlphaPow47 | Blocktapping | 0.3191199 | 0.68088015 | 0.5 | 0.4686873 | 0.1921406293 | 0.04776027825 | 0.03621694 |
| AlphaPow47 | Sum_PC | 0.2390584 | 0.76094163 | 0.5 | 0.3141612 | 0.5320254298 | 0.01415131771 | 0.02356401 |
| AlphaPow48 | Blocktapping | 0.2232919 | 0.77670809 | 0.5 | 0.2874850 | 0.3974720971 | 0.02301430731 | 0.02773547 |
| AlphaPow48 | Sum_PC | 0.1932924 | 0.80670758 | 0.5 | 0.2396066 | 0.7223924396 | 0.00691109119 | 0.02038542 |
| AlphaPow49 | Blocktapping | 0.2936501 | 0.70634985 | 0.5 | 0.4157290 | 0.2242507624 | 0.03935308290 | 0.03631500 |
| AlphaPow49 | Sum_PC | 0.2250363 | 0.77496374 | 0.5 | 0.2903829 | 0.6521751664 | -0.00832779286 | 0.02186053 |
| AlphaPow5 | Blocktapping | 0.3152861 | 0.68471388 | 0.5 | 0.4604640 | 0.1974876084 | 0.04221558911 | 0.03683505 |
| AlphaPow5 | Sum_PC | 0.2455700 | 0.75442997 | 0.5 | 0.3255041 | 0.5096678155 | -0.01390217191 | 0.02396076 |
| AlphaPow50 | Blocktapping | 0.3800794 | 0.61992059 | 0.5 | 0.6131098 | 0.1307153745 | 0.06210419107 | 0.04358927 |
| AlphaPow50 | Sum_PC | 0.2788357 | 0.72116430 | 0.5 | 0.3866466 | 0.4351482614 | -0.01764554897 | 0.02553031 |
| AlphaPow51 | Blocktapping | 0.2640483 | 0.73595166 | 0.5 | 0.3587849 | 0.2861275763 | 0.03020534018 | 0.03231695 |
| AlphaPow51 | Sum_PC | 0.2112054 | 0.78879462 | 0.5 | 0.2677571 | 0.7017122011 | -0.00701758301 | 0.02160604 |
| AlphaPow52 | Blocktapping | 0.2767682 | 0.72323178 | 0.5 | 0.3826826 | 0.2710557881 | 0.03380941773 | 0.03290042 |
| AlphaPow52 | Sum_PC | 0.2158167 | 0.78418326 | 0.5 | 0.2752121 | 0.7068468624 | -0.00620092503 | 0.02208714 |
| AlphaPow53 | Blocktapping | 0.1873977 | 0.81260226 | 0.5 | 0.2306143 | 0.7367965850 | 0.00685933674 | 0.02324165 |
| AlphaPow53 | Sum_PC | 0.1839694 | 0.81603060 | 0.5 | 0.2254442 | 0.9145727826 | 0.00131583478 | 0.02030560 |
| AlphaPow54 | Blocktapping | 0.3359182 | 0.66408182 | 0.5 | 0.5058385 | 0.1670690827 | 0.04887797462 | 0.03848922 |
| AlphaPow54 | Sum_PC | 0.2355304 | 0.76446955 | 0.5 | 0.3080966 | 0.7015671166 | -0.00783329029 | 0.02139461 |
| AlphaPow55 | Blocktapping | 0.3386274 | 0.66137262 | 0.5 | 0.5120070 | 0.1657282239 | 0.04898112036 | 0.03958078 |
| AlphaPow55 | Sum_PC | 0.2513973 | 0.74860269 | 0.5 | 0.3358221 | 0.5150496691 | -0.01302416716 | 0.02345139 |
| AlphaPow56 | Blocktapping | 0.3541660 | 0.64583403 | 0.5 | 0.5483854 | 0.1795301912 | 0.04776190612 | 0.04017459 |
| AlphaPow56 | Sum_PC | 0.2568286 | 0.74317144 | 0.5 | 0.3455845 | 0.4472033472 | -0.01660030958 | 0.02419100 |
| AlphaPow57 | Blocktapping | 0.2768900 | 0.72310996 | 0.5 | 0.3829155 | 0.2556391970 | 0.03394074721 | 0.03247775 |
| AlphaPow57 | Sum_PC | 0.2205090 | 0.77949096 | 0.5 | 0.2828885 | 0.6377925258 | -0.00951058785 | 0.02150033 |
| AlphaPow58 | Blocktapping | 0.4093253 | 0.59067474 | 0.5 | 0.6929791 | 0.1138545001 | 0.06498454564 | 0.04571035 |
| AlphaPow58 | Sum_PC | 0.3033503 | 0.69664973 | 0.5 | 0.4354416 | 0.3584302906 | -0.02264810907 | 0.02675670 |

| DV | Predictor | Sum_PostModProb_Incl | Sum_PostModProb_Excl | Sum_PriorModProb_Incl | BF_Incl | Frequentist_p_value | postMean | postStd |
|---|---|---|---|---|---|---|---|---|
| AlphaPow59 | Blocktapping | 0.1988467 | 0.80115333 | 0.5 | 0.2482005 | 0.5113786052 | 0.01462318679 | 0.02509337 |
| AlphaPow59 | Sum_PC | 0.1771014 | 0.82289863 | 0.5 | 0.2152165 | 0.9332568842 | -0.00143483870 | 0.01960429 |
| AlphaPow6 | Blocktapping | 0.2576562 | 0.74234376 | 0.5 | 0.3470848 | 0.2938163926 | 0.02961181273 | 0.03108143 |
| AlphaPow6 | Sum_PC | 0.2070243 | 0.79297571 | 0.5 | 0.2610727 | 0.6808278102 | -0.00765329174 | 0.02144061 |
| AlphaPow60 | Blocktapping | 0.2231414 | 0.77685865 | 0.5 | 0.2872355 | 0.3688527992 | 0.02387401291 | 0.02809246 |
| AlphaPow60 | Sum_PC | 0.1880096 | 0.81199044 | 0.5 | 0.2315416 | 0.8539282029 | -0.00328751487 | 0.02042562 |
| AlphaPow61 | Blocktapping | 0.1866324 | 0.81336755 | 0.5 | 0.2294565 | 0.6175203829 | 0.01026908366 | 0.02284549 |
| AlphaPow61 | Sum_PC | 0.1761274 | 0.82387261 | 0.5 | 0.2137799 | 0.8312671507 | 0.00406612244 | 0.01909551 |
| AlphaPow62 | Blocktapping | 0.1983738 | 0.80162623 | 0.5 | 0.2474642 | 0.4842084194 | 0.01577180164 | 0.02478548 |
| AlphaPow62 | Sum_PC | 0.1761038 | 0.82389618 | 0.5 | 0.2137452 | 0.9352672410 | -0.00202429469 | 0.01896804 |
| AlphaPow63 | Blocktapping | 0.2269162 | 0.77308379 | 0.5 | 0.2935208 | 0.3663446176 | 0.02201693906 | 0.02800613 |
| AlphaPow63 | Sum_PC | 0.1869574 | 0.81304263 | 0.5 | 0.2299478 | 0.8476302800 | -0.00331112387 | 0.02000489 |
| AlphaPow64 | Blocktapping | 0.2946612 | 0.70533883 | 0.5 | 0.4177583 | 0.2081002792 | 0.03993399575 | 0.03591454 |
| AlphaPow64 | Sum_PC | 0.2176800 | 0.78232003 | 0.5 | 0.2782493 | 0.6855480293 | -0.00751086276 | 0.02138143 |
| AlphaPow65 | Blocktapping | 0.3769466 | 0.62305341 | 0.5 | 0.6049988 | 0.1274803595 | 0.05948731232 | 0.04454350 |
| AlphaPow65 | Sum_PC | 0.2734972 | 0.72650284 | 0.5 | 0.3764571 | 0.4261744151 | -0.01760844754 | 0.02529052 |
| AlphaPow66 | Blocktapping | 0.4138801 | 0.58611989 | 0.5 | 0.7061356 | 0.1018880237 | 0.07167321734 | 0.04887102 |
| AlphaPow66 | Sum_PC | 0.2416032 | 0.75839681 | 0.5 | 0.3185709 | 0.9449120740 | 0.00369511639 | 0.02018113 |
| AlphaPow67 | Blocktapping | 0.2127750 | 0.78722505 | 0.5 | 0.2702848 | 0.4688805483 | 0.01674515161 | 0.02613867 |
| AlphaPow67 | Sum_PC | 0.1905251 | 0.80947488 | 0.5 | 0.2353688 | 0.9466830515 | -0.00196557355 | 0.02100856 |
| AlphaPow68 | Blocktapping | 0.2212633 | 0.77873665 | 0.5 | 0.2841312 | 0.3676744922 | 0.02226237084 | 0.02667130 |
| AlphaPow68 | Sum_PC | 0.1954718 | 0.80452820 | 0.5 | 0.2429645 | 0.6794254754 | -0.00873499010 | 0.02097937 |
| AlphaPow69 | Blocktapping | 0.3414712 | 0.65852881 | 0.5 | 0.5185364 | 0.1816445092 | 0.04588461213 | 0.03713436 |
| AlphaPow69 | Sum_PC | 0.2947502 | 0.70524976 | 0.5 | 0.4179374 | 0.2860454630 | -0.02653615515 | 0.02857751 |
| AlphaPow7 | Blocktapping | 0.2714897 | 0.72851029 | 0.5 | 0.3726642 | 0.2699165683 | 0.03276487961 | 0.03269251 |
| AlphaPow7 | Sum_PC | 0.2090542 | 0.79094580 | 0.5 | 0.2643091 | 0.8783055831 | -0.00235352179 | 0.02044005 |
| AlphaPow70 | Blocktapping | 0.3244577 | 0.67554226 | 0.5 | 0.4802923 | 0.2066964454 | 0.03983935493 | 0.03679775 |
| AlphaPow70 | Sum_PC | 0.2746426 | 0.72535740 | 0.5 | 0.3786307 | 0.3805964083 | -0.02167601357 | 0.02632736 |
| AlphaPow71 | Blocktapping | 0.3708780 | 0.62912200 | 0.5 | 0.5895168 | 0.1327844323 | 0.06093997019 | 0.04344291 |
| AlphaPow71 | Sum_PC | 0.2465181 | 0.75348192 | 0.5 | 0.3271719 | 0.6568184383 | -0.00874902432 | 0.02229547 |
| AlphaPow72 | Blocktapping | 0.2327722 | 0.76722785 | 0.5 | 0.3033938 | 0.3386814350 | 0.02555064346 | 0.02929276 |
| AlphaPow72 | Sum_PC | 0.1902981 | 0.80970189 | 0.5 | 0.2350224 | 0.8104337847 | -0.00376703245 | 0.01988826 |
| AlphaPow73 | Blocktapping | 0.2654866 | 0.73451336 | 0.5 | 0.3614456 | 0.2834240766 | 0.03242875725 | 0.03263284 |
| AlphaPow73 | Sum_PC | 0.2048776 | 0.79512242 | 0.5 | 0.2576680 | 0.8233893079 | 0.00427869925 | 0.02098377 |
| AlphaPow74 | Blocktapping | 0.2015026 | 0.79849742 | 0.5 | 0.2523522 | 0.5023168403 | 0.01475208064 | 0.02511838 |
| AlphaPow74 | Sum_PC | 0.1868652 | 0.81313479 | 0.5 | 0.2298084 | 0.8875964340 | -0.00281266740 | 0.02032658 |
| AlphaPow75 | Blocktapping | 0.1880203 | 0.81197967 | 0.5 | 0.2315579 | 0.5535179801 | 0.01230016651 | 0.02339780 |
| AlphaPow75 | Sum_PC | 0.1783937 | 0.82160630 | 0.5 | 0.2171280 | 0.8115679426 | -0.00330583369 | 0.01913914 |
| AlphaPow76 | Blocktapping | 0.2246419 | 0.77535808 | 0.5 | 0.2897267 | 0.3935477327 | 0.02115882776 | 0.02849598 |
| AlphaPow76 | Sum_PC | 0.1848680 | 0.81513202 | 0.5 | 0.2267951 | 0.8136911660 | 0.00465340708 | 0.01956632 |
| AlphaPow77 | Blocktapping | 0.3917308 | 0.60826918 | 0.5 | 0.6440090 | 0.1191684955 | 0.06323078168 | 0.04520969 |

| DV | Predictor | Sum_PostModProb_Incl | Sum_PostModProb_Excl | Sum_PriorModProb_Incl | BF_Incl | Frequentist_p_value | postMean | postStd |
|---|---|---|---|---|---|---|---|---|
| AlphaPow77 | Sum_PC | 0.2608771 | 0.73912291 | 0.5 | 0.3529550 | 0.5702579302 | -0.01164241535 | 0.02318180 |
| AlphaPow78 | Blocktapping | 0.2796022 | 0.72039782 | 0.5 | 0.3881219 | 0.2415654532 | 0.03769618841 | 0.03382840 |
| AlphaPow78 | Sum_PC | 0.2029891 | 0.79701087 | 0.5 | 0.2546880 | 0.8743893478 | 0.00400338531 | 0.02024661 |
| AlphaPow79 | Blocktapping | 0.2040398 | 0.79596017 | 0.5 | 0.2563443 | 0.4501648970 | 0.01867734079 | 0.02507615 |
| AlphaPow79 | Sum_PC | 0.1793695 | 0.82063049 | 0.5 | 0.2185752 | 0.9749762537 | 0.00088797027 | 0.01961003 |
| AlphaPow8 | Blocktapping | 0.3515968 | 0.64840324 | 0.5 | 0.5422502 | 0.1616964817 | 0.05088956465 | 0.04161330 |
| AlphaPow8 | Sum_PC | 0.2589414 | 0.74105859 | 0.5 | 0.3494210 | 0.4987607926 | -0.01460152842 | 0.02456306 |
| AlphaPow80 | Blocktapping | 0.2374124 | 0.76258758 | 0.5 | 0.3113248 | 0.3512119740 | 0.02428371201 | 0.02888622 |
| AlphaPow80 | Sum_PC | 0.1925676 | 0.80743240 | 0.5 | 0.2384938 | 0.9995667880 | -0.00009956201 | 0.02041918 |
| AlphaPow81 | Blocktapping | 0.2172855 | 0.78271451 | 0.5 | 0.2776050 | 0.4449013698 | 0.01760231282 | 0.02546810 |
| AlphaPow81 | Sum_PC | 0.2113699 | 0.78863005 | 0.5 | 0.2680217 | 0.5287460317 | -0.01244826117 | 0.02169633 |
| AlphaPow82 | Blocktapping | 0.2737811 | 0.72621890 | 0.5 | 0.3769953 | 0.2466046312 | 0.03440290255 | 0.03291603 |
| AlphaPow82 | Sum_PC | 0.2238808 | 0.77611921 | 0.5 | 0.2884619 | 0.7960072152 | -0.00479183571 | 0.02321210 |
| AlphaPow83 | Blocktapping | 0.2106641 | 0.78933585 | 0.5 | 0.2668878 | 0.4526899004 | 0.01877569689 | 0.02582703 |
| AlphaPow83 | Sum_PC | 0.1865121 | 0.81348791 | 0.5 | 0.2292746 | 0.7841588593 | 0.00588364993 | 0.02036866 |
| AlphaPow84 | Blocktapping | 0.1765587 | 0.82344134 | 0.5 | 0.2144156 | 0.6061719388 | 0.00980080928 | 0.02205695 |
| AlphaPow84 | Sum_PC | 0.1849860 | 0.81501403 | 0.5 | 0.2269727 | 0.6387509213 | -0.00952869955 | 0.02120594 |
| AlphaPow85 | Blocktapping | 0.1817099 | 0.81829007 | 0.5 | 0.2220605 | 0.6866145526 | 0.00883666537 | 0.02262114 |
| AlphaPow85 | Sum_PC | 0.1793052 | 0.82069479 | 0.5 | 0.2184798 | 0.7460764088 | -0.00705208599 | 0.02067282 |
| AlphaPow86 | Blocktapping | 0.2645792 | 0.73542080 | 0.5 | 0.3597657 | 0.2641475776 | 0.03279023972 | 0.03258790 |
| AlphaPow86 | Sum_PC | 0.2032554 | 0.79674465 | 0.5 | 0.2551073 | 0.8064989712 | 0.00523763544 | 0.02105913 |
| AlphaPow87 | Blocktapping | 0.4036045 | 0.59639552 | 0.5 | 0.6767396 | 0.1073973660 | 0.06749341322 | 0.04595474 |
| AlphaPow87 | Sum_PC | 0.2639734 | 0.73602658 | 0.5 | 0.3586466 | 0.6279727460 | -0.00999037057 | 0.02322725 |
| AlphaPow88 | Blocktapping | 0.1997202 | 0.80027984 | 0.5 | 0.2495629 | 0.5091848819 | 0.01582292739 | 0.02429698 |
| AlphaPow88 | Sum_PC | 0.1820821 | 0.81791788 | 0.5 | 0.2226166 | 0.8934769145 | 0.00289239848 | 0.01975296 |
| AlphaPow89 | Blocktapping | 0.3526189 | 0.64738109 | 0.5 | 0.5446852 | 0.1658105549 | 0.04790678740 | 0.03822234 |
| AlphaPow89 | Sum_PC | 0.3026032 | 0.69739684 | 0.5 | 0.4339038 | 0.2977998889 | -0.02655630740 | 0.02927257 |
| AlphaPow9 | Blocktapping | 0.3296353 | 0.67036467 | 0.5 | 0.4917254 | 0.1829634485 | 0.04540268945 | 0.03787729 |
| AlphaPow9 | Sum_PC | 0.2410008 | 0.75899923 | 0.5 | 0.3175244 | 0.5738127697 | -0.01129763944 | 0.02229114 |
| AlphaPow90 | Blocktapping | 0.2440589 | 0.75594113 | 0.5 | 0.3228543 | 0.3555803978 | 0.02366139696 | 0.02819960 |
| AlphaPow90 | Sum_PC | 0.2210795 | 0.77892050 | 0.5 | 0.2838281 | 0.5248634719 | -0.01332241255 | 0.02343997 |
| AlphaPow91 | Blocktapping | 0.1868595 | 0.81314055 | 0.5 | 0.2297997 | 0.7042395196 | 0.00704569002 | 0.02287564 |
| AlphaPow91 | Sum_PC | 0.1824912 | 0.81750882 | 0.5 | 0.2232284 | 0.8806559507 | 0.00280449264 | 0.01972496 |
| AlphaPow92 | Blocktapping | 0.1994050 | 0.80059503 | 0.5 | 0.2490710 | 0.5269733564 | 0.01496243132 | 0.02447599 |
| AlphaPow92 | Sum_PC | 0.1813728 | 0.81862721 | 0.5 | 0.2215572 | 0.8568332530 | 0.00365569703 | 0.02015364 |
| AlphaPow93 | Blocktapping | 0.1966270 | 0.80337298 | 0.5 | 0.2447518 | 0.5664948770 | 0.01152903416 | 0.02480235 |
| AlphaPow93 | Sum_PC | 0.1797059 | 0.82029413 | 0.5 | 0.2190749 | 0.9312961466 | -0.00104889048 | 0.01922756 |
| AlphaPow94 | Blocktapping | 0.2234515 | 0.77654853 | 0.5 | 0.2877495 | 0.3785688266 | 0.02168525659 | 0.02739786 |
| AlphaPow94 | Sum_PC | 0.1847143 | 0.81528568 | 0.5 | 0.2265639 | 0.9334392810 | 0.00240522533 | 0.02007881 |
| AlphaPow95 | Blocktapping | 0.2463933 | 0.75360667 | 0.5 | 0.3269522 | 0.3623301244 | 0.02349739852 | 0.02819644 |
| AlphaPow95 | Sum_PC | 0.2329252 | 0.76707479 | 0.5 | 0.3036538 | 0.4613608318 | -0.01508560227 | 0.02465243 |

| DV | Predictor | Sum_PostModProb_Incl | Sum_PostModProb_Excl | Sum_PriorModProb_Incl | BF_Incl | Frequentist_p_value | postMean | postStd |
|---|---|---|---|---|---|---|---|---|
| AlphaPow96 | Blocktapping | 0.1951112 | 0.80488881 | 0.5 | 0.2424076 | 0.4900433266 | 0.01589420166 | 0.02467828 |
| AlphaPow96 | Sum_PC | 0.1764369 | 0.82356313 | 0.5 | 0.2142360 | 0.7903132457 | -0.00449646163 | 0.01939991 |
| AlphaPow97 | Blocktapping | 0.1722925 | 0.82770751 | 0.5 | 0.2081562 | 0.7601127569 | 0.00619720288 | 0.02216979 |
| AlphaPow97 | Sum_PC | 0.1688954 | 0.83110458 | 0.5 | 0.2032180 | 0.8101709073 | -0.00393115227 | 0.01943108 |
| AlphaPow98 | Blocktapping | 0.2159904 | 0.78400960 | 0.5 | 0.2754946 | 0.3894423020 | 0.02218565591 | 0.02770217 |
| AlphaPow98 | Sum_PC | 0.1810002 | 0.81899979 | 0.5 | 0.2210015 | 0.8532667016 | 0.00371761665 | 0.01990813 |
| AlphaPow99 | Blocktapping | 0.3588920 | 0.64110805 | 0.5 | 0.5597995 | 0.1600907687 | 0.04880834527 | 0.04074738 |
| AlphaPow99 | Sum_PC | 0.2957018 | 0.70429822 | 0.5 | 0.4198531 | 0.3418709711 | -0.02397575425 | 0.02728672 |
| beta_ROI1_AECnodestrength | Blocktapping | 0.3042849 | 0.69571515 | 0.5 | 0.4373699 | 0.3032402154 | -0.02837309505 | 0.03108861 |
| beta_ROI1_AECnodestrength | Sum_PC | 0.3755201 | 0.62447987 | 0.5 | 0.6013326 | 0.1360246689 | 0.05099058355 | 0.03855493 |
| beta_ROI10_AECnodestrength | Blocktapping | 0.3409171 | 0.65908286 | 0.5 | 0.5172599 | 0.9768923711 | 0.00002950200 | 0.02304910 |
| beta_ROI10_AECnodestrength | Sum_PC | 0.8334951 | 0.16650486 | 0.5 | 5.0058305 | 0.0077879544 | 0.19081921814 | 0.07871861 |
| beta_ROI100_AECnodestrength | Blocktapping | 0.2268859 | 0.77311406 | 0.5 | 0.2934702 | 0.9710953947 | 0.00001638448 | 0.02325830 |
| beta_ROI100_AECnodestrength | Sum_PC | 0.3458951 | 0.65410491 | 0.5 | 0.5288068 | 0.1562702203 | 0.04980765915 | 0.03696129 |
| beta_ROI11_AECnodestrength | Blocktapping | 0.2535926 | 0.74640744 | 0.5 | 0.3397508 | 0.6740305957 | -0.00869983880 | 0.02432762 |
| beta_ROI11_AECnodestrength | Sum_PC | 0.4189375 | 0.58106253 | 0.5 | 0.7209852 | 0.0951330682 | 0.06738266907 | 0.04379281 |
| beta_ROI12_AECnodestrength | Blocktapping | 0.3351094 | 0.66489056 | 0.5 | 0.5040069 | 0.6347517785 | -0.01014476218 | 0.02458758 |
| beta_ROI12_AECnodestrength | Sum_PC | 0.7170626 | 0.28293744 | 0.5 | 2.5343502 | 0.0176788076 | 0.15147580040 | 0.06940521 |
| beta_ROI13_AECnodestrength | Blocktapping | 0.2339102 | 0.76608977 | 0.5 | 0.3053301 | 0.5882177797 | -0.01158022189 | 0.02423713 |
| beta_ROI13_AECnodestrength | Sum_PC | 0.3111960 | 0.68880395 | 0.5 | 0.4517919 | 0.1946675925 | 0.03773213168 | 0.03422021 |
| beta_ROI14_AECnodestrength | Blocktapping | 0.1982827 | 0.80171732 | 0.5 | 0.2473224 | 0.8916620978 | -0.00301003459 | 0.02335983 |
| beta_ROI14_AECnodestrength | Sum_PC | 0.2380214 | 0.76197860 | 0.5 | 0.3123728 | 0.3576127220 | 0.02191272969 | 0.02623798 |
| beta_ROI15_AECnodestrength | Blocktapping | 0.2792582 | 0.72074179 | 0.5 | 0.3874594 | 0.3612738626 | 0.02591965747 | 0.02932238 |
| beta_ROI15_AECnodestrength | Sum_PC | 0.3274721 | 0.67252789 | 0.5 | 0.4869272 | 0.2050049033 | 0.04031123708 | 0.03375410 |
| beta_ROI16_AECnodestrength | Blocktapping | 0.1890361 | 0.81096389 | 0.5 | 0.2331005 | 0.7582240678 | -0.00672432382 | 0.02281639 |
| beta_ROI16_AECnodestrength | Sum_PC | 0.2284972 | 0.77150280 | 0.5 | 0.2961716 | 0.3461705887 | 0.02276915938 | 0.02514149 |
| beta_ROI17_AECnodestrength | Blocktapping | 0.2090087 | 0.79099125 | 0.5 | 0.2642365 | 0.6549500986 | 0.01026038590 | 0.02460152 |
| beta_ROI17_AECnodestrength | Sum_PC | 0.2426000 | 0.75739996 | 0.5 | 0.3203064 | 0.3594501837 | 0.02320887225 | 0.02694557 |
| beta_ROI18_AECnodestrength | Blocktapping | 0.1955675 | 0.80443251 | 0.5 | 0.2431124 | 0.7942366739 | -0.00522250311 | 0.02379086 |
| beta_ROI18_AECnodestrength | Sum_PC | 0.2199896 | 0.78001037 | 0.5 | 0.2820342 | 0.4701244685 | 0.01738324563 | 0.02544933 |
| beta_ROI19_AECnodestrength | Blocktapping | 0.1899982 | 0.81000181 | 0.5 | 0.2345651 | 0.9163866315 | 0.00194101391 | 0.02323764 |

| DV | Predictor | Sum_PostModProb_Incl | Sum_PostModProb_Excl | Sum_PriorModProb_Incl | BF_Incl | Frequentist_p_value | postMean | postStd |
|---|---|---|---|---|---|---|---|---|
| beta_ROI19_AECnodestrength | Sum_PC | 0.2096561 | 0.79034389 | 0.5 | 0.2652720 | 0.5030927915 | 0.01405778026 | 0.02346449 |
| beta_ROI2_AECnodestrength | Blocktapping | 0.2346597 | 0.76534033 | 0.5 | 0.3066083 | 0.3668076556 | -0.02299609890 | 0.02908614 |
| beta_ROI2_AECnodestrength | Sum_PC | 0.1969054 | 0.80309465 | 0.5 | 0.2451832 | 0.7400936199 | 0.00619195208 | 0.02094175 |
| beta_ROI20_AECnodestrength | Blocktapping | 0.1989945 | 0.80100551 | 0.5 | 0.2484309 | 0.8308763890 | -0.00506997668 | 0.02271595 |
| beta_ROI20_AECnodestrength | Sum_PC | 0.2371065 | 0.76289355 | 0.5 | 0.3107989 | 0.3574428757 | 0.02249684899 | 0.02650813 |
| beta_ROI21_AECnodestrength | Blocktapping | 0.1888376 | 0.81116236 | 0.5 | 0.2327988 | 0.9701911582 | 0.00032836921 | 0.02370240 |
| beta_ROI21_AECnodestrength | Sum_PC | 0.2089137 | 0.79108628 | 0.5 | 0.2640846 | 0.5144690604 | 0.01488658209 | 0.02354265 |
| beta_ROI22_AECnodestrength | Blocktapping | 0.2451664 | 0.75483356 | 0.5 | 0.3247954 | 0.6728948639 | -0.00757941059 | 0.02462455 |
| beta_ROI22_AECnodestrength | Sum_PC | 0.3816012 | 0.61839884 | 0.5 | 0.6170794 | 0.1208956719 | 0.05642910987 | 0.04109132 |
| beta_ROI23_AECnodestrength | Blocktapping | 0.2519807 | 0.74801933 | 0.5 | 0.3368639 | 0.4981648739 | -0.01568398724 | 0.02695072 |
| beta_ROI23_AECnodestrength | Sum_PC | 0.3265764 | 0.67342355 | 0.5 | 0.4849495 | 0.1896987802 | 0.04178193210 | 0.03577236 |
| beta_ROI24_AECnodestrength | Blocktapping | 0.3190690 | 0.68093100 | 0.5 | 0.4685776 | 0.3499955594 | 0.02625031285 | 0.03006929 |
| beta_ROI24_AECnodestrength | Sum_PC | 0.4441195 | 0.55588045 | 0.5 | 0.7989479 | 0.1020943850 | 0.06756338475 | 0.04462202 |
| beta_ROI25_AECnodestrength | Blocktapping | 0.3758636 | 0.62413639 | 0.5 | 0.6022139 | 0.3658538929 | -0.02306210897 | 0.02942342 |
| beta_ROI25_AECnodestrength | Sum_PC | 0.6690434 | 0.33095658 | 0.5 | 2.0215444 | 0.0259884744 | 0.13312608529 | 0.06635641 |
| beta_ROI26_AECnodestrength | Blocktapping | 0.3316247 | 0.66837531 | 0.5 | 0.4961654 | 0.5507086530 | -0.01409939964 | 0.02560783 |
| beta_ROI26_AECnodestrength | Sum_PC | 0.6361907 | 0.36380925 | 0.5 | 1.7486931 | 0.0296024997 | 0.12655175544 | 0.06365675 |
| beta_ROI27_AECnodestrength | Blocktapping | 0.3936345 | 0.60636555 | 0.5 | 0.6491702 | 0.4422755454 | 0.02019930225 | 0.02816358 |
| beta_ROI27_AECnodestrength | Sum_PC | 0.7769464 | 0.22305360 | 0.5 | 3.4832273 | 0.0150117300 | 0.16703872073 | 0.07313399 |
| beta_ROI28_AECnodestrength | Blocktapping | 0.3310658 | 0.66893415 | 0.5 | 0.4949154 | 0.8975984008 | -0.00298576751 | 0.02362437 |
| beta_ROI28_AECnodestrength | Sum_PC | 0.7566773 | 0.24332274 | 0.5 | 3.1097679 | 0.0140560301 | 0.16521581234 | 0.07184318 |
| beta_ROI29_AECnodestrength | Blocktapping | 0.2119325 | 0.78806750 | 0.5 | 0.2689268 | 0.5406550360 | 0.01422236247 | 0.02604439 |
| beta_ROI29_AECnodestrength | Sum_PC | 0.2064987 | 0.79350129 | 0.5 | 0.2602374 | 0.6188816253 | 0.01070355807 | 0.02245718 |
| beta_ROI3_AECnodestrength | Blocktapping | 0.2562192 | 0.74378083 | 0.5 | 0.3444821 | 0.3190951010 | -0.02546081722 | 0.03071617 |
| beta_ROI3_AECnodestrength | Sum_PC | 0.2361723 | 0.76382769 | 0.5 | 0.3091958 | 0.4118389684 | 0.01727610522 | 0.02329257 |
| beta_ROI30_AECnodestrength | Blocktapping | 0.2461984 | 0.75380155 | 0.5 | 0.3266091 | 0.7041707180 | -0.00755637010 | 0.02419367 |
| beta_ROI30_AECnodestrength | Sum_PC | 0.3768574 | 0.62314257 | 0.5 | 0.6047692 | 0.1278322591 | 0.05420488443 | 0.03937125 |
| beta_ROI31_AECnodestrength | Blocktapping | 0.2988015 | 0.70119854 | 0.5 | 0.4261296 | 0.8405350668 | -0.00411451994 | 0.02360488 |
| beta_ROI31_AECnodestrength | Sum_PC | 0.6090978 | 0.39090217 | 0.5 | 1.5581848 | 0.0326763884 | 0.11878019126 | 0.06046703 |

| DV | Predictor | Sum_PostModProb_Incl | Sum_PostModProb_Excl | Sum_PriorModProb_Incl | BF_Incl | Frequentist_p_value | postMean | postStd |
|---|---|---|---|---|---|---|---|---|
| beta_ROI32_AECnodestrength | Blocktapping | 0.3261914 | 0.67380859 | 0.5 | 0.4841010 | 0.3723005604 | -0.02447146111 | 0.02953346 |
| beta_ROI32_AECnodestrength | Sum_PC | 0.4912151 | 0.50878486 | 0.5 | 0.9654673 | 0.0694149537 | 0.08224474367 | 0.05004538 |
| beta_ROI33_AECnodestrength | Blocktapping | 0.2705388 | 0.72946120 | 0.5 | 0.3708748 | 0.9703818134 | 0.00038547878 | 0.02196693 |
| beta_ROI33_AECnodestrength | Sum_PC | 0.5530460 | 0.44695396 | 0.5 | 1.2373669 | 0.0454475451 | 0.10470656383 | 0.05916261 |
| beta_ROI34_AECnodestrength | Blocktapping | 0.1481875 | 0.85181255 | 0.5 | 0.1739672 | 0.9998885679 | 0.00019757105 | 0.01885437 |
| beta_ROI34_AECnodestrength | Sum_PC | 0.1722671 | 0.82773289 | 0.5 | 0.2081192 | 0.4362036666 | 0.01510115358 | 0.02056772 |
| beta_ROI35_AECnodestrength | Blocktapping | 0.3668500 | 0.63315001 | 0.5 | 0.5794045 | 0.7520717551 | 0.00647664070 | 0.02265804 |
| beta_ROI35_AECnodestrength | Sum_PC | 0.9092855 | 0.09071452 | 0.5 | 10.0235933 | 0.0035721215 | 0.23137508837 | 0.08352159 |
| beta_ROI36_AECnodestrength | Blocktapping | 0.2103788 | 0.78962121 | 0.5 | 0.2664300 | 0.6526472673 | -0.00957792311 | 0.02508669 |
| beta_ROI36_AECnodestrength | Sum_PC | 0.2350102 | 0.76498977 | 0.5 | 0.3072070 | 0.3941439531 | 0.02009424979 | 0.02552000 |
| beta_ROI37_AECnodestrength | Blocktapping | 0.1953160 | 0.80468403 | 0.5 | 0.2427238 | 0.9130327881 | 0.00242193373 | 0.02175759 |
| beta_ROI37_AECnodestrength | Sum_PC | 0.2549269 | 0.74507307 | 0.5 | 0.3421502 | 0.2895084132 | 0.02859075966 | 0.02821294 |
| beta_ROI38_AECnodestrength | Blocktapping | 0.3220536 | 0.67794643 | 0.5 | 0.4750428 | 0.8146743994 | -0.00430897566 | 0.02345546 |
| beta_ROI38_AECnodestrength | Sum_PC | 0.7416376 | 0.25836243 | 0.5 | 2.8705318 | 0.0151628733 | 0.16005844412 | 0.07340314 |
| beta_ROI39_AECnodestrength | Blocktapping | 0.3364907 | 0.66350929 | 0.5 | 0.5071379 | 0.7118750605 | 0.00821089229 | 0.02325274 |
| beta_ROI39_AECnodestrength | Sum_PC | 0.7760461 | 0.22395391 | 0.5 | 3.4652045 | 0.0125663962 | 0.17388741042 | 0.07401769 |
| beta_ROI4_AECnodestrength | Blocktapping | 0.2047830 | 0.79521700 | 0.5 | 0.2575184 | 0.5441785389 | -0.01480683948 | 0.02503274 |
| beta_ROI4_AECnodestrength | Sum_PC | 0.1862384 | 0.81376161 | 0.5 | 0.2288611 | 0.9002338928 | 0.00203823725 | 0.02052624 |
| beta_ROI40_AECnodestrength | Blocktapping | 0.2040897 | 0.79591032 | 0.5 | 0.2564230 | 0.9765896037 | 0.00124039591 | 0.02162126 |
| beta_ROI40_AECnodestrength | Sum_PC | 0.2895547 | 0.71044531 | 0.5 | 0.4075679 | 0.2074914108 | 0.03802685824 | 0.03225717 |
| beta_ROI41_AECnodestrength | Blocktapping | 0.2400624 | 0.75993760 | 0.5 | 0.3158975 | 0.8771562444 | 0.00428264391 | 0.02263519 |
| beta_ROI41_AECnodestrength | Sum_PC | 0.4010447 | 0.59895528 | 0.5 | 0.6695737 | 0.1092636194 | 0.06394600937 | 0.04292812 |
| beta_ROI42_AECnodestrength | Blocktapping | 0.3387353 | 0.66126474 | 0.5 | 0.5122536 | 0.8969268167 | 0.00170497108 | 0.02261053 |
| beta_ROI42_AECnodestrength | Sum_PC | 0.8228916 | 0.17710839 | 0.5 | 4.6462597 | 0.0084364183 | 0.19161497300 | 0.07756835 |
| beta_ROI43_AECnodestrength | Blocktapping | 0.3837372 | 0.61626283 | 0.5 | 0.6226843 | 0.4870501059 | 0.01741994051 | 0.02551811 |
| beta_ROI43_AECnodestrength | Sum_PC | 0.8483676 | 0.15163238 | 0.5 | 5.5948974 | 0.0075974644 | 0.19758146326 | 0.08029769 |
| beta_ROI44_AECnodestrength | Blocktapping | 0.3456175 | 0.65438252 | 0.5 | 0.5281582 | 0.6651392224 | -0.00914203263 | 0.02598273 |
| beta_ROI44_AECnodestrength | Sum_PC | 0.7067859 | 0.29321414 | 0.5 | 2.4104768 | 0.0193580837 | 0.14386344569 | 0.06856352 |
| beta_ROI45_AECnodestrength | Blocktapping | 0.4048892 | 0.59511079 | 0.5 | 0.6803594 | 0.5565361347 | -0.01371573985 | 0.02363018 |

| DV | Predictor | Sum_PostModProb_Incl | Sum_PostModProb_Excl | Sum_PriorModProb_Incl | BF_Incl | Frequentist_p_value | postMean | postStd |
|---|---|---|---|---|---|---|---|---|
| beta_ROI45_AECnodestrength | Sum_PC | 0.9879151 | 0.01208490 | 0.5 | 81.7478738 | 0.0002393285 | 0.30078635060 | 0.08581494 |
| beta_ROI46_AECnodestrength | Blocktapping | 0.3333916 | 0.66660843 | 0.5 | 0.5001311 | 0.5924469346 | 0.01315409202 | 0.02485183 |
| beta_ROI46_AECnodestrength | Sum_PC | 0.6727525 | 0.32724750 | 0.5 | 2.0557911 | 0.0258891546 | 0.13385372490 | 0.06549635 |
| beta_ROI47_AECnodestrength | Blocktapping | 0.2836227 | 0.71637729 | 0.5 | 0.3959125 | 0.8054928028 | 0.00560672420 | 0.02291006 |
| beta_ROI47_AECnodestrength | Sum_PC | 0.5589773 | 0.44102270 | 0.5 | 1.2674570 | 0.0469017272 | 0.10372313499 | 0.05561641 |
| beta_ROI48_AECnodestrength | Blocktapping | 0.2641729 | 0.73582712 | 0.5 | 0.3590149 | 0.8546861890 | 0.00501369839 | 0.02316259 |
| beta_ROI48_AECnodestrength | Sum_PC | 0.4929058 | 0.50709420 | 0.5 | 0.9720202 | 0.0726179974 | 0.08464402374 | 0.05077077 |
| beta_ROI49_AECnodestrength | Blocktapping | 0.2431406 | 0.75685936 | 0.5 | 0.3212494 | 0.3606663712 | -0.02372628159 | 0.02909214 |
| beta_ROI49_AECnodestrength | Sum_PC | 0.2319747 | 0.76802531 | 0.5 | 0.3020404 | 0.4442772620 | 0.01740039871 | 0.02429427 |
| beta_ROI5_AECnodestrength | Blocktapping | 0.2320983 | 0.76790168 | 0.5 | 0.3022500 | 0.4077134831 | -0.02086991323 | 0.02837415 |
| beta_ROI5_AECnodestrength | Sum_PC | 0.2087769 | 0.79122314 | 0.5 | 0.2638660 | 0.6161826915 | 0.00923908224 | 0.02240325 |
| beta_ROI50_AECnodestrength | Blocktapping | 0.1943352 | 0.80566477 | 0.5 | 0.2412110 | 0.9531299601 | -0.00119335316 | 0.02260668 |
| beta_ROI50_AECnodestrength | Sum_PC | 0.2354052 | 0.76459481 | 0.5 | 0.3078823 | 0.3585333958 | 0.02204324568 | 0.02698649 |
| beta_ROI51_AECnodestrength | Blocktapping | 0.2262320 | 0.77376801 | 0.5 | 0.2923770 | 0.5010926814 | -0.01466395011 | 0.02591864 |
| beta_ROI51_AECnodestrength | Sum_PC | 0.2506676 | 0.74933239 | 0.5 | 0.3345212 | 0.3500487712 | 0.02335346400 | 0.02750402 |
| beta_ROI52_AECnodestrength | Blocktapping | 0.2373749 | 0.76262506 | 0.5 | 0.3112603 | 0.4729132687 | -0.01717630016 | 0.02538519 |
| beta_ROI52_AECnodestrength | Sum_PC | 0.2845629 | 0.71543709 | 0.5 | 0.3977469 | 0.2849010800 | 0.02829933639 | 0.03095408 |
| beta_ROI53_AECnodestrength | Blocktapping | 0.2330984 | 0.76690159 | 0.5 | 0.3039483 | 0.8707152333 | -0.00349757173 | 0.02324913 |
| beta_ROI53_AECnodestrength | Sum_PC | 0.3615302 | 0.63846976 | 0.5 | 0.5662449 | 0.1370316394 | 0.05340917513 | 0.03753140 |
| beta_ROI54_AECnodestrength | Blocktapping | 0.2727735 | 0.72722648 | 0.5 | 0.3750874 | 0.7487709952 | -0.00623080854 | 0.02417337 |
| beta_ROI54_AECnodestrength | Sum_PC | 0.4956832 | 0.50431682 | 0.5 | 0.9828805 | 0.0651879574 | 0.08855388681 | 0.05200143 |
| beta_ROI55_AECnodestrength | Blocktapping | 0.3245528 | 0.67544716 | 0.5 | 0.4805007 | 0.3119442066 | -0.02906773742 | 0.03103607 |
| beta_ROI55_AECnodestrength | Sum_PC | 0.4043875 | 0.59561253 | 0.5 | 0.6789439 | 0.1286488732 | 0.05474162401 | 0.04074201 |
| beta_ROI56_AECnodestrength | Blocktapping | 0.2962839 | 0.70371611 | 0.5 | 0.4210276 | 0.3165837361 | -0.02760471951 | 0.03181126 |
| beta_ROI56_AECnodestrength | Sum_PC | 0.3526118 | 0.64738818 | 0.5 | 0.5446683 | 0.1725745051 | 0.04352408491 | 0.03538187 |
| beta_ROI57_AECnodestrength | Blocktapping | 0.2110753 | 0.78892471 | 0.5 | 0.2675481 | 0.6022211274 | -0.01114192497 | 0.02542156 |
| beta_ROI57_AECnodestrength | Sum_PC | 0.2287456 | 0.77125442 | 0.5 | 0.2965890 | 0.4528688777 | 0.01917566439 | 0.02498187 |
| beta_ROI58_AECnodestrength | Blocktapping | 0.2437259 | 0.75627412 | 0.5 | 0.3222719 | 0.3550018079 | -0.02261915800 | 0.02921431 |
| beta_ROI58_AECnodestrength | Sum_PC | 0.2459824 | 0.75401760 | 0.5 | 0.3262290 | 0.4169377827 | 0.01920185232 | 0.02689728 |

| DV | Predictor | Sum_PostModProb_Incl | Sum_PostModProb_Excl | Sum_PriorModProb_Incl | BF_Incl | Frequentist_p_value | postMean | postStd |
|---|---|---|---|---|---|---|---|---|
| beta_ROI59_AECnodestrength | Blocktapping | 0.2758028 | 0.72419725 | 0.5 | 0.3808393 | 0.6660676463 | 0.01090192584 | 0.02519572 |
| beta_ROI59_AECnodestrength | Sum_PC | 0.4591057 | 0.54089432 | 0.5 | 0.8487900 | 0.0853927823 | 0.07383471293 | 0.04804160 |
| beta_ROI6_AECnodestrength | Blocktapping | 0.2642012 | 0.73579876 | 0.5 | 0.3590673 | 0.3111240560 | -0.02830244552 | 0.03093384 |
| beta_ROI6_AECnodestrength | Sum_PC | 0.2430999 | 0.75690013 | 0.5 | 0.3211783 | 0.4152399578 | 0.01905636944 | 0.02497058 |
| beta_ROI60_AECnodestrength | Blocktapping | 0.3286445 | 0.67135554 | 0.5 | 0.4895237 | 0.6648604965 | 0.01069353126 | 0.02427365 |
| beta_ROI60_AECnodestrength | Sum_PC | 0.6753625 | 0.32463752 | 0.5 | 2.0803586 | 0.0254965539 | 0.13640430447 | 0.06424587 |
| beta_ROI61_AECnodestrength | Blocktapping | 0.2905879 | 0.70941206 | 0.5 | 0.4096180 | 0.7159007254 | 0.00840832054 | 0.02450464 |
| beta_ROI61_AECnodestrength | Sum_PC | 0.5536760 | 0.44632400 | 0.5 | 1.2405248 | 0.0505506842 | 0.10091064286 | 0.05572052 |
| beta_ROI62_AECnodestrength | Blocktapping | 0.3078836 | 0.69211641 | 0.5 | 0.4448436 | 0.6504383556 | 0.01053452166 | 0.02538917 |
| beta_ROI62_AECnodestrength | Sum_PC | 0.5795398 | 0.42046017 | 0.5 | 1.3783466 | 0.0440975244 | 0.10666041578 | 0.05774152 |
| beta_ROI63_AECnodestrength | Blocktapping | 0.3447838 | 0.65521622 | 0.5 | 0.5262137 | 0.5044639619 | 0.01646946265 | 0.02540758 |
| beta_ROI63_AECnodestrength | Sum_PC | 0.6455336 | 0.35446643 | 0.5 | 1.8211416 | 0.0307805257 | 0.12212477719 | 0.06043479 |
| beta_ROI64_AECnodestrength | Blocktapping | 0.2735360 | 0.72646398 | 0.5 | 0.3765307 | 0.6114357960 | 0.01141281402 | 0.02487962 |
| beta_ROI64_AECnodestrength | Sum_PC | 0.4495298 | 0.55047024 | 0.5 | 0.8166286 | 0.0859571432 | 0.07257141582 | 0.04555657 |
| beta_ROI65_AECnodestrength | Blocktapping | 0.2557005 | 0.74429946 | 0.5 | 0.3435452 | 0.4532498543 | 0.01891642788 | 0.02641875 |
| beta_ROI65_AECnodestrength | Sum_PC | 0.3218825 | 0.67811752 | 0.5 | 0.4746706 | 0.2026786168 | 0.04010500462 | 0.03278321 |
| beta_ROI66_AECnodestrength | Blocktapping | 0.3530820 | 0.64691803 | 0.5 | 0.5457909 | 0.2516215676 | 0.03774414120 | 0.03535857 |
| beta_ROI66_AECnodestrength | Sum_PC | 0.4290455 | 0.57095451 | 0.5 | 0.7514530 | 0.1234197309 | 0.06056861371 | 0.04259701 |
| beta_ROI67_AECnodestrength | Blocktapping | 0.2443058 | 0.75569421 | 0.5 | 0.3232866 | 0.7022456508 | -0.00666528334 | 0.02501541 |
| beta_ROI67_AECnodestrength | Sum_PC | 0.3662173 | 0.63378266 | 0.5 | 0.5778280 | 0.1338599417 | 0.05411825524 | 0.03944566 |
| beta_ROI68_AECnodestrength | Blocktapping | 0.4323187 | 0.56768134 | 0.5 | 0.7615517 | 0.2269607830 | 0.03861137795 | 0.03449046 |
| beta_ROI68_AECnodestrength | Sum_PC | 0.6354879 | 0.36451207 | 0.5 | 1.7433934 | 0.0403737553 | 0.10632704516 | 0.05615558 |
| beta_ROI69_AECnodestrength | Blocktapping | 0.2513983 | 0.74860166 | 0.5 | 0.3358239 | 0.4441574478 | 0.02018214990 | 0.02805435 |
| beta_ROI69_AECnodestrength | Sum_PC | 0.2726353 | 0.72736473 | 0.5 | 0.3748261 | 0.3091743584 | 0.02633950140 | 0.02775720 |
| beta_ROI7_AECnodestrength | Blocktapping | 0.1714919 | 0.82850805 | 0.5 | 0.2069889 | 0.8873346961 | 0.00239297064 | 0.02167809 |
| beta_ROI7_AECnodestrength | Sum_PC | 0.1773079 | 0.82269212 | 0.5 | 0.2155216 | 0.8583699283 | 0.00321335923 | 0.02043914 |
| beta_ROI70_AECnodestrength | Blocktapping | 0.1631037 | 0.83689630 | 0.5 | 0.1948912 | 0.8661140029 | 0.00297715105 | 0.01731539 |
| beta_ROI70_AECnodestrength | Sum_PC | 0.3493310 | 0.65066897 | 0.5 | 0.5368798 | 0.3830914777 | 0.03386449246 | 0.03950792 |
| beta_ROI71_AECnodestrength | Blocktapping | 0.2413086 | 0.75869135 | 0.5 | 0.3180590 | 0.5671851701 | 0.01370728376 | 0.02587144 |

| DV | Predictor | Sum_PostModProb_Incl | Sum_PostModProb_Excl | Sum_PriorModProb_Incl | BF_Incl | Frequentist_p_value | postMean | postStd |
|---|---|---|---|---|---|---|---|---|
| beta_ROI71_AECnodestrength | Sum_PC | 0.3132095 | 0.68679046 | 0.5 | 0.4560482 | 0.2008484241 | 0.04011005618 | 0.03260665 |
| beta_ROI72_AECnodestrength | Blocktapping | 0.3524593 | 0.64754069 | 0.5 | 0.5443045 | 0.7539108842 | 0.00741287865 | 0.02213393 |
| beta_ROI72_AECnodestrength | Sum_PC | 0.8523857 | 0.14761431 | 0.5 | 5.7744108 | 0.0066565535 | 0.19833165376 | 0.07907205 |
| beta_ROI73_AECnodestrength | Blocktapping | 0.3502995 | 0.64970053 | 0.5 | 0.5391707 | 0.7012002524 | 0.00824784778 | 0.02418754 |
| beta_ROI73_AECnodestrength | Sum_PC | 0.7952339 | 0.20476608 | 0.5 | 3.8836215 | 0.0111814466 | 0.17956147195 | 0.07756755 |
| beta_ROI74_AECnodestrength | Blocktapping | 0.2325597 | 0.76744025 | 0.5 | 0.3030330 | 0.9649955294 | 0.00064033655 | 0.02152843 |
| beta_ROI74_AECnodestrength | Sum_PC | 0.4013285 | 0.59867151 | 0.5 | 0.6703651 | 0.0982573198 | 0.06505034911 | 0.04397533 |
| beta_ROI75_AECnodestrength | Blocktapping | 0.2393023 | 0.76069772 | 0.5 | 0.3145826 | 0.8602390473 | 0.00377823976 | 0.02301297 |
| beta_ROI75_AECnodestrength | Sum_PC | 0.3897577 | 0.61024226 | 0.5 | 0.6386934 | 0.1104968239 | 0.05891723961 | 0.04164207 |
| beta_ROI76_AECnodestrength | Blocktapping | 0.3217606 | 0.67823936 | 0.5 | 0.4744057 | 0.8930365252 | -0.00346217684 | 0.02186691 |
| beta_ROI76_AECnodestrength | Sum_PC | 0.7934381 | 0.20656192 | 0.5 | 3.8411634 | 0.0128667807 | 0.17282688230 | 0.07500975 |
| beta_ROI77_AECnodestrength | Blocktapping | 0.2402811 | 0.75971886 | 0.5 | 0.3162764 | 0.7078591601 | 0.00856107484 | 0.02370137 |
| beta_ROI77_AECnodestrength | Sum_PC | 0.3656477 | 0.63435234 | 0.5 | 0.5764110 | 0.1406051928 | 0.05188222938 | 0.03915880 |
| beta_ROI78_AECnodestrength | Blocktapping | 0.2996074 | 0.70039261 | 0.5 | 0.4277706 | 0.9144908563 | -0.00232617102 | 0.02349828 |
| beta_ROI78_AECnodestrength | Sum_PC | 0.6268055 | 0.37319454 | 0.5 | 1.6795676 | 0.0309310468 | 0.12306211285 | 0.06072413 |
| beta_ROI79_AECnodestrength | Blocktapping | 0.2533141 | 0.74668591 | 0.5 | 0.3392512 | 0.9968906252 | -0.00043136993 | 0.02357371 |
| beta_ROI79_AECnodestrength | Sum_PC | 0.4511315 | 0.54886846 | 0.5 | 0.8219302 | 0.0819588259 | 0.07390258854 | 0.04671806 |
| beta_ROI8_AECnodestrength | Blocktapping | 0.1837694 | 0.81623058 | 0.5 | 0.2251440 | 0.8480176361 | -0.00422572744 | 0.02330404 |
| beta_ROI8_AECnodestrength | Sum_PC | 0.1934214 | 0.80657859 | 0.5 | 0.2398048 | 0.5885303038 | -0.01098025524 | 0.02193882 |
| beta_ROI80_AECnodestrength | Blocktapping | 0.2688762 | 0.73112383 | 0.5 | 0.3677574 | 0.9829508486 | -0.00047495091 | 0.02251187 |
| beta_ROI80_AECnodestrength | Sum_PC | 0.5271161 | 0.47288394 | 0.5 | 1.1146838 | 0.0553091494 | 0.09696305707 | 0.05440230 |
| beta_ROI81_AECnodestrength | Blocktapping | 0.2406278 | 0.75937224 | 0.5 | 0.3168772 | 0.5121787626 | 0.01546741958 | 0.02586229 |
| beta_ROI81_AECnodestrength | Sum_PC | 0.2962873 | 0.70371270 | 0.5 | 0.4210345 | 0.2418278899 | 0.03240674342 | 0.03074251 |
| beta_ROI82_AECnodestrength | Blocktapping | 0.1951480 | 0.80485201 | 0.5 | 0.2424644 | 0.9892506708 | 0.00009915418 | 0.02187812 |
| beta_ROI82_AECnodestrength | Sum_PC | 0.2441662 | 0.75583375 | 0.5 | 0.3230423 | 0.3409303574 | 0.02386780464 | 0.02811825 |
| beta_ROI83_AECnodestrength | Blocktapping | 0.4315981 | 0.56840186 | 0.5 | 0.7593187 | 0.3645839774 | 0.02268584122 | 0.02860239 |
| beta_ROI83_AECnodestrength | Sum_PC | 0.8725797 | 0.12742026 | 0.5 | 6.8480457 | 0.0063297431 | 0.19822898206 | 0.07821806 |
| beta_ROI84_AECnodestrength | Blocktapping | 0.3414913 | 0.65850872 | 0.5 | 0.5185828 | 0.8253370043 | -0.00542143569 | 0.02264102 |
| beta_ROI84_AECnodestrength | Sum_PC | 0.7851769 | 0.21482307 | 0.5 | 3.6549935 | 0.0112795285 | 0.17232553098 | 0.07323592 |

| DV | Predictor | Sum_PostModProb_Incl | Sum_PostModProb_Excl | Sum_PriorModProb_Incl | BF_Incl | Frequentist_p_value | postMean | postStd |
|---|---|---|---|---|---|---|---|---|
| beta_ROI85_AECnodestrength | Blocktapping | 0.2708617 | 0.72913830 | 0.5 | 0.3714819 | 0.9073573954 | 0.00438185976 | 0.02349069 |
| beta_ROI85_AECnodestrength | Sum_PC | 0.5111730 | 0.48882705 | 0.5 | 1.0457133 | 0.0609363390 | 0.09169533165 | 0.05394430 |
| beta_ROI86_AECnodestrength | Blocktapping | 0.3989209 | 0.60107909 | 0.5 | 0.6636746 | 0.5284270179 | 0.01521106965 | 0.02545135 |
| beta_ROI86_AECnodestrength | Sum_PC | 0.9187710 | 0.08122904 | 0.5 | 11.3108689 | 0.0032444318 | 0.23280708780 | 0.08353165 |
| beta_ROI87_AECnodestrength | Blocktapping | 0.2236704 | 0.77632959 | 0.5 | 0.2881127 | 0.9503638779 | 0.00260711494 | 0.02252807 |
| beta_ROI87_AECnodestrength | Sum_PC | 0.3451759 | 0.65482413 | 0.5 | 0.5271276 | 0.1571231916 | 0.04846030280 | 0.03682517 |
| beta_ROI88_AECnodestrength | Blocktapping | 0.3483331 | 0.65166688 | 0.5 | 0.5345263 | 0.8164774796 | -0.00505575993 | 0.02266525 |
| beta_ROI88_AECnodestrength | Sum_PC | 0.8268269 | 0.17317310 | 0.5 | 4.7745690 | 0.0077883126 | 0.19079749642 | 0.07748993 |
| beta_ROI89_AECnodestrength | Blocktapping | 0.2291304 | 0.77086963 | 0.5 | 0.2972362 | 0.9072009041 | -0.00195396708 | 0.02167391 |
| beta_ROI89_AECnodestrength | Sum_PC | 0.3615881 | 0.63841190 | 0.5 | 0.5663868 | 0.1331003507 | 0.05368758914 | 0.03977731 |
| beta_ROI9_AECnodestrength | Blocktapping | 0.2263841 | 0.77361591 | 0.5 | 0.2926311 | 0.4248472980 | -0.01925691881 | 0.02757419 |
| beta_ROI9_AECnodestrength | Sum_PC | 0.2233716 | 0.77662839 | 0.5 | 0.2876171 | 0.4479719276 | 0.01677365860 | 0.02393046 |
| beta_ROI90_AECnodestrength | Blocktapping | 0.2161293 | 0.78387067 | 0.5 | 0.2757206 | 0.8105132309 | 0.00582233716 | 0.02245918 |
| beta_ROI90_AECnodestrength | Sum_PC | 0.2987560 | 0.70124404 | 0.5 | 0.4260371 | 0.2111979179 | 0.03716921267 | 0.03282471 |
| beta_ROI91_AECnodestrength | Blocktapping | 0.2178752 | 0.78212477 | 0.5 | 0.2785684 | 0.6318886272 | 0.01163865234 | 0.02424851 |
| beta_ROI91_AECnodestrength | Sum_PC | 0.2598352 | 0.74016479 | 0.5 | 0.3510505 | 0.3116842187 | 0.02569400914 | 0.02801578 |
| beta_ROI92_AECnodestrength | Blocktapping | 0.2991106 | 0.70088939 | 0.5 | 0.4267586 | 0.7383598527 | 0.00805070927 | 0.02416234 |
| beta_ROI92_AECnodestrength | Sum_PC | 0.5782997 | 0.42170027 | 0.5 | 1.3713525 | 0.0439835326 | 0.11047364056 | 0.05742014 |
| beta_ROI93_AECnodestrength | Blocktapping | 0.2342680 | 0.76573199 | 0.5 | 0.3059399 | 0.7453844753 | 0.00734850861 | 0.02421297 |
| beta_ROI93_AECnodestrength | Sum_PC | 0.3444597 | 0.65554035 | 0.5 | 0.5254591 | 0.1645744542 | 0.04806558313 | 0.03634708 |
| beta_ROI94_AECnodestrength | Blocktapping | 0.3204412 | 0.67955883 | 0.5 | 0.4715430 | 0.9852101224 | 0.00089596546 | 0.02188607 |
| beta_ROI94_AECnodestrength | Sum_PC | 0.7239448 | 0.27605516 | 0.5 | 2.6224645 | 0.0182113721 | 0.15679396072 | 0.06916461 |
| beta_ROI95_AECnodestrength | Blocktapping | 0.3059800 | 0.69402000 | 0.5 | 0.4408807 | 0.9730281187 | 0.00181980762 | 0.02333895 |
| beta_ROI95_AECnodestrength | Sum_PC | 0.6618944 | 0.33810561 | 0.5 | 1.9576557 | 0.0256795838 | 0.13258192703 | 0.06664513 |
| beta_ROI96_AECnodestrength | Blocktapping | 0.3052947 | 0.69470532 | 0.5 | 0.4394592 | 0.6820416283 | -0.00822176714 | 0.02411014 |
| beta_ROI96_AECnodestrength | Sum_PC | 0.6067741 | 0.39322594 | 0.5 | 1.5430672 | 0.0343572291 | 0.11774796527 | 0.05996473 |
| beta_ROI97_AECnodestrength | Blocktapping | 0.3370712 | 0.66292880 | 0.5 | 0.5084576 | 0.6762234656 | 0.01187961466 | 0.02751201 |
| beta_ROI97_AECnodestrength | Sum_PC | 0.5664084 | 0.43359160 | 0.5 | 1.3063177 | 0.0535535162 | 0.10068867924 | 0.05627787 |
| beta_ROI98_AECnodestrength | Blocktapping | 0.3774377 | 0.62256231 | 0.5 | 0.6062649 | 0.5064581083 | 0.01555741638 | 0.02635063 |

| DV | Predictor | Sum_PostModProb_Incl | Sum_PostModProb_Excl | Sum_PriorModProb_Incl | BF_Incl | Frequentist_p_value | postMean | postStd |
|---|---|---|---|---|---|---|---|---|
| beta_ROI98_AECnodestrength | Sum_PC | 0.8036712 | 0.19632880 | 0.5 | 4.0934961 | 0.0115917652 | 0.17503473055 | 0.07518282 |
| beta_ROI99_AECnodestrength | Blocktapping | 0.2399325 | 0.76006747 | 0.5 | 0.3156727 | 0.5485569370 | -0.01351984822 | 0.02481310 |
| beta_ROI99_AECnodestrength | Sum_PC | 0.3195155 | 0.68048449 | 0.5 | 0.4695412 | 0.1849632684 | 0.04217654986 | 0.03437213 |
| BetaPow1 | Blocktapping | 0.2147317 | 0.78526827 | 0.5 | 0.2734502 | 0.6803098397 | 0.00891336880 | 0.02390191 |
| BetaPow1 | Sum_PC | 0.2436173 | 0.75638274 | 0.5 | 0.3220820 | 0.3797899236 | 0.02050455367 | 0.02530436 |
| BetaPow10 | Blocktapping | 0.2301100 | 0.76989004 | 0.5 | 0.2988868 | 0.5374697121 | 0.01451007037 | 0.02546881 |
| BetaPow10 | Sum_PC | 0.2451043 | 0.75489566 | 0.5 | 0.3246864 | 0.3846441001 | 0.02107353699 | 0.02548978 |
| BetaPow100 | Blocktapping | 0.2455165 | 0.75448353 | 0.5 | 0.3254100 | 0.4173197723 | 0.02093915921 | 0.02767328 |
| BetaPow100 | Sum_PC | 0.2513064 | 0.74869358 | 0.5 | 0.3356599 | 0.3873734307 | 0.01956148995 | 0.02548734 |
| BetaPow11 | Blocktapping | 0.2521050 | 0.74789500 | 0.5 | 0.3370861 | 0.4754286078 | 0.01850623195 | 0.02656673 |
| BetaPow11 | Sum_PC | 0.2890068 | 0.71099320 | 0.5 | 0.4064832 | 0.2560181617 | 0.03101608090 | 0.02884786 |
| BetaPow12 | Blocktapping | 0.2123993 | 0.78760068 | 0.5 | 0.2696789 | 0.6059423566 | 0.01161577653 | 0.02431976 |
| BetaPow12 | Sum_PC | 0.2383191 | 0.76168090 | 0.5 | 0.3128857 | 0.3854477454 | 0.01963657554 | 0.02522933 |
| BetaPow13 | Blocktapping | 0.2259475 | 0.77405255 | 0.5 | 0.2919020 | 0.4462752367 | 0.01909777915 | 0.02606763 |
| BetaPow13 | Sum_PC | 0.2093337 | 0.79066626 | 0.5 | 0.2647561 | 0.5887097280 | 0.01034038662 | 0.02168509 |
| BetaPow14 | Blocktapping | 0.4404860 | 0.55951403 | 0.5 | 0.7872653 | 0.1131398616 | 0.06748174737 | 0.04540804 |
| BetaPow14 | Sum_PC | 0.3481554 | 0.65184456 | 0.5 | 0.5341081 | 0.2626218313 | 0.03127179859 | 0.03022399 |
| BetaPow15 | Blocktapping | 0.3679291 | 0.63207088 | 0.5 | 0.5821010 | 0.1536781477 | 0.04989568620 | 0.03551524 |
| BetaPow15 | Sum_PC | 0.3649643 | 0.63503575 | 0.5 | 0.5747145 | 0.1622213781 | 0.04369891287 | 0.03287044 |
| BetaPow16 | Blocktapping | 0.2039049 | 0.79609512 | 0.5 | 0.2561313 | 0.5805662018 | 0.01243473544 | 0.02254463 |
| BetaPow16 | Sum_PC | 0.2006126 | 0.79938741 | 0.5 | 0.2509579 | 0.6514503264 | 0.00928525590 | 0.02134236 |
| BetaPow17 | Blocktapping | 0.2567730 | 0.74322697 | 0.5 | 0.3454840 | 0.3180186995 | 0.02637980943 | 0.03006783 |
| BetaPow17 | Sum_PC | 0.2154641 | 0.78453591 | 0.5 | 0.2746389 | 0.6599088440 | 0.00986064533 | 0.02186484 |
| BetaPow18 | Blocktapping | 0.3090160 | 0.69098401 | 0.5 | 0.4472115 | 0.2171340568 | 0.03860118978 | 0.03453959 |
| BetaPow18 | Sum_PC | 0.2393581 | 0.76064193 | 0.5 | 0.3146790 | 0.5611388829 | 0.01265734892 | 0.02157359 |
| BetaPow19 | Blocktapping | 0.4347214 | 0.56527862 | 0.5 | 0.7690391 | 0.1089105854 | 0.06861077773 | 0.04684796 |
| BetaPow19 | Sum_PC | 0.3209091 | 0.67909094 | 0.5 | 0.4725568 | 0.3460278008 | 0.02456203691 | 0.02779433 |
| BetaPow2 | Blocktapping | 0.2225704 | 0.77742960 | 0.5 | 0.2862901 | 0.5307889364 | 0.01432517765 | 0.02442376 |
| BetaPow2 | Sum_PC | 0.2386659 | 0.76133408 | 0.5 | 0.3134838 | 0.4182696916 | 0.01886895873 | 0.02397025 |
| BetaPow20 | Blocktapping | 0.3326920 | 0.66730800 | 0.5 | 0.4985584 | 0.2024769883 | 0.04534489851 | 0.03756086 |
| BetaPow20 | Sum_PC | 0.2653597 | 0.73464035 | 0.5 | 0.3612103 | 0.4473243036 | 0.01715178025 | 0.02465447 |
| BetaPow21 | Blocktapping | 0.4002729 | 0.59972710 | 0.5 | 0.6674251 | 0.1367923013 | 0.05849667664 | 0.04268009 |
| BetaPow21 | Sum_PC | 0.3047618 | 0.69523819 | 0.5 | 0.4383560 | 0.3640537531 | 0.02291589649 | 0.02711901 |
| BetaPow22 | Blocktapping | 0.2198279 | 0.78017206 | 0.5 | 0.2817685 | 0.6008122172 | 0.01157928109 | 0.02380401 |
| BetaPow22 | Sum_PC | 0.2759599 | 0.72404008 | 0.5 | 0.3811390 | 0.2709732552 | 0.03036625616 | 0.02880896 |
| BetaPow23 | Blocktapping | 0.3700314 | 0.62996856 | 0.5 | 0.5873808 | 0.1919807445 | 0.04705657463 | 0.03810781 |
| BetaPow23 | Sum_PC | 0.3484326 | 0.65156744 | 0.5 | 0.5347605 | 0.2270817976 | 0.03605431315 | 0.03177719 |
| BetaPow24 | Blocktapping | 0.2618856 | 0.73811437 | 0.5 | 0.3548036 | 0.3053067200 | 0.02892215037 | 0.03060295 |
| BetaPow24 | Sum_PC | 0.2266022 | 0.77339783 | 0.5 | 0.2929956 | 0.6586587092 | 0.01000700225 | 0.02258085 |
| BetaPow25 | Blocktapping | 0.2161771 | 0.78382293 | 0.5 | 0.2757983 | 0.7617728237 | 0.00732912745 | 0.02332734 |

| DV | Predictor | Sum_PostModProb_Incl | Sum_PostModProb_Excl | Sum_PriorModProb_Incl | BF_Incl | Frequentist_p_value | postMean | postStd |
|---|---|---|---|---|---|---|---|---|
| BetaPow25 | Sum_PC | 0.2773754 | 0.72262457 | 0.5 | 0.3838444 | 0.2717431594 | 0.03016306331 | 0.02831366 |
| BetaPow26 | Blocktapping | 0.2097980 | 0.79020200 | 0.5 | 0.2654992 | 0.9024405946 | 0.00298201542 | 0.02221392 |
| BetaPow26 | Sum_PC | 0.2864208 | 0.71357917 | 0.5 | 0.4013862 | 0.2348063759 | 0.03334463058 | 0.03034257 |
| BetaPow27 | Blocktapping | 0.1724736 | 0.82752639 | 0.5 | 0.2084207 | 0.8883835446 | -0.00215787069 | 0.02129363 |
| BetaPow27 | Sum_PC | 0.1807452 | 0.81925478 | 0.5 | 0.2206215 | 0.6204913953 | 0.00923093110 | 0.02038767 |
| BetaPow28 | Blocktapping | 0.2026088 | 0.79739119 | 0.5 | 0.2540896 | 0.7053024115 | 0.00697448999 | 0.02284259 |
| BetaPow28 | Sum_PC | 0.2494060 | 0.75059396 | 0.5 | 0.3322782 | 0.3048796021 | 0.02600160967 | 0.02742735 |
| BetaPow29 | Blocktapping | 0.3637208 | 0.63627920 | 0.5 | 0.5716371 | 0.2013342388 | 0.04652350117 | 0.03735812 |
| BetaPow29 | Sum_PC | 0.3601831 | 0.63981692 | 0.5 | 0.5629471 | 0.2065284744 | 0.03835882135 | 0.03441842 |
| BetaPow3 | Blocktapping | 0.2229836 | 0.77701640 | 0.5 | 0.2869741 | 0.6244720659 | 0.01114203413 | 0.02361993 |
| BetaPow3 | Sum_PC | 0.2719387 | 0.72806132 | 0.5 | 0.3735107 | 0.2822680773 | 0.02795261106 | 0.02873752 |
| BetaPow30 | Blocktapping | 0.3731818 | 0.62681820 | 0.5 | 0.5953589 | 0.1859280880 | 0.04668188001 | 0.03916374 |
| BetaPow30 | Sum_PC | 0.3621192 | 0.63788081 | 0.5 | 0.5676910 | 0.2111259388 | 0.03935731255 | 0.03383343 |
| BetaPow31 | Blocktapping | 0.1877648 | 0.81223517 | 0.5 | 0.2311705 | 0.7411772745 | -0.00692599595 | 0.02295065 |
| BetaPow31 | Sum_PC | 0.2003632 | 0.79963682 | 0.5 | 0.2505677 | 0.5448097678 | 0.01152834985 | 0.02178772 |
| BetaPow32 | Blocktapping | 0.1995974 | 0.80040256 | 0.5 | 0.2493713 | 0.8489440679 | 0.00465967701 | 0.02234538 |
| BetaPow32 | Sum_PC | 0.2333164 | 0.76668363 | 0.5 | 0.3043190 | 0.4035286251 | 0.01743206271 | 0.02508036 |
| BetaPow33 | Blocktapping | 0.1970465 | 0.80295353 | 0.5 | 0.2454021 | 0.6899924492 | 0.00796735053 | 0.02303203 |
| BetaPow33 | Sum_PC | 0.2009838 | 0.79901618 | 0.5 | 0.2515391 | 0.6185602862 | 0.00982359922 | 0.02189814 |
| BetaPow34 | Blocktapping | 0.4455770 | 0.55442298 | 0.5 | 0.8036771 | 0.1123061784 | 0.06872552057 | 0.04623296 |
| BetaPow34 | Sum_PC | 0.3453288 | 0.65467117 | 0.5 | 0.5274844 | 0.2865409158 | 0.02775493046 | 0.02868127 |
| BetaPow35 | Blocktapping | 0.1932512 | 0.80674885 | 0.5 | 0.2395431 | 0.8402448736 | -0.00354340655 | 0.02112984 |
| BetaPow35 | Sum_PC | 0.2513282 | 0.74867183 | 0.5 | 0.3356987 | 0.2755260508 | 0.02713483233 | 0.02678228 |
| BetaPow36 | Blocktapping | 0.3317670 | 0.66823298 | 0.5 | 0.4964841 | 0.2004003432 | 0.04328458450 | 0.03628878 |
| BetaPow36 | Sum_PC | 0.2700650 | 0.72993505 | 0.5 | 0.3699849 | 0.4242686344 | 0.01910714490 | 0.02503410 |
| BetaPow37 | Blocktapping | 0.3113621 | 0.68863788 | 0.5 | 0.4521420 | 0.3823750567 | 0.02519048629 | 0.03071713 |
| BetaPow37 | Sum_PC | 0.3674593 | 0.63254070 | 0.5 | 0.5809259 | 0.1821950375 | 0.04521705100 | 0.03433920 |
| BetaPow38 | Blocktapping | 0.3163973 | 0.68360271 | 0.5 | 0.4628380 | 0.7842615191 | 0.00747489189 | 0.03537001 |
| BetaPow38 | Sum_PC | 0.2235659 | 0.77643406 | 0.5 | 0.2879394 | 0.3230352002 | 0.01964938400 | 0.02161687 |
| BetaPow39 | Blocktapping | 0.2127888 | 0.78721120 | 0.5 | 0.2703071 | 0.5392993100 | 0.01367754980 | 0.02443640 |
| BetaPow39 | Sum_PC | 0.2072063 | 0.79279370 | 0.5 | 0.2613622 | 0.5888974639 | 0.01111882569 | 0.02164546 |
| BetaPow4 | Blocktapping | 0.3496664 | 0.65033359 | 0.5 | 0.5376724 | 0.1546343010 | 0.05002012692 | 0.03841465 |
| BetaPow4 | Sum_PC | 0.2418754 | 0.75812458 | 0.5 | 0.3190444 | 0.6545673331 | 0.00911394608 | 0.02115360 |
| BetaPow40 | Blocktapping | 0.2603577 | 0.73964233 | 0.5 | 0.3520048 | 0.3227674070 | 0.02682064752 | 0.02908955 |
| BetaPow40 | Sum_PC | 0.2166563 | 0.78334366 | 0.5 | 0.2765789 | 0.6559259904 | 0.00758525550 | 0.02062879 |
| BetaPow41 | Blocktapping | 0.4399237 | 0.56007631 | 0.5 | 0.7854710 | 0.1146086728 | 0.06581071926 | 0.04531706 |
| BetaPow41 | Sum_PC | 0.3428389 | 0.65716106 | 0.5 | 0.5216970 | 0.2946851703 | 0.02693822000 | 0.02915252 |
| BetaPow42 | Blocktapping | 0.1955971 | 0.80440293 | 0.5 | 0.2431581 | 0.9029426483 | -0.00250938219 | 0.02240275 |
| BetaPow42 | Sum_PC | 0.2348830 | 0.76511697 | 0.5 | 0.3069897 | 0.3684831522 | 0.02068376294 | 0.02543805 |
| BetaPow43 | Blocktapping | 0.1806010 | 0.81939901 | 0.5 | 0.2204067 | 0.8959793236 | -0.00222422226 | 0.02161984 |
| BetaPow43 | Sum_PC | 0.1936219 | 0.80637811 | 0.5 | 0.2401130 | 0.5575459396 | 0.01159874289 | 0.02151941 |

| DV | Predictor | Sum_PostModProb_Incl | Sum_PostModProb_Excl | Sum_PriorModProb_Incl | BF_Incl | Frequentist_p_value | postMean | postStd |
|---|---|---|---|---|---|---|---|---|
| BetaPow44 | Blocktapping | 0.1681343 | 0.83186571 | 0.5 | 0.2021171 | 0.8535410060 | -0.00320395688 | 0.02188906 |
| BetaPow44 | Sum_PC | 0.1660998 | 0.83390017 | 0.5 | 0.1991843 | 0.9829853976 | 0.00058541276 | 0.01858817 |
| BetaPow45 | Blocktapping | 0.1795505 | 0.82044950 | 0.5 | 0.2188441 | 0.5643782866 | -0.01281305753 | 0.02248526 |
| BetaPow45 | Sum_PC | 0.1635920 | 0.83640800 | 0.5 | 0.1955888 | 0.9291686750 | -0.00225498715 | 0.01738353 |
| BetaPow46 | Blocktapping | 0.1582171 | 0.84178289 | 0.5 | 0.1879548 | 0.9838009536 | -0.00038856053 | 0.01959296 |
| BetaPow46 | Sum_PC | 0.1599074 | 0.84009264 | 0.5 | 0.1903449 | 0.8801110024 | 0.00331725848 | 0.01863906 |
| BetaPow47 | Blocktapping | 0.2356780 | 0.76432204 | 0.5 | 0.3083490 | 0.4089568921 | 0.02059645872 | 0.02623831 |
| BetaPow47 | Sum_PC | 0.2548985 | 0.74510146 | 0.5 | 0.3420991 | 0.3169558371 | 0.02574485553 | 0.02728908 |
| BetaPow48 | Blocktapping | 0.2217215 | 0.77827847 | 0.5 | 0.2848871 | 0.5010305935 | 0.01523607628 | 0.02512967 |
| BetaPow48 | Sum_PC | 0.2484344 | 0.75156562 | 0.5 | 0.3305558 | 0.3340759967 | 0.02544961001 | 0.02670603 |
| BetaPow49 | Blocktapping | 0.2552542 | 0.74474583 | 0.5 | 0.3427400 | 0.4564948233 | 0.01964712091 | 0.02609655 |
| BetaPow49 | Sum_PC | 0.3070628 | 0.69293715 | 0.5 | 0.4431323 | 0.2294740300 | 0.03412745996 | 0.03089464 |
| BetaPow5 | Blocktapping | 0.2568118 | 0.74318822 | 0.5 | 0.3455542 | 0.3312171013 | 0.02619531826 | 0.02812982 |
| BetaPow5 | Sum_PC | 0.2300156 | 0.76998445 | 0.5 | 0.2987275 | 0.5044395281 | 0.01444286368 | 0.02228204 |
| BetaPow50 | Blocktapping | 0.2855471 | 0.71445289 | 0.5 | 0.3996724 | 0.3040395281 | 0.02931531944 | 0.03090106 |
| BetaPow50 | Sum_PC | 0.2816270 | 0.71837301 | 0.5 | 0.3920345 | 0.3431747554 | 0.02517660159 | 0.02746032 |
| BetaPow51 | Blocktapping | 0.1962817 | 0.80371828 | 0.5 | 0.2442171 | 0.8758145825 | 0.00314725398 | 0.02296498 |
| BetaPow51 | Sum_PC | 0.2236088 | 0.77639122 | 0.5 | 0.2880104 | 0.4256571927 | 0.01855142230 | 0.02423677 |
| BetaPow52 | Blocktapping | 0.2062582 | 0.79374181 | 0.5 | 0.2598555 | 0.6755704392 | 0.00907507625 | 0.02365569 |
| BetaPow52 | Sum_PC | 0.2261808 | 0.77381919 | 0.5 | 0.2922915 | 0.4668419567 | 0.01624471384 | 0.02364418 |
| BetaPow53 | Blocktapping | 0.2043562 | 0.79564383 | 0.5 | 0.2568438 | 0.9119805697 | 0.00237136853 | 0.02121669 |
| BetaPow53 | Sum_PC | 0.2563398 | 0.74366016 | 0.5 | 0.3447002 | 0.3003208759 | 0.02591111853 | 0.02718768 |
| BetaPow54 | Blocktapping | 0.2596773 | 0.74032275 | 0.5 | 0.3507622 | 0.3391731451 | 0.02637309736 | 0.02840494 |
| BetaPow54 | Sum_PC | 0.2343617 | 0.76563826 | 0.5 | 0.3060998 | 0.4950991570 | 0.01406037553 | 0.02218237 |
| BetaPow55 | Blocktapping | 0.2301374 | 0.76986260 | 0.5 | 0.2989331 | 0.4712928906 | 0.01696726946 | 0.02516693 |
| BetaPow55 | Sum_PC | 0.2368776 | 0.76312242 | 0.5 | 0.3104057 | 0.4308905682 | 0.01734736837 | 0.02424651 |
| BetaPow56 | Blocktapping | 0.2149539 | 0.78504610 | 0.5 | 0.2738105 | 0.6051955805 | 0.01135506632 | 0.02347369 |
| BetaPow56 | Sum_PC | 0.2313356 | 0.76866441 | 0.5 | 0.3009579 | 0.4511835620 | 0.01703359624 | 0.02444150 |
| BetaPow57 | Blocktapping | 0.2567197 | 0.74328030 | 0.5 | 0.3453875 | 0.3501213061 | 0.02437328982 | 0.02825608 |
| BetaPow57 | Sum_PC | 0.2361707 | 0.76382934 | 0.5 | 0.3091930 | 0.4927041325 | 0.01488302946 | 0.02308643 |
| BetaPow58 | Blocktapping | 0.2896073 | 0.71039273 | 0.5 | 0.4076721 | 0.2595350542 | 0.03341620898 | 0.03020893 |
| BetaPow58 | Sum_PC | 0.2279927 | 0.77200727 | 0.5 | 0.2953246 | 0.6920502045 | 0.00835988156 | 0.02134690 |
| BetaPow59 | Blocktapping | 0.2018804 | 0.79811963 | 0.5 | 0.2529450 | 0.9809671923 | 0.00067333979 | 0.02162681 |
| BetaPow59 | Sum_PC | 0.2631101 | 0.73688986 | 0.5 | 0.3570549 | 0.2769086818 | 0.02827353005 | 0.02744670 |
| BetaPow6 | Blocktapping | 0.2599319 | 0.74006811 | 0.5 | 0.3512270 | 0.4415392989 | 0.01988760812 | 0.02755084 |
| BetaPow6 | Sum_PC | 0.3087592 | 0.69124078 | 0.5 | 0.4466739 | 0.2374235356 | 0.03266652357 | 0.03060189 |
| BetaPow60 | Blocktapping | 0.2129216 | 0.78707837 | 0.5 | 0.2705215 | 0.8510506780 | 0.00427262049 | 0.02215892 |
| BetaPow60 | Sum_PC | 0.2819620 | 0.71803803 | 0.5 | 0.3926839 | 0.2458699211 | 0.03134961826 | 0.03080378 |
| BetaPow61 | Blocktapping | 0.1997127 | 0.80028733 | 0.5 | 0.2495512 | 0.7256735293 | -0.00749113411 | 0.02257279 |
| BetaPow61 | Sum_PC | 0.2109600 | 0.78904005 | 0.5 | 0.2673628 | 0.4629481730 | 0.01343683311 | 0.02189949 |
| BetaPow62 | Blocktapping | 0.1840584 | 0.81594163 | 0.5 | 0.2255779 | 0.8607466832 | -0.00321353412 | 0.02176863 |

| DV | Predictor | Sum_PostModProb_Incl | Sum_PostModProb_Excl | Sum_PriorModProb_Incl | BF_Incl | Frequentist_p_value | postMean | postStd |
|---|---|---|---|---|---|---|---|---|
| BetaPow62 | Sum_PC | 0.1933807 | 0.80661930 | 0.5 | 0.2397422 | 0.5785149242 | 0.01024427760 | 0.02039062 |
| BetaPow63 | Blocktapping | 0.2165401 | 0.78345994 | 0.5 | 0.2763895 | 0.5687433971 | 0.01324290228 | 0.02328953 |
| BetaPow63 | Sum_PC | 0.2620124 | 0.73798760 | 0.5 | 0.3550363 | 0.2862926271 | 0.02668713529 | 0.02693178 |
| BetaPow64 | Blocktapping | 0.2952389 | 0.70476109 | 0.5 | 0.4189206 | 0.2555151337 | 0.03271772217 | 0.03211332 |
| BetaPow64 | Sum_PC | 0.2698276 | 0.73017235 | 0.5 | 0.3695397 | 0.3530528808 | 0.02165139608 | 0.02599045 |
| BetaPow65 | Blocktapping | 0.3019048 | 0.69809523 | 0.5 | 0.4324693 | 0.2551781347 | 0.03485486856 | 0.03200574 |
| BetaPow65 | Sum_PC | 0.2808986 | 0.71910137 | 0.5 | 0.3906245 | 0.3398139385 | 0.02404091019 | 0.02599746 |
| BetaPow66 | Blocktapping | 0.4020250 | 0.59797497 | 0.5 | 0.6723108 | 0.1878808156 | 0.04557888499 | 0.03870090 |
| BetaPow66 | Sum_PC | 0.4413220 | 0.55867804 | 0.5 | 0.7899397 | 0.1313494275 | 0.05684762740 | 0.04113666 |
| BetaPow67 | Blocktapping | 0.2260164 | 0.77398361 | 0.5 | 0.2920170 | 0.5804574672 | 0.01178488695 | 0.02317976 |
| BetaPow67 | Sum_PC | 0.2832854 | 0.71671460 | 0.5 | 0.3952555 | 0.2612020841 | 0.02960703378 | 0.02964232 |
| BetaPow68 | Blocktapping | 0.2309987 | 0.76900125 | 0.5 | 0.3003880 | 0.4235495725 | 0.01945876758 | 0.02525241 |
| BetaPow68 | Sum_PC | 0.2321927 | 0.76780734 | 0.5 | 0.3024100 | 0.4260891074 | 0.01690876011 | 0.02290237 |
| BetaPow69 | Blocktapping | 0.2213460 | 0.77865401 | 0.5 | 0.2842674 | 0.4877378453 | 0.01630172259 | 0.02422745 |
| BetaPow69 | Sum_PC | 0.2195839 | 0.78041606 | 0.5 | 0.2813678 | 0.5324672340 | 0.01270005912 | 0.02277109 |
| BetaPow7 | Blocktapping | 0.3032632 | 0.69673680 | 0.5 | 0.4352622 | 0.2356435775 | 0.03570893171 | 0.03307228 |
| BetaPow7 | Sum_PC | 0.2465247 | 0.75347533 | 0.5 | 0.3271835 | 0.5276768201 | 0.01324702222 | 0.02307544 |
| BetaPow70 | Blocktapping | 0.2006624 | 0.79933761 | 0.5 | 0.2510358 | 0.7277828766 | 0.00751691582 | 0.02335644 |
| BetaPow70 | Sum_PC | 0.2166162 | 0.78338378 | 0.5 | 0.2765135 | 0.4785808451 | 0.01509773964 | 0.02278705 |
| BetaPow71 | Blocktapping | 0.2782511 | 0.72174885 | 0.5 | 0.3855235 | 0.3540797864 | 0.02483915856 | 0.02847840 |
| BetaPow71 | Sum_PC | 0.2974255 | 0.70257447 | 0.5 | 0.4233367 | 0.2767982048 | 0.02919225234 | 0.02950400 |
| BetaPow72 | Blocktapping | 0.1865678 | 0.81343220 | 0.5 | 0.2293588 | 0.9261402768 | -0.00176480874 | 0.02128550 |
| BetaPow72 | Sum_PC | 0.2261972 | 0.77380278 | 0.5 | 0.2923190 | 0.3641711879 | 0.02051669426 | 0.02475236 |
| BetaPow73 | Blocktapping | 0.2554829 | 0.74451706 | 0.5 | 0.3431526 | 0.4883080241 | 0.01724735630 | 0.02635055 |
| BetaPow73 | Sum_PC | 0.3370905 | 0.66290953 | 0.5 | 0.5085015 | 0.1772024089 | 0.04307460230 | 0.03474234 |
| BetaPow74 | Blocktapping | 0.2368500 | 0.76315003 | 0.5 | 0.3103583 | 0.6789411316 | 0.00946907147 | 0.02327060 |
| BetaPow74 | Sum_PC | 0.3296061 | 0.67039387 | 0.5 | 0.4916604 | 0.1838046779 | 0.04049801108 | 0.03357505 |
| BetaPow75 | Blocktapping | 0.1980165 | 0.80198349 | 0.5 | 0.2469085 | 0.7874715940 | 0.00581840517 | 0.02093110 |
| BetaPow75 | Sum_PC | 0.2475659 | 0.75243406 | 0.5 | 0.3290201 | 0.3133973111 | 0.02416581148 | 0.02578670 |
| BetaPow76 | Blocktapping | 0.1972182 | 0.80278175 | 0.5 | 0.2456686 | 0.9094515706 | -0.00253493361 | 0.02163724 |
| BetaPow76 | Sum_PC | 0.2365346 | 0.76346539 | 0.5 | 0.3098171 | 0.3583190402 | 0.02185077678 | 0.02629326 |
| BetaPow77 | Blocktapping | 0.2883900 | 0.71161003 | 0.5 | 0.4052641 | 0.3516724346 | 0.02573630272 | 0.02877583 |
| BetaPow77 | Sum_PC | 0.3244285 | 0.67557152 | 0.5 | 0.4802282 | 0.2238427878 | 0.03658376281 | 0.03185589 |
| BetaPow78 | Blocktapping | 0.2889580 | 0.71104202 | 0.5 | 0.4063866 | 0.3131087720 | 0.02826034451 | 0.03052875 |
| BetaPow78 | Sum_PC | 0.3176651 | 0.68233492 | 0.5 | 0.4655560 | 0.2298018074 | 0.03477343077 | 0.03286325 |
| BetaPow79 | Blocktapping | 0.1807878 | 0.81921217 | 0.5 | 0.2206850 | 0.9842721667 | -0.00033782342 | 0.02185468 |
| BetaPow79 | Sum_PC | 0.1943606 | 0.80563941 | 0.5 | 0.2412501 | 0.5650262006 | 0.01129912391 | 0.02227227 |
| BetaPow8 | Blocktapping | 0.3132370 | 0.68676297 | 0.5 | 0.4561065 | 0.2096342240 | 0.04125539726 | 0.03476259 |
| BetaPow8 | Sum_PC | 0.2452353 | 0.75476465 | 0.5 | 0.3249163 | 0.5446524228 | 0.01296664654 | 0.02209991 |
| BetaPow80 | Blocktapping | 0.1998371 | 0.80016288 | 0.5 | 0.2497455 | 0.9932606970 | 0.00027146424 | 0.02206507 |
| BetaPow80 | Sum_PC | 0.2357389 | 0.76426107 | 0.5 | 0.3084534 | 0.4072346891 | 0.01881982196 | 0.02576462 |

| DV | Predictor | Sum_PostModProb_Incl | Sum_PostModProb_Excl | Sum_PriorModProb_Incl | BF_Incl | Frequentist_p_value | postMean | postStd |
|---|---|---|---|---|---|---|---|---|
| BetaPow81 | Blocktapping | 0.2204474 | 0.77955261 | 0.5 | 0.2827871 | 0.4906283936 | 0.01631241075 | 0.02478756 |
| BetaPow81 | Sum_PC | 0.2102477 | 0.78975233 | 0.5 | 0.2662198 | 0.5904459503 | 0.01024477651 | 0.02157235 |
| BetaPow82 | Blocktapping | 0.2007362 | 0.79926380 | 0.5 | 0.2511514 | 0.7165920626 | 0.00796480757 | 0.02336361 |
| BetaPow82 | Sum_PC | 0.2311526 | 0.76884740 | 0.5 | 0.3006482 | 0.3950598975 | 0.01845927138 | 0.02470775 |
| BetaPow83 | Blocktapping | 0.1689503 | 0.83104970 | 0.5 | 0.2032975 | 0.9466955795 | -0.00111222908 | 0.02122637 |
| BetaPow83 | Sum_PC | 0.1685936 | 0.83140641 | 0.5 | 0.2027812 | 0.9803172133 | 0.00075920177 | 0.01946872 |
| BetaPow84 | Blocktapping | 0.1733626 | 0.82663745 | 0.5 | 0.2097202 | 0.8408531426 | -0.00446457175 | 0.02117706 |
| BetaPow84 | Sum_PC | 0.1745435 | 0.82545649 | 0.5 | 0.2114509 | 0.8101431405 | 0.00371804991 | 0.02050657 |
| BetaPow85 | Blocktapping | 0.1671263 | 0.83287369 | 0.5 | 0.2006623 | 0.8969550170 | -0.00359215710 | 0.02167739 |
| BetaPow85 | Sum_PC | 0.1646755 | 0.83532449 | 0.5 | 0.1971396 | 0.9361237348 | 0.00180857572 | 0.01859887 |
| BetaPow86 | Blocktapping | 0.1837770 | 0.81622296 | 0.5 | 0.2251554 | 0.8340331982 | 0.00460520863 | 0.02190250 |
| BetaPow86 | Sum_PC | 0.2227150 | 0.77728503 | 0.5 | 0.2865294 | 0.3623749507 | 0.02107303895 | 0.02457389 |
| BetaPow87 | Blocktapping | 0.2632330 | 0.73676704 | 0.5 | 0.3572811 | 0.4402775996 | 0.02071166128 | 0.02851037 |
| BetaPow87 | Sum_PC | 0.3247866 | 0.67521339 | 0.5 | 0.4810133 | 0.2096995162 | 0.03814731072 | 0.03341864 |
| BetaPow88 | Blocktapping | 0.1793522 | 0.82064783 | 0.5 | 0.2185495 | 0.9751207184 | 0.00046017047 | 0.02156938 |
| BetaPow88 | Sum_PC | 0.2127042 | 0.78729578 | 0.5 | 0.2701707 | 0.4166945643 | 0.01686173269 | 0.02440089 |
| BetaPow89 | Blocktapping | 0.2580842 | 0.74191582 | 0.5 | 0.3478618 | 0.3484757538 | 0.02596708934 | 0.02892450 |
| BetaPow89 | Sum_PC | 0.2413776 | 0.75862239 | 0.5 | 0.3181789 | 0.4386330759 | 0.01672398112 | 0.02424349 |
| BetaPow9 | Blocktapping | 0.3366250 | 0.66337495 | 0.5 | 0.5074431 | 0.1986177703 | 0.04355370837 | 0.03721276 |
| BetaPow9 | Sum_PC | 0.2833689 | 0.71663108 | 0.5 | 0.3954181 | 0.4001469296 | 0.02142634722 | 0.02550629 |
| BetaPow90 | Blocktapping | 0.2249372 | 0.77506283 | 0.5 | 0.2902180 | 0.5952627287 | 0.01055372265 | 0.02313329 |
| BetaPow90 | Sum_PC | 0.2598905 | 0.74010947 | 0.5 | 0.3511515 | 0.3439981730 | 0.02196211142 | 0.02543286 |
| BetaPow91 | Blocktapping | 0.2086464 | 0.79135365 | 0.5 | 0.2636575 | 0.9859101735 | 0.00101844501 | 0.02100444 |
| BetaPow91 | Sum_PC | 0.2759706 | 0.72402935 | 0.5 | 0.3811595 | 0.2591676153 | 0.02966505669 | 0.02961190 |
| BetaPow92 | Blocktapping | 0.1936425 | 0.80635748 | 0.5 | 0.2401448 | 0.8835580006 | -0.00290595838 | 0.02191054 |
| BetaPow92 | Sum_PC | 0.2258691 | 0.77413085 | 0.5 | 0.2917713 | 0.3975818196 | 0.01900494495 | 0.02439880 |
| BetaPow93 | Blocktapping | 0.2141944 | 0.78580559 | 0.5 | 0.2725794 | 0.8808111448 | 0.00403373535 | 0.02197230 |
| BetaPow93 | Sum_PC | 0.2955873 | 0.70441271 | 0.5 | 0.4196223 | 0.2260819434 | 0.03378973484 | 0.03080528 |
| BetaPow94 | Blocktapping | 0.1695670 | 0.83043304 | 0.5 | 0.2041910 | 0.9023812563 | -0.00231623424 | 0.02058347 |
| BetaPow94 | Sum_PC | 0.1860552 | 0.81394483 | 0.5 | 0.2285845 | 0.4946638921 | 0.01290485008 | 0.02094535 |
| BetaPow95 | Blocktapping | 0.1820805 | 0.81791953 | 0.5 | 0.2226142 | 0.7937567397 | -0.00463571879 | 0.02189649 |
| BetaPow95 | Sum_PC | 0.1843293 | 0.81567073 | 0.5 | 0.2259849 | 0.6969166689 | 0.00606062212 | 0.02077312 |
| BetaPow96 | Blocktapping | 0.1683607 | 0.83163928 | 0.5 | 0.2024444 | 0.9957304605 | 0.00010822581 | 0.02119873 |
| BetaPow96 | Sum_PC | 0.1701065 | 0.82989350 | 0.5 | 0.2049739 | 0.9577469066 | 0.00059044362 | 0.01996038 |
| BetaPow97 | Blocktapping | 0.1657611 | 0.83423887 | 0.5 | 0.1986974 | 0.7803737282 | -0.00568453766 | 0.02119229 |
| BetaPow97 | Sum_PC | 0.1648138 | 0.83518618 | 0.5 | 0.1973378 | 0.9803104377 | -0.00103197791 | 0.01832066 |
| BetaPow98 | Blocktapping | 0.1739022 | 0.82609776 | 0.5 | 0.2105105 | 0.8732256676 | 0.00340090249 | 0.02061740 |
| BetaPow98 | Sum_PC | 0.1938179 | 0.80618207 | 0.5 | 0.2404146 | 0.5113136381 | 0.01260459493 | 0.02237991 |
| BetaPow99 | Blocktapping | 0.2178377 | 0.78216228 | 0.5 | 0.2785071 | 0.5532252888 | 0.01250944396 | 0.02469683 |
| BetaPow99 | Sum_PC | 0.2334039 | 0.76659611 | 0.5 | 0.3044679 | 0.4247350327 | 0.01816679941 | 0.02441369 |

| DV | Predictor | Sum_PostModProb_Incl | Sum_PostModProb_Excl | Sum_PriorModProb_Incl | BF_Incl | Frequentist_p_value | postMean | postStd |
|---|---|---|---|---|---|---|---|---|
| gamma_ROI1_AECnodestrength | Blocktapping | 0.1767096 | 0.82329041 | 0.5 | 0.2146382 | 0.9747257563 | 0.00154193548 | 0.02147298 |
| gamma_ROI1_AECnodestrength | Sum_PC | 0.1780463 | 0.82195372 | 0.5 | 0.2166135 | 0.8912966941 | 0.00268296912 | 0.02052896 |
| gamma_ROI10_AECnodestrength | Blocktapping | 0.1940670 | 0.80593298 | 0.5 | 0.2407980 | 0.6956470415 | 0.00770582031 | 0.02282702 |
| gamma_ROI10_AECnodestrength | Sum_PC | 0.2177006 | 0.78229944 | 0.5 | 0.2782829 | 0.4176144599 | -0.01718215955 | 0.02430623 |
| gamma_ROI100_AECnodestrength | Blocktapping | 0.2117501 | 0.78824990 | 0.5 | 0.2686332 | 0.6206181134 | -0.01207866504 | 0.02493333 |
| gamma_ROI100_AECnodestrength | Sum_PC | 0.2170579 | 0.78294210 | 0.5 | 0.2772337 | 0.5217599807 | -0.01410422211 | 0.02372874 |
| gamma_ROI11_AECnodestrength | Blocktapping | 0.1861665 | 0.81383352 | 0.5 | 0.2287525 | 0.9085796542 | -0.00305354641 | 0.02398353 |
| gamma_ROI11_AECnodestrength | Sum_PC | 0.1912660 | 0.80873398 | 0.5 | 0.2365005 | 0.7368379750 | -0.00742579608 | 0.02175658 |
| gamma_ROI12_AECnodestrength | Blocktapping | 0.2012710 | 0.79872901 | 0.5 | 0.2519891 | 0.6096808521 | 0.01175234181 | 0.02404119 |
| gamma_ROI12_AECnodestrength | Sum_PC | 0.2063277 | 0.79367227 | 0.5 | 0.2599659 | 0.5424442691 | -0.01204316573 | 0.02337037 |
| gamma_ROI13_AECnodestrength | Blocktapping | 0.1834974 | 0.81650261 | 0.5 | 0.2247358 | 0.9679166942 | -0.00100005832 | 0.02266541 |
| gamma_ROI13_AECnodestrength | Sum_PC | 0.2052605 | 0.79473947 | 0.5 | 0.2582740 | 0.5514157656 | -0.01263089751 | 0.02338941 |
| gamma_ROI14_AECnodestrength | Blocktapping | 0.2135326 | 0.78646735 | 0.5 | 0.2715086 | 0.5671800756 | -0.01511456925 | 0.02610892 |
| gamma_ROI14_AECnodestrength | Sum_PC | 0.2092498 | 0.79075022 | 0.5 | 0.2646218 | 0.6279629084 | -0.01176216858 | 0.02252388 |
| gamma_ROI15_AECnodestrength | Blocktapping | 0.2306564 | 0.76934360 | 0.5 | 0.2998093 | 0.5780314754 | -0.01465315936 | 0.02559370 |
| gamma_ROI15_AECnodestrength | Sum_PC | 0.2759957 | 0.72400427 | 0.5 | 0.3812073 | 0.3062488869 | -0.02956475388 | 0.02978028 |
| gamma_ROI16_AECnodestrength | Blocktapping | 0.2001349 | 0.79986510 | 0.5 | 0.2502108 | 0.8335061818 | 0.00441794422 | 0.02361466 |
| gamma_ROI16_AECnodestrength | Sum_PC | 0.2481292 | 0.75187084 | 0.5 | 0.3300157 | 0.3355029746 | -0.02422492130 | 0.02808384 |
| gamma_ROI17_AECnodestrength | Blocktapping | 0.1828456 | 0.81715439 | 0.5 | 0.2237589 | 0.6455047439 | -0.00994847840 | 0.02109083 |
| gamma_ROI17_AECnodestrength | Sum_PC | 0.2590757 | 0.74092431 | 0.5 | 0.3496655 | 0.5753143045 | -0.01586443522 | 0.03001818 |
| gamma_ROI18_AECnodestrength | Blocktapping | 0.2072582 | 0.79274176 | 0.5 | 0.2614448 | 0.6741862803 | -0.01005786382 | 0.02415563 |
| gamma_ROI18_AECnodestrength | Sum_PC | 0.2357338 | 0.76426616 | 0.5 | 0.3084447 | 0.4211807979 | -0.01956653088 | 0.02660531 |
| gamma_ROI19_AECnodestrength | Blocktapping | 0.2885737 | 0.71142628 | 0.5 | 0.4056270 | 0.3522132061 | -0.02811605706 | 0.03154081 |
| gamma_ROI19_AECnodestrength | Sum_PC | 0.3212154 | 0.67878463 | 0.5 | 0.4732213 | 0.2512449934 | -0.03645398735 | 0.03225647 |
| gamma_ROI2_AECnodestrength | Blocktapping | 0.1930918 | 0.80690820 | 0.5 | 0.2392984 | 0.7131524356 | -0.01016223297 | 0.02347869 |
| gamma_ROI2_AECnodestrength | Sum_PC | 0.1894174 | 0.81058258 | 0.5 | 0.2336806 | 0.7986005633 | -0.00597286010 | 0.02097721 |
| gamma_ROI20_AECnodestrength | Blocktapping | 0.3417846 | 0.65821535 | 0.5 | 0.5192596 | 0.3663870978 | -0.02573749726 | 0.03019925 |
| gamma_ROI20_AECnodestrength | Sum_PC | 0.5164454 | 0.48355464 | 0.5 | 1.0680186 | 0.0726236485 | -0.08607050315 | 0.05009142 |
| gamma_ROI21_AECnodestrength | Blocktapping | 0.2388240 | 0.76117600 | 0.5 | 0.3137566 | 0.9149211361 | -0.00330849957 | 0.02432886 |

| DV | Predictor | Sum_PostModProb_Incl | Sum_PostModProb_Excl | Sum_PriorModProb_Incl | BF_Incl | Frequentist_p_value | postMean | postStd |
|---|---|---|---|---|---|---|---|---|
| gamma_ROI21_AECnodestrength | Sum_PC | 0.3700346 | 0.62996542 | 0.5 | 0.5873887 | 0.1417295178 | -0.05507586732 | 0.03907856 |
| gamma_ROI22_AECnodestrength | Blocktapping | 0.1929768 | 0.80702324 | 0.5 | 0.2391217 | 0.9671762290 | 0.00188231715 | 0.02364936 |
| gamma_ROI22_AECnodestrength | Sum_PC | 0.2158090 | 0.78419101 | 0.5 | 0.2751995 | 0.4735511913 | 0.01622086818 | 0.02556352 |
| gamma_ROI23_AECnodestrength | Blocktapping | 0.2041718 | 0.79582816 | 0.5 | 0.2565527 | 0.7515734353 | 0.00622693005 | 0.02443608 |
| gamma_ROI23_AECnodestrength | Sum_PC | 0.2315934 | 0.76840657 | 0.5 | 0.3013944 | 0.4012193558 | -0.02066641084 | 0.02490202 |
| gamma_ROI24_AECnodestrength | Blocktapping | 0.3010547 | 0.69894532 | 0.5 | 0.4307271 | 0.3587485141 | -0.02812818803 | 0.03077952 |
| gamma_ROI24_AECnodestrength | Sum_PC | 0.3740534 | 0.62594660 | 0.5 | 0.5975804 | 0.1641470032 | -0.04984608623 | 0.03768967 |
| gamma_ROI25_AECnodestrength | Blocktapping | 0.1848672 | 0.81513283 | 0.5 | 0.2267939 | 0.9143628165 | -0.00218526822 | 0.02228138 |
| gamma_ROI25_AECnodestrength | Sum_PC | 0.1954897 | 0.80451032 | 0.5 | 0.2429921 | 0.6335186492 | 0.00959605335 | 0.02276240 |
| gamma_ROI26_AECnodestrength | Blocktapping | 0.2187245 | 0.78127554 | 0.5 | 0.2799582 | 0.8251241219 | -0.00378198721 | 0.02285173 |
| gamma_ROI26_AECnodestrength | Sum_PC | 0.3167681 | 0.68323192 | 0.5 | 0.4636319 | 0.1998274714 | 0.04475247850 | 0.03525775 |
| gamma_ROI27_AECnodestrength | Blocktapping | 0.2906427 | 0.70935734 | 0.5 | 0.4097267 | 0.3149688297 | -0.02773464410 | 0.02953172 |
| gamma_ROI27_AECnodestrength | Sum_PC | 0.3587758 | 0.64122415 | 0.5 | 0.5595170 | 0.1596685271 | 0.04532019785 | 0.03676048 |
| gamma_ROI28_AECnodestrength | Blocktapping | 0.2330599 | 0.76694010 | 0.5 | 0.3038828 | 0.4034047165 | -0.02312245932 | 0.02924633 |
| gamma_ROI28_AECnodestrength | Sum_PC | 0.2012956 | 0.79870443 | 0.5 | 0.2520276 | 0.9384753381 | -0.00284179390 | 0.02143816 |
| gamma_ROI29_AECnodestrength | Blocktapping | 0.2172220 | 0.78277795 | 0.5 | 0.2775015 | 0.5945713344 | -0.01406551292 | 0.02528262 |
| gamma_ROI29_AECnodestrength | Sum_PC | 0.2247935 | 0.77520650 | 0.5 | 0.2899789 | 0.5110688345 | -0.01541250866 | 0.02478574 |
| gamma_ROI3_AECnodestrength | Blocktapping | 0.2062177 | 0.79378228 | 0.5 | 0.2597913 | 0.5008794481 | -0.01601043855 | 0.02561192 |
| gamma_ROI3_AECnodestrength | Sum_PC | 0.1817148 | 0.81828525 | 0.5 | 0.2220677 | 0.9170382399 | 0.00059125676 | 0.01997136 |
| gamma_ROI30_AECnodestrength | Blocktapping | 0.2155204 | 0.78447959 | 0.5 | 0.2747304 | 0.5627214415 | -0.01436785305 | 0.02490855 |
| gamma_ROI30_AECnodestrength | Sum_PC | 0.2201528 | 0.77984720 | 0.5 | 0.2823025 | 0.5160565738 | -0.01646035874 | 0.02468014 |
| gamma_ROI31_AECnodestrength | Blocktapping | 0.2018981 | 0.79810189 | 0.5 | 0.2529729 | 0.6093398015 | -0.01083744239 | 0.02448811 |
| gamma_ROI31_AECnodestrength | Sum_PC | 0.2290979 | 0.77090207 | 0.5 | 0.2971816 | 0.3956765710 | 0.01966594080 | 0.02497997 |
| gamma_ROI32_AECnodestrength | Blocktapping | 0.1976003 | 0.80239972 | 0.5 | 0.2462617 | 0.6224769706 | -0.01135654664 | 0.02474774 |
| gamma_ROI32_AECnodestrength | Sum_PC | 0.1866948 | 0.81330520 | 0.5 | 0.2295507 | 0.9191535882 | 0.00069954542 | 0.02103908 |
| gamma_ROI33_AECnodestrength | Blocktapping | 0.1873345 | 0.81266552 | 0.5 | 0.2305186 | 0.9418536249 | -0.00218248040 | 0.02345770 |
| gamma_ROI33_AECnodestrength | Sum_PC | 0.2056063 | 0.79439366 | 0.5 | 0.2588217 | 0.5676489990 | -0.01198377940 | 0.02327946 |
| gamma_ROI34_AECnodestrength | Blocktapping | 0.2196747 | 0.78032526 | 0.5 | 0.2815169 | 0.9070775197 | -0.00361571887 | 0.02370777 |
| gamma_ROI34_AECnodestrength | Sum_PC | 0.3055151 | 0.69448492 | 0.5 | 0.4399161 | 0.2198148963 | -0.03902958890 | 0.03337770 |

| DV | Predictor | Sum_PostModProb_Incl | Sum_PostModProb_Excl | Sum_PriorModProb_Incl | BF_Incl | Frequentist_p_value | postMean | postStd |
|---|---|---|---|---|---|---|---|---|
| gamma_ROI35_AECnodestrength | Blocktapping | 0.2342433 | 0.76575674 | 0.5 | 0.3058977 | 0.9601384526 | 0.00201782179 | 0.02297501 |
| gamma_ROI35_AECnodestrength | Sum_PC | 0.3870588 | 0.61294124 | 0.5 | 0.6314778 | 0.1163608045 | 0.06226746571 | 0.04144994 |
| gamma_ROI36_AECnodestrength | Blocktapping | 0.2250101 | 0.77498992 | 0.5 | 0.2903394 | 0.4995382509 | -0.01883327363 | 0.02739268 |
| gamma_ROI36_AECnodestrength | Sum_PC | 0.2141637 | 0.78583627 | 0.5 | 0.2725297 | 0.6121684850 | -0.01233005051 | 0.02364892 |
| gamma_ROI37_AECnodestrength | Blocktapping | 0.1998622 | 0.80013780 | 0.5 | 0.2497847 | 0.6344700042 | -0.01216478307 | 0.02419928 |
| gamma_ROI37_AECnodestrength | Sum_PC | 0.2036867 | 0.79631330 | 0.5 | 0.2557871 | 0.6184086759 | -0.01169984428 | 0.02266772 |
| gamma_ROI38_AECnodestrength | Blocktapping | 0.1884094 | 0.81159064 | 0.5 | 0.2321483 | 0.6787160177 | -0.00936047274 | 0.02303377 |
| gamma_ROI38_AECnodestrength | Sum_PC | 0.1913317 | 0.80866825 | 0.5 | 0.2366010 | 0.7572059757 | -0.00619253264 | 0.02193295 |
| gamma_ROI39_AECnodestrength | Blocktapping | 0.1971535 | 0.80284645 | 0.5 | 0.2455682 | 0.8181905326 | 0.00499742083 | 0.02337104 |
| gamma_ROI39_AECnodestrength | Sum_PC | 0.2304895 | 0.76951051 | 0.5 | 0.2995274 | 0.4197857735 | -0.01997429573 | 0.02682420 |
| gamma_ROI4_AECnodestrength | Blocktapping | 0.1896697 | 0.81033034 | 0.5 | 0.2340646 | 0.8416278499 | -0.00455830503 | 0.02359084 |
| gamma_ROI4_AECnodestrength | Sum_PC | 0.2057234 | 0.79427660 | 0.5 | 0.2590073 | 0.5431520091 | -0.01321336695 | 0.02273424 |
| gamma_ROI40_AECnodestrength | Blocktapping | 0.1918072 | 0.80819281 | 0.5 | 0.2373285 | 0.8816732708 | 0.00215206891 | 0.02305851 |
| gamma_ROI40_AECnodestrength | Sum_PC | 0.2165010 | 0.78349901 | 0.5 | 0.2763258 | 0.4701304974 | -0.01623341101 | 0.02484329 |
| gamma_ROI41_AECnodestrength | Blocktapping | 0.2648396 | 0.73516044 | 0.5 | 0.3602473 | 0.9087051936 | 0.00205371298 | 0.02317260 |
| gamma_ROI41_AECnodestrength | Sum_PC | 0.4934488 | 0.50655121 | 0.5 | 0.9741341 | 0.0669825147 | -0.08738329420 | 0.05046862 |
| gamma_ROI42_AECnodestrength | Blocktapping | 0.2454852 | 0.75451482 | 0.5 | 0.3253550 | 0.5160859078 | -0.01474944324 | 0.02603677 |
| gamma_ROI42_AECnodestrength | Sum_PC | 0.3257161 | 0.67428389 | 0.5 | 0.4830549 | 0.1935515482 | 0.04161631580 | 0.03374501 |
| gamma_ROI43_AECnodestrength | Blocktapping | 0.2357182 | 0.76428180 | 0.5 | 0.3084179 | 0.4683224653 | -0.01726094264 | 0.02654256 |
| gamma_ROI43_AECnodestrength | Sum_PC | 0.2762935 | 0.72370646 | 0.5 | 0.3817757 | 0.2905307562 | 0.02830050846 | 0.02979159 |
| gamma_ROI44_AECnodestrength | Blocktapping | 0.2564763 | 0.74352366 | 0.5 | 0.3449471 | 0.8571088222 | 0.00544616177 | 0.02417093 |
| gamma_ROI44_AECnodestrength | Sum_PC | 0.4305821 | 0.56941792 | 0.5 | 0.7561794 | 0.0988898644 | 0.06842046167 | 0.04642479 |
| gamma_ROI45_AECnodestrength | Blocktapping | 0.2235048 | 0.77649516 | 0.5 | 0.2878380 | 0.7924346434 | -0.00416733644 | 0.02360116 |
| gamma_ROI45_AECnodestrength | Sum_PC | 0.3391336 | 0.66086643 | 0.5 | 0.5131651 | 0.1551113392 | 0.04896574582 | 0.03640398 |
| gamma_ROI46_AECnodestrength | Blocktapping | 0.3268709 | 0.67312914 | 0.5 | 0.4855990 | 0.1906742839 | -0.04941358297 | 0.03848575 |
| gamma_ROI46_AECnodestrength | Sum_PC | 0.2322408 | 0.76775916 | 0.5 | 0.3024918 | 0.7012443930 | -0.00786171773 | 0.02214037 |
| gamma_ROI47_AECnodestrength | Blocktapping | 0.2105795 | 0.78942045 | 0.5 | 0.2667521 | 0.5892619033 | -0.01086176392 | 0.02497633 |
| gamma_ROI47_AECnodestrength | Sum_PC | 0.2287910 | 0.77120905 | 0.5 | 0.2966653 | 0.4485238417 | 0.01709368474 | 0.02547002 |
| gamma_ROI48_AECnodestrength | Blocktapping | 0.2368866 | 0.76311345 | 0.5 | 0.3104211 | 0.3715144014 | -0.02430092385 | 0.02975848 |

| DV | Predictor | Sum_PostModProb_Incl | Sum_PostModProb_Excl | Sum_PriorModProb_Incl | BF_Incl | Frequentist_p_value | postMean | postStd |
|---|---|---|---|---|---|---|---|---|
| gamma_ROI48_AECnodestrength | Sum_PC | 0.1963466 | 0.80365340 | 0.5 | 0.2443175 | 0.9327948209 | 0.00157857831 | 0.02097072 |
| gamma_ROI49_AECnodestrength | Blocktapping | 0.2129921 | 0.78700789 | 0.5 | 0.2706353 | 0.5751621520 | -0.01409117248 | 0.02636233 |
| gamma_ROI49_AECnodestrength | Sum_PC | 0.2019247 | 0.79807535 | 0.5 | 0.2530145 | 0.6005127803 | -0.01103920576 | 0.02169263 |
| gamma_ROI5_AECnodestrength | Blocktapping | 0.4281453 | 0.57185468 | 0.5 | 0.7486960 | 0.0950381926 | -0.08116281188 | 0.05201249 |
| gamma_ROI5_AECnodestrength | Sum_PC | 0.2488174 | 0.75118261 | 0.5 | 0.3312342 | 0.8595907011 | -0.00451278748 | 0.02135519 |
| gamma_ROI50_AECnodestrength | Blocktapping | 0.2322473 | 0.76775266 | 0.5 | 0.3025028 | 0.5246819539 | -0.01734777290 | 0.02628181 |
| gamma_ROI50_AECnodestrength | Sum_PC | 0.2496938 | 0.75030615 | 0.5 | 0.3327893 | 0.4047436156 | -0.02147094607 | 0.02694774 |
| gamma_ROI51_AECnodestrength | Blocktapping | 0.1898254 | 0.81017459 | 0.5 | 0.2343019 | 0.6921700521 | 0.00756402988 | 0.02359568 |
| gamma_ROI51_AECnodestrength | Sum_PC | 0.1935894 | 0.80641055 | 0.5 | 0.2400631 | 0.6183015219 | -0.01003043647 | 0.02245388 |
| gamma_ROI52_AECnodestrength | Blocktapping | 0.1871499 | 0.81285010 | 0.5 | 0.2302391 | 0.8493002195 | -0.00461471756 | 0.02309118 |
| gamma_ROI52_AECnodestrength | Sum_PC | 0.1862121 | 0.81378791 | 0.5 | 0.2288214 | 0.9339469050 | -0.00133755772 | 0.02137592 |
| gamma_ROI53_AECnodestrength | Blocktapping | 0.2220732 | 0.77792682 | 0.5 | 0.2854680 | 0.5171538685 | 0.01698105474 | 0.02653497 |
| gamma_ROI53_AECnodestrength | Sum_PC | 0.2167198 | 0.78328020 | 0.5 | 0.2766823 | 0.5508606607 | 0.01323634127 | 0.02366084 |
| gamma_ROI54_AECnodestrength | Blocktapping | 0.2708831 | 0.72911692 | 0.5 | 0.3715221 | 0.2814576650 | -0.03310245253 | 0.03309018 |
| gamma_ROI54_AECnodestrength | Sum_PC | 0.2109586 | 0.78904142 | 0.5 | 0.2673606 | 0.8037428634 | 0.00436154706 | 0.02150518 |
| gamma_ROI55_AECnodestrength | Blocktapping | 0.4214778 | 0.57852223 | 0.5 | 0.7285420 | 0.1014876823 | -0.07501184669 | 0.04961486 |
| gamma_ROI55_AECnodestrength | Sum_PC | 0.2477882 | 0.75221182 | 0.5 | 0.3294128 | 0.8376350216 | 0.00193205651 | 0.02177966 |
| gamma_ROI56_AECnodestrength | Blocktapping | 0.1858830 | 0.81411700 | 0.5 | 0.2283247 | 0.7963684972 | -0.00562158490 | 0.02352761 |
| gamma_ROI56_AECnodestrength | Sum_PC | 0.1821802 | 0.81781983 | 0.5 | 0.2227632 | 0.8825184045 | -0.00396153097 | 0.02126229 |
| gamma_ROI57_AECnodestrength | Blocktapping | 0.2393076 | 0.76069239 | 0.5 | 0.3145918 | 0.3891239979 | -0.02151134084 | 0.02875038 |
| gamma_ROI57_AECnodestrength | Sum_PC | 0.2231155 | 0.77688451 | 0.5 | 0.2871926 | 0.5351124463 | 0.01360008775 | 0.02410659 |
| gamma_ROI58_AECnodestrength | Blocktapping | 0.3686243 | 0.63137573 | 0.5 | 0.5838429 | 0.1410045361 | -0.06098070742 | 0.04535194 |
| gamma_ROI58_AECnodestrength | Sum_PC | 0.2311510 | 0.76884902 | 0.5 | 0.3006455 | 0.8058388828 | 0.00265822865 | 0.02140479 |
| gamma_ROI59_AECnodestrength | Blocktapping | 0.2409087 | 0.75909135 | 0.5 | 0.3173645 | 0.6482211481 | -0.01226086818 | 0.02550930 |
| gamma_ROI59_AECnodestrength | Sum_PC | 0.3382710 | 0.66172901 | 0.5 | 0.5111926 | 0.1759977512 | -0.04569025481 | 0.03695651 |
| gamma_ROI6_AECnodestrength | Blocktapping | 0.2667822 | 0.73321782 | 0.5 | 0.3638512 | 0.3256218217 | -0.02991023334 | 0.03330366 |
| gamma_ROI6_AECnodestrength | Sum_PC | 0.2071689 | 0.79283111 | 0.5 | 0.2613027 | 0.8300092985 | -0.00477746598 | 0.02121108 |
| gamma_ROI60_AECnodestrength | Blocktapping | 0.2411998 | 0.75880018 | 0.5 | 0.3178700 | 0.4515931533 | -0.02124608660 | 0.02822467 |
| gamma_ROI60_AECnodestrength | Sum_PC | 0.2392365 | 0.76076354 | 0.5 | 0.3144689 | 0.4376805900 | -0.01861505717 | 0.02488359 |

| DV | Predictor | Sum_PostModProb_Incl | Sum_PostModProb_Excl | Sum_PriorModProb_Incl | BF_Incl | Frequentist_p_value | postMean | postStd |
|---|---|---|---|---|---|---|---|---|
| gamma_ROI61_AECnodestrength | Blocktapping | 0.1996157 | 0.80038428 | 0.5 | 0.2493998 | 0.8561962507 | -0.00400139583 | 0.02362471 |
| gamma_ROI61_AECnodestrength | Sum_PC | 0.2277777 | 0.77222229 | 0.5 | 0.2949639 | 0.4409647117 | -0.01932582300 | 0.02573293 |
| gamma_ROI62_AECnodestrength | Blocktapping | 0.2513825 | 0.74861753 | 0.5 | 0.3357956 | 0.3886130575 | -0.02478498513 | 0.02975416 |
| gamma_ROI62_AECnodestrength | Sum_PC | 0.2341156 | 0.76588444 | 0.5 | 0.3056800 | 0.5220114861 | -0.01555550066 | 0.02461605 |
| gamma_ROI63_AECnodestrength | Blocktapping | 0.2481961 | 0.75180391 | 0.5 | 0.3301341 | 0.7229106103 | -0.00914715178 | 0.02399226 |
| gamma_ROI63_AECnodestrength | Sum_PC | 0.3877617 | 0.61223832 | 0.5 | 0.6333509 | 0.1332787270 | -0.05876285718 | 0.04182810 |
| gamma_ROI64_AECnodestrength | Blocktapping | 0.3700607 | 0.62993930 | 0.5 | 0.5874545 | 0.3209647288 | -0.03195593104 | 0.03313891 |
| gamma_ROI64_AECnodestrength | Sum_PC | 0.5454259 | 0.45457413 | 0.5 | 1.1998612 | 0.0643354590 | -0.09388223750 | 0.05291749 |
| gamma_ROI65_AECnodestrength | Blocktapping | 0.3146870 | 0.68531302 | 0.5 | 0.4591872 | 0.4634401658 | -0.02153348727 | 0.02994525 |
| gamma_ROI65_AECnodestrength | Sum_PC | 0.4711410 | 0.52885902 | 0.5 | 0.8908631 | 0.0909955228 | -0.07464712405 | 0.04736864 |
| gamma_ROI66_AECnodestrength | Blocktapping | 0.2390635 | 0.76093653 | 0.5 | 0.3141700 | 0.5180967739 | -0.01751559329 | 0.02760054 |
| gamma_ROI66_AECnodestrength | Sum_PC | 0.2582786 | 0.74172141 | 0.5 | 0.3482151 | 0.3704386064 | -0.02307277560 | 0.02756515 |
| gamma_ROI67_AECnodestrength | Blocktapping | 0.1879602 | 0.81203977 | 0.5 | 0.2314668 | 0.7901738698 | 0.00555676172 | 0.02356434 |
| gamma_ROI67_AECnodestrength | Sum_PC | 0.1891391 | 0.81086089 | 0.5 | 0.2332572 | 0.6949157098 | 0.00789291238 | 0.02168429 |
| gamma_ROI68_AECnodestrength | Blocktapping | 0.4719936 | 0.52800637 | 0.5 | 0.8939165 | 0.1295019213 | -0.06934975248 | 0.04776776 |
| gamma_ROI68_AECnodestrength | Sum_PC | 0.4524217 | 0.54757834 | 0.5 | 0.8262227 | 0.1501588186 | -0.05524539481 | 0.04253927 |
| gamma_ROI69_AECnodestrength | Blocktapping | 0.3362805 | 0.66371952 | 0.5 | 0.5066605 | 0.2974944781 | -0.03510751802 | 0.03364190 |
| gamma_ROI69_AECnodestrength | Sum_PC | 0.3954613 | 0.60453875 | 0.5 | 0.6541537 | 0.1635038183 | -0.04926961058 | 0.03824258 |
| gamma_ROI7_AECnodestrength | Blocktapping | 0.2441184 | 0.75588157 | 0.5 | 0.3229586 | 0.9257843394 | -0.00285180354 | 0.02397793 |
| gamma_ROI7_AECnodestrength | Sum_PC | 0.4111879 | 0.58881210 | 0.5 | 0.6983347 | 0.1097253653 | -0.06550597993 | 0.04386451 |
| gamma_ROI70_AECnodestrength | Blocktapping | 0.2665766 | 0.73342340 | 0.5 | 0.3634689 | 0.5539628185 | -0.01558594101 | 0.02688228 |
| gamma_ROI70_AECnodestrength | Sum_PC | 0.3900160 | 0.60998402 | 0.5 | 0.6393872 | 0.1365466033 | -0.05939413144 | 0.03963767 |
| gamma_ROI71_AECnodestrength | Blocktapping | 0.4226222 | 0.57737777 | 0.5 | 0.7319683 | 0.2100602704 | -0.04701391309 | 0.03989420 |
| gamma_ROI71_AECnodestrength | Sum_PC | 0.5221395 | 0.47786054 | 0.5 | 1.0926608 | 0.0868012194 | -0.07878075762 | 0.04891707 |
| gamma_ROI72_AECnodestrength | Blocktapping | 0.2734985 | 0.72650153 | 0.5 | 0.3764596 | 0.3352903167 | -0.03048510170 | 0.03163875 |
| gamma_ROI72_AECnodestrength | Sum_PC | 0.2605901 | 0.73940990 | 0.5 | 0.3524298 | 0.3940689780 | -0.02333292670 | 0.02714931 |
| gamma_ROI73_AECnodestrength | Blocktapping | 0.2190697 | 0.78093028 | 0.5 | 0.2805240 | 0.5364303745 | -0.01609873121 | 0.02570427 |
| gamma_ROI73_AECnodestrength | Sum_PC | 0.2224000 | 0.77759995 | 0.5 | 0.2860083 | 0.5153490509 | -0.01600282652 | 0.02474537 |
| gamma_ROI74_AECnodestrength | Blocktapping | 0.2023936 | 0.79760638 | 0.5 | 0.2537512 | 0.9900567022 | -0.00077838804 | 0.02430744 |

| DV | Predictor | Sum_PostModProb_Incl | Sum_PostModProb_Excl | Sum_PriorModProb_Incl | BF_Incl | Frequentist_p_value | postMean | postStd |
|---|---|---|---|---|---|---|---|---|
| gamma_ROI74_AECnodestrength | Sum_PC | 0.2418607 | 0.75813930 | 0.5 | 0.3190188 | 0.3637714286 | -0.02291708394 | 0.02661992 |
| gamma_ROI75_AECnodestrength | Blocktapping | 0.2770957 | 0.72290431 | 0.5 | 0.3833089 | 0.2991514885 | -0.03346059610 | 0.03331728 |
| gamma_ROI75_AECnodestrength | Sum_PC | 0.2357162 | 0.76428376 | 0.5 | 0.3084146 | 0.5265382166 | -0.01501065315 | 0.02391876 |
| gamma_ROI76_AECnodestrength | Blocktapping | 0.1867453 | 0.81325468 | 0.5 | 0.2296271 | 0.8679100754 | 0.00286824650 | 0.02293766 |
| gamma_ROI76_AECnodestrength | Sum_PC | 0.2001421 | 0.79985791 | 0.5 | 0.2502221 | 0.5736868375 | -0.01280374938 | 0.02228909 |
| gamma_ROI77_AECnodestrength | Blocktapping | 0.2327668 | 0.76723319 | 0.5 | 0.3033847 | 0.5886549617 | -0.01409327505 | 0.02581712 |
| gamma_ROI77_AECnodestrength | Sum_PC | 0.2929502 | 0.70704981 | 0.5 | 0.4143275 | 0.2584990203 | -0.03419858205 | 0.03142981 |
| gamma_ROI78_AECnodestrength | Blocktapping | 0.2415374 | 0.75846262 | 0.5 | 0.3184565 | 0.4332961987 | -0.02115104458 | 0.02912746 |
| gamma_ROI78_AECnodestrength | Sum_PC | 0.2298696 | 0.77013042 | 0.5 | 0.2984814 | 0.5359812850 | -0.01560946322 | 0.02435846 |
| gamma_ROI79_AECnodestrength | Blocktapping | 0.1933374 | 0.80666263 | 0.5 | 0.2396756 | 0.9687429612 | 0.00245457607 | 0.02212542 |
| gamma_ROI79_AECnodestrength | Sum_PC | 0.2346294 | 0.76537058 | 0.5 | 0.3065566 | 0.3643415105 | 0.02249492692 | 0.02598457 |
| gamma_ROI8_AECnodestrength | Blocktapping | 0.2082167 | 0.79178328 | 0.5 | 0.2629719 | 0.6841088085 | -0.00929955457 | 0.02500529 |
| gamma_ROI8_AECnodestrength | Sum_PC | 0.2239655 | 0.77603454 | 0.5 | 0.2886024 | 0.5087473511 | -0.01539865949 | 0.02480828 |
| gamma_ROI80_AECnodestrength | Blocktapping | 0.2044863 | 0.79551368 | 0.5 | 0.2570494 | 0.8583018362 | 0.00290918158 | 0.02257743 |
| gamma_ROI80_AECnodestrength | Sum_PC | 0.2626504 | 0.73734965 | 0.5 | 0.3562087 | 0.2955795860 | -0.02674256298 | 0.02873017 |
| gamma_ROI81_AECnodestrength | Blocktapping | 0.6413525 | 0.35864755 | 0.5 | 1.7882527 | 0.0603666373 | -0.10951273584 | 0.06235450 |
| gamma_ROI81_AECnodestrength | Sum_PC | 0.5735875 | 0.42641246 | 0.5 | 1.3451472 | 0.1237590058 | -0.06729616292 | 0.04581491 |
| gamma_ROI82_AECnodestrength | Blocktapping | 0.2114499 | 0.78855010 | 0.5 | 0.2681502 | 0.7801343601 | -0.00715378445 | 0.02453934 |
| gamma_ROI82_AECnodestrength | Sum_PC | 0.2654551 | 0.73454491 | 0.5 | 0.3613871 | 0.3231053721 | -0.02802607861 | 0.02992682 |
| gamma_ROI83_AECnodestrength | Blocktapping | 0.2035841 | 0.79641586 | 0.5 | 0.2556254 | 0.4564595410 | 0.01762748334 | 0.02542301 |
| gamma_ROI83_AECnodestrength | Sum_PC | 0.1945593 | 0.80544069 | 0.5 | 0.2415563 | 0.5251306140 | 0.01282106106 | 0.02169707 |
| gamma_ROI84_AECnodestrength | Blocktapping | 0.1781033 | 0.82189671 | 0.5 | 0.2166979 | 0.9944068037 | 0.00009489178 | 0.02210741 |
| gamma_ROI84_AECnodestrength | Sum_PC | 0.1822847 | 0.81771534 | 0.5 | 0.2229195 | 0.7403552574 | 0.00602523893 | 0.02108124 |
| gamma_ROI85_AECnodestrength | Blocktapping | 0.1830950 | 0.81690501 | 0.5 | 0.2241325 | 0.7263647039 | -0.00728124567 | 0.02339766 |
| gamma_ROI85_AECnodestrength | Sum_PC | 0.1845355 | 0.81546446 | 0.5 | 0.2262950 | 0.6576968233 | 0.00844943564 | 0.02090427 |
| gamma_ROI86_AECnodestrength | Blocktapping | 0.2540864 | 0.74591358 | 0.5 | 0.3406379 | 0.4493084832 | -0.02080557385 | 0.02805205 |
| gamma_ROI86_AECnodestrength | Sum_PC | 0.2747293 | 0.72527075 | 0.5 | 0.3787954 | 0.3379337206 | -0.02540987136 | 0.02826014 |
| gamma_ROI87_AECnodestrength | Blocktapping | 0.1994197 | 0.80058027 | 0.5 | 0.2490940 | 0.6560540764 | -0.01024073399 | 0.02512930 |
| gamma_ROI87_AECnodestrength | Sum_PC | 0.1981224 | 0.80187757 | 0.5 | 0.2470732 | 0.6552057879 | -0.01023214243 | 0.02214352 |

| DV | Predictor | Sum_PostModProb_Incl | Sum_PostModProb_Excl | Sum_PriorModProb_Incl | BF_Incl | Frequentist_p_value | postMean | postStd |
|---|---|---|---|---|---|---|---|---|
| gamma_ROI88_AECnodestrength | Blocktapping | 0.2286645 | 0.77133553 | 0.5 | 0.2964527 | 0.4705899806 | -0.01942503413 | 0.02841557 |
| gamma_ROI88_AECnodestrength | Sum_PC | 0.2025069 | 0.79749307 | 0.5 | 0.2539294 | 0.8791730082 | -0.00434674077 | 0.02173767 |
| gamma_ROI89_AECnodestrength | Blocktapping | 0.2587280 | 0.74127205 | 0.5 | 0.3490324 | 0.4057524204 | -0.02394778937 | 0.02866340 |
| gamma_ROI89_AECnodestrength | Sum_PC | 0.2697101 | 0.73028994 | 0.5 | 0.3693191 | 0.3488665517 | -0.02575122613 | 0.02851090 |
| gamma_ROI9_AECnodestrength | Blocktapping | 0.2707521 | 0.72924794 | 0.5 | 0.3712757 | 0.2955936818 | -0.03454936080 | 0.03389259 |
| gamma_ROI9_AECnodestrength | Sum_PC | 0.2087775 | 0.79122251 | 0.5 | 0.2638670 | 0.9800497475 | -0.00089418530 | 0.02134342 |
| gamma_ROI90_AECnodestrength | Blocktapping | 0.2415717 | 0.75842834 | 0.5 | 0.3185161 | 0.4217860513 | -0.02295416347 | 0.02895832 |
| gamma_ROI90_AECnodestrength | Sum_PC | 0.2260452 | 0.77395483 | 0.5 | 0.2920651 | 0.5296286592 | -0.01412746741 | 0.02486716 |
| gamma_ROI91_AECnodestrength | Blocktapping | 0.2208071 | 0.77919289 | 0.5 | 0.2833793 | 0.7479586448 | -0.00677111302 | 0.02247093 |
| gamma_ROI91_AECnodestrength | Sum_PC | 0.3297393 | 0.67026071 | 0.5 | 0.4919568 | 0.1715569162 | -0.04707531138 | 0.03524105 |
| gamma_ROI92_AECnodestrength | Blocktapping | 0.1999465 | 0.80005354 | 0.5 | 0.2499163 | 0.9910413835 | -0.00094592667 | 0.02266808 |
| gamma_ROI92_AECnodestrength | Sum_PC | 0.2495585 | 0.75044146 | 0.5 | 0.3325490 | 0.3371695756 | -0.02527152921 | 0.02820254 |
| gamma_ROI93_AECnodestrength | Blocktapping | 0.2227484 | 0.77725160 | 0.5 | 0.2865847 | 0.6921367971 | -0.01068413085 | 0.02471510 |
| gamma_ROI93_AECnodestrength | Sum_PC | 0.3008170 | 0.69918300 | 0.5 | 0.4302407 | 0.2257075379 | -0.03903132109 | 0.03237004 |
| gamma_ROI94_AECnodestrength | Blocktapping | 0.1806358 | 0.81936418 | 0.5 | 0.2204585 | 0.9087478522 | 0.00270126557 | 0.02243250 |
| gamma_ROI94_AECnodestrength | Sum_PC | 0.1840096 | 0.81599044 | 0.5 | 0.2255045 | 0.7891921905 | -0.00530877527 | 0.02141692 |
| gamma_ROI95_AECnodestrength | Blocktapping | 0.1782489 | 0.82175113 | 0.5 | 0.2169134 | 0.8902087094 | -0.00335644792 | 0.02240273 |
| gamma_ROI95_AECnodestrength | Sum_PC | 0.1838141 | 0.81618586 | 0.5 | 0.2252111 | 0.7577439134 | -0.00653042623 | 0.02066859 |
| gamma_ROI96_AECnodestrength | Blocktapping | 0.2158668 | 0.78413317 | 0.5 | 0.2752936 | 0.9438824790 | -0.00143241083 | 0.02297824 |
| gamma_ROI96_AECnodestrength | Sum_PC | 0.3067651 | 0.69323495 | 0.5 | 0.4425124 | 0.2163443160 | 0.03975840293 | 0.03435287 |
| gamma_ROI97_AECnodestrength | Blocktapping | 0.1806019 | 0.81939805 | 0.5 | 0.2204081 | 0.9083676125 | -0.00316811323 | 0.02281746 |
| gamma_ROI97_AECnodestrength | Sum_PC | 0.1815185 | 0.81848154 | 0.5 | 0.2217746 | 0.7768457105 | -0.00607385163 | 0.02080063 |
| gamma_ROI98_AECnodestrength | Blocktapping | 0.2722744 | 0.72772562 | 0.5 | 0.3741443 | 0.3506798893 | -0.02979957104 | 0.03000356 |
| gamma_ROI98_AECnodestrength | Sum_PC | 0.2764776 | 0.72352237 | 0.5 | 0.3821273 | 0.3299762993 | -0.02707629969 | 0.02900468 |
| gamma_ROI99_AECnodestrength | Blocktapping | 0.1991809 | 0.80081907 | 0.5 | 0.2487215 | 0.5710060850 | -0.01309098334 | 0.02392924 |
| gamma_ROI99_AECnodestrength | Sum_PC | 0.1919633 | 0.80803671 | 0.5 | 0.2375675 | 0.6888865835 | -0.00797468436 | 0.02129922 |
| GammaPow1 | Blocktapping | 0.2092146 | 0.79078535 | 0.5 | 0.2645657 | 0.5182598376 | -0.01503515109 | 0.02519933 |
| GammaPow1 | Sum_PC | 0.2020621 | 0.79793793 | 0.5 | 0.2532303 | 0.6194463336 | -0.00995524400 | 0.02103943 |
| GammaPow10 | Blocktapping | 0.3266827 | 0.67331731 | 0.5 | 0.4851839 | 0.4125763657 | -0.02243960916 | 0.02879799 |
| GammaPow10 | Sum_PC | 0.5117400 | 0.48826002 | 0.5 | 1.0480891 | 0.0742192244 | -0.08282813003 | 0.04999181 |

| DV | Predictor | Sum_PostModProb_Incl | Sum_PostModProb_Excl | Sum_PriorModProb_Incl | BF_Incl | Frequentist_p_value | postMean | postStd |
|---|---|---|---|---|---|---|---|---|
| GammaPow100 | Blocktapping | 0.2116619 | 0.78833815 | 0.5 | 0.2684912 | 0.4302460905 | -0.01903013253 | 0.02480553 |
| GammaPow100 | Sum_PC | 0.1821316 | 0.81786838 | 0.5 | 0.2226906 | 0.8234026017 | -0.00467235483 | 0.01868997 |
| GammaPow11 | Blocktapping | 0.2824704 | 0.71752956 | 0.5 | 0.3936708 | 0.4577710698 | -0.02061030345 | 0.02686491 |
| GammaPow11 | Sum_PC | 0.3838991 | 0.61610093 | 0.5 | 0.6231107 | 0.1472265182 | -0.05338750602 | 0.03875168 |
| GammaPow12 | Blocktapping | 0.3597643 | 0.64023568 | 0.5 | 0.5619248 | 0.3598603597 | -0.02731051854 | 0.02929943 |
| GammaPow12 | Sum_PC | 0.5641145 | 0.43588548 | 0.5 | 1.2941806 | 0.0592638475 | -0.09585963528 | 0.05249874 |
| GammaPow13 | Blocktapping | 0.3180456 | 0.68195444 | 0.5 | 0.4663736 | 0.5661940293 | -0.01565094070 | 0.02586461 |
| GammaPow13 | Sum_PC | 0.5612562 | 0.43874379 | 0.5 | 1.2792345 | 0.0542613211 | -0.10098999168 | 0.05503753 |
| GammaPow14 | Blocktapping | 0.2567888 | 0.74321118 | 0.5 | 0.3455126 | 0.7430115215 | 0.00584752658 | 0.02256711 |
| GammaPow14 | Sum_PC | 0.4434733 | 0.55652670 | 0.5 | 0.7968590 | 0.0886354324 | -0.07142487550 | 0.04513424 |
| GammaPow15 | Blocktapping | 0.2125845 | 0.78741553 | 0.5 | 0.2699775 | 0.6870394316 | -0.00906412980 | 0.02425628 |
| GammaPow15 | Sum_PC | 0.2455714 | 0.75442863 | 0.5 | 0.3255064 | 0.3714401263 | -0.02272145798 | 0.02546460 |
| GammaPow16 | Blocktapping | 0.1899056 | 0.81009436 | 0.5 | 0.2344241 | 0.7738895457 | -0.00697031676 | 0.02084674 |
| GammaPow16 | Sum_PC | 0.2242923 | 0.77570767 | 0.5 | 0.2891454 | 0.4067911206 | -0.01921548996 | 0.02395314 |
| GammaPow17 | Blocktapping | 0.2657206 | 0.73427941 | 0.5 | 0.3618794 | 0.7968984515 | -0.00641510449 | 0.02312693 |
| GammaPow17 | Sum_PC | 0.4628502 | 0.53714981 | 0.5 | 0.8616780 | 0.0861910498 | -0.07786914764 | 0.04788164 |
| GammaPow18 | Blocktapping | 0.1926203 | 0.80737965 | 0.5 | 0.2385747 | 0.7462314128 | -0.00746470638 | 0.02133716 |
| GammaPow18 | Sum_PC | 0.2157849 | 0.78421512 | 0.5 | 0.2751603 | 0.4522228676 | -0.01665353814 | 0.02340352 |
| GammaPow19 | Blocktapping | 0.2409171 | 0.75908289 | 0.5 | 0.3173792 | 0.9906073007 | -0.00203054544 | 0.02250126 |
| GammaPow19 | Sum_PC | 0.4082101 | 0.59178995 | 0.5 | 0.6897888 | 0.1169157109 | -0.06245414209 | 0.04367889 |
| GammaPow2 | Blocktapping | 0.1804820 | 0.81951803 | 0.5 | 0.2202294 | 0.5995775922 | -0.01061470725 | 0.02156431 |
| GammaPow2 | Sum_PC | 0.1681614 | 0.83183860 | 0.5 | 0.2021563 | 0.8734727286 | 0.00208522353 | 0.01787800 |
| GammaPow20 | Blocktapping | 0.1979740 | 0.80202595 | 0.5 | 0.2468424 | 0.7683491090 | -0.00759561660 | 0.02155540 |
| GammaPow20 | Sum_PC | 0.2466376 | 0.75336240 | 0.5 | 0.3273824 | 0.3335699039 | -0.02438580034 | 0.02540060 |
| GammaPow21 | Blocktapping | 0.2155992 | 0.78440082 | 0.5 | 0.2748584 | 0.9214393523 | -0.00358400704 | 0.02192120 |
| GammaPow21 | Sum_PC | 0.3122300 | 0.68776997 | 0.5 | 0.4539745 | 0.2049936516 | -0.04114624583 | 0.03405994 |
| GammaPow22 | Blocktapping | 0.2759928 | 0.72400715 | 0.5 | 0.3812018 | 0.5146043882 | -0.01703275067 | 0.02571192 |
| GammaPow22 | Sum_PC | 0.4226828 | 0.57731719 | 0.5 | 0.7321500 | 0.1140317104 | -0.06334218615 | 0.04182227 |
| GammaPow23 | Blocktapping | 0.2226757 | 0.77732428 | 0.5 | 0.2864644 | 0.9983893292 | -0.00108491204 | 0.02275826 |
| GammaPow23 | Sum_PC | 0.3345167 | 0.66548326 | 0.5 | 0.5026674 | 0.1786723217 | -0.04368293013 | 0.03561557 |
| GammaPow24 | Blocktapping | 0.2631267 | 0.73687326 | 0.5 | 0.3570855 | 0.9045874097 | -0.00385856346 | 0.02298765 |
| GammaPow24 | Sum_PC | 0.4603917 | 0.53960832 | 0.5 | 0.8531960 | 0.0832611510 | -0.07625620748 | 0.04666461 |
| GammaPow25 | Blocktapping | 0.3426122 | 0.65738782 | 0.5 | 0.5211721 | 0.3036965300 | -0.03173545559 | 0.03202607 |
| GammaPow25 | Sum_PC | 0.4376094 | 0.56239062 | 0.5 | 0.7781236 | 0.1219730847 | -0.06308536101 | 0.04092915 |
| GammaPow26 | Blocktapping | 0.3538996 | 0.64610037 | 0.5 | 0.5477472 | 0.2927925526 | -0.03455357065 | 0.03351421 |
| GammaPow26 | Sum_PC | 0.4581916 | 0.54180840 | 0.5 | 0.8456709 | 0.1108074443 | -0.06680978223 | 0.04320925 |
| GammaPow27 | Blocktapping | 0.2429296 | 0.75707036 | 0.5 | 0.3208812 | 0.6150239788 | -0.01209369595 | 0.02448597 |
| GammaPow27 | Sum_PC | 0.3475418 | 0.65245820 | 0.5 | 0.5326652 | 0.1698123877 | -0.04772202090 | 0.03537930 |
| GammaPow28 | Blocktapping | 0.2275018 | 0.77249818 | 0.5 | 0.2945014 | 0.7490685780 | -0.00888491709 | 0.02397623 |
| GammaPow28 | Sum_PC | 0.3153458 | 0.68465418 | 0.5 | 0.4605914 | 0.1954249723 | -0.04310108922 | 0.03359473 |
| GammaPow29 | Blocktapping | 0.2164454 | 0.78355459 | 0.5 | 0.2762353 | 0.6619335057 | -0.01033017991 | 0.02335413 |

| DV | Predictor | Sum_PostModProb_Incl | Sum_PostModProb_Excl | Sum_PriorModProb_Incl | BF_Incl | Frequentist_p_value | postMean | postStd |
|---|---|---|---|---|---|---|---|---|
| GammaPow29 | Sum_PC | 0.2619498 | 0.73805024 | 0.5 | 0.3549213 | 0.3151559026 | -0.02635282290 | 0.02791736 |
| GammaPow3 | Blocktapping | 0.1988212 | 0.80117876 | 0.5 | 0.2481609 | 0.5078682371 | -0.01517930427 | 0.02399392 |
| GammaPow3 | Sum_PC | 0.1749757 | 0.82502427 | 0.5 | 0.2120856 | 0.9626434493 | 0.00014406601 | 0.01823979 |
| GammaPow30 | Blocktapping | 0.2125413 | 0.78745870 | 0.5 | 0.2699079 | 0.7633066805 | -0.00729031487 | 0.02285069 |
| GammaPow30 | Sum_PC | 0.2903940 | 0.70960604 | 0.5 | 0.4092326 | 0.2418487797 | -0.03425449700 | 0.03075869 |
| GammaPow31 | Blocktapping | 0.3172223 | 0.68277768 | 0.5 | 0.4646056 | 0.2785255770 | -0.03564626924 | 0.03350112 |
| GammaPow31 | Sum_PC | 0.3375795 | 0.66242047 | 0.5 | 0.5096152 | 0.2343627559 | -0.03827871281 | 0.03321107 |
| GammaPow32 | Blocktapping | 0.2736130 | 0.72638700 | 0.5 | 0.3766766 | 0.3495575467 | -0.02699922991 | 0.02884119 |
| GammaPow32 | Sum_PC | 0.3021467 | 0.69785328 | 0.5 | 0.4329660 | 0.2602681217 | -0.03325350056 | 0.02964832 |
| GammaPow33 | Blocktapping | 0.2511996 | 0.74880037 | 0.5 | 0.3354694 | 0.5056467338 | -0.01781537503 | 0.02577076 |
| GammaPow33 | Sum_PC | 0.3379191 | 0.66208086 | 0.5 | 0.5103895 | 0.1873354350 | -0.04208657291 | 0.03287693 |
| GammaPow34 | Blocktapping | 0.2297940 | 0.77020604 | 0.5 | 0.2983539 | 0.8597204124 | -0.00497568434 | 0.02350536 |
| GammaPow34 | Sum_PC | 0.3385505 | 0.66144945 | 0.5 | 0.5118313 | 0.1779414668 | -0.04496165184 | 0.03551194 |
| GammaPow35 | Blocktapping | 0.2910272 | 0.70897285 | 0.5 | 0.4104912 | 0.3576653796 | -0.02740646306 | 0.02974864 |
| GammaPow35 | Sum_PC | 0.3555933 | 0.64440675 | 0.5 | 0.5518149 | 0.1804101263 | -0.04569569070 | 0.03497989 |
| GammaPow36 | Blocktapping | 0.1897825 | 0.81021746 | 0.5 | 0.2342366 | 0.5463101387 | -0.01337561005 | 0.02281818 |
| GammaPow36 | Sum_PC | 0.1743864 | 0.82561357 | 0.5 | 0.2112204 | 0.8842332216 | -0.00309066673 | 0.01841600 |
| GammaPow37 | Blocktapping | 0.2009722 | 0.79902777 | 0.5 | 0.2515210 | 0.5480976930 | -0.01412886698 | 0.02350415 |
| GammaPow37 | Sum_PC | 0.2121727 | 0.78782730 | 0.5 | 0.2693137 | 0.4421283507 | -0.01679822728 | 0.02237368 |
| GammaPow38 | Blocktapping | 0.3186507 | 0.68134930 | 0.5 | 0.4676760 | 0.3454441881 | -0.02857847630 | 0.03067046 |
| GammaPow38 | Sum_PC | 0.4137932 | 0.58620678 | 0.5 | 0.7058827 | 0.1309146750 | -0.05740332116 | 0.04000218 |
| GammaPow39 | Blocktapping | 0.3008816 | 0.69911843 | 0.5 | 0.4303728 | 0.4818435879 | -0.01977664409 | 0.02617258 |
| GammaPow39 | Sum_PC | 0.4727449 | 0.52725506 | 0.5 | 0.8966153 | 0.0867483889 | -0.07295842256 | 0.04607624 |
| GammaPow4 | Blocktapping | 0.1751457 | 0.82485426 | 0.5 | 0.2123354 | 0.9127857437 | -0.00230888389 | 0.02026355 |
| GammaPow4 | Sum_PC | 0.1708665 | 0.82913351 | 0.5 | 0.2060784 | 0.9718762414 | 0.00084948187 | 0.01912115 |
| GammaPow40 | Blocktapping | 0.2351844 | 0.76481559 | 0.5 | 0.3075047 | 0.9984721026 | -0.00150822591 | 0.02188018 |
| GammaPow40 | Sum_PC | 0.3861251 | 0.61387494 | 0.5 | 0.6289963 | 0.1283515075 | -0.05522498296 | 0.03961017 |
| GammaPow41 | Blocktapping | 0.2152068 | 0.78479323 | 0.5 | 0.2742210 | 0.7946481547 | 0.00453856630 | 0.02310783 |
| GammaPow41 | Sum_PC | 0.2940092 | 0.70599077 | 0.5 | 0.4164491 | 0.2413588692 | -0.03405055636 | 0.03185660 |
| GammaPow42 | Blocktapping | 0.3773303 | 0.62266971 | 0.5 | 0.6059879 | 0.2461902761 | -0.04180916470 | 0.03583930 |
| GammaPow42 | Sum_PC | 0.4349322 | 0.56506775 | 0.5 | 0.7696993 | 0.1252182580 | -0.06085843138 | 0.03993296 |
| GammaPow43 | Blocktapping | 0.3721940 | 0.62780600 | 0.5 | 0.5928487 | 0.2708697425 | -0.03690275243 | 0.03522073 |
| GammaPow43 | Sum_PC | 0.4877077 | 0.51229234 | 0.5 | 0.9520104 | 0.0969936347 | -0.07447858787 | 0.04546951 |
| GammaPow44 | Blocktapping | 0.3001943 | 0.69980568 | 0.5 | 0.4289681 | 0.3193494346 | -0.02934031667 | 0.03153261 |
| GammaPow44 | Sum_PC | 0.3503092 | 0.64969075 | 0.5 | 0.5391938 | 0.1914709753 | -0.04291041040 | 0.03504367 |
| GammaPow45 | Blocktapping | 0.3469155 | 0.65308454 | 0.5 | 0.5311953 | 0.3357418064 | -0.02750303418 | 0.02987972 |
| GammaPow45 | Sum_PC | 0.5318788 | 0.46812118 | 0.5 | 1.1361990 | 0.0682645946 | -0.08805096961 | 0.05037354 |
| GammaPow46 | Blocktapping | 0.2235969 | 0.77640309 | 0.5 | 0.2879907 | 0.6677222078 | -0.00986947806 | 0.02236166 |
| GammaPow46 | Sum_PC | 0.3441545 | 0.65584550 | 0.5 | 0.5247493 | 0.1403468826 | -0.05184688334 | 0.03569857 |
| GammaPow47 | Blocktapping | 0.2076735 | 0.79232649 | 0.5 | 0.2621060 | 0.8381858636 | -0.00557990201 | 0.02238932 |
| GammaPow47 | Sum_PC | 0.2991345 | 0.70086553 | 0.5 | 0.4268072 | 0.2057646033 | -0.03958624871 | 0.03236329 |

| DV | Predictor | Sum_PostModProb_Incl | Sum_PostModProb_Excl | Sum_PriorModProb_Incl | BF_Incl | Frequentist_p_value | postMean | postStd |
|---|---|---|---|---|---|---|---|---|
| GammaPow48 | Blocktapping | 0.2074031 | 0.79259694 | 0.5 | 0.2616753 | 0.9380765692 | -0.00329376040 | 0.02199718 |
| GammaPow48 | Sum_PC | 0.2921155 | 0.70788445 | 0.5 | 0.4126599 | 0.2153223035 | -0.03762915396 | 0.03091568 |
| GammaPow49 | Blocktapping | 0.1987538 | 0.80124623 | 0.5 | 0.2480558 | 0.4749278971 | -0.01611642045 | 0.02403216 |
| GammaPow49 | Sum_PC | 0.1814202 | 0.81857976 | 0.5 | 0.2216281 | 0.9434774853 | -0.00220567025 | 0.01909122 |
| GammaPow5 | Blocktapping | 0.1872431 | 0.81275689 | 0.5 | 0.2303802 | 0.5293473466 | -0.01403778623 | 0.02187128 |
| GammaPow5 | Sum_PC | 0.1734047 | 0.82659531 | 0.5 | 0.2097818 | 0.7106715270 | 0.00523043227 | 0.01781655 |
| GammaPow50 | Blocktapping | 0.2005972 | 0.79940284 | 0.5 | 0.2509338 | 0.5208324827 | -0.01462949679 | 0.02361136 |
| GammaPow50 | Sum_PC | 0.1882774 | 0.81172263 | 0.5 | 0.2319479 | 0.6865270235 | -0.00787242312 | 0.01920350 |
| GammaPow51 | Blocktapping | 0.2087435 | 0.79125650 | 0.5 | 0.2638127 | 0.4642162912 | -0.01605645021 | 0.02441807 |
| GammaPow51 | Sum_PC | 0.1866362 | 0.81336383 | 0.5 | 0.2294621 | 0.9817040281 | -0.00072261130 | 0.01935425 |
| GammaPow52 | Blocktapping | 0.2161072 | 0.78389277 | 0.5 | 0.2756847 | 0.3976404960 | -0.01964143190 | 0.02525385 |
| GammaPow52 | Sum_PC | 0.1831260 | 0.81687400 | 0.5 | 0.2241790 | 0.7231770886 | 0.00481694896 | 0.01776725 |
| GammaPow53 | Blocktapping | 0.1949932 | 0.80500684 | 0.5 | 0.2422255 | 0.5649127469 | -0.01292390986 | 0.02345875 |
| GammaPow53 | Sum_PC | 0.1826347 | 0.81736528 | 0.5 | 0.2234432 | 0.7654594175 | 0.00380107763 | 0.01868399 |
| GammaPow54 | Blocktapping | 0.2048185 | 0.79518150 | 0.5 | 0.2575745 | 0.4897526631 | -0.01443098578 | 0.02316346 |
| GammaPow54 | Sum_PC | 0.1965569 | 0.80344306 | 0.5 | 0.2446433 | 0.5679560996 | 0.00927149594 | 0.02033149 |
| GammaPow55 | Blocktapping | 0.2373353 | 0.76266467 | 0.5 | 0.3111922 | 0.3037461794 | -0.02707591988 | 0.02833741 |
| GammaPow55 | Sum_PC | 0.1823469 | 0.81765312 | 0.5 | 0.2230125 | 0.7774139298 | 0.00355237572 | 0.01761570 |
| GammaPow56 | Blocktapping | 0.2283813 | 0.77161873 | 0.5 | 0.2959768 | 0.3353390656 | -0.02485804333 | 0.02625258 |
| GammaPow56 | Sum_PC | 0.1827910 | 0.81720899 | 0.5 | 0.2236772 | 0.8596124614 | 0.00223644852 | 0.01804714 |
| GammaPow57 | Blocktapping | 0.2144698 | 0.78553023 | 0.5 | 0.2730255 | 0.4421220493 | -0.01691000074 | 0.02353933 |
| GammaPow57 | Sum_PC | 0.2087211 | 0.79127892 | 0.5 | 0.2637769 | 0.4862621542 | 0.01237294319 | 0.02014494 |
| GammaPow58 | Blocktapping | 0.2137459 | 0.78625412 | 0.5 | 0.2718534 | 0.3821786424 | -0.02105977670 | 0.02451205 |
| GammaPow58 | Sum_PC | 0.1790727 | 0.82092728 | 0.5 | 0.2181347 | 0.7973228894 | 0.00287860551 | 0.01772569 |
| GammaPow59 | Blocktapping | 0.2018590 | 0.79814101 | 0.5 | 0.2529114 | 0.6189230426 | -0.01238075808 | 0.02340108 |
| GammaPow59 | Sum_PC | 0.1996888 | 0.80031117 | 0.5 | 0.2495140 | 0.6568607535 | -0.00850249992 | 0.02154724 |
| GammaPow6 | Blocktapping | 0.2009284 | 0.79907159 | 0.5 | 0.2514523 | 0.4588342148 | -0.01762037150 | 0.02349701 |
| GammaPow6 | Sum_PC | 0.1730683 | 0.82693166 | 0.5 | 0.2092898 | 0.8956197796 | 0.00218968860 | 0.01824999 |
| GammaPow60 | Blocktapping | 0.2091691 | 0.79083091 | 0.5 | 0.2644928 | 0.5068055778 | -0.01702597088 | 0.02465074 |
| GammaPow60 | Sum_PC | 0.1953444 | 0.80465558 | 0.5 | 0.2427677 | 0.6923589083 | -0.00820046182 | 0.02019735 |
| GammaPow61 | Blocktapping | 0.2137164 | 0.78628364 | 0.5 | 0.2718057 | 0.5986665119 | -0.01251916027 | 0.02408555 |
| GammaPow61 | Sum_PC | 0.2366791 | 0.76332093 | 0.5 | 0.3100650 | 0.4072278524 | -0.02133524460 | 0.02427179 |
| GammaPow62 | Blocktapping | 0.2127478 | 0.78725220 | 0.5 | 0.2702410 | 0.6637949741 | -0.01013637100 | 0.02272759 |
| GammaPow62 | Sum_PC | 0.2572088 | 0.74279122 | 0.5 | 0.3462733 | 0.3160505983 | -0.02628466422 | 0.02800034 |
| GammaPow63 | Blocktapping | 0.1993187 | 0.80068134 | 0.5 | 0.2489363 | 0.7497046856 | -0.00796283747 | 0.02187743 |
| GammaPow63 | Sum_PC | 0.2400788 | 0.75992118 | 0.5 | 0.3159259 | 0.3453148868 | -0.02299923414 | 0.02585513 |
| GammaPow64 | Blocktapping | 0.1993333 | 0.80066669 | 0.5 | 0.2489592 | 0.6463207056 | -0.01134661745 | 0.02213568 |
| GammaPow64 | Sum_PC | 0.2291612 | 0.77083881 | 0.5 | 0.2972881 | 0.4032929999 | -0.01853991120 | 0.02428224 |
| GammaPow65 | Blocktapping | 0.2277303 | 0.77226974 | 0.5 | 0.2948844 | 0.4437840605 | -0.01828623979 | 0.02644711 |
| GammaPow65 | Sum_PC | 0.2178526 | 0.78214738 | 0.5 | 0.2785314 | 0.5269388195 | -0.01410490437 | 0.02268960 |
| GammaPow66 | Blocktapping | 0.2132602 | 0.78673976 | 0.5 | 0.2710683 | 0.5947887687 | -0.01323928363 | 0.02370788 |

| DV | Predictor | Sum_PostModProb_Incl | Sum_PostModProb_Excl | Sum_PriorModProb_Incl | BF_Incl | Frequentist_p_value | postMean | postStd |
|---|---|---|---|---|---|---|---|---|
| GammaPow66 | Sum_PC | 0.2324614 | 0.76753860 | 0.5 | 0.3028661 | 0.4167122951 | -0.01965130324 | 0.02386878 |
| GammaPow67 | Blocktapping | 0.2031449 | 0.79685509 | 0.5 | 0.2549333 | 0.4868866114 | -0.01668435658 | 0.02311651 |
| GammaPow67 | Sum_PC | 0.1822187 | 0.81778126 | 0.5 | 0.2228209 | 0.8276900229 | 0.00282376063 | 0.01897640 |
| GammaPow68 | Blocktapping | 0.2021408 | 0.79785919 | 0.5 | 0.2533540 | 0.6257087374 | -0.01222477490 | 0.02336458 |
| GammaPow68 | Sum_PC | 0.2197056 | 0.78029436 | 0.5 | 0.2815676 | 0.4298488270 | -0.01754207400 | 0.02418121 |
| GammaPow69 | Blocktapping | 0.2437027 | 0.75629731 | 0.5 | 0.3222313 | 0.3818506944 | -0.02435015714 | 0.02733454 |
| GammaPow69 | Sum_PC | 0.2151261 | 0.78487386 | 0.5 | 0.2740901 | 0.5731281543 | -0.01177316504 | 0.02147501 |
| GammaPow7 | Blocktapping | 0.1895657 | 0.81043434 | 0.5 | 0.2339063 | 0.9589001613 | -0.00232631368 | 0.02090656 |
| GammaPow7 | Sum_PC | 0.2254655 | 0.77453449 | 0.5 | 0.2910981 | 0.4006452766 | -0.01908108920 | 0.02401105 |
| GammaPow70 | Blocktapping | 0.2187363 | 0.78126367 | 0.5 | 0.2799776 | 0.4346347514 | -0.01969791730 | 0.02436331 |
| GammaPow70 | Sum_PC | 0.1856168 | 0.81438317 | 0.5 | 0.2279232 | 0.9119252708 | -0.00256402856 | 0.01831487 |
| GammaPow71 | Blocktapping | 0.2112010 | 0.78879895 | 0.5 | 0.2677502 | 0.6567215907 | -0.01107149796 | 0.02249848 |
| GammaPow71 | Sum_PC | 0.2378875 | 0.76211247 | 0.5 | 0.3121423 | 0.3985701602 | -0.02108358097 | 0.02398156 |
| GammaPow72 | Blocktapping | 0.2214075 | 0.77859249 | 0.5 | 0.2843689 | 0.5909627697 | -0.01364008590 | 0.02413703 |
| GammaPow72 | Sum_PC | 0.2548523 | 0.74514773 | 0.5 | 0.3420158 | 0.3460768282 | -0.02400756478 | 0.02675574 |
| GammaPow73 | Blocktapping | 0.2035444 | 0.79645560 | 0.5 | 0.2555628 | 0.6360657829 | -0.01115642498 | 0.02396378 |
| GammaPow73 | Sum_PC | 0.2236706 | 0.77632945 | 0.5 | 0.2881129 | 0.4316188017 | -0.01847817011 | 0.02314815 |
| GammaPow74 | Blocktapping | 0.1612912 | 0.83870876 | 0.5 | 0.1923090 | 0.6711904368 | -0.00879895930 | 0.01962224 |
| GammaPow74 | Sum_PC | 0.1529692 | 0.84703077 | 0.5 | 0.1805947 | 0.9154767123 | -0.00250239327 | 0.01729538 |
| GammaPow75 | Blocktapping | 0.1928769 | 0.80712311 | 0.5 | 0.2389684 | 0.6715920821 | -0.01017273299 | 0.02202666 |
| GammaPow75 | Sum_PC | 0.1979123 | 0.80208767 | 0.5 | 0.2467465 | 0.5910586499 | -0.01143517370 | 0.02144076 |
| GammaPow76 | Blocktapping | 0.2264306 | 0.77356936 | 0.5 | 0.2927089 | 0.4702027516 | -0.01826681123 | 0.02628610 |
| GammaPow76 | Sum_PC | 0.2201393 | 0.77986067 | 0.5 | 0.2822803 | 0.5515586828 | -0.01361549232 | 0.02393739 |
| GammaPow77 | Blocktapping | 0.2209800 | 0.77902005 | 0.5 | 0.2836640 | 0.5049758425 | -0.01637087088 | 0.02536934 |
| GammaPow77 | Sum_PC | 0.2142161 | 0.78578394 | 0.5 | 0.2726145 | 0.5091280545 | -0.01418857545 | 0.02262231 |
| GammaPow78 | Blocktapping | 0.2086043 | 0.79139572 | 0.5 | 0.2635904 | 0.7201406057 | -0.00883197838 | 0.02328503 |
| GammaPow78 | Sum_PC | 0.2632872 | 0.73671282 | 0.5 | 0.3573810 | 0.2935487462 | -0.02870707590 | 0.02852718 |
| GammaPow79 | Blocktapping | 0.2776525 | 0.72234749 | 0.5 | 0.3843753 | 0.3167694095 | -0.03104796101 | 0.03113744 |
| GammaPow79 | Sum_PC | 0.2648178 | 0.73518217 | 0.5 | 0.3602071 | 0.3935482494 | -0.02225445468 | 0.02576927 |
| GammaPow8 | Blocktapping | 0.1742640 | 0.82573595 | 0.5 | 0.2110409 | 0.9261554956 | -0.00261114366 | 0.02109250 |
| GammaPow8 | Sum_PC | 0.1724283 | 0.82757167 | 0.5 | 0.2083546 | 0.9921872050 | -0.00083767943 | 0.01770251 |
| GammaPow80 | Blocktapping | 0.2260932 | 0.77390677 | 0.5 | 0.2921453 | 0.4411095230 | -0.01874610528 | 0.02760337 |
| GammaPow80 | Sum_PC | 0.2062340 | 0.79376599 | 0.5 | 0.2598171 | 0.6532620688 | -0.01048887266 | 0.02124221 |
| GammaPow81 | Blocktapping | 0.2199374 | 0.78006261 | 0.5 | 0.2819484 | 0.4822826666 | -0.01721653313 | 0.02529513 |
| GammaPow81 | Sum_PC | 0.2096250 | 0.79037503 | 0.5 | 0.2652222 | 0.5834395690 | -0.01205084508 | 0.02183823 |
| GammaPow82 | Blocktapping | 0.2460275 | 0.75397252 | 0.5 | 0.3263083 | 0.3206200963 | -0.02559297427 | 0.02730544 |
| GammaPow82 | Sum_PC | 0.2140720 | 0.78592798 | 0.5 | 0.2723812 | 0.8939319591 | -0.00375508579 | 0.01999881 |
| GammaPow83 | Blocktapping | 0.3367769 | 0.66322307 | 0.5 | 0.5077883 | 0.2932713653 | -0.03689876268 | 0.03442588 |
| GammaPow83 | Sum_PC | 0.3592166 | 0.64078342 | 0.5 | 0.5605897 | 0.1796660863 | -0.04465538924 | 0.03497727 |
| GammaPow84 | Blocktapping | 0.2886683 | 0.71133174 | 0.5 | 0.4058138 | 0.3725470734 | -0.02639561196 | 0.02927394 |
| GammaPow84 | Sum_PC | 0.3461002 | 0.65389976 | 0.5 | 0.5292864 | 0.1969446251 | -0.04223599498 | 0.03541993 |

| DV | Predictor | Sum_PostModProb_Incl | Sum_PostModProb_Excl | Sum_PriorModProb_Incl | BF_Incl | Frequentist_p_value | postMean | postStd |
|---|---|---|---|---|---|---|---|---|
| GammaPow85 | Blocktapping | 0.3338285 | 0.66617154 | 0.5 | 0.5011149 | 0.3538868781 | -0.02787640493 | 0.03023843 |
| GammaPow85 | Sum_PC | 0.4806742 | 0.51932581 | 0.5 | 0.9255735 | 0.0911265746 | -0.07326733109 | 0.04730033 |
| GammaPow86 | Blocktapping | 0.2059565 | 0.79404351 | 0.5 | 0.2593768 | 0.7290720347 | -0.00835082585 | 0.02183073 |
| GammaPow86 | Sum_PC | 0.2790513 | 0.72094871 | 0.5 | 0.3870612 | 0.2348569029 | -0.03444377575 | 0.02954643 |
| GammaPow87 | Blocktapping | 0.2166257 | 0.78337434 | 0.5 | 0.2765289 | 0.5038148713 | -0.01684600342 | 0.02589142 |
| GammaPow87 | Sum_PC | 0.2117544 | 0.78824558 | 0.5 | 0.2686402 | 0.5435386727 | -0.01445302895 | 0.02196814 |
| GammaPow88 | Blocktapping | 0.2378144 | 0.76218557 | 0.5 | 0.3120164 | 0.5829018991 | -0.01433145185 | 0.02466159 |
| GammaPow88 | Sum_PC | 0.3167171 | 0.68328290 | 0.5 | 0.4635227 | 0.2066072698 | -0.04171931494 | 0.03316071 |
| GammaPow89 | Blocktapping | 0.2005944 | 0.79940558 | 0.5 | 0.2509295 | 0.4982241756 | -0.01556602147 | 0.02300831 |
| GammaPow89 | Sum_PC | 0.1777130 | 0.82228698 | 0.5 | 0.2161204 | 0.8531609394 | -0.00463450301 | 0.01779629 |
| GammaPow9 | Blocktapping | 0.1947098 | 0.80529020 | 0.5 | 0.2417884 | 0.5054944202 | -0.01563184232 | 0.02323350 |
| GammaPow9 | Sum_PC | 0.1743827 | 0.82561735 | 0.5 | 0.2112149 | 0.8829989467 | 0.00180104076 | 0.01819915 |
| GammaPow90 | Blocktapping | 0.2270918 | 0.77290819 | 0.5 | 0.2938147 | 0.4019281757 | -0.02020004847 | 0.02620996 |
| GammaPow90 | Sum_PC | 0.1892490 | 0.81075099 | 0.5 | 0.2334243 | 0.9070952657 | -0.00313636904 | 0.01848772 |
| GammaPow91 | Blocktapping | 0.1970500 | 0.80295000 | 0.5 | 0.2454076 | 0.7402642424 | -0.00895374760 | 0.02375449 |
| GammaPow91 | Sum_PC | 0.1789417 | 0.82105826 | 0.5 | 0.2179404 | 0.8365437394 | -0.00422850366 | 0.01911153 |
| GammaPow92 | Blocktapping | 0.2197467 | 0.78025327 | 0.5 | 0.2816351 | 0.5199620782 | -0.01582887220 | 0.02674775 |
| GammaPow92 | Sum_PC | 0.2162946 | 0.78370537 | 0.5 | 0.2759897 | 0.5267232932 | -0.01386707689 | 0.02357180 |
| GammaPow93 | Blocktapping | 0.1878550 | 0.81214500 | 0.5 | 0.2313072 | 0.7110893567 | -0.00819875913 | 0.02315789 |
| GammaPow93 | Sum_PC | 0.1832191 | 0.81678086 | 0.5 | 0.2243186 | 0.8359621137 | -0.00438494712 | 0.01868564 |
| GammaPow94 | Blocktapping | 0.2810794 | 0.71892058 | 0.5 | 0.3909742 | 0.2937787896 | -0.03271752448 | 0.03192117 |
| GammaPow94 | Sum_PC | 0.2485240 | 0.75147600 | 0.5 | 0.3307145 | 0.4616461022 | -0.01742868896 | 0.02481402 |
| GammaPow95 | Blocktapping | 0.2313298 | 0.76867020 | 0.5 | 0.3009481 | 0.4023340517 | -0.02133410416 | 0.02568247 |
| GammaPow95 | Sum_PC | 0.2331962 | 0.76680379 | 0.5 | 0.3041146 | 0.3771431618 | -0.01998975416 | 0.02477620 |
| GammaPow96 | Blocktapping | 0.2906404 | 0.70935964 | 0.5 | 0.4097222 | 0.3370174167 | -0.02889160199 | 0.03035834 |
| GammaPow96 | Sum_PC | 0.3306282 | 0.66937182 | 0.5 | 0.4939380 | 0.2235539343 | -0.03825598701 | 0.03316200 |
| GammaPow97 | Blocktapping | 0.3101756 | 0.68982441 | 0.5 | 0.4496443 | 0.4054042896 | -0.02303882892 | 0.02811745 |
| GammaPow97 | Sum_PC | 0.4683293 | 0.53167074 | 0.5 | 0.8808633 | 0.0901375059 | -0.07631969981 | 0.04614917 |
| GammaPow98 | Blocktapping | 0.2283865 | 0.77161354 | 0.5 | 0.2959855 | 0.7195754909 | -0.00943466553 | 0.02271505 |
| GammaPow98 | Sum_PC | 0.3289069 | 0.67109306 | 0.5 | 0.4901063 | 0.1836720793 | -0.04493387823 | 0.03452041 |
| GammaPow99 | Blocktapping | 0.2289824 | 0.77101758 | 0.5 | 0.2969873 | 0.3451636585 | -0.02350886721 | 0.02595245 |
| GammaPow99 | Sum_PC | 0.1853952 | 0.81460485 | 0.5 | 0.2275891 | 0.9513292337 | 0.00009337420 | 0.01834663 |
| global_absPower_alpha | Blocktapping | 0.3404952 | 0.65950482 | 0.5 | 0.5162892 | 0.1613862882 | 0.05170038083 | 0.04168718 |
| global_absPower_alpha | Sum_PC | 0.2333687 | 0.76663130 | 0.5 | 0.3044080 | 0.7104695182 | -0.00625006914 | 0.02273884 |
| global_absPower_beta | Blocktapping | 0.2645550 | 0.73544502 | 0.5 | 0.3597210 | 0.3419556136 | 0.02730311761 | 0.03177811 |
| global_absPower_beta | Sum_PC | 0.1941598 | 0.80584022 | 0.5 | 0.2409408 | 0.7906512504 | 0.00533548341 | 0.01905729 |
| global_absPower_gamma | Blocktapping | 0.2708658 | 0.72913415 | 0.5 | 0.3714897 | 0.6479421589 | -0.01010108369 | 0.02262788 |
| global_absPower_gamma | Sum_PC | 0.4910244 | 0.50897563 | 0.5 | 0.9647306 | 0.0703009593 | -0.07865674854 | 0.04776198 |

| DV | Predictor | Sum_PostModProb_Incl | Sum_PostModProb_Excl | Sum_PriorModProb_Incl | BF_Incl | Frequentist_p_value | postMean | postStd |
|---|---|---|---|---|---|---|---|---|
| global_absPower_theta | Blocktapping | 0.1726282 | 0.82737180 | 0.5 | 0.2086465 | 0.6803171800 | 0.00903860832 | 0.02221399 |
| global_absPower_theta | Sum_PC | 0.1659184 | 0.83408160 | 0.5 | 0.1989235 | 0.7798056404 | -0.00466907978 | 0.01976401 |
| global_PAF_Cog | Blocktapping | 0.1301832 | 0.86981680 | 0.5 | 0.1496674 | 0.7684304469 | 0.00455983770 | 0.01695475 |
| global_PAF_Cog | Sum_PC | 0.1275835 | 0.87241650 | 0.5 | 0.1462415 | 0.9940518550 | -0.00016655328 | 0.01511375 |
| PAF1 | Blocktapping | 0.1562681 | 0.84373191 | 0.5 | 0.1852106 | 0.5440304359 | -0.01203277451 | 0.01942141 |
| PAF1 | Sum_PC | 0.1655765 | 0.83442349 | 0.5 | 0.1984322 | 0.4272110441 | 0.01363325677 | 0.01874414 |
| PAF10 | Blocktapping | 0.1963103 | 0.80368970 | 0.5 | 0.2442613 | 0.9839222514 | 0.00073410720 | 0.01960791 |
| PAF10 | Sum_PC | 0.3370336 | 0.66296641 | 0.5 | 0.5083721 | 0.1178263103 | 0.05588417841 | 0.03707934 |
| PAF100 | Blocktapping | 0.1736155 | 0.82638454 | 0.5 | 0.2100904 | 0.4098669598 | 0.01614889703 | 0.02310644 |
| PAF100 | Sum_PC | 0.1474753 | 0.85252466 | 0.5 | 0.1729866 | 0.8044139927 | 0.00404374821 | 0.01687176 |
| PAF11 | Blocktapping | 0.1810985 | 0.81890153 | 0.5 | 0.2211480 | 0.9289867601 | 0.00374001174 | 0.01966618 |
| PAF11 | Sum_PC | 0.2665825 | 0.73341748 | 0.5 | 0.3634799 | 0.2135854657 | 0.03643606101 | 0.03066016 |
| PAF12 | Blocktapping | 0.2449599 | 0.75504005 | 0.5 | 0.3244330 | 0.7000640722 | -0.00764899212 | 0.02140408 |
| PAF12 | Sum_PC | 0.4638702 | 0.53612985 | 0.5 | 0.8652198 | 0.0636781611 | 0.08416632471 | 0.04823963 |
| PAF13 | Blocktapping | 0.1922355 | 0.80776447 | 0.5 | 0.2379846 | 0.8993985563 | -0.00207816797 | 0.02125574 |
| PAF13 | Sum_PC | 0.2557418 | 0.74425818 | 0.5 | 0.3436198 | 0.2779630747 | 0.02904219529 | 0.02965257 |
| PAF14 | Blocktapping | 0.1846542 | 0.81534580 | 0.5 | 0.2264735 | 0.6163151046 | 0.00999612115 | 0.02295228 |
| PAF14 | Sum_PC | 0.1791098 | 0.82089022 | 0.5 | 0.2181897 | 0.7336115748 | 0.00604302766 | 0.02044278 |
| PAF15 | Blocktapping | 0.2002372 | 0.79976284 | 0.5 | 0.2503707 | 0.4279065142 | 0.01735266045 | 0.02424771 |
| PAF15 | Sum_PC | 0.1798564 | 0.82014361 | 0.5 | 0.2192987 | 0.6532377529 | -0.00746683459 | 0.01911678 |
| PAF16 | Blocktapping | 0.1736063 | 0.82639373 | 0.5 | 0.2100769 | 0.4616497997 | -0.01400136259 | 0.02056533 |
| PAF16 | Sum_PC | 0.1765471 | 0.82345289 | 0.5 | 0.2143986 | 0.4059110637 | 0.01435282107 | 0.01904586 |
| PAF17 | Blocktapping | 0.1629204 | 0.83707964 | 0.5 | 0.1946295 | 0.9624719441 | 0.00002729954 | 0.02078038 |
| PAF17 | Sum_PC | 0.1673569 | 0.83264308 | 0.5 | 0.2009948 | 0.7358518628 | 0.00622462327 | 0.01985380 |
| PAF18 | Blocktapping | 0.1758555 | 0.82414448 | 0.5 | 0.2133795 | 0.4977579965 | 0.01320301175 | 0.02181436 |
| PAF18 | Sum_PC | 0.1573763 | 0.84262373 | 0.5 | 0.1867693 | 0.8082205330 | -0.00385399492 | 0.01793679 |
| PAF19 | Blocktapping | 0.1800412 | 0.81995876 | 0.5 | 0.2195735 | 0.6167622063 | 0.01097810252 | 0.02310908 |
| PAF19 | Sum_PC | 0.1749192 | 0.82508076 | 0.5 | 0.2120026 | 0.7206311176 | 0.00695126290 | 0.02015933 |
| PAF2 | Blocktapping | 0.1382346 | 0.86176539 | 0.5 | 0.1604086 | 0.6630278206 | -0.00729098149 | 0.01690568 |
| PAF2 | Sum_PC | 0.1381569 | 0.86184305 | 0.5 | 0.1603041 | 0.6136243534 | 0.00620539134 | 0.01587800 |
| PAF20 | Blocktapping | 0.2000384 | 0.79996158 | 0.5 | 0.2500600 | 0.3439438341 | 0.02154390241 | 0.02477126 |
| PAF20 | Sum_PC | 0.1673652 | 0.83263476 | 0.5 | 0.2010068 | 0.7283688157 | -0.00586547299 | 0.01800597 |
| PAF21 | Blocktapping | 0.1895480 | 0.81045200 | 0.5 | 0.2338794 | 0.6138900740 | 0.00952386958 | 0.02286100 |
| PAF21 | Sum_PC | 0.2049963 | 0.79500366 | 0.5 | 0.2578558 | 0.4715437367 | -0.01524596042 | 0.02235172 |
| PAF22 | Blocktapping | 0.2006024 | 0.79939755 | 0.5 | 0.2509420 | 0.8524818563 | -0.00304352331 | 0.02260471 |
| PAF22 | Sum_PC | 0.2648140 | 0.73518604 | 0.5 | 0.3601999 | 0.2811988414 | 0.02936390188 | 0.02916479 |
| PAF23 | Blocktapping | 0.1873950 | 0.81260497 | 0.5 | 0.2306102 | 0.7506316304 | 0.00678924283 | 0.02391127 |
| PAF23 | Sum_PC | 0.1807500 | 0.81925003 | 0.5 | 0.2206286 | 0.9879725361 | 0.00104447518 | 0.02055145 |
| PAF24 | Blocktapping | 0.1594768 | 0.84052320 | 0.5 | 0.1897352 | 0.8591612767 | 0.00342225686 | 0.02092182 |
| PAF24 | Sum_PC | 0.1588428 | 0.84115718 | 0.5 | 0.1888384 | 0.8523923573 | 0.00388572309 | 0.01874357 |

| DV | Predictor | Sum_PostModProb_Incl | Sum_PostModProb_Excl | Sum_PriorModProb_Incl | BF_Incl | Frequentist_p_value | postMean | postStd |
|---|---|---|---|---|---|---|---|---|
| PAF25 | Blocktapping | 0.2536636 | 0.74633637 | 0.5 | 0.3398784 | 0.5307942422 | -0.01265060680 | 0.02336063 |
| PAF25 | Sum_PC | 0.4441590 | 0.55584099 | 0.5 | 0.7990757 | 0.0696621666 | 0.07594715201 | 0.04646896 |
| PAF26 | Blocktapping | 0.2436267 | 0.75637327 | 0.5 | 0.3220985 | 0.5473557747 | -0.01206558612 | 0.02463419 |
| PAF26 | Sum_PC | 0.3858910 | 0.61410899 | 0.5 | 0.6283755 | 0.1015045971 | 0.06232995385 | 0.04176361 |
| PAF27 | Blocktapping | 0.1968329 | 0.80316709 | 0.5 | 0.2450709 | 0.6531019536 | -0.01047602282 | 0.02530021 |
| PAF27 | Sum_PC | 0.1863708 | 0.81362918 | 0.5 | 0.2290611 | 0.9702700999 | -0.00106845328 | 0.02061654 |
| PAF28 | Blocktapping | 0.1939643 | 0.80603572 | 0.5 | 0.2406398 | 0.6775055112 | 0.00998775023 | 0.02423217 |
| PAF28 | Sum_PC | 0.1926139 | 0.80738614 | 0.5 | 0.2385647 | 0.6994778456 | 0.00853882768 | 0.02164374 |
| PAF29 | Blocktapping | 0.1992148 | 0.80078520 | 0.5 | 0.2487743 | 0.4063054616 | 0.01898210792 | 0.02549255 |
| PAF29 | Sum_PC | 0.1678978 | 0.83210216 | 0.5 | 0.2017755 | 0.9560657907 | 0.00086570908 | 0.01840204 |
| PAF3 | Blocktapping | 0.1408412 | 0.85915884 | 0.5 | 0.1639291 | 0.6657154719 | -0.00705130171 | 0.01761296 |
| PAF3 | Sum_PC | 0.1400566 | 0.85994337 | 0.5 | 0.1628673 | 0.6340592174 | 0.00629060542 | 0.01596930 |
| PAF30 | Blocktapping | 0.2216872 | 0.77831278 | 0.5 | 0.2848305 | 0.4905248898 | 0.01699683743 | 0.02740718 |
| PAF30 | Sum_PC | 0.2068349 | 0.79316508 | 0.5 | 0.2607716 | 0.7316674687 | 0.00700473075 | 0.02211472 |
| PAF31 | Blocktapping | 0.2272053 | 0.77279473 | 0.5 | 0.2940047 | 0.4493085144 | -0.01596530080 | 0.02491578 |
| PAF31 | Sum_PC | 0.2806653 | 0.71933473 | 0.5 | 0.3901734 | 0.2132339916 | 0.03476164845 | 0.03037826 |
| PAF32 | Blocktapping | 0.2181551 | 0.78184494 | 0.5 | 0.2790260 | 0.3350178476 | -0.02133406692 | 0.02461480 |
| PAF32 | Sum_PC | 0.2389921 | 0.76100791 | 0.5 | 0.3140468 | 0.2473003452 | 0.02737969980 | 0.02572676 |
| PAF33 | Blocktapping | 0.1754292 | 0.82457082 | 0.5 | 0.2127521 | 0.4665308307 | -0.01403485966 | 0.02044163 |
| PAF33 | Sum_PC | 0.2110963 | 0.78890368 | 0.5 | 0.2675819 | 0.2646758391 | 0.02378668652 | 0.02260115 |
| PAF34 | Blocktapping | 0.1970165 | 0.80298352 | 0.5 | 0.2453556 | 0.4826304755 | 0.01609140276 | 0.02318437 |
| PAF34 | Sum_PC | 0.1919236 | 0.80807636 | 0.5 | 0.2375068 | 0.5277233895 | 0.01266720424 | 0.02170839 |
| PAF35 | Blocktapping | 0.2041667 | 0.79583328 | 0.5 | 0.2565446 | 0.5387201772 | -0.01411770259 | 0.02542096 |
| PAF35 | Sum_PC | 0.1975439 | 0.80245612 | 0.5 | 0.2461741 | 0.5838816070 | 0.01142527675 | 0.02169629 |
| PAF36 | Blocktapping | 0.1435178 | 0.85648219 | 0.5 | 0.1675666 | 0.9618291572 | 0.00061702176 | 0.01813470 |
| PAF36 | Sum_PC | 0.1418305 | 0.85816954 | 0.5 | 0.1652709 | 0.8850630629 | -0.00186957740 | 0.01659925 |
| PAF37 | Blocktapping | 0.1916472 | 0.80835284 | 0.5 | 0.2370835 | 0.4998043720 | 0.01523190309 | 0.02360706 |
| PAF37 | Sum_PC | 0.1865638 | 0.81343619 | 0.5 | 0.2293527 | 0.5650333309 | 0.01102597963 | 0.02089263 |
| PAF38 | Blocktapping | 0.1860792 | 0.81392080 | 0.5 | 0.2286208 | 0.6298223698 | -0.00884519380 | 0.02048716 |
| PAF38 | Sum_PC | 0.2629340 | 0.73706602 | 0.5 | 0.3567306 | 0.1937142709 | 0.03568387758 | 0.02980280 |
| PAF39 | Blocktapping | 0.1600636 | 0.83993642 | 0.5 | 0.1905663 | 0.8271689951 | -0.00335213979 | 0.01812941 |
| PAF39 | Sum_PC | 0.2328722 | 0.76712780 | 0.5 | 0.3035638 | 0.2157422231 | 0.03031914488 | 0.02773410 |
| PAF4 | Blocktapping | 0.1515734 | 0.84842661 | 0.5 | 0.1786523 | 0.5810703353 | -0.01009641898 | 0.01868960 |
| PAF4 | Sum_PC | 0.1517139 | 0.84828605 | 0.5 | 0.1788476 | 0.5568424896 | 0.00821141376 | 0.01717138 |
| PAF40 | Blocktapping | 0.1536516 | 0.84634844 | 0.5 | 0.1815465 | 0.7211404877 | -0.00511779359 | 0.01839879 |
| PAF40 | Sum_PC | 0.1837230 | 0.81627696 | 0.5 | 0.2250744 | 0.3699877932 | 0.01720282886 | 0.02124808 |
| PAF41 | Blocktapping | 0.1426360 | 0.85736402 | 0.5 | 0.1663657 | 0.9502209625 | -0.00123292720 | 0.01796848 |
| PAF41 | Sum_PC | 0.1492606 | 0.85073939 | 0.5 | 0.1754481 | 0.6144062769 | 0.00791099983 | 0.01763742 |
| PAF42 | Blocktapping | 0.2596300 | 0.74036996 | 0.5 | 0.3506761 | 0.3574269675 | -0.02207497388 | 0.02617963 |
| PAF42 | Sum_PC | 0.3516291 | 0.64837087 | 0.5 | 0.5423272 | 0.1501677383 | 0.04772423887 | 0.03703795 |
| PAF43 | Blocktapping | 0.2287571 | 0.77124289 | 0.5 | 0.2966084 | 0.4073955893 | -0.01978166099 | 0.02613780 |

| DV | Predictor | Sum_PostModProb_Incl | Sum_PostModProb_Excl | Sum_PriorModProb_Incl | BF_Incl | Frequentist_p_value | postMean | postStd |
|---|---|---|---|---|---|---|---|---|
| PAF43 | Sum_PC | 0.2375915 | 0.76240853 | 0.5 | 0.3116328 | 0.3468953698 | 0.02170843714 | 0.02543878 |
| PAF44 | Blocktapping | 0.2181632 | 0.78183677 | 0.5 | 0.2790394 | 0.5254530132 | -0.01439055690 | 0.02547028 |
| PAF44 | Sum_PC | 0.2489803 | 0.75101974 | 0.5 | 0.3315229 | 0.3251677457 | 0.02236921129 | 0.02702528 |
| PAF45 | Blocktapping | 0.1858557 | 0.81414429 | 0.5 | 0.2282835 | 0.7282042628 | -0.00731081016 | 0.02355216 |
| PAF45 | Sum_PC | 0.1795356 | 0.82046437 | 0.5 | 0.2188220 | 0.9153902742 | 0.00186082909 | 0.01977913 |
| PAF46 | Blocktapping | 0.1839746 | 0.81602544 | 0.5 | 0.2254520 | 0.8693435722 | 0.00375670665 | 0.02334295 |
| PAF46 | Sum_PC | 0.1863906 | 0.81360938 | 0.5 | 0.2290910 | 0.7622459467 | 0.00609107601 | 0.02142038 |
| PAF47 | Blocktapping | 0.1867360 | 0.81326405 | 0.5 | 0.2296129 | 0.8466383426 | -0.00413975191 | 0.02311679 |
| PAF47 | Sum_PC | 0.1871394 | 0.81286063 | 0.5 | 0.2302232 | 0.8731751277 | 0.00305654940 | 0.02102035 |
| PAF48 | Blocktapping | 0.1960622 | 0.80393781 | 0.5 | 0.2438773 | 0.6777700265 | 0.00816250396 | 0.02396016 |
| PAF48 | Sum_PC | 0.1908822 | 0.80911785 | 0.5 | 0.2359139 | 0.8197481853 | -0.00433420945 | 0.02168820 |
| PAF49 | Blocktapping | 0.1392667 | 0.86073326 | 0.5 | 0.1618001 | 0.9930967066 | 0.00044966703 | 0.01828463 |
| PAF49 | Sum_PC | 0.1403004 | 0.85969963 | 0.5 | 0.1631970 | 0.7135610921 | 0.00553476089 | 0.01664142 |
| PAF5 | Blocktapping | 0.1424323 | 0.85756768 | 0.5 | 0.1660887 | 0.6073335264 | -0.00878838825 | 0.01809403 |
| PAF5 | Sum_PC | 0.1419091 | 0.85809088 | 0.5 | 0.1653777 | 0.5715450213 | 0.00884816288 | 0.01606161 |
| PAF50 | Blocktapping | 0.1598923 | 0.84010773 | 0.5 | 0.1903235 | 0.5419545113 | 0.01131129839 | 0.01999109 |
| PAF50 | Sum_PC | 0.1476919 | 0.85230805 | 0.5 | 0.1732847 | 0.9490233442 | 0.00041961026 | 0.01709008 |
| PAF51 | Blocktapping | 0.1444251 | 0.85557492 | 0.5 | 0.1688047 | 0.5815571636 | -0.00857228757 | 0.01741356 |
| PAF51 | Sum_PC | 0.1647334 | 0.83526657 | 0.5 | 0.1972226 | 0.3585339727 | 0.01486379237 | 0.01806063 |
| PAF52 | Blocktapping | 0.1388733 | 0.86112667 | 0.5 | 0.1612693 | 0.6103735470 | -0.00883688021 | 0.01804393 |
| PAF52 | Sum_PC | 0.1389767 | 0.86102334 | 0.5 | 0.1614087 | 0.5756672608 | 0.00844298343 | 0.01644073 |
| PAF53 | Blocktapping | 0.1218414 | 0.87815857 | 0.5 | 0.1387465 | 0.9243894704 | -0.00153212495 | 0.01602037 |
| PAF53 | Sum_PC | 0.1230876 | 0.87691240 | 0.5 | 0.1403648 | 0.9536479130 | 0.00036129527 | 0.01506755 |
| PAF54 | Blocktapping | 0.1430328 | 0.85696720 | 0.5 | 0.1669058 | 0.6302008895 | -0.00805125159 | 0.01935184 |
| PAF54 | Sum_PC | 0.1337960 | 0.86620404 | 0.5 | 0.1544624 | 0.7736141676 | 0.00339754928 | 0.01578624 |
| PAF55 | Blocktapping | 0.1513538 | 0.84864616 | 0.5 | 0.1783474 | 0.5280205404 | -0.01103437592 | 0.01931392 |
| PAF55 | Sum_PC | 0.1452197 | 0.85478031 | 0.5 | 0.1698912 | 0.5683434817 | 0.00867912493 | 0.01644311 |
| PAF56 | Blocktapping | 0.1436799 | 0.85632012 | 0.5 | 0.1677876 | 0.7105224295 | -0.00583058788 | 0.01825138 |
| PAF56 | Sum_PC | 0.1589890 | 0.84101098 | 0.5 | 0.1890451 | 0.4445566695 | 0.01263447382 | 0.01874592 |
| PAF57 | Blocktapping | 0.1284279 | 0.87157210 | 0.5 | 0.1473520 | 0.7889111381 | -0.00460748527 | 0.01671101 |
| PAF57 | Sum_PC | 0.1302370 | 0.86976301 | 0.5 | 0.1497385 | 0.7161087857 | 0.00552360044 | 0.01499728 |
| PAF58 | Blocktapping | 0.1438203 | 0.85617966 | 0.5 | 0.1679792 | 0.5494810515 | -0.01118550608 | 0.01884462 |
| PAF58 | Sum_PC | 0.1319593 | 0.86804073 | 0.5 | 0.1520197 | 0.7627095678 | 0.00341080294 | 0.01520167 |
| PAF59 | Blocktapping | 0.1572355 | 0.84276446 | 0.5 | 0.1865712 | 0.6361735474 | -0.00761499727 | 0.01913609 |
| PAF59 | Sum_PC | 0.1851902 | 0.81480985 | 0.5 | 0.2272802 | 0.3552741650 | 0.01737126497 | 0.02144027 |
| PAF6 | Blocktapping | 0.1412147 | 0.85878534 | 0.5 | 0.1644353 | 0.7471247434 | -0.00525660769 | 0.01817622 |
| PAF6 | Sum_PC | 0.1478845 | 0.85211549 | 0.5 | 0.1735498 | 0.5641675902 | 0.00918767577 | 0.01699848 |
| PAF60 | Blocktapping | 0.1708673 | 0.82913270 | 0.5 | 0.2060796 | 0.8688137225 | -0.00256415658 | 0.01880886 |
| PAF60 | Sum_PC | 0.2541316 | 0.74586843 | 0.5 | 0.3407190 | 0.2014272709 | 0.03416768582 | 0.02923665 |
| PAF61 | Blocktapping | 0.1840254 | 0.81597460 | 0.5 | 0.2255283 | 0.5015974226 | -0.01358383636 | 0.02306816 |
| PAF61 | Sum_PC | 0.1884668 | 0.81153323 | 0.5 | 0.2322354 | 0.4763320284 | 0.01349272074 | 0.02101014 |

| DV | Predictor | Sum_PostModProb_Incl | Sum_PostModProb_Excl | Sum_PriorModProb_Incl | BF_Incl | Frequentist_p_value | postMean | postStd |
|---|---|---|---|---|---|---|---|---|
| PAF62 | Blocktapping | 0.1693197 | 0.83068029 | 0.5 | 0.2038326 | 0.7839055281 | -0.00545226262 | 0.02070971 |
| PAF62 | Sum_PC | 0.1986590 | 0.80134099 | 0.5 | 0.2479082 | 0.4174012904 | 0.01692874097 | 0.02348165 |
| PAF63 | Blocktapping | 0.1807943 | 0.81920570 | 0.5 | 0.2206946 | 0.9684296866 | 0.00150091801 | 0.02086015 |
| PAF63 | Sum_PC | 0.2421660 | 0.75783400 | 0.5 | 0.3195502 | 0.2795072871 | 0.02820197958 | 0.02713607 |
| PAF64 | Blocktapping | 0.1709577 | 0.82904225 | 0.5 | 0.2062111 | 0.7247627095 | 0.00685936673 | 0.02063601 |
| PAF64 | Sum_PC | 0.1953058 | 0.80469416 | 0.5 | 0.2427082 | 0.4310036909 | 0.01646318181 | 0.02154661 |
| PAF65 | Blocktapping | 0.1660716 | 0.83392836 | 0.5 | 0.1991438 | 0.5722356299 | 0.01107825335 | 0.02151026 |
| PAF65 | Sum_PC | 0.1641874 | 0.83581262 | 0.5 | 0.1964404 | 0.6323170367 | 0.00798288972 | 0.01905371 |
| PAF66 | Blocktapping | 0.1970870 | 0.80291297 | 0.5 | 0.2454650 | 0.3878163880 | 0.01977866092 | 0.02402972 |
| PAF66 | Sum_PC | 0.1738678 | 0.82613216 | 0.5 | 0.2104601 | 0.8132043745 | -0.00320325371 | 0.01933738 |
| PAF67 | Blocktapping | 0.1279064 | 0.87209361 | 0.5 | 0.1466659 | 0.9185498099 | -0.00151836035 | 0.01663048 |
| PAF67 | Sum_PC | 0.1268941 | 0.87310594 | 0.5 | 0.1453364 | 0.8618898964 | 0.00274384683 | 0.01493964 |
| PAF68 | Blocktapping | 0.1618761 | 0.83812388 | 0.5 | 0.1931410 | 0.9221075494 | -0.00148587193 | 0.01806106 |
| PAF68 | Sum_PC | 0.2380169 | 0.76198309 | 0.5 | 0.3123651 | 0.3012531234 | 0.02653557881 | 0.02718415 |
| PAF69 | Blocktapping | 0.1595035 | 0.84049653 | 0.5 | 0.1897729 | 0.8561472214 | 0.00401247937 | 0.02007920 |
| PAF69 | Sum_PC | 0.1826690 | 0.81733095 | 0.5 | 0.2234946 | 0.4569357365 | 0.01393869351 | 0.02091476 |
| PAF7 | Blocktapping | 0.1430786 | 0.85692136 | 0.5 | 0.1669682 | 0.6423648464 | -0.00837785018 | 0.01824324 |
| PAF7 | Sum_PC | 0.1363910 | 0.86360897 | 0.5 | 0.1579315 | 0.7511549265 | 0.00385579012 | 0.01535014 |
| PAF70 | Blocktapping | 0.1481330 | 0.85186701 | 0.5 | 0.1738921 | 0.6508418297 | 0.00786463046 | 0.01946295 |
| PAF70 | Sum_PC | 0.1393703 | 0.86062974 | 0.5 | 0.1619399 | 0.9115087434 | 0.00144580985 | 0.01627066 |
| PAF71 | Blocktapping | 0.1783500 | 0.82164998 | 0.5 | 0.2170633 | 0.4699658188 | 0.01499945779 | 0.02193045 |
| PAF71 | Sum_PC | 0.1615718 | 0.83842823 | 0.5 | 0.1927079 | 0.7318893025 | -0.00610751978 | 0.01800604 |
| PAF72 | Blocktapping | 0.1932818 | 0.80671819 | 0.5 | 0.2395902 | 0.7580859329 | -0.00529534641 | 0.02191230 |
| PAF72 | Sum_PC | 0.2616285 | 0.73837150 | 0.5 | 0.3543318 | 0.2284432619 | 0.03189749460 | 0.02981154 |
| PAF73 | Blocktapping | 0.1946051 | 0.80539490 | 0.5 | 0.2416269 | 0.6396353858 | 0.01149372895 | 0.02351097 |
| PAF73 | Sum_PC | 0.2011427 | 0.79885726 | 0.5 | 0.2517881 | 0.5029793269 | 0.01504745706 | 0.02217160 |
| PAF74 | Blocktapping | 0.1337495 | 0.86625053 | 0.5 | 0.1544004 | 0.9110951026 | -0.00142934786 | 0.01698327 |
| PAF74 | Sum_PC | 0.1539827 | 0.84601726 | 0.5 | 0.1820090 | 0.4719334763 | 0.01267533898 | 0.01819769 |
| PAF75 | Blocktapping | 0.1668514 | 0.83314865 | 0.5 | 0.2002660 | 0.6659129310 | -0.00918650326 | 0.02093166 |
| PAF75 | Sum_PC | 0.2062153 | 0.79378470 | 0.5 | 0.2597874 | 0.3131587799 | 0.02146416408 | 0.02292006 |
| PAF76 | Blocktapping | 0.1979295 | 0.80207049 | 0.5 | 0.2467732 | 0.4985598919 | -0.01371337255 | 0.02179704 |
| PAF76 | Sum_PC | 0.2563755 | 0.74362454 | 0.5 | 0.3447647 | 0.2059271728 | 0.03237924738 | 0.02812817 |
| PAF77 | Blocktapping | 0.2055791 | 0.79442092 | 0.5 | 0.2587785 | 0.3655024059 | 0.02215278536 | 0.02599835 |
| PAF77 | Sum_PC | 0.1718633 | 0.82813671 | 0.5 | 0.2075301 | 0.7104134729 | 0.00703134583 | 0.01828004 |
| PAF78 | Blocktapping | 0.2228444 | 0.77715561 | 0.5 | 0.2867436 | 0.5116175094 | 0.01676481962 | 0.02751050 |
| PAF78 | Sum_PC | 0.2021780 | 0.79782200 | 0.5 | 0.2534124 | 0.6969231621 | 0.00792769458 | 0.02175849 |
| PAF79 | Blocktapping | 0.2327681 | 0.76723191 | 0.5 | 0.3033869 | 0.3682299886 | -0.02235031976 | 0.02669795 |
| PAF79 | Sum_PC | 0.2461278 | 0.75387221 | 0.5 | 0.3264848 | 0.2922267225 | 0.02426157731 | 0.02657478 |
| PAF8 | Blocktapping | 0.1291259 | 0.87087409 | 0.5 | 0.1482716 | 0.9611217395 | -0.00046785053 | 0.01571830 |
| PAF8 | Sum_PC | 0.1432024 | 0.85679759 | 0.5 | 0.1671368 | 0.5538981987 | 0.00949576395 | 0.01675314 |
| PAF80 | Blocktapping | 0.1935114 | 0.80648862 | 0.5 | 0.2399431 | 0.4356611137 | -0.01415082766 | 0.02125840 |

| DV | Predictor | Sum_PostModProb_Incl | Sum_PostModProb_Excl | Sum_PriorModProb_Incl | BF_Incl | Frequentist_p_value | postMean | postStd |
|---|---|---|---|---|---|---|---|---|
| PAF80 | Sum_PC | 0.2538739 | 0.74612611 | 0.5 | 0.3402560 | 0.1795087718 | 0.03287722380 | 0.02767371 |
| PAF81 | Blocktapping | 0.1758020 | 0.82419801 | 0.5 | 0.2133007 | 0.8172485681 | -0.00329015364 | 0.01964243 |
| PAF81 | Sum_PC | 0.2475572 | 0.75244280 | 0.5 | 0.3290047 | 0.2167390781 | 0.03183740469 | 0.02857771 |
| PAF82 | Blocktapping | 0.1413105 | 0.85868951 | 0.5 | 0.1645653 | 0.8491108641 | -0.00328032139 | 0.01820752 |
| PAF82 | Sum_PC | 0.1488149 | 0.85118512 | 0.5 | 0.1748326 | 0.6242673098 | 0.00752968400 | 0.01762641 |
| PAF83 | Blocktapping | 0.1945732 | 0.80542677 | 0.5 | 0.2415778 | 0.5892297144 | -0.01337610657 | 0.02446612 |
| PAF83 | Sum_PC | 0.1876535 | 0.81234652 | 0.5 | 0.2310018 | 0.6555435823 | 0.00849134131 | 0.02059174 |
| PAF84 | Blocktapping | 0.2101266 | 0.78987339 | 0.5 | 0.2660257 | 0.4509840840 | -0.01858622165 | 0.02543452 |
| PAF84 | Sum_PC | 0.2030657 | 0.79693432 | 0.5 | 0.2548086 | 0.4847337714 | 0.01397522962 | 0.02222318 |
| PAF85 | Blocktapping | 0.2078005 | 0.79219950 | 0.5 | 0.2623083 | 0.5060495886 | -0.01630962807 | 0.02648403 |
| PAF85 | Sum_PC | 0.1939813 | 0.80601868 | 0.5 | 0.2406660 | 0.6960082914 | 0.00733674497 | 0.02134836 |
| PAF86 | Blocktapping | 0.1971190 | 0.80288103 | 0.5 | 0.2455145 | 0.9932987973 | -0.00001742507 | 0.02322724 |
| PAF86 | Sum_PC | 0.2389273 | 0.76107273 | 0.5 | 0.3139349 | 0.3799656163 | 0.02244927585 | 0.02627017 |
| PAF87 | Blocktapping | 0.1954216 | 0.80457841 | 0.5 | 0.2428869 | 0.4469375334 | 0.01706921071 | 0.02411364 |
| PAF87 | Sum_PC | 0.1905174 | 0.80948264 | 0.5 | 0.2353569 | 0.4900600206 | 0.01439596518 | 0.02124429 |
| PAF88 | Blocktapping | 0.1845732 | 0.81542685 | 0.5 | 0.2263516 | 0.9168047771 | -0.00226535051 | 0.02332254 |
| PAF88 | Sum_PC | 0.1960227 | 0.80397732 | 0.5 | 0.2438162 | 0.6336893721 | 0.00943142036 | 0.02253775 |
| PAF89 | Blocktapping | 0.1460097 | 0.85399032 | 0.5 | 0.1709735 | 0.5629282372 | 0.01022533007 | 0.01828977 |
| PAF89 | Sum_PC | 0.1335230 | 0.86647697 | 0.5 | 0.1540988 | 0.9365926170 | -0.00118055965 | 0.01511776 |
| PAF9 | Blocktapping | 0.1423041 | 0.85769588 | 0.5 | 0.1659144 | 0.6883938332 | -0.00660514109 | 0.01768158 |
| PAF9 | Sum_PC | 0.1427527 | 0.85724729 | 0.5 | 0.1665245 | 0.6352664169 | 0.00620155647 | 0.01632615 |
| PAF90 | Blocktapping | 0.1574464 | 0.84255362 | 0.5 | 0.1868681 | 0.7767382739 | -0.00455125840 | 0.01833237 |
| PAF90 | Sum_PC | 0.1992273 | 0.80077273 | 0.5 | 0.2487938 | 0.3080535541 | 0.02260388274 | 0.02261426 |
| PAF91 | Blocktapping | 0.1386413 | 0.86135868 | 0.5 | 0.1609565 | 0.6319904105 | -0.00769390251 | 0.01797398 |
| PAF91 | Sum_PC | 0.1452947 | 0.85470531 | 0.5 | 0.1699939 | 0.5553923770 | 0.00926305101 | 0.01683659 |
| PAF92 | Blocktapping | 0.1800101 | 0.81998993 | 0.5 | 0.2195272 | 0.4699345024 | -0.01340765682 | 0.02028870 |
| PAF92 | Sum_PC | 0.2208293 | 0.77917065 | 0.5 | 0.2834159 | 0.2626367795 | 0.02483083289 | 0.02398397 |
| PAF93 | Blocktapping | 0.1420199 | 0.85798007 | 0.5 | 0.1655282 | 0.6683620922 | -0.00686316436 | 0.01835015 |
| PAF93 | Sum_PC | 0.1551808 | 0.84481920 | 0.5 | 0.1836852 | 0.4672389317 | 0.01200575794 | 0.01807057 |
| PAF94 | Blocktapping | 0.2240978 | 0.77590225 | 0.5 | 0.2888221 | 0.3349856366 | -0.02303121617 | 0.02535312 |
| PAF94 | Sum_PC | 0.2400717 | 0.75992826 | 0.5 | 0.3159137 | 0.2564492106 | 0.02633386814 | 0.02509120 |
| PAF95 | Blocktapping | 0.2178447 | 0.78215534 | 0.5 | 0.2785184 | 0.3740975996 | -0.02020847573 | 0.02501404 |
| PAF95 | Sum_PC | 0.2479518 | 0.75204815 | 0.5 | 0.3297021 | 0.2366659183 | 0.02903108039 | 0.02605256 |
| PAF96 | Blocktapping | 0.2245247 | 0.77547530 | 0.5 | 0.2895317 | 0.4593926083 | -0.01683025910 | 0.02573762 |
| PAF96 | Sum_PC | 0.2432194 | 0.75678057 | 0.5 | 0.3213870 | 0.3491710959 | 0.02193291141 | 0.02573241 |
| PAF97 | Blocktapping | 0.1983491 | 0.80165094 | 0.5 | 0.2474257 | 0.6782243744 | -0.00905428239 | 0.02385105 |
| PAF97 | Sum_PC | 0.2073512 | 0.79264880 | 0.5 | 0.2615928 | 0.5575587365 | 0.01313288509 | 0.02365157 |
| PAF98 | Blocktapping | 0.1810032 | 0.81899675 | 0.5 | 0.2210061 | 0.9855925719 | -0.00039309284 | 0.02302399 |
| PAF98 | Sum_PC | 0.1974684 | 0.80253157 | 0.5 | 0.2460569 | 0.8059461966 | 0.00512994083 | 0.02314313 |
| PAF99 | Blocktapping | 0.1375446 | 0.86245538 | 0.5 | 0.1594803 | 0.9660304195 | -0.00132428380 | 0.01695721 |
| PAF99 | Sum_PC | 0.1533165 | 0.84668352 | 0.5 | 0.1810789 | 0.5366642068 | 0.01023586266 | 0.01750537 |

| DV | Predictor | Sum_PostModProb_Incl | Sum_PostModProb_Excl | Sum_PriorModProb_Incl | BF_Incl | Frequentist_p_value | postMean | postStd |
|---|---|---|---|---|---|---|---|---|
| theta_ROI1_AECnodestrength | Blocktapping | 0.2147996 | 0.78520041 | 0.5 | 0.2735602 | 0.4894464970 | -0.01682342458 | 0.02627610 |
| theta_ROI1_AECnodestrength | Sum_PC | 0.2041215 | 0.79587849 | 0.5 | 0.2564732 | 0.6151976816 | 0.00964074973 | 0.02273114 |
| theta_ROI10_AECnodestrength | Blocktapping | 0.1996466 | 0.80035345 | 0.5 | 0.2494480 | 0.7019976241 | -0.00786546761 | 0.02373741 |
| theta_ROI10_AECnodestrength | Sum_PC | 0.2168316 | 0.78316835 | 0.5 | 0.2768647 | 0.4791582582 | 0.01341633727 | 0.02331925 |
| theta_ROI100_AECnodestrength | Blocktapping | 0.2750323 | 0.72496771 | 0.5 | 0.3793718 | 0.2741000054 | -0.03384255298 | 0.03281440 |
| theta_ROI100_AECnodestrength | Sum_PC | 0.2235518 | 0.77644816 | 0.5 | 0.2879160 | 0.5477569888 | 0.01101948327 | 0.02293292 |
| theta_ROI11_AECnodestrength | Blocktapping | 0.2266897 | 0.77331033 | 0.5 | 0.2931419 | 0.5145012129 | -0.01536836880 | 0.02686381 |
| theta_ROI11_AECnodestrength | Sum_PC | 0.2288152 | 0.77118479 | 0.5 | 0.2967061 | 0.5204907998 | 0.01402079805 | 0.02479783 |
| theta_ROI12_AECnodestrength | Blocktapping | 0.1983491 | 0.80165094 | 0.5 | 0.2474257 | 0.6760696168 | -0.00824869938 | 0.02427826 |
| theta_ROI12_AECnodestrength | Sum_PC | 0.2243176 | 0.77568243 | 0.5 | 0.2891874 | 0.3985575399 | 0.01905398245 | 0.02544755 |
| theta_ROI13_AECnodestrength | Blocktapping | 0.1807847 | 0.81921534 | 0.5 | 0.2206803 | 0.7864882255 | -0.00486540421 | 0.02184736 |
| theta_ROI13_AECnodestrength | Sum_PC | 0.2016636 | 0.79833639 | 0.5 | 0.2526048 | 0.4908420878 | 0.01339585208 | 0.02387909 |
| theta_ROI14_AECnodestrength | Blocktapping | 0.2884808 | 0.71151923 | 0.5 | 0.4054434 | 0.2366808687 | -0.03699702775 | 0.03320494 |
| theta_ROI14_AECnodestrength | Sum_PC | 0.2432124 | 0.75678756 | 0.5 | 0.3213748 | 0.4105257934 | 0.01821620277 | 0.02409045 |
| theta_ROI15_AECnodestrength | Blocktapping | 0.3000730 | 0.69992696 | 0.5 | 0.4287205 | 0.2109771881 | -0.04091721237 | 0.03532567 |
| theta_ROI15_AECnodestrength | Sum_PC | 0.2408541 | 0.75914590 | 0.5 | 0.3172698 | 0.4438470065 | 0.01529404906 | 0.02345691 |
| theta_ROI16_AECnodestrength | Blocktapping | 0.2321183 | 0.76788171 | 0.5 | 0.3022839 | 0.4078401339 | -0.02110293095 | 0.02770807 |
| theta_ROI16_AECnodestrength | Sum_PC | 0.2093978 | 0.79060223 | 0.5 | 0.2648586 | 0.6254554189 | 0.00973953153 | 0.02211129 |
| theta_ROI17_AECnodestrength | Blocktapping | 0.2037823 | 0.79621765 | 0.5 | 0.2559380 | 0.5058368246 | -0.01674973866 | 0.02603738 |
| theta_ROI17_AECnodestrength | Sum_PC | 0.1823713 | 0.81762867 | 0.5 | 0.2230491 | 0.9725198767 | -0.00150419242 | 0.02054586 |
| theta_ROI18_AECnodestrength | Blocktapping | 0.2681741 | 0.73182592 | 0.5 | 0.3664452 | 0.3135340250 | -0.02867758511 | 0.03149810 |
| theta_ROI18_AECnodestrength | Sum_PC | 0.2428266 | 0.75717344 | 0.5 | 0.3207014 | 0.4069479437 | 0.01813718707 | 0.02475747 |
| theta_ROI19_AECnodestrength | Blocktapping | 0.2768305 | 0.72316951 | 0.5 | 0.3828017 | 0.2601124197 | -0.03355875222 | 0.03094750 |
| theta_ROI19_AECnodestrength | Sum_PC | 0.2460925 | 0.75390747 | 0.5 | 0.3264227 | 0.3828955207 | 0.01833502362 | 0.02441808 |
| theta_ROI2_AECnodestrength | Blocktapping | 0.2203647 | 0.77963530 | 0.5 | 0.2826510 | 0.3783990553 | -0.02172740662 | 0.02843877 |
| theta_ROI2_AECnodestrength | Sum_PC | 0.1822572 | 0.81774281 | 0.5 | 0.2228784 | 0.9052962538 | 0.00142182898 | 0.01982887 |
| theta_ROI20_AECnodestrength | Blocktapping | 0.2527204 | 0.74727955 | 0.5 | 0.3381873 | 0.3538944067 | -0.02536838845 | 0.02895484 |
| theta_ROI20_AECnodestrength | Sum_PC | 0.2526272 | 0.74737281 | 0.5 | 0.3380203 | 0.3415082355 | 0.02246230952 | 0.02679084 |
| theta_ROI21_AECnodestrength | Blocktapping | 0.2501205 | 0.74987950 | 0.5 | 0.3335476 | 0.3460926342 | -0.02510881838 | 0.02991903 |

| DV | Predictor | Sum_PostModProb_Incl | Sum_PostModProb_Excl | Sum_PriorModProb_Incl | BF_Incl | Frequentist_p_value | postMean | postStd |
|---|---|---|---|---|---|---|---|---|
| theta_ROI21_AECnodestrength | Sum_PC | 0.2317504 | 0.76824964 | 0.5 | 0.3016602 | 0.4599613386 | 0.01548346148 | 0.02435756 |
| theta_ROI22_AECnodestrength | Blocktapping | 0.2007202 | 0.79927977 | 0.5 | 0.2511264 | 0.8121590740 | -0.00420630467 | 0.02252667 |
| theta_ROI22_AECnodestrength | Sum_PC | 0.2681287 | 0.73187130 | 0.5 | 0.3663605 | 0.2540345448 | 0.03126498988 | 0.03104732 |
| theta_ROI23_AECnodestrength | Blocktapping | 0.2465560 | 0.75344395 | 0.5 | 0.3272387 | 0.4348822830 | -0.01951785059 | 0.02709907 |
| theta_ROI23_AECnodestrength | Sum_PC | 0.2871630 | 0.71283699 | 0.5 | 0.4028453 | 0.2418806109 | 0.03155845391 | 0.02974735 |
| theta_ROI24_AECnodestrength | Blocktapping | 0.1944679 | 0.80553213 | 0.5 | 0.2414154 | 0.6297252397 | -0.01159547467 | 0.02414422 |
| theta_ROI24_AECnodestrength | Sum_PC | 0.1835326 | 0.81646743 | 0.5 | 0.2247886 | 0.7758425946 | 0.00549623811 | 0.02104058 |
| theta_ROI25_AECnodestrength | Blocktapping | 0.2073537 | 0.79264629 | 0.5 | 0.2615968 | 0.5700739105 | -0.01262324122 | 0.02409205 |
| theta_ROI25_AECnodestrength | Sum_PC | 0.2226787 | 0.77732131 | 0.5 | 0.2864693 | 0.4141288454 | 0.01784027391 | 0.02549460 |
| theta_ROI26_AECnodestrength | Blocktapping | 0.2077314 | 0.79226861 | 0.5 | 0.2621982 | 0.7936500732 | -0.00575562563 | 0.02308407 |
| theta_ROI26_AECnodestrength | Sum_PC | 0.2715754 | 0.72842461 | 0.5 | 0.3728257 | 0.2574561940 | 0.02907841561 | 0.03085786 |
| theta_ROI27_AECnodestrength | Blocktapping | 0.2576464 | 0.74235363 | 0.5 | 0.3470669 | 0.4167552231 | -0.01995752362 | 0.02732956 |
| theta_ROI27_AECnodestrength | Sum_PC | 0.3259843 | 0.67401573 | 0.5 | 0.4836449 | 0.1767303286 | 0.04097435865 | 0.03544161 |
| theta_ROI28_AECnodestrength | Blocktapping | 0.2192776 | 0.78072236 | 0.5 | 0.2808651 | 0.3974942778 | -0.01990077666 | 0.02604679 |
| theta_ROI28_AECnodestrength | Sum_PC | 0.2407218 | 0.75927816 | 0.5 | 0.3170404 | 0.3580983632 | 0.02060908697 | 0.02554917 |
| theta_ROI29_AECnodestrength | Blocktapping | 0.2456377 | 0.75436231 | 0.5 | 0.3256230 | 0.3525951370 | -0.02588087102 | 0.03010069 |
| theta_ROI29_AECnodestrength | Sum_PC | 0.2122771 | 0.78772287 | 0.5 | 0.2694820 | 0.5609488635 | 0.01142451332 | 0.02281086 |
| theta_ROI3_AECnodestrength | Blocktapping | 0.2391902 | 0.76080975 | 0.5 | 0.3143890 | 0.4098423394 | -0.02142283695 | 0.02892560 |
| theta_ROI3_AECnodestrength | Sum_PC | 0.2220752 | 0.77792477 | 0.5 | 0.2854713 | 0.5322148611 | 0.01314105437 | 0.02379826 |
| theta_ROI30_AECnodestrength | Blocktapping | 0.2213144 | 0.77868564 | 0.5 | 0.2842153 | 0.4545688636 | -0.01707848601 | 0.02492346 |
| theta_ROI30_AECnodestrength | Sum_PC | 0.2233049 | 0.77669509 | 0.5 | 0.2875065 | 0.4341767619 | 0.01559118789 | 0.02345107 |
| theta_ROI31_AECnodestrength | Blocktapping | 0.2370755 | 0.76292448 | 0.5 | 0.3107457 | 0.6566445983 | -0.00953906603 | 0.02417363 |
| theta_ROI31_AECnodestrength | Sum_PC | 0.3496152 | 0.65038477 | 0.5 | 0.5375514 | 0.1496156463 | 0.04942093296 | 0.03659776 |
| theta_ROI32_AECnodestrength | Blocktapping | 0.1905195 | 0.80948054 | 0.5 | 0.2353602 | 0.7021744760 | -0.00808628566 | 0.02333688 |
| theta_ROI32_AECnodestrength | Sum_PC | 0.2023084 | 0.79769155 | 0.5 | 0.2536174 | 0.5353581188 | 0.01332996539 | 0.02283094 |
| theta_ROI33_AECnodestrength | Blocktapping | 0.2035049 | 0.79649510 | 0.5 | 0.2555005 | 0.6600648036 | -0.01090846313 | 0.02444781 |
| theta_ROI33_AECnodestrength | Sum_PC | 0.2193099 | 0.78069007 | 0.5 | 0.2809180 | 0.4603244611 | 0.01675762984 | 0.02475457 |
| theta_ROI34_AECnodestrength | Blocktapping | 0.5146805 | 0.48531954 | 0.5 | 1.0604981 | 0.2126402621 | -0.07097520028 | 0.06012834 |
| theta_ROI34_AECnodestrength | Sum_PC | 0.1711583 | 0.82884168 | 0.5 | 0.2065030 | 0.4721258020 | 0.00883275272 | 0.01403443 |

| DV | Predictor | Sum_PostModProb_Incl | Sum_PostModProb_Excl | Sum_PriorModProb_Incl | BF_Incl | Frequentist_p_value | postMean | postStd |
|---|---|---|---|---|---|---|---|---|
| theta_ROI35_AECnodestrength | Blocktapping | 0.2207710 | 0.77922900 | 0.5 | 0.2833198 | 0.7652127442 | -0.00630369237 | 0.02348503 |
| theta_ROI35_AECnodestrength | Sum_PC | 0.3204765 | 0.67952353 | 0.5 | 0.4716194 | 0.1832973908 | 0.04231716799 | 0.03467938 |
| theta_ROI36_AECnodestrength | Blocktapping | 0.1799661 | 0.82003395 | 0.5 | 0.2194617 | 0.3481140951 | -0.01860015822 | 0.02252648 |
| theta_ROI36_AECnodestrength | Sum_PC | 0.1681259 | 0.83187412 | 0.5 | 0.2021050 | 0.4148871335 | 0.01337624975 | 0.01914906 |
| theta_ROI37_AECnodestrength | Blocktapping | 0.2164757 | 0.78352431 | 0.5 | 0.2762846 | 0.5111851771 | -0.01414161367 | 0.02615434 |
| theta_ROI37_AECnodestrength | Sum_PC | 0.2179770 | 0.78202304 | 0.5 | 0.2787347 | 0.4622628075 | 0.01607149938 | 0.02394719 |
| theta_ROI38_AECnodestrength | Blocktapping | 0.2002275 | 0.79977249 | 0.5 | 0.2503556 | 0.7266784211 | -0.00694411133 | 0.02360738 |
| theta_ROI38_AECnodestrength | Sum_PC | 0.2373773 | 0.76262271 | 0.5 | 0.3112644 | 0.3590899522 | 0.02211591475 | 0.02640351 |
| theta_ROI39_AECnodestrength | Blocktapping | 0.1969342 | 0.80306575 | 0.5 | 0.2452280 | 0.7373993162 | -0.00724339140 | 0.02369771 |
| theta_ROI39_AECnodestrength | Sum_PC | 0.2127033 | 0.78729671 | 0.5 | 0.2701692 | 0.4935728657 | 0.01487062487 | 0.02384854 |
| theta_ROI4_AECnodestrength | Blocktapping | 0.2138977 | 0.78610227 | 0.5 | 0.2720991 | 0.5042362417 | -0.01585145256 | 0.02523943 |
| theta_ROI4_AECnodestrength | Sum_PC | 0.2118024 | 0.78819761 | 0.5 | 0.2687174 | 0.5163649862 | 0.01244614462 | 0.02341930 |
| theta_ROI40_AECnodestrength | Blocktapping | 0.1931751 | 0.80682485 | 0.5 | 0.2394264 | 0.6593554582 | -0.01007083662 | 0.02467893 |
| theta_ROI40_AECnodestrength | Sum_PC | 0.1889323 | 0.81106766 | 0.5 | 0.2329428 | 0.7420059018 | 0.00563157817 | 0.02028360 |
| theta_ROI41_AECnodestrength | Blocktapping | 0.2631627 | 0.73683733 | 0.5 | 0.3571516 | 0.3020715036 | -0.02923843690 | 0.03272017 |
| theta_ROI41_AECnodestrength | Sum_PC | 0.2122472 | 0.78775279 | 0.5 | 0.2694338 | 0.6876529187 | 0.00739675699 | 0.02177972 |
| theta_ROI42_AECnodestrength | Blocktapping | 0.2198572 | 0.78014279 | 0.5 | 0.2818166 | 0.5776830740 | -0.01262376255 | 0.02462810 |
| theta_ROI42_AECnodestrength | Sum_PC | 0.2640375 | 0.73596253 | 0.5 | 0.3587648 | 0.3067302785 | 0.02702465187 | 0.02928278 |
| theta_ROI43_AECnodestrength | Blocktapping | 0.2101708 | 0.78982924 | 0.5 | 0.2660965 | 0.7261742570 | -0.00730720446 | 0.02366499 |
| theta_ROI43_AECnodestrength | Sum_PC | 0.2612344 | 0.73876563 | 0.5 | 0.3536093 | 0.2981003542 | 0.02892903177 | 0.02954077 |
| theta_ROI44_AECnodestrength | Blocktapping | 0.2557796 | 0.74422043 | 0.5 | 0.3436879 | 0.4325367860 | -0.01666126366 | 0.02625207 |
| theta_ROI44_AECnodestrength | Sum_PC | 0.3456356 | 0.65436444 | 0.5 | 0.5282004 | 0.1454656955 | 0.04793599541 | 0.03609313 |
| theta_ROI45_AECnodestrength | Blocktapping | 0.1849227 | 0.81507729 | 0.5 | 0.2268775 | 0.8414201579 | -0.00347676945 | 0.02233479 |
| theta_ROI45_AECnodestrength | Sum_PC | 0.2241031 | 0.77589691 | 0.5 | 0.2888310 | 0.4183311506 | 0.01925117968 | 0.02559061 |
| theta_ROI46_AECnodestrength | Blocktapping | 0.2966257 | 0.70337432 | 0.5 | 0.4217181 | 0.2145251841 | -0.03880536092 | 0.03291490 |
| theta_ROI46_AECnodestrength | Sum_PC | 0.2672080 | 0.73279203 | 0.5 | 0.3646437 | 0.2811448980 | 0.02511930848 | 0.02607384 |
| theta_ROI47_AECnodestrength | Blocktapping | 0.2940655 | 0.70593449 | 0.5 | 0.4165620 | 0.4141826618 | -0.01893094573 | 0.02783142 |
| theta_ROI47_AECnodestrength | Sum_PC | 0.4494462 | 0.55055376 | 0.5 | 0.8163530 | 0.0802764192 | 0.07185881990 | 0.04591695 |
| theta_ROI48_AECnodestrength | Blocktapping | 0.2488428 | 0.75115720 | 0.5 | 0.3312793 | 0.3893794754 | -0.02156546467 | 0.02766538 |

| DV | Predictor | Sum_PostModProb_Incl | Sum_PostModProb_Excl | Sum_PriorModProb_Incl | BF_Incl | Frequentist_p_value | postMean | postStd |
|---|---|---|---|---|---|---|---|---|
| theta_ROI48_AECnodestrength | Sum_PC | 0.2710522 | 0.72894780 | 0.5 | 0.3718403 | 0.2668661046 | 0.02751074732 | 0.02748596 |
| theta_ROI49_AECnodestrength | Blocktapping | 0.2509370 | 0.74906301 | 0.5 | 0.3350012 | 0.3619577021 | -0.02342370290 | 0.02986461 |
| theta_ROI49_AECnodestrength | Sum_PC | 0.2276670 | 0.77233297 | 0.5 | 0.2947783 | 0.4946667613 | 0.01359692899 | 0.02418202 |
| theta_ROI5_AECnodestrength | Blocktapping | 0.2212925 | 0.77870752 | 0.5 | 0.2841792 | 0.4044711605 | -0.02231660966 | 0.02795910 |
| theta_ROI5_AECnodestrength | Sum_PC | 0.1986475 | 0.80135245 | 0.5 | 0.2478904 | 0.5781271723 | 0.01052790505 | 0.02152133 |
| theta_ROI50_AECnodestrength | Blocktapping | 0.2429574 | 0.75704262 | 0.5 | 0.3209296 | 0.3528407112 | -0.02484666664 | 0.02991427 |
| theta_ROI50_AECnodestrength | Sum_PC | 0.2028573 | 0.79714266 | 0.5 | 0.2544806 | 0.7332387089 | 0.00603145164 | 0.02117542 |
| theta_ROI51_AECnodestrength | Blocktapping | 0.2345387 | 0.76546130 | 0.5 | 0.3064018 | 0.4123835250 | -0.02077753178 | 0.02748750 |
| theta_ROI51_AECnodestrength | Sum_PC | 0.2279581 | 0.77204193 | 0.5 | 0.2952664 | 0.4472333828 | 0.01571782605 | 0.02525931 |
| theta_ROI52_AECnodestrength | Blocktapping | 0.2727678 | 0.72723224 | 0.5 | 0.3750765 | 0.2566910861 | -0.03420917351 | 0.03289119 |
| theta_ROI52_AECnodestrength | Sum_PC | 0.2183222 | 0.78167776 | 0.5 | 0.2792995 | 0.5954134663 | 0.00951682664 | 0.02183821 |
| theta_ROI53_AECnodestrength | Blocktapping | 0.2163769 | 0.78362308 | 0.5 | 0.2761237 | 0.5173191614 | -0.01610448976 | 0.02615166 |
| theta_ROI53_AECnodestrength | Sum_PC | 0.2004000 | 0.79959999 | 0.5 | 0.2506253 | 0.7180608082 | 0.00602916523 | 0.02143441 |
| theta_ROI54_AECnodestrength | Blocktapping | 0.2866895 | 0.71331049 | 0.5 | 0.4019141 | 0.2805220839 | -0.03452014705 | 0.03290600 |
| theta_ROI54_AECnodestrength | Sum_PC | 0.2481051 | 0.75189488 | 0.5 | 0.3299732 | 0.3952441724 | 0.01889368378 | 0.02476493 |
| theta_ROI55_AECnodestrength | Blocktapping | 0.3474769 | 0.65252306 | 0.5 | 0.5325129 | 0.1585460387 | -0.05289725165 | 0.04201311 |
| theta_ROI55_AECnodestrength | Sum_PC | 0.2397786 | 0.76022142 | 0.5 | 0.3154062 | 0.6388639703 | 0.00942243177 | 0.02207472 |
| theta_ROI56_AECnodestrength | Blocktapping | 0.2982367 | 0.70176333 | 0.5 | 0.4249818 | 0.2298258286 | -0.03913634535 | 0.03438515 |
| theta_ROI56_AECnodestrength | Sum_PC | 0.2371927 | 0.76280729 | 0.5 | 0.3109471 | 0.5198558856 | 0.01242202590 | 0.02397293 |
| theta_ROI57_AECnodestrength | Blocktapping | 0.3078367 | 0.69216328 | 0.5 | 0.4447458 | 0.2081404726 | -0.04247928931 | 0.03826558 |
| theta_ROI57_AECnodestrength | Sum_PC | 0.2222166 | 0.77778336 | 0.5 | 0.2857051 | 0.7425301553 | 0.00635794943 | 0.02180366 |
| theta_ROI58_AECnodestrength | Blocktapping | 0.3262891 | 0.67371092 | 0.5 | 0.4843162 | 0.1705953957 | -0.04949934995 | 0.03947525 |
| theta_ROI58_AECnodestrength | Sum_PC | 0.2362691 | 0.76373085 | 0.5 | 0.3093618 | 0.5464001381 | 0.01083923979 | 0.02270084 |
| theta_ROI59_AECnodestrength | Blocktapping | 0.1974499 | 0.80255014 | 0.5 | 0.2460281 | 0.9775781308 | 0.00014859595 | 0.02286141 |
| theta_ROI59_AECnodestrength | Sum_PC | 0.2414059 | 0.75859409 | 0.5 | 0.3182280 | 0.3578311329 | 0.02267382883 | 0.02817785 |
| theta_ROI6_AECnodestrength | Blocktapping | 0.2449977 | 0.75500234 | 0.5 | 0.3244992 | 0.3349107359 | -0.02714698201 | 0.03107647 |
| theta_ROI6_AECnodestrength | Sum_PC | 0.2098710 | 0.79012896 | 0.5 | 0.2656162 | 0.5655190890 | 0.00979632064 | 0.02197058 |
| theta_ROI60_AECnodestrength | Blocktapping | 0.2033832 | 0.79661680 | 0.5 | 0.2553087 | 0.6881963608 | -0.00818821622 | 0.02356312 |
| theta_ROI60_AECnodestrength | Sum_PC | 0.2244249 | 0.77557513 | 0.5 | 0.2893657 | 0.4350637161 | 0.01686685882 | 0.02522442 |

| DV | Predictor | Sum_PostModProb_Incl | Sum_PostModProb_Excl | Sum_PriorModProb_Incl | BF_Incl | Frequentist_p_value | postMean | postStd |
|---|---|---|---|---|---|---|---|---|
| theta_ROI61_AECnodestrength | Blocktapping | 0.2103448 | 0.78965524 | 0.5 | 0.2663754 | 0.8217687357 | 0.00521198966 | 0.02333463 |
| theta_ROI61_AECnodestrength | Sum_PC | 0.2767769 | 0.72322306 | 0.5 | 0.3826993 | 0.2587470897 | 0.03156489099 | 0.03108964 |
| theta_ROI62_AECnodestrength | Blocktapping | 0.2506071 | 0.74939293 | 0.5 | 0.3344134 | 0.9203479491 | 0.00152257497 | 0.02307482 |
| theta_ROI62_AECnodestrength | Sum_PC | 0.4646870 | 0.53531301 | 0.5 | 0.8680659 | 0.0738666117 | 0.08196353755 | 0.04978689 |
| theta_ROI63_AECnodestrength | Blocktapping | 0.2517431 | 0.74825693 | 0.5 | 0.3364393 | 0.5058948465 | -0.01679940420 | 0.02644489 |
| theta_ROI63_AECnodestrength | Sum_PC | 0.3284216 | 0.67157837 | 0.5 | 0.4890295 | 0.1657303451 | 0.04395968806 | 0.03552667 |
| theta_ROI64_AECnodestrength | Blocktapping | 0.3576025 | 0.64239747 | 0.5 | 0.5566686 | 0.1552543265 | -0.05225323772 | 0.04063065 |
| theta_ROI64_AECnodestrength | Sum_PC | 0.2844071 | 0.71559285 | 0.5 | 0.3974427 | 0.3235703921 | 0.02289805499 | 0.02652213 |
| theta_ROI65_AECnodestrength | Blocktapping | 0.3495301 | 0.65046990 | 0.5 | 0.5373502 | 0.1619015546 | -0.05111630133 | 0.03890607 |
| theta_ROI65_AECnodestrength | Sum_PC | 0.2924901 | 0.70750990 | 0.5 | 0.4134078 | 0.2961384293 | 0.02551698426 | 0.02825053 |
| theta_ROI66_AECnodestrength | Blocktapping | 0.3550590 | 0.64494097 | 0.5 | 0.5505295 | 0.1505439873 | -0.05391422666 | 0.04140220 |
| theta_ROI66_AECnodestrength | Sum_PC | 0.2571696 | 0.74283037 | 0.5 | 0.3462024 | 0.4531762283 | 0.01495793838 | 0.02341690 |
| theta_ROI67_AECnodestrength | Blocktapping | 0.2614987 | 0.73850128 | 0.5 | 0.3540938 | 0.3154223424 | -0.02901115065 | 0.03248409 |
| theta_ROI67_AECnodestrength | Sum_PC | 0.2198346 | 0.78016536 | 0.5 | 0.2817796 | 0.5943912384 | 0.01024958206 | 0.02242179 |
| theta_ROI68_AECnodestrength | Blocktapping | 0.2220151 | 0.77798487 | 0.5 | 0.2853720 | 0.5077220214 | -0.01542844182 | 0.02560887 |
| theta_ROI68_AECnodestrength | Sum_PC | 0.2427794 | 0.75722056 | 0.5 | 0.3206192 | 0.3315522008 | 0.02306089764 | 0.02571923 |
| theta_ROI69_AECnodestrength | Blocktapping | 0.4649442 | 0.53505576 | 0.5 | 0.8689641 | 0.1041334721 | -0.06984380992 | 0.04679275 |
| theta_ROI69_AECnodestrength | Sum_PC | 0.4370695 | 0.56293054 | 0.5 | 0.7764181 | 0.1312942478 | 0.05150732415 | 0.03827648 |
| theta_ROI7_AECnodestrength | Blocktapping | 0.2740246 | 0.72597535 | 0.5 | 0.3774572 | 0.2650308499 | -0.03453151899 | 0.03339092 |
| theta_ROI7_AECnodestrength | Sum_PC | 0.2191028 | 0.78089722 | 0.5 | 0.2805783 | 0.6715473182 | 0.00725634738 | 0.02249600 |
| theta_ROI70_AECnodestrength | Blocktapping | 0.3369998 | 0.66300018 | 0.5 | 0.5082952 | 0.1816675845 | -0.04620885451 | 0.03675813 |
| theta_ROI70_AECnodestrength | Sum_PC | 0.2828984 | 0.71710156 | 0.5 | 0.3945026 | 0.3236141694 | 0.02322968543 | 0.02717080 |
| theta_ROI71_AECnodestrength | Blocktapping | 0.3393262 | 0.66067382 | 0.5 | 0.5136062 | 0.1570747322 | -0.04906228339 | 0.03783472 |
| theta_ROI71_AECnodestrength | Sum_PC | 0.3020399 | 0.69796015 | 0.5 | 0.4327466 | 0.2327822495 | 0.03102725830 | 0.02903487 |
| theta_ROI72_AECnodestrength | Blocktapping | 0.2714018 | 0.72859823 | 0.5 | 0.3724985 | 0.5202396010 | -0.01420257041 | 0.02468646 |
| theta_ROI72_AECnodestrength | Sum_PC | 0.4461000 | 0.55390001 | 0.5 | 0.8053800 | 0.0814647384 | 0.07365968136 | 0.04604453 |
| theta_ROI73_AECnodestrength | Blocktapping | 0.2379518 | 0.76204821 | 0.5 | 0.3122529 | 0.4330440984 | -0.01820924058 | 0.02634609 |
| theta_ROI73_AECnodestrength | Sum_PC | 0.2715414 | 0.72845856 | 0.5 | 0.3727617 | 0.2612652307 | 0.02940382211 | 0.02871024 |
| theta_ROI74_AECnodestrength | Blocktapping | 0.2074820 | 0.79251798 | 0.5 | 0.2618010 | 0.6968021074 | -0.00882602978 | 0.02491053 |

| DV | Predictor | Sum_PostModProb_Incl | Sum_PostModProb_Excl | Sum_PriorModProb_Incl | BF_Incl | Frequentist_p_value | postMean | postStd |
|---|---|---|---|---|---|---|---|---|
| theta_ROI74_AECnodestrength | Sum_PC | 0.2195519 | 0.78044812 | 0.5 | 0.2813151 | 0.4843645650 | 0.01591813801 | 0.02433267 |
| theta_ROI75_AECnodestrength | Blocktapping | 0.2016068 | 0.79839317 | 0.5 | 0.2525157 | 0.6321274482 | -0.00970181606 | 0.02364567 |
| theta_ROI75_AECnodestrength | Sum_PC | 0.2321356 | 0.76786443 | 0.5 | 0.3023132 | 0.3734896387 | 0.02051709954 | 0.02589907 |
| theta_ROI76_AECnodestrength | Blocktapping | 0.2233139 | 0.77668607 | 0.5 | 0.2875215 | 0.5912156023 | -0.01184032275 | 0.02442347 |
| theta_ROI76_AECnodestrength | Sum_PC | 0.2736356 | 0.72636435 | 0.5 | 0.3767195 | 0.2607866499 | 0.03114882239 | 0.03092117 |
| theta_ROI77_AECnodestrength | Blocktapping | 0.3022066 | 0.69779335 | 0.5 | 0.4330890 | 0.2177934886 | -0.03970949041 | 0.03541669 |
| theta_ROI77_AECnodestrength | Sum_PC | 0.2362988 | 0.76370124 | 0.5 | 0.3094126 | 0.5152547741 | 0.01247015326 | 0.02339606 |
| theta_ROI78_AECnodestrength | Blocktapping | 0.2102002 | 0.78979981 | 0.5 | 0.2661436 | 0.3738002074 | -0.02021863472 | 0.02435492 |
| theta_ROI78_AECnodestrength | Sum_PC | 0.2615553 | 0.73844473 | 0.5 | 0.3541975 | 0.5715098897 | 0.01420023526 | 0.02912849 |
| theta_ROI79_AECnodestrength | Blocktapping | 0.4140623 | 0.58593774 | 0.5 | 0.7066660 | 0.1156057120 | -0.06512925361 | 0.04641441 |
| theta_ROI79_AECnodestrength | Sum_PC | 0.3111775 | 0.68882251 | 0.5 | 0.4517528 | 0.2643780239 | 0.02751387405 | 0.02788012 |
| theta_ROI8_AECnodestrength | Blocktapping | 0.3387506 | 0.66124943 | 0.5 | 0.5122886 | 0.1748740521 | -0.04670085281 | 0.03825212 |
| theta_ROI8_AECnodestrength | Sum_PC | 0.2730006 | 0.72699940 | 0.5 | 0.3755170 | 0.3820884509 | 0.01979079420 | 0.02696011 |
| theta_ROI80_AECnodestrength | Blocktapping | 0.2237127 | 0.77628732 | 0.5 | 0.2881828 | 0.4873750901 | -0.01778526200 | 0.02737117 |
| theta_ROI80_AECnodestrength | Sum_PC | 0.2219312 | 0.77806884 | 0.5 | 0.2852333 | 0.4844127553 | 0.01577081955 | 0.02336912 |
| theta_ROI81_AECnodestrength | Blocktapping | 0.2698386 | 0.73016145 | 0.5 | 0.3695601 | 0.2685753295 | -0.03030835175 | 0.03181291 |
| theta_ROI81_AECnodestrength | Sum_PC | 0.2582046 | 0.74179544 | 0.5 | 0.3480805 | 0.2982077473 | 0.02440004969 | 0.02619477 |
| theta_ROI82_AECnodestrength | Blocktapping | 0.3255416 | 0.67445839 | 0.5 | 0.4826712 | 0.1841610153 | -0.04580631608 | 0.03717255 |
| theta_ROI82_AECnodestrength | Sum_PC | 0.2442866 | 0.75571344 | 0.5 | 0.3232529 | 0.5183713826 | 0.01240444333 | 0.02350620 |
| theta_ROI83_AECnodestrength | Blocktapping | 0.3242080 | 0.67579203 | 0.5 | 0.4797452 | 0.3450651144 | -0.02570924828 | 0.03016763 |
| theta_ROI83_AECnodestrength | Sum_PC | 0.4731870 | 0.52681297 | 0.5 | 0.8982069 | 0.0756314937 | 0.07705015817 | 0.04730048 |
| theta_ROI84_AECnodestrength | Blocktapping | 0.3427407 | 0.65725930 | 0.5 | 0.5214695 | 0.3596074468 | -0.02349395275 | 0.02996569 |
| theta_ROI84_AECnodestrength | Sum_PC | 0.5488794 | 0.45112057 | 0.5 | 1.2167023 | 0.0467225863 | 0.09702663345 | 0.05369822 |
| theta_ROI85_AECnodestrength | Blocktapping | 0.3432038 | 0.65679620 | 0.5 | 0.5225423 | 0.2015663125 | -0.04066666305 | 0.03644790 |
| theta_ROI85_AECnodestrength | Sum_PC | 0.3586067 | 0.64139333 | 0.5 | 0.5591057 | 0.1569717463 | 0.04364407722 | 0.03416443 |
| theta_ROI86_AECnodestrength | Blocktapping | 0.2579590 | 0.74204099 | 0.5 | 0.3476344 | 0.4203758931 | -0.01939892197 | 0.02651316 |
| theta_ROI86_AECnodestrength | Sum_PC | 0.3402190 | 0.65978101 | 0.5 | 0.5156544 | 0.1558989110 | 0.04684507014 | 0.03579690 |
| theta_ROI87_AECnodestrength | Blocktapping | 0.2553840 | 0.74461598 | 0.5 | 0.3429741 | 0.3546950085 | -0.02697757433 | 0.03075660 |
| theta_ROI87_AECnodestrength | Sum_PC | 0.2298009 | 0.77019913 | 0.5 | 0.2983655 | 0.5062041059 | 0.01392704827 | 0.02383446 |

| DV | Predictor | Sum_PostModProb_Incl | Sum_PostModProb_Excl | Sum_PriorModProb_Incl | BF_Incl | Frequentist_p_value | postMean | postStd |
|---|---|---|---|---|---|---|---|---|
| theta_ROI88_AECnodestrength | Blocktapping | 0.2200429 | 0.77995707 | 0.5 | 0.2821218 | 0.4247354309 | -0.01974745436 | 0.02567661 |
| theta_ROI88_AECnodestrength | Sum_PC | 0.2272717 | 0.77272825 | 0.5 | 0.2941160 | 0.3609564430 | 0.02026963323 | 0.02479530 |
| theta_ROI89_AECnodestrength | Blocktapping | 0.2468699 | 0.75313006 | 0.5 | 0.3277919 | 0.3641725887 | -0.02483805788 | 0.02889298 |
| theta_ROI89_AECnodestrength | Sum_PC | 0.2183881 | 0.78161186 | 0.5 | 0.2794074 | 0.5151704739 | 0.01297273807 | 0.02265512 |
| theta_ROI9_AECnodestrength | Blocktapping | 0.3678131 | 0.63218691 | 0.5 | 0.5818107 | 0.1304460369 | -0.05999725095 | 0.04302530 |
| theta_ROI9_AECnodestrength | Sum_PC | 0.2586154 | 0.74138464 | 0.5 | 0.3488275 | 0.4493419672 | 0.01572271107 | 0.02318225 |
| theta_ROI90_AECnodestrength | Blocktapping | 0.2257356 | 0.77426435 | 0.5 | 0.2915485 | 0.4849981772 | -0.01557814699 | 0.02732701 |
| theta_ROI90_AECnodestrength | Sum_PC | 0.2342577 | 0.76574235 | 0.5 | 0.3059223 | 0.4109114904 | 0.01941398389 | 0.02618663 |
| theta_ROI91_AECnodestrength | Blocktapping | 0.1884937 | 0.81150632 | 0.5 | 0.2322763 | 0.9834084754 | -0.00004507522 | 0.02261107 |
| theta_ROI91_AECnodestrength | Sum_PC | 0.2042024 | 0.79579759 | 0.5 | 0.2566009 | 0.5433160153 | 0.01330482673 | 0.02341251 |
| theta_ROI92_AECnodestrength | Blocktapping | 0.1953280 | 0.80467197 | 0.5 | 0.2427424 | 0.9397465758 | -0.00028378642 | 0.02231185 |
| theta_ROI92_AECnodestrength | Sum_PC | 0.2517989 | 0.74820109 | 0.5 | 0.3365391 | 0.3051359881 | 0.02738536958 | 0.02936498 |
| theta_ROI93_AECnodestrength | Blocktapping | 0.1968567 | 0.80314333 | 0.5 | 0.2451078 | 0.8693321728 | 0.00451108296 | 0.02354834 |
| theta_ROI93_AECnodestrength | Sum_PC | 0.2177919 | 0.78220813 | 0.5 | 0.2784321 | 0.5055719110 | 0.01559601051 | 0.02563666 |
| theta_ROI94_AECnodestrength | Blocktapping | 0.3012011 | 0.69879887 | 0.5 | 0.4310269 | 0.1704434906 | -0.04562142109 | 0.03603248 |
| theta_ROI94_AECnodestrength | Sum_PC | 0.2264143 | 0.77358567 | 0.5 | 0.2926817 | 0.4407116456 | 0.01544218347 | 0.02202656 |
| theta_ROI95_AECnodestrength | Blocktapping | 0.2921146 | 0.70788540 | 0.5 | 0.4126580 | 0.4249819519 | -0.01894666522 | 0.02688878 |
| theta_ROI95_AECnodestrength | Sum_PC | 0.4518863 | 0.54811370 | 0.5 | 0.8244390 | 0.0792642884 | 0.07254128401 | 0.04567929 |
| theta_ROI96_AECnodestrength | Blocktapping | 0.4349376 | 0.56506237 | 0.5 | 0.7697161 | 0.1104341839 | -0.06738307794 | 0.04536998 |
| theta_ROI96_AECnodestrength | Sum_PC | 0.3803772 | 0.61962284 | 0.5 | 0.6138850 | 0.1824257721 | 0.04024078236 | 0.03402110 |
| theta_ROI97_AECnodestrength | Blocktapping | 0.2790106 | 0.72098935 | 0.5 | 0.3869830 | 0.2644224299 | -0.03073328901 | 0.03137108 |
| theta_ROI97_AECnodestrength | Sum_PC | 0.2897355 | 0.71026455 | 0.5 | 0.4079261 | 0.2291236673 | 0.03208636490 | 0.03022591 |
| theta_ROI98_AECnodestrength | Blocktapping | 0.2295959 | 0.77040410 | 0.5 | 0.2980201 | 0.3912399779 | -0.02144889850 | 0.02680498 |
| theta_ROI98_AECnodestrength | Sum_PC | 0.2189777 | 0.78102231 | 0.5 | 0.2803732 | 0.4491809739 | 0.01578102460 | 0.02355585 |
| theta_ROI99_AECnodestrength | Blocktapping | 0.2501753 | 0.74982472 | 0.5 | 0.3336450 | 0.3689806976 | -0.02322905038 | 0.03027570 |
| theta_ROI99_AECnodestrength | Sum_PC | 0.2291588 | 0.77084123 | 0.5 | 0.2972840 | 0.4916524788 | 0.01274461776 | 0.02363837 |
| ThetaPow1 | Blocktapping | 0.2242579 | 0.77574209 | 0.5 | 0.2890882 | 0.3947007729 | 0.02252256881 | 0.02759054 |
| ThetaPow1 | Sum_PC | 0.1870776 | 0.81292243 | 0.5 | 0.2301297 | 0.9653179919 | -0.00111922092 | 0.02035991 |
| ThetaPow10 | Blocktapping | 0.1853950 | 0.81460498 | 0.5 | 0.2275889 | 0.6373845827 | 0.00983452708 | 0.02360738 |
| ThetaPow10 | Sum_PC | 0.1746829 | 0.82531710 | 0.5 | 0.2116555 | 0.9829024714 | 0.00069988121 | 0.01928938 |

| DV | Predictor | Sum_PostModProb_Incl | Sum_PostModProb_Excl | Sum_PriorModProb_Incl | BF_Incl | Frequentist_p_value | postMean | postStd |
|---|---|---|---|---|---|---|---|---|
| ThetaPow100 | Blocktapping | 0.2028497 | 0.79715027 | 0.5 | 0.2544686 | 0.5591123342 | 0.01376128452 | 0.02580896 |
| ThetaPow100 | Sum_PC | 0.1961496 | 0.80385041 | 0.5 | 0.2440126 | 0.6652885729 | -0.00808954764 | 0.02172297 |
| ThetaPow11 | Blocktapping | 0.1976057 | 0.80239428 | 0.5 | 0.2462701 | 0.5671248554 | 0.01362814985 | 0.02357297 |
| ThetaPow11 | Sum_PC | 0.1807315 | 0.81926852 | 0.5 | 0.2206010 | 0.8552757274 | 0.00341532853 | 0.02057913 |
| ThetaPow12 | Blocktapping | 0.1929195 | 0.80708050 | 0.5 | 0.2390338 | 0.5676750869 | 0.01208117477 | 0.02398629 |
| ThetaPow12 | Sum_PC | 0.1790989 | 0.82090111 | 0.5 | 0.2181735 | 0.8745122215 | -0.00294600051 | 0.01969322 |
| ThetaPow13 | Blocktapping | 0.2051087 | 0.79489128 | 0.5 | 0.2580337 | 0.5079417740 | 0.01657801593 | 0.02582754 |
| ThetaPow13 | Sum_PC | 0.1905734 | 0.80942658 | 0.5 | 0.2354425 | 0.7241112253 | -0.00629693648 | 0.02049005 |
| ThetaPow14 | Blocktapping | 0.2026699 | 0.79733008 | 0.5 | 0.2541857 | 0.6197695159 | 0.01312892929 | 0.02592373 |
| ThetaPow14 | Sum_PC | 0.1934387 | 0.80656135 | 0.5 | 0.2398313 | 0.6948610016 | 0.00763119605 | 0.02188196 |
| ThetaPow15 | Blocktapping | 0.1928657 | 0.80713432 | 0.5 | 0.2389512 | 0.8170986336 | 0.00492621576 | 0.02339432 |
| ThetaPow15 | Sum_PC | 0.2144229 | 0.78557711 | 0.5 | 0.2729495 | 0.4942929939 | 0.01478317782 | 0.02350256 |
| ThetaPow16 | Blocktapping | 0.2295479 | 0.77045214 | 0.5 | 0.2979391 | 0.3701242659 | 0.02390079463 | 0.02834464 |
| ThetaPow16 | Sum_PC | 0.1909945 | 0.80900551 | 0.5 | 0.2360855 | 0.8884899152 | -0.00226235546 | 0.02017191 |
| ThetaPow17 | Blocktapping | 0.1838255 | 0.81617448 | 0.5 | 0.2252282 | 0.7754601562 | 0.00612708684 | 0.02311110 |
| ThetaPow17 | Sum_PC | 0.1826900 | 0.81730998 | 0.5 | 0.2235260 | 0.8309044533 | -0.00458079785 | 0.02103001 |
| ThetaPow18 | Blocktapping | 0.2582462 | 0.74175385 | 0.5 | 0.3481561 | 0.2952010055 | 0.03006026164 | 0.03169852 |
| ThetaPow18 | Sum_PC | 0.2020247 | 0.79797526 | 0.5 | 0.2531717 | 0.8771718008 | -0.00419902788 | 0.02086705 |
| ThetaPow19 | Blocktapping | 0.1890963 | 0.81090367 | 0.5 | 0.2331921 | 0.6403599177 | 0.00985490653 | 0.02286394 |
| ThetaPow19 | Sum_PC | 0.1833824 | 0.81661759 | 0.5 | 0.2245634 | 0.7987209651 | 0.00539415129 | 0.02074479 |
| ThetaPow2 | Blocktapping | 0.2401716 | 0.75982839 | 0.5 | 0.3160867 | 0.3256485464 | 0.02737636420 | 0.02995123 |
| ThetaPow2 | Sum_PC | 0.1894464 | 0.81055360 | 0.5 | 0.2337247 | 0.9654762099 | 0.00112087115 | 0.01975502 |
| ThetaPow20 | Blocktapping | 0.2106067 | 0.78939329 | 0.5 | 0.2667957 | 0.5167188553 | 0.01479156405 | 0.02696223 |
| ThetaPow20 | Sum_PC | 0.1936609 | 0.80633912 | 0.5 | 0.2401730 | 0.8872571342 | -0.00189534555 | 0.02088357 |
| ThetaPow21 | Blocktapping | 0.2104573 | 0.78954273 | 0.5 | 0.2665559 | 0.5234883906 | 0.01495387116 | 0.02710985 |
| ThetaPow21 | Sum_PC | 0.1935960 | 0.80640399 | 0.5 | 0.2400732 | 0.8077055022 | 0.00492695360 | 0.02134652 |
| ThetaPow22 | Blocktapping | 0.2005828 | 0.79941722 | 0.5 | 0.2509113 | 0.5361679888 | 0.01312457849 | 0.02528847 |
| ThetaPow22 | Sum_PC | 0.1810970 | 0.81890297 | 0.5 | 0.2211459 | 0.9259331581 | 0.00200318827 | 0.02005384 |
| ThetaPow23 | Blocktapping | 0.2032440 | 0.79675604 | 0.5 | 0.2550893 | 0.5861419160 | 0.01347703785 | 0.02492322 |
| ThetaPow23 | Sum_PC | 0.2007923 | 0.79920769 | 0.5 | 0.2512392 | 0.6458339059 | 0.01052294391 | 0.02223245 |
| ThetaPow24 | Blocktapping | 0.1847342 | 0.81526581 | 0.5 | 0.2265938 | 0.7558461834 | 0.00468596252 | 0.02358158 |
| ThetaPow24 | Sum_PC | 0.1823042 | 0.81769584 | 0.5 | 0.2229486 | 0.8377213716 | -0.00404709202 | 0.02052450 |
| ThetaPow25 | Blocktapping | 0.2457462 | 0.75425383 | 0.5 | 0.3258136 | 0.4512392731 | 0.02091084044 | 0.03054776 |
| ThetaPow25 | Sum_PC | 0.1743705 | 0.82562951 | 0.5 | 0.2111970 | 0.9936609271 | 0.00011528929 | 0.01837254 |
| ThetaPow26 | Blocktapping | 0.1895069 | 0.81049310 | 0.5 | 0.2338168 | 0.6255415309 | 0.01065834677 | 0.02286886 |
| ThetaPow26 | Sum_PC | 0.1814040 | 0.81859605 | 0.5 | 0.2216038 | 0.9197491520 | -0.00258924029 | 0.02032347 |
| ThetaPow27 | Blocktapping | 0.1800988 | 0.81990116 | 0.5 | 0.2196592 | 0.8813908755 | -0.00438225728 | 0.02307191 |
| ThetaPow27 | Sum_PC | 0.1796636 | 0.82033639 | 0.5 | 0.2190121 | 0.9131709523 | -0.00259220742 | 0.02033961 |
| ThetaPow28 | Blocktapping | 0.1871456 | 0.81285435 | 0.5 | 0.2302327 | 0.8895890289 | 0.00304149851 | 0.02359667 |
| ThetaPow28 | Sum_PC | 0.1913900 | 0.80861000 | 0.5 | 0.2366901 | 0.7990967470 | 0.00599019100 | 0.02120741 |
| ThetaPow29 | Blocktapping | 0.2018811 | 0.79811886 | 0.5 | 0.2529462 | 0.5622932587 | 0.01324913567 | 0.02562831 |

| DV | Predictor | Sum_PostModProb_Incl | Sum_PostModProb_Excl | Sum_PriorModProb_Incl | BF_Incl | Frequentist_p_value | postMean | postStd |
|---|---|---|---|---|---|---|---|---|
| ThetaPow29 | Sum_PC | 0.1869455 | 0.81305450 | 0.5 | 0.2299299 | 0.9287024187 | 0.00201285269 | 0.02078193 |
| ThetaPow3 | Blocktapping | 0.2276818 | 0.77231818 | 0.5 | 0.2948031 | 0.3677316972 | 0.02463003334 | 0.02789643 |
| ThetaPow3 | Sum_PC | 0.1894286 | 0.81057135 | 0.5 | 0.2336977 | 0.9695817182 | -0.00051053302 | 0.01984544 |
| ThetaPow30 | Blocktapping | 0.2097090 | 0.79029098 | 0.5 | 0.2653567 | 0.5447205048 | 0.01329415366 | 0.02524758 |
| ThetaPow30 | Sum_PC | 0.1958035 | 0.80419652 | 0.5 | 0.2434772 | 0.7299182153 | 0.00713177807 | 0.02125979 |
| ThetaPow31 | Blocktapping | 0.1805813 | 0.81941866 | 0.5 | 0.2203774 | 0.9726167135 | 0.00006174023 | 0.02228162 |
| ThetaPow31 | Sum_PC | 0.1845839 | 0.81541607 | 0.5 | 0.2263678 | 0.7493108700 | -0.00649620041 | 0.02112106 |
| ThetaPow32 | Blocktapping | 0.2099766 | 0.79002342 | 0.5 | 0.2657853 | 0.4237568913 | 0.01960331887 | 0.02628291 |
| ThetaPow32 | Sum_PC | 0.1856559 | 0.81434409 | 0.5 | 0.2279821 | 0.8417600360 | -0.00369066163 | 0.02020159 |
| ThetaPow33 | Blocktapping | 0.2097445 | 0.79025554 | 0.5 | 0.2654135 | 0.4435705570 | 0.01879985546 | 0.02609609 |
| ThetaPow33 | Sum_PC | 0.1851291 | 0.81487085 | 0.5 | 0.2271883 | 0.8464664221 | -0.00153285820 | 0.01965434 |
| ThetaPow34 | Blocktapping | 0.2312333 | 0.76876666 | 0.5 | 0.3007848 | 0.4144422419 | 0.02264561698 | 0.02830827 |
| ThetaPow34 | Sum_PC | 0.2137249 | 0.78627505 | 0.5 | 0.2718196 | 0.7684016278 | 0.00748318406 | 0.02291171 |
| ThetaPow35 | Blocktapping | 0.1749836 | 0.82501645 | 0.5 | 0.2120970 | 0.9475666345 | -0.00138404266 | 0.02154983 |
| ThetaPow35 | Sum_PC | 0.1815152 | 0.81848476 | 0.5 | 0.2217699 | 0.6905275773 | 0.00741533537 | 0.02085224 |
| ThetaPow36 | Blocktapping | 0.2382464 | 0.76175359 | 0.5 | 0.3127605 | 0.3473566388 | 0.02566457794 | 0.02923231 |
| ThetaPow36 | Sum_PC | 0.1951710 | 0.80482898 | 0.5 | 0.2425000 | 0.8681992279 | -0.00242679522 | 0.02032086 |
| ThetaPow37 | Blocktapping | 0.1992753 | 0.80072473 | 0.5 | 0.2488686 | 0.5989540536 | 0.01220151732 | 0.02532162 |
| ThetaPow37 | Sum_PC | 0.1868502 | 0.81314981 | 0.5 | 0.2297857 | 0.9823505498 | 0.00147306604 | 0.02082610 |
| ThetaPow38 | Blocktapping | 0.2133636 | 0.78663641 | 0.5 | 0.2712353 | 0.4359594576 | 0.01856524929 | 0.02590410 |
| ThetaPow38 | Sum_PC | 0.1884918 | 0.81150823 | 0.5 | 0.2322734 | 0.9585389070 | 0.00147681075 | 0.02007455 |
| ThetaPow39 | Blocktapping | 0.1870219 | 0.81297808 | 0.5 | 0.2300455 | 0.5977509678 | 0.01159587847 | 0.02455021 |
| ThetaPow39 | Sum_PC | 0.1764165 | 0.82358347 | 0.5 | 0.2142060 | 0.8916205882 | -0.00211029207 | 0.01979295 |
| ThetaPow4 | Blocktapping | 0.3184620 | 0.68153804 | 0.5 | 0.4672695 | 0.1787364757 | 0.04706968198 | 0.03876836 |
| ThetaPow4 | Sum_PC | 0.2153749 | 0.78462506 | 0.5 | 0.2744941 | 0.8327909155 | 0.00426772287 | 0.02055662 |
| ThetaPow40 | Blocktapping | 0.2115929 | 0.78840705 | 0.5 | 0.2683803 | 0.4594588915 | 0.01669796018 | 0.02599920 |
| ThetaPow40 | Sum_PC | 0.1925214 | 0.80747862 | 0.5 | 0.2384229 | 0.7690521166 | -0.00551875124 | 0.02153491 |
| ThetaPow41 | Blocktapping | 0.2389242 | 0.76107583 | 0.5 | 0.3139295 | 0.3761297990 | 0.02418699441 | 0.03004093 |
| ThetaPow41 | Sum_PC | 0.1982309 | 0.80176910 | 0.5 | 0.2472419 | 0.7837298977 | 0.00636189485 | 0.02073548 |
| ThetaPow42 | Blocktapping | 0.1956038 | 0.80439620 | 0.5 | 0.2431685 | 0.6531697732 | 0.00958258866 | 0.02402943 |
| ThetaPow42 | Sum_PC | 0.1882985 | 0.81170153 | 0.5 | 0.2319799 | 0.7615872670 | -0.00574189305 | 0.01993061 |
| ThetaPow43 | Blocktapping | 0.1839747 | 0.81602535 | 0.5 | 0.2254521 | 0.6958885181 | 0.00839160053 | 0.02256397 |
| ThetaPow43 | Sum_PC | 0.1800985 | 0.81990152 | 0.5 | 0.2196587 | 0.8061076945 | -0.00487685458 | 0.02038620 |
| ThetaPow44 | Blocktapping | 0.1836889 | 0.81631110 | 0.5 | 0.2250232 | 0.8572964465 | 0.00295886462 | 0.02197337 |
| ThetaPow44 | Sum_PC | 0.2108885 | 0.78911147 | 0.5 | 0.2672481 | 0.4769707836 | -0.01539869615 | 0.02407212 |
| ThetaPow45 | Blocktapping | 0.1665820 | 0.83341803 | 0.5 | 0.1998781 | 0.8813228650 | 0.00360234329 | 0.02083389 |
| ThetaPow45 | Sum_PC | 0.1653057 | 0.83469426 | 0.5 | 0.1980435 | 0.9520385551 | -0.00073027831 | 0.01878500 |
| ThetaPow46 | Blocktapping | 0.1885167 | 0.81148335 | 0.5 | 0.2323112 | 0.7525438508 | -0.00719302523 | 0.02340091 |
| ThetaPow46 | Sum_PC | 0.1906956 | 0.80930444 | 0.5 | 0.2356290 | 0.6949007178 | -0.00899085098 | 0.02130228 |
| ThetaPow47 | Blocktapping | 0.1998055 | 0.80019446 | 0.5 | 0.2496962 | 0.6116357127 | 0.01189801680 | 0.02496208 |
| ThetaPow47 | Sum_PC | 0.1895808 | 0.81041922 | 0.5 | 0.2339293 | 0.7350922749 | 0.00645393057 | 0.02084230 |

| DV | Predictor | Sum_PostModProb_Incl | Sum_PostModProb_Excl | Sum_PriorModProb_Incl | BF_Incl | Frequentist_p_value | postMean | postStd |
|---|---|---|---|---|---|---|---|---|
| ThetaPow48 | Blocktapping | 0.1848965 | 0.81510351 | 0.5 | 0.2268380 | 0.8496868871 | 0.00439163908 | 0.02344725 |
| ThetaPow48 | Sum_PC | 0.1894879 | 0.81051209 | 0.5 | 0.2337879 | 0.7686213995 | 0.00674167826 | 0.02148347 |
| ThetaPow49 | Blocktapping | 0.2251711 | 0.77482887 | 0.5 | 0.2906076 | 0.4002763988 | 0.02094168255 | 0.02860286 |
| ThetaPow49 | Sum_PC | 0.1857548 | 0.81424520 | 0.5 | 0.2281313 | 0.8893623947 | -0.00220060920 | 0.01980060 |
| ThetaPow5 | Blocktapping | 0.3068363 | 0.69316366 | 0.5 | 0.4426607 | 0.2053806985 | 0.04477056902 | 0.03858706 |
| ThetaPow5 | Sum_PC | 0.2106256 | 0.78937444 | 0.5 | 0.2668259 | 0.7879654945 | 0.00537768628 | 0.01977569 |
| ThetaPow50 | Blocktapping | 0.2247292 | 0.77527079 | 0.5 | 0.2898719 | 0.4366884341 | 0.01981589525 | 0.02801408 |
| ThetaPow50 | Sum_PC | 0.2003230 | 0.79967702 | 0.5 | 0.2505049 | 0.7644382471 | -0.00527521701 | 0.02180431 |
| ThetaPow51 | Blocktapping | 0.1736878 | 0.82631223 | 0.5 | 0.2101963 | 0.9816669648 | 0.00086544408 | 0.02228046 |
| ThetaPow51 | Sum_PC | 0.1744145 | 0.82558547 | 0.5 | 0.2112616 | 0.8466705434 | -0.00345004729 | 0.01961322 |
| ThetaPow52 | Blocktapping | 0.1836540 | 0.81634604 | 0.5 | 0.2249707 | 0.6672162952 | 0.00997857505 | 0.02363586 |
| ThetaPow52 | Sum_PC | 0.1771801 | 0.82281994 | 0.5 | 0.2153327 | 0.9987728580 | 0.00003896918 | 0.01972106 |
| ThetaPow53 | Blocktapping | 0.1806879 | 0.81931209 | 0.5 | 0.2205361 | 0.9002645758 | -0.00172951299 | 0.02247625 |
| ThetaPow53 | Sum_PC | 0.1821458 | 0.81785419 | 0.5 | 0.2227118 | 0.6472936898 | 0.00965015678 | 0.02116914 |
| ThetaPow54 | Blocktapping | 0.2419954 | 0.75800460 | 0.5 | 0.3192532 | 0.3412199637 | 0.02622607601 | 0.02882253 |
| ThetaPow54 | Sum_PC | 0.2006116 | 0.79938844 | 0.5 | 0.2509563 | 0.7591925257 | 0.00657060663 | 0.02048331 |
| ThetaPow55 | Blocktapping | 0.2189452 | 0.78105479 | 0.5 | 0.2803199 | 0.4035845462 | 0.01989939798 | 0.02725327 |
| ThetaPow55 | Sum_PC | 0.1847304 | 0.81526965 | 0.5 | 0.2265880 | 0.9926176000 | 0.00007764019 | 0.01937502 |
| ThetaPow56 | Blocktapping | 0.1860789 | 0.81392109 | 0.5 | 0.2286203 | 0.6060906263 | 0.01147174288 | 0.02344925 |
| ThetaPow56 | Sum_PC | 0.1767850 | 0.82321496 | 0.5 | 0.2147495 | 0.7952786945 | -0.00436825455 | 0.01978897 |
| ThetaPow57 | Blocktapping | 0.1904275 | 0.80957248 | 0.5 | 0.2352198 | 0.5534433870 | 0.01257235523 | 0.02426108 |
| ThetaPow57 | Sum_PC | 0.1732494 | 0.82675062 | 0.5 | 0.2095546 | 0.9578210604 | -0.00088745867 | 0.01837993 |
| ThetaPow58 | Blocktapping | 0.2664770 | 0.73352301 | 0.5 | 0.3632837 | 0.2597559297 | 0.03299112579 | 0.03238661 |
| ThetaPow58 | Sum_PC | 0.1990084 | 0.80099161 | 0.5 | 0.2484525 | 0.9415922129 | 0.00215317467 | 0.02010910 |
| ThetaPow59 | Blocktapping | 0.1724268 | 0.82757322 | 0.5 | 0.2083523 | 0.8739714438 | -0.00363415462 | 0.02143825 |
| ThetaPow59 | Sum_PC | 0.1785852 | 0.82141476 | 0.5 | 0.2174118 | 0.6822744879 | -0.00817358542 | 0.02045233 |
| ThetaPow6 | Blocktapping | 0.2361223 | 0.76387770 | 0.5 | 0.3091101 | 0.3685744213 | 0.02356546009 | 0.02962244 |
| ThetaPow6 | Sum_PC | 0.1883531 | 0.81164688 | 0.5 | 0.2320629 | 0.9681233723 | 0.00159619542 | 0.02001650 |
| ThetaPow60 | Blocktapping | 0.1755205 | 0.82447946 | 0.5 | 0.2128865 | 0.9363395290 | -0.00151323982 | 0.02218325 |
| ThetaPow60 | Sum_PC | 0.1787998 | 0.82120023 | 0.5 | 0.2177298 | 0.7250228360 | -0.00695611023 | 0.02032091 |
| ThetaPow61 | Blocktapping | 0.1789150 | 0.82108498 | 0.5 | 0.2179007 | 0.8776639604 | -0.00423928930 | 0.02226552 |
| ThetaPow61 | Sum_PC | 0.1923203 | 0.80767971 | 0.5 | 0.2381146 | 0.6073516722 | -0.01076557874 | 0.02147631 |
| ThetaPow62 | Blocktapping | 0.1810240 | 0.81897602 | 0.5 | 0.2210370 | 0.9209985618 | 0.00088754033 | 0.02187034 |
| ThetaPow62 | Sum_PC | 0.1861693 | 0.81383074 | 0.5 | 0.2287567 | 0.6970261789 | -0.00768696469 | 0.02187426 |
| ThetaPow63 | Blocktapping | 0.1724161 | 0.82758393 | 0.5 | 0.2083367 | 0.9285565744 | 0.00076471578 | 0.02121702 |
| ThetaPow63 | Sum_PC | 0.1697598 | 0.83024020 | 0.5 | 0.2044707 | 0.9226535619 | -0.00251174271 | 0.01901425 |
| ThetaPow64 | Blocktapping | 0.1707307 | 0.82926934 | 0.5 | 0.2058808 | 0.8742103501 | 0.00191846647 | 0.02127733 |
| ThetaPow64 | Sum_PC | 0.1718862 | 0.82811384 | 0.5 | 0.2075634 | 0.9178095584 | -0.00140971616 | 0.01998400 |
| ThetaPow65 | Blocktapping | 0.2168832 | 0.78311678 | 0.5 | 0.2769488 | 0.8531243246 | 0.00420399572 | 0.02752918 |
| ThetaPow65 | Sum_PC | 0.1609155 | 0.83908453 | 0.5 | 0.1917750 | 0.8326543783 | -0.00314631635 | 0.01735962 |
| ThetaPow66 | Blocktapping | 0.1876962 | 0.81230382 | 0.5 | 0.2310665 | 0.8643389704 | 0.00434487657 | 0.02285572 |

| DV | Predictor | Sum_PostModProb_Incl | Sum_PostModProb_Excl | Sum_PriorModProb_Incl | BF_Incl | Frequentist_p_value | postMean | postStd |
|---|---|---|---|---|---|---|---|---|
| ThetaPow66 | Sum_PC | 0.2016608 | 0.79833924 | 0.5 | 0.2526003 | 0.5573976719 | 0.01273276951 | 0.02299086 |
| ThetaPow67 | Blocktapping | 0.1723311 | 0.82766885 | 0.5 | 0.2082127 | 0.9580647559 | 0.00017473350 | 0.02119773 |
| ThetaPow67 | Sum_PC | 0.1783113 | 0.82168871 | 0.5 | 0.2170059 | 0.7453431251 | 0.00670619538 | 0.02072677 |
| ThetaPow68 | Blocktapping | 0.1651784 | 0.83482165 | 0.5 | 0.1978607 | 0.9969081696 | 0.00026332768 | 0.02110518 |
| ThetaPow68 | Sum_PC | 0.1690425 | 0.83095746 | 0.5 | 0.2034310 | 0.7831564726 | -0.00494767989 | 0.01995977 |
| ThetaPow69 | Blocktapping | 0.1812324 | 0.81876760 | 0.5 | 0.2213478 | 0.9076123277 | -0.00363771240 | 0.02114395 |
| ThetaPow69 | Sum_PC | 0.1877583 | 0.81224171 | 0.5 | 0.2311606 | 0.6807306929 | -0.00761609043 | 0.02152468 |
| ThetaPow7 | Blocktapping | 0.2369834 | 0.76301665 | 0.5 | 0.3105874 | 0.3415193838 | 0.02507691552 | 0.03049608 |
| ThetaPow7 | Sum_PC | 0.1897357 | 0.81026426 | 0.5 | 0.2341653 | 0.9665872215 | 0.00129608594 | 0.02030852 |
| ThetaPow70 | Blocktapping | 0.1877120 | 0.81228802 | 0.5 | 0.2310904 | 0.8451378186 | 0.00313713203 | 0.02328406 |
| ThetaPow70 | Sum_PC | 0.1881136 | 0.81188645 | 0.5 | 0.2316993 | 0.7971159652 | -0.00552153171 | 0.01997043 |
| ThetaPow71 | Blocktapping | 0.1860846 | 0.81391542 | 0.5 | 0.2286289 | 0.8401434555 | 0.00420047486 | 0.02350317 |
| ThetaPow71 | Sum_PC | 0.1818190 | 0.81818105 | 0.5 | 0.2222234 | 0.9841684393 | 0.00048938260 | 0.02021842 |
| ThetaPow72 | Blocktapping | 0.1758652 | 0.82413475 | 0.5 | 0.2133938 | 0.7905224601 | 0.00569971232 | 0.02206715 |
| ThetaPow72 | Sum_PC | 0.1707824 | 0.82921760 | 0.5 | 0.2059561 | 0.9493030595 | -0.00071159379 | 0.01957762 |
| ThetaPow73 | Blocktapping | 0.1784399 | 0.82156010 | 0.5 | 0.2171964 | 0.9190257903 | 0.00256401184 | 0.02182298 |
| ThetaPow73 | Sum_PC | 0.1820641 | 0.81793587 | 0.5 | 0.2225897 | 0.7789658366 | 0.00446928016 | 0.02086436 |
| ThetaPow74 | Blocktapping | 0.1640171 | 0.83598287 | 0.5 | 0.1961968 | 0.9298343869 | -0.00257280527 | 0.02140025 |
| ThetaPow74 | Sum_PC | 0.1633022 | 0.83669781 | 0.5 | 0.1951746 | 0.8211930593 | 0.00407612543 | 0.01857870 |
| ThetaPow75 | Blocktapping | 0.1720040 | 0.82799598 | 0.5 | 0.2077353 | 0.8623585430 | -0.00377714106 | 0.02113385 |
| ThetaPow75 | Sum_PC | 0.1715843 | 0.82841568 | 0.5 | 0.2071235 | 0.7766155588 | -0.00563979519 | 0.01933858 |
| ThetaPow76 | Blocktapping | 0.1741576 | 0.82584242 | 0.5 | 0.2108848 | 0.9372593869 | 0.00177702970 | 0.02188293 |
| ThetaPow76 | Sum_PC | 0.1814429 | 0.81855708 | 0.5 | 0.2216619 | 0.7323360704 | -0.00653758948 | 0.02009109 |
| ThetaPow77 | Blocktapping | 0.1869430 | 0.81305703 | 0.5 | 0.2299260 | 0.7311670854 | 0.00685007816 | 0.02352935 |
| ThetaPow77 | Sum_PC | 0.1801027 | 0.81989734 | 0.5 | 0.2196649 | 0.8915062557 | -0.00226032002 | 0.01995222 |
| ThetaPow78 | Blocktapping | 0.1966400 | 0.80335999 | 0.5 | 0.2447720 | 0.6534042982 | 0.01029438072 | 0.02530951 |
| ThetaPow78 | Sum_PC | 0.1898603 | 0.81013967 | 0.5 | 0.2343551 | 0.7921041400 | 0.00497327650 | 0.02115253 |
| ThetaPow79 | Blocktapping | 0.1865583 | 0.81344169 | 0.5 | 0.2293444 | 0.8838122897 | 0.00239565127 | 0.02284274 |
| ThetaPow79 | Sum_PC | 0.1914046 | 0.80859542 | 0.5 | 0.2367124 | 0.7189802975 | -0.00730159008 | 0.02180235 |
| ThetaPow8 | Blocktapping | 0.2725983 | 0.72740170 | 0.5 | 0.3747562 | 0.2521757947 | 0.03503714573 | 0.03383367 |
| ThetaPow8 | Sum_PC | 0.2024017 | 0.79759831 | 0.5 | 0.2537639 | 0.8397121550 | 0.00395773876 | 0.02087258 |
| ThetaPow80 | Blocktapping | 0.1798431 | 0.82015692 | 0.5 | 0.2192789 | 0.9781332933 | 0.00015128976 | 0.02297814 |
| ThetaPow80 | Sum_PC | 0.1932124 | 0.80678761 | 0.5 | 0.2394836 | 0.6134368704 | -0.01114277934 | 0.02217441 |
| ThetaPow81 | Blocktapping | 0.1660172 | 0.83398276 | 0.5 | 0.1990656 | 0.7526212812 | -0.00742621565 | 0.02127881 |
| ThetaPow81 | Sum_PC | 0.1645658 | 0.83543420 | 0.5 | 0.1969824 | 0.7980937088 | -0.00499119774 | 0.01868904 |
| ThetaPow82 | Blocktapping | 0.1797182 | 0.82028177 | 0.5 | 0.2190933 | 0.9498107782 | -0.00066220221 | 0.02260920 |
| ThetaPow82 | Sum_PC | 0.1819078 | 0.81809221 | 0.5 | 0.2223561 | 0.9302678417 | -0.00182874015 | 0.02038214 |
| ThetaPow83 | Blocktapping | 0.1780760 | 0.82192400 | 0.5 | 0.2166575 | 0.9819052085 | 0.00111154854 | 0.02170766 |
| ThetaPow83 | Sum_PC | 0.1802779 | 0.81972215 | 0.5 | 0.2199256 | 0.8783930269 | 0.00192378047 | 0.01980195 |
| ThetaPow84 | Blocktapping | 0.1810549 | 0.81894509 | 0.5 | 0.2210831 | 0.9220961117 | -0.00218581220 | 0.02200819 |
| ThetaPow84 | Sum_PC | 0.2044204 | 0.79557962 | 0.5 | 0.2569452 | 0.5506679736 | -0.01391733728 | 0.02302057 |

| DV | Predictor | Sum_PostModProb_Incl | Sum_PostModProb_Excl | Sum_PriorModProb_Incl | BF_Incl | Frequentist_p_value | postMean | postStd |
|---|---|---|---|---|---|---|---|---|
| ThetaPow85 | Blocktapping | 0.1759064 | 0.82409365 | 0.5 | 0.2134543 | 0.9062761101 | -0.00274494991 | 0.02157269 |
| ThetaPow85 | Sum_PC | 0.1782667 | 0.82173331 | 0.5 | 0.2169398 | 0.7170753072 | -0.00703768001 | 0.01959794 |
| ThetaPow86 | Blocktapping | 0.1972915 | 0.80270854 | 0.5 | 0.2457822 | 0.6388787461 | 0.01012131402 | 0.02421956 |
| ThetaPow86 | Sum_PC | 0.1835131 | 0.81648686 | 0.5 | 0.2247594 | 0.8374003581 | 0.00328833171 | 0.02087797 |
| ThetaPow87 | Blocktapping | 0.1938637 | 0.80613626 | 0.5 | 0.2404851 | 0.6599134491 | 0.00893908796 | 0.02414695 |
| ThetaPow87 | Sum_PC | 0.1826817 | 0.81731827 | 0.5 | 0.2235136 | 0.9094015075 | -0.00086310509 | 0.02084707 |
| ThetaPow88 | Blocktapping | 0.1801764 | 0.81982364 | 0.5 | 0.2197745 | 0.9446834990 | -0.00123693965 | 0.02326519 |
| ThetaPow88 | Sum_PC | 0.1797232 | 0.82027675 | 0.5 | 0.2191007 | 0.8751302803 | 0.00231299152 | 0.02071481 |
| ThetaPow89 | Blocktapping | 0.2041974 | 0.79580256 | 0.5 | 0.2565931 | 0.5632463104 | 0.01268519464 | 0.02574749 |
| ThetaPow89 | Sum_PC | 0.1958788 | 0.80412120 | 0.5 | 0.2435936 | 0.7213055906 | -0.00670773771 | 0.02170691 |
| ThetaPow9 | Blocktapping | 0.2758088 | 0.72419116 | 0.5 | 0.3808509 | 0.2482544382 | 0.03849118311 | 0.03375157 |
| ThetaPow9 | Sum_PC | 0.2007850 | 0.79921499 | 0.5 | 0.2512278 | 0.8348144738 | 0.00431808399 | 0.01985809 |
| ThetaPow90 | Blocktapping | 0.1705198 | 0.82948023 | 0.5 | 0.2055742 | 0.8921752230 | -0.00333150124 | 0.02121869 |
| ThetaPow90 | Sum_PC | 0.1696550 | 0.83034502 | 0.5 | 0.2043187 | 0.9695486523 | -0.00086866245 | 0.01940593 |
| ThetaPow91 | Blocktapping | 0.1689176 | 0.83108242 | 0.5 | 0.2032501 | 0.8864417904 | -0.00345987711 | 0.02113495 |
| ThetaPow91 | Sum_PC | 0.1686236 | 0.83137639 | 0.5 | 0.2028246 | 0.9597687244 | 0.00042018012 | 0.01988066 |
| ThetaPow92 | Blocktapping | 0.1789324 | 0.82106761 | 0.5 | 0.2179265 | 0.9016769253 | -0.00301534139 | 0.02155761 |
| ThetaPow92 | Sum_PC | 0.1910679 | 0.80893212 | 0.5 | 0.2361977 | 0.5957991500 | -0.01069003047 | 0.02134016 |
| ThetaPow93 | Blocktapping | 0.1665565 | 0.83344352 | 0.5 | 0.1998414 | 0.9147167726 | -0.00220426179 | 0.02174785 |
| ThetaPow93 | Sum_PC | 0.1658714 | 0.83412864 | 0.5 | 0.1988559 | 0.9571564655 | -0.00168767929 | 0.01956661 |
| ThetaPow94 | Blocktapping | 0.1760076 | 0.82399238 | 0.5 | 0.2136035 | 0.8735243579 | 0.00249392086 | 0.02215021 |
| ThetaPow94 | Sum_PC | 0.1848226 | 0.81517736 | 0.5 | 0.2267269 | 0.7084044802 | -0.00824526654 | 0.02097550 |
| ThetaPow95 | Blocktapping | 0.1811641 | 0.81883594 | 0.5 | 0.2212459 | 0.9609241160 | -0.00029326302 | 0.02214122 |
| ThetaPow95 | Sum_PC | 0.1893769 | 0.81062307 | 0.5 | 0.2336190 | 0.6161636617 | -0.00989143141 | 0.02132705 |
| ThetaPow96 | Blocktapping | 0.1803082 | 0.81969179 | 0.5 | 0.2199707 | 0.8549038121 | 0.00202885758 | 0.02195930 |
| ThetaPow96 | Sum_PC | 0.1988595 | 0.80114050 | 0.5 | 0.2482205 | 0.5288864100 | -0.01336349501 | 0.02274317 |
| ThetaPow97 | Blocktapping | 0.1871587 | 0.81284129 | 0.5 | 0.2302525 | 0.7802712247 | -0.00747516861 | 0.02309566 |
| ThetaPow97 | Sum_PC | 0.1851491 | 0.81485091 | 0.5 | 0.2272184 | 0.7479901622 | -0.00660255299 | 0.02154603 |
| ThetaPow98 | Blocktapping | 0.1818548 | 0.81814524 | 0.5 | 0.2222769 | 0.9311495822 | 0.00241446884 | 0.02310282 |
| ThetaPow98 | Sum_PC | 0.1855549 | 0.81444510 | 0.5 | 0.2278298 | 0.8328522469 | 0.00383147037 | 0.02161435 |
| ThetaPow99 | Blocktapping | 0.2033661 | 0.79663392 | 0.5 | 0.2552817 | 0.5450348880 | 0.01427903234 | 0.02537654 |
| ThetaPow99 | Sum_PC | 0.1917451 | 0.80825486 | 0.5 | 0.2372335 | 0.7026884617 | -0.00671336895 | 0.02030471 |

**Table S5** Full Summary of Regression Models of EEG features (DV) and Subjective/Cognitive Functioning (Predictors), with Component Scores for Affective and Physical Burden added to the Null Model. ROI numbering refers to the labeling of the 100 parcel 7 network version of the Schaefer-Yeo atlases (2018)

| DV | Predictor | Sum_PostModProb_Incl | Sum_PostModProb_Excl | Sum_PriorModProb_Incl | BF_Incl | Frequentist_p_value | postMean | postStd |
|---|---|---|---|---|---|---|---|---|
| AEC_average_alpha | Blocktapping | 0.2871187 | 0.7128813 | 0.5 | 0.4027582 | 0.65926577 | 0.009620233886 | 0.02804896 |
| AEC_average_alpha | Sum_PC | 0.3717655 | 0.6282345 | 0.5 | 0.5917623 | 0.21098172 | -0.047335307671 | 0.04366738 |
| AEC_average_beta | Blocktapping | 0.2129283 | 0.7870717 | 0.5 | 0.2705322 | 0.90330802 | 0.002983548439 | 0.02389553 |
| AEC_average_beta | Sum_PC | 0.2325357 | 0.7674643 | 0.5 | 0.3029922 | 0.53808509 | 0.016431688935 | 0.02850493 |
| AEC_average_gamma | Blocktapping | 0.2722903 | 0.7277097 | 0.5 | 0.3741744 | 0.52212972 | -0.018762277123 | 0.02939515 |
| AEC_average_gamma | Sum_PC | 0.2621293 | 0.7378707 | 0.5 | 0.3552509 | 0.62786805 | -0.014556752771 | 0.02986921 |
| AEC_average_theta | Blocktapping | 0.2949349 | 0.7050651 | 0.5 | 0.4183088 | 0.39480497 | -0.024020169923 | 0.03207671 |
| AEC_average_theta | Sum_PC | 0.2726687 | 0.7273313 | 0.5 | 0.3748892 | 0.56603681 | 0.014799398570 | 0.02931493 |
| alpha_ROI1_AECnodestrength | Blocktapping | 0.3286323 | 0.6713677 | 0.5 | 0.4894968 | 0.51939751 | 0.015808469892 | 0.03007407 |
| alpha_ROI1_AECnodestrength | Sum_PC | 0.4545875 | 0.5454125 | 0.5 | 0.8334746 | 0.12700622 | -0.066802010667 | 0.04976967 |
| alpha_ROI10_AECnodestrength | Blocktapping | 0.3092638 | 0.6907362 | 0.5 | 0.4477307 | 0.34513510 | 0.028260157253 | 0.03424712 |
| alpha_ROI10_AECnodestrength | Sum_PC | 0.2756161 | 0.7243839 | 0.5 | 0.3804835 | 0.56174919 | -0.016037115026 | 0.02968193 |
| alpha_ROI100_AECnodestrength | Blocktapping | 0.2956390 | 0.7043610 | 0.5 | 0.4197265 | 0.61251603 | 0.013287796046 | 0.02984374 |
| alpha_ROI100_AECnodestrength | Sum_PC | 0.3670890 | 0.6329110 | 0.5 | 0.5800011 | 0.22756869 | -0.042911803616 | 0.04089115 |
| alpha_ROI11_AECnodestrength | Blocktapping | 0.2595848 | 0.7404152 | 0.5 | 0.3505936 | 0.67485121 | 0.009945116492 | 0.02868789 |
| alpha_ROI11_AECnodestrength | Sum_PC | 0.2630675 | 0.7369325 | 0.5 | 0.3569764 | 0.61136997 | -0.013789313558 | 0.03011120 |
| alpha_ROI12_AECnodestrength | Blocktapping | 0.3662095 | 0.6337905 | 0.5 | 0.5778084 | 0.21210320 | 0.045210054554 | 0.03962462 |
| alpha_ROI12_AECnodestrength | Sum_PC | 0.2872249 | 0.7127751 | 0.5 | 0.4029671 | 0.59604690 | -0.013124851074 | 0.02957762 |
| alpha_ROI13_AECnodestrength | Blocktapping | 0.3671636 | 0.6328364 | 0.5 | 0.5801872 | 0.24147415 | 0.038427888619 | 0.03799974 |
| alpha_ROI13_AECnodestrength | Sum_PC | 0.3385655 | 0.6614345 | 0.5 | 0.5118655 | 0.34252005 | -0.027244822489 | 0.03475750 |
| alpha_ROI14_AECnodestrength | Blocktapping | 0.2685485 | 0.7314515 | 0.5 | 0.3671446 | 0.64660085 | 0.010658844748 | 0.02927684 |
| alpha_ROI14_AECnodestrength | Sum_PC | 0.2833118 | 0.7166882 | 0.5 | 0.3953070 | 0.48095202 | -0.020648751066 | 0.03273364 |
| alpha_ROI15_AECnodestrength | Blocktapping | 0.2776911 | 0.7223089 | 0.5 | 0.3844492 | 0.80798264 | 0.004847447790 | 0.02791117 |
| alpha_ROI15_AECnodestrength | Sum_PC | 0.3728029 | 0.6271971 | 0.5 | 0.5943952 | 0.20928389 | -0.048875755745 | 0.04543611 |

| DV | Predictor | Sum_PostModProb_Incl | Sum_PostModProb_Excl | Sum_PriorModProb_Incl | BF_Incl | Frequentist_p_value | postMean | postStd |
|---|---|---|---|---|---|---|---|---|
| alpha_ROI16_AECnodestrength | Blocktapping | 0.4286608 | 0.5713392 | 0.5 | 0.7502737 | 0.39078524 | 0.023539193858 | 0.03354511 |
| alpha_ROI16_AECnodestrength | Sum_PC | 0.7037336 | 0.2962664 | 0.5 | 2.3753410 | 0.02745358 | -0.138823569158 | 0.07212257 |
| alpha_ROI17_AECnodestrength | Blocktapping | 0.3011398 | 0.6988602 | 0.5 | 0.4309014 | 0.46541523 | 0.018981384975 | 0.03079050 |
| alpha_ROI17_AECnodestrength | Sum_PC | 0.3481203 | 0.6518797 | 0.5 | 0.5340253 | 0.26030579 | -0.038831265107 | 0.03971720 |
| alpha_ROI18_AECnodestrength | Blocktapping | 0.3770231 | 0.6229769 | 0.5 | 0.6051960 | 0.22666911 | 0.040874275383 | 0.03913025 |
| alpha_ROI18_AECnodestrength | Sum_PC | 0.3458249 | 0.6541751 | 0.5 | 0.5286428 | 0.35557951 | -0.028266102084 | 0.03531993 |
| alpha_ROI19_AECnodestrength | Blocktapping | 0.2616811 | 0.7383189 | 0.5 | 0.3544282 | 0.78497988 | 0.006049947184 | 0.02802912 |
| alpha_ROI19_AECnodestrength | Sum_PC | 0.3017610 | 0.6982390 | 0.5 | 0.4321743 | 0.37605510 | -0.026919097611 | 0.03402783 |
| alpha_ROI2_AECnodestrength | Blocktapping | 0.3526683 | 0.6473317 | 0.5 | 0.5448030 | 0.77993177 | 0.005185847740 | 0.02780497 |
| alpha_ROI2_AECnodestrength | Sum_PC | 0.6422745 | 0.3577255 | 0.5 | 1.7954396 | 0.04069352 | -0.129166519578 | 0.07067289 |
| alpha_ROI20_AECnodestrength | Blocktapping | 0.3749400 | 0.6250600 | 0.5 | 0.5998465 | 0.27620231 | 0.035008815414 | 0.03566599 |
| alpha_ROI20_AECnodestrength | Sum_PC | 0.4067091 | 0.5932909 | 0.5 | 0.6855137 | 0.19209913 | -0.048168520067 | 0.04273690 |
| alpha_ROI21_AECnodestrength | Blocktapping | 0.4900068 | 0.5099932 | 0.5 | 0.9608104 | 0.12830472 | 0.060941747558 | 0.04621462 |
| alpha_ROI21_AECnodestrength | Sum_PC | 0.4799594 | 0.5200406 | 0.5 | 0.9229266 | 0.14315392 | -0.060956427243 | 0.04757685 |
| alpha_ROI22_AECnodestrength | Blocktapping | 0.3269618 | 0.6730382 | 0.5 | 0.4857998 | 0.32743576 | 0.027947546427 | 0.03422118 |
| alpha_ROI22_AECnodestrength | Sum_PC | 0.3213415 | 0.6786585 | 0.5 | 0.4734951 | 0.35970591 | -0.028139215563 | 0.03583998 |
| alpha_ROI23_AECnodestrength | Blocktapping | 0.2599627 | 0.7400373 | 0.5 | 0.3512832 | 0.85173375 | 0.001899131312 | 0.02653340 |
| alpha_ROI23_AECnodestrength | Sum_PC | 0.3151800 | 0.6848200 | 0.5 | 0.4602377 | 0.30670679 | -0.033427349235 | 0.03771804 |
| alpha_ROI24_AECnodestrength | Blocktapping | 0.3425033 | 0.6574967 | 0.5 | 0.5209201 | 0.33989727 | 0.025723326836 | 0.03408847 |
| alpha_ROI24_AECnodestrength | Sum_PC | 0.3994897 | 0.6005103 | 0.5 | 0.6652505 | 0.17925937 | -0.049718766902 | 0.04281760 |
| alpha_ROI25_AECnodestrength | Blocktapping | 0.2779733 | 0.7220267 | 0.5 | 0.3849903 | 0.47317048 | 0.018050805687 | 0.02973383 |
| alpha_ROI25_AECnodestrength | Sum_PC | 0.2702407 | 0.7297593 | 0.5 | 0.3703149 | 0.62745366 | -0.014102120921 | 0.03056514 |
| alpha_ROI26_AECnodestrength | Blocktapping | 0.2713094 | 0.7286906 | 0.5 | 0.3723246 | 0.60506852 | 0.012233919483 | 0.02921573 |
| alpha_ROI26_AECnodestrength | Sum_PC | 0.2856858 | 0.7143142 | 0.5 | 0.3999441 | 0.43607448 | -0.022536415762 | 0.03338341 |
| alpha_ROI27_AECnodestrength | Blocktapping | 0.2348820 | 0.7651180 | 0.5 | 0.3069879 | 0.78373502 | 0.006711064120 | 0.02699561 |
| alpha_ROI27_AECnodestrength | Sum_PC | 0.2371627 | 0.7628373 | 0.5 | 0.3108955 | 0.75631026 | -0.006793872465 | 0.02765438 |
| alpha_ROI28_AECnodestrength | Blocktapping | 0.2735861 | 0.7264139 | 0.5 | 0.3766256 | 0.56795353 | -0.015996603780 | 0.03043292 |
| alpha_ROI28_AECnodestrength | Sum_PC | 0.2795255 | 0.7204745 | 0.5 | 0.3879743 | 0.59109677 | -0.016198378436 | 0.03191988 |
| alpha_ROI29_AECnodestrength | Blocktapping | 0.3313295 | 0.6686705 | 0.5 | 0.4955048 | 0.47618850 | 0.018260635599 | 0.03160261 |

| DV | Predictor | Sum_PostModProb_Incl | Sum_PostModProb_Excl | Sum_PriorModProb_Incl | BF_Incl | Frequentist_p_value | postMean | postStd |
|---|---|---|---|---|---|---|---|---|
| alpha_ROI29_AECnodestrength | Sum_PC | 0.4434832 | 0.5565168 | 0.5 | 0.7968909 | 0.13942921 | -0.064758098540 | 0.04834974 |
| alpha_ROI3_AECnodestrength | Blocktapping | 0.2939613 | 0.7060387 | 0.5 | 0.4163529 | 0.83316410 | 0.003791374859 | 0.02697816 |
| alpha_ROI3_AECnodestrength | Sum_PC | 0.4236211 | 0.5763789 | 0.5 | 0.7349699 | 0.14853817 | -0.062673062158 | 0.04945795 |
| alpha_ROI30_AECnodestrength | Blocktapping | 0.4113910 | 0.5886090 | 0.5 | 0.6989208 | 0.30093246 | -0.037791474840 | 0.03658408 |
| alpha_ROI30_AECnodestrength | Sum_PC | 0.5032661 | 0.4967339 | 0.5 | 1.0131505 | 0.12741673 | -0.076452364133 | 0.05307257 |
| alpha_ROI31_AECnodestrength | Blocktapping | 0.2534933 | 0.7465067 | 0.5 | 0.3395728 | 0.64515702 | 0.010901895358 | 0.02866025 |
| alpha_ROI31_AECnodestrength | Sum_PC | 0.2520161 | 0.7479839 | 0.5 | 0.3369272 | 0.66769711 | -0.010626348397 | 0.02922579 |
| alpha_ROI32_AECnodestrength | Blocktapping | 0.2698278 | 0.7301722 | 0.5 | 0.3695400 | 0.49600517 | 0.017482790760 | 0.03030343 |
| alpha_ROI32_AECnodestrength | Sum_PC | 0.2522952 | 0.7477048 | 0.5 | 0.3374263 | 0.72494370 | -0.008780081344 | 0.02816744 |
| alpha_ROI33_AECnodestrength | Blocktapping | 0.3441521 | 0.6558479 | 0.5 | 0.5247438 | 0.42991013 | 0.018880246093 | 0.03094552 |
| alpha_ROI33_AECnodestrength | Sum_PC | 0.4598415 | 0.5401585 | 0.5 | 0.8513084 | 0.11759707 | -0.068990864974 | 0.05015061 |
| alpha_ROI34_AECnodestrength | Blocktapping | 0.3823203 | 0.6176797 | 0.5 | 0.6189621 | 0.39592215 | 0.022693106615 | 0.03229647 |
| alpha_ROI34_AECnodestrength | Sum_PC | 0.5503900 | 0.4496100 | 0.5 | 1.2241499 | 0.07276145 | -0.091909456562 | 0.05898870 |
| alpha_ROI35_AECnodestrength | Blocktapping | 0.3307369 | 0.6692631 | 0.5 | 0.4941806 | 0.35884360 | 0.023624501128 | 0.03286946 |
| alpha_ROI35_AECnodestrength | Sum_PC | 0.3740601 | 0.6259399 | 0.5 | 0.5975975 | 0.21305212 | -0.043429112756 | 0.04170580 |
| alpha_ROI36_AECnodestrength | Blocktapping | 0.2653734 | 0.7346266 | 0.5 | 0.3612357 | 0.76862426 | 0.004777135806 | 0.02616223 |
| alpha_ROI36_AECnodestrength | Sum_PC | 0.3517868 | 0.6482132 | 0.5 | 0.5427024 | 0.20852070 | -0.046806036691 | 0.04200013 |
| alpha_ROI37_AECnodestrength | Blocktapping | 0.3649121 | 0.6350879 | 0.5 | 0.5745851 | 0.32800675 | 0.027392198831 | 0.03357912 |
| alpha_ROI37_AECnodestrength | Sum_PC | 0.4600467 | 0.5399533 | 0.5 | 0.8520121 | 0.13767863 | -0.064643705124 | 0.05111265 |
| alpha_ROI38_AECnodestrength | Blocktapping | 0.2517656 | 0.7482344 | 0.5 | 0.3364795 | 0.61278616 | 0.013080314814 | 0.02853595 |
| alpha_ROI38_AECnodestrength | Sum_PC | 0.2392088 | 0.7607912 | 0.5 | 0.3144211 | 0.99935011 | 0.000353603443 | 0.02710680 |
| alpha_ROI39_AECnodestrength | Blocktapping | 0.3813501 | 0.6186499 | 0.5 | 0.6164230 | 0.25811358 | 0.036122239294 | 0.03725714 |
| alpha_ROI39_AECnodestrength | Sum_PC | 0.4158276 | 0.5841724 | 0.5 | 0.7118235 | 0.18220795 | -0.050967985417 | 0.04493524 |
| alpha_ROI4_AECnodestrength | Blocktapping | 0.2666507 | 0.7333493 | 0.5 | 0.3636066 | 0.92572602 | -0.003184125295 | 0.02719538 |
| alpha_ROI4_AECnodestrength | Sum_PC | 0.3280849 | 0.6719151 | 0.5 | 0.4882833 | 0.30822153 | -0.037440853933 | 0.03798938 |
| alpha_ROI40_AECnodestrength | Blocktapping | 0.4552023 | 0.5447977 | 0.5 | 0.8355438 | 0.40121646 | 0.023531763517 | 0.03191432 |
| alpha_ROI40_AECnodestrength | Sum_PC | 0.8238226 | 0.1761774 | 0.5 | 4.6760969 | 0.01108312 | -0.192897220133 | 0.08749801 |
| alpha_ROI41_AECnodestrength | Blocktapping | 0.3586950 | 0.6413050 | 0.5 | 0.5593205 | 0.52253369 | 0.017077623394 | 0.03003810 |
| alpha_ROI41_AECnodestrength | Sum_PC | 0.5406623 | 0.4593377 | 0.5 | 1.1770476 | 0.07652838 | -0.092037715413 | 0.05826204 |

| DV | Predictor | Sum_PostModProb_Incl | Sum_PostModProb_Excl | Sum_PriorModProb_Incl | BF_Incl | Frequentist_p_value | postMean | postStd |
|---|---|---|---|---|---|---|---|---|
| alpha_ROI42_AEC nodestrength | Blocktapping | 0.2737627 | 0.7262373 | 0.5 | 0.3769603 | 0.56979901 | 0.014319963370 | 0.02962020 |
| alpha_ROI42_AEC nodestrength | Sum_PC | 0.2948675 | 0.7051325 | 0.5 | 0.4181732 | 0.40857764 | -0.024475479285 | 0.03408696 |
| alpha_ROI43_AEC nodestrength | Blocktapping | 0.2536573 | 0.7463427 | 0.5 | 0.3398670 | 0.53983235 | 0.015848388377 | 0.02789397 |
| alpha_ROI43_AEC nodestrength | Sum_PC | 0.2639664 | 0.7360336 | 0.5 | 0.3586337 | 0.62149890 | -0.012769435557 | 0.03081192 |
| alpha_ROI44_AEC nodestrength | Blocktapping | 0.2780927 | 0.7219073 | 0.5 | 0.3852194 | 0.54086992 | -0.017883047386 | 0.03047521 |
| alpha_ROI44_AEC nodestrength | Sum_PC | 0.2683405 | 0.7316595 | 0.5 | 0.3667561 | 0.64889818 | -0.014485447396 | 0.03086552 |
| alpha_ROI45_AEC nodestrength | Blocktapping | 0.2483247 | 0.7516753 | 0.5 | 0.3303617 | 0.78809385 | -0.007519074420 | 0.02722104 |
| alpha_ROI45_AEC nodestrength | Sum_PC | 0.2635964 | 0.7364036 | 0.5 | 0.3579510 | 0.61113773 | -0.014201824242 | 0.03038208 |
| alpha_ROI46_AEC nodestrength | Blocktapping | 0.2635428 | 0.7364572 | 0.5 | 0.3578521 | 0.52747705 | -0.014718775234 | 0.02968244 |
| alpha_ROI46_AEC nodestrength | Sum_PC | 0.2560359 | 0.7439641 | 0.5 | 0.3441509 | 0.61151489 | 0.013291165970 | 0.02929676 |
| alpha_ROI47_AEC nodestrength | Blocktapping | 0.2575070 | 0.7424930 | 0.5 | 0.3468141 | 0.82548444 | 0.003887947528 | 0.02727810 |
| alpha_ROI47_AEC nodestrength | Sum_PC | 0.2935854 | 0.7064146 | 0.5 | 0.4155992 | 0.42087447 | -0.023334173435 | 0.03315044 |
| alpha_ROI48_AEC nodestrength | Blocktapping | 0.2677639 | 0.7322361 | 0.5 | 0.3656798 | 0.81367679 | 0.004556121566 | 0.02729841 |
| alpha_ROI48_AEC nodestrength | Sum_PC | 0.3433885 | 0.6566115 | 0.5 | 0.5229707 | 0.25654569 | -0.040847845526 | 0.04120193 |
| alpha_ROI49_AEC nodestrength | Blocktapping | 0.3080988 | 0.6919012 | 0.5 | 0.4452931 | 0.69510148 | 0.007841795546 | 0.02809487 |
| alpha_ROI49_AEC nodestrength | Sum_PC | 0.4405700 | 0.5594300 | 0.5 | 0.7875337 | 0.13892109 | -0.066256853403 | 0.05098182 |
| alpha_ROI5_AECn odestrength | Blocktapping | 0.2974574 | 0.7025426 | 0.5 | 0.4234013 | 0.97919395 | -0.002513436216 | 0.02811689 |
| alpha_ROI5_AECn odestrength | Sum_PC | 0.4261129 | 0.5738871 | 0.5 | 0.7425030 | 0.14892355 | -0.063660885956 | 0.04923165 |
| alpha_ROI50_AEC nodestrength | Blocktapping | 0.3460688 | 0.6539312 | 0.5 | 0.5292129 | 0.45713912 | 0.019512512842 | 0.03181034 |
| alpha_ROI50_AEC nodestrength | Sum_PC | 0.4666005 | 0.5333995 | 0.5 | 0.8747674 | 0.12223140 | -0.068207172459 | 0.05058018 |
| alpha_ROI51_AEC nodestrength | Blocktapping | 0.3337324 | 0.6662676 | 0.5 | 0.5008984 | 0.35504260 | 0.026060588002 | 0.03285586 |
| alpha_ROI51_AEC nodestrength | Sum_PC | 0.3638298 | 0.6361702 | 0.5 | 0.5719065 | 0.24975429 | -0.038404501233 | 0.04055028 |
| alpha_ROI52_AEC nodestrength | Blocktapping | 0.2993079 | 0.7006921 | 0.5 | 0.4271603 | 0.48011485 | 0.019233757451 | 0.03105903 |
| alpha_ROI52_AEC nodestrength | Sum_PC | 0.3081787 | 0.6918213 | 0.5 | 0.4454600 | 0.40088378 | -0.025164410513 | 0.03469951 |
| alpha_ROI53_AEC nodestrength | Blocktapping | 0.2703137 | 0.7296863 | 0.5 | 0.3704518 | 0.71060540 | 0.007393085934 | 0.02690090 |
| alpha_ROI53_AEC nodestrength | Sum_PC | 0.3515859 | 0.6484141 | 0.5 | 0.5422242 | 0.26039892 | -0.040035563099 | 0.04033237 |
| alpha_ROI54_AEC nodestrength | Blocktapping | 0.2640858 | 0.7359142 | 0.5 | 0.3588540 | 0.63377454 | 0.009566886403 | 0.02808602 |
| alpha_ROI54_AEC nodestrength | Sum_PC | 0.3362363 | 0.6637637 | 0.5 | 0.5065602 | 0.38043078 | -0.030447268437 | 0.03984744 |
| alpha_ROI55_AEC nodestrength | Blocktapping | 0.2696879 | 0.7303121 | 0.5 | 0.3692776 | 0.95881335 | -0.002581147025 | 0.02737000 |

| DV | Predictor | Sum_PostModProb_Incl | Sum_PostModProb_Excl | Sum_PriorModProb_Incl | BF_Incl | Frequentist_p_value | postMean | postStd |
|---|---|---|---|---|---|---|---|---|
| alpha_ROI55_AEC nodestrength | Sum_PC | 0.3484333 | 0.6515667 | 0.5 | 0.5347622 | 0.26828637 | -0.042390807068 | 0.04147517 |
| alpha_ROI56_AEC nodestrength | Blocktapping | 0.3042602 | 0.6957398 | 0.5 | 0.4373190 | 0.49931289 | 0.017879152734 | 0.03134057 |
| alpha_ROI56_AEC nodestrength | Sum_PC | 0.3064431 | 0.6935569 | 0.5 | 0.4418428 | 0.50646305 | -0.019777445668 | 0.03433468 |
| alpha_ROI57_AEC nodestrength | Blocktapping | 0.2576208 | 0.7423792 | 0.5 | 0.3470205 | 0.99177280 | 0.000085972067 | 0.02670873 |
| alpha_ROI57_AEC nodestrength | Sum_PC | 0.3044545 | 0.6955455 | 0.5 | 0.4377205 | 0.38902585 | -0.027195797696 | 0.03575602 |
| alpha_ROI58_AEC nodestrength | Blocktapping | 0.2659556 | 0.7340444 | 0.5 | 0.3623154 | 0.85308931 | -0.006474227999 | 0.02725198 |
| alpha_ROI58_AEC nodestrength | Sum_PC | 0.3357684 | 0.6642316 | 0.5 | 0.5054991 | 0.28607312 | -0.040517126034 | 0.03909890 |
| alpha_ROI59_AEC nodestrength | Blocktapping | 0.2923918 | 0.7076082 | 0.5 | 0.4132115 | 0.52021712 | 0.014981789467 | 0.02908156 |
| alpha_ROI59_AEC nodestrength | Sum_PC | 0.3519083 | 0.6480917 | 0.5 | 0.5429914 | 0.24081060 | -0.040186485251 | 0.03993308 |
| alpha_ROI6_AECn odestrength | Blocktapping | 0.3079899 | 0.6920101 | 0.5 | 0.4450656 | 0.87732728 | -0.005660688375 | 0.02736887 |
| alpha_ROI6_AECn odestrength | Sum_PC | 0.4623825 | 0.5376175 | 0.5 | 0.8600585 | 0.12480228 | -0.074065012595 | 0.05252313 |
| alpha_ROI60_AEC nodestrength | Blocktapping | 0.2898762 | 0.7101238 | 0.5 | 0.4082051 | 0.51618540 | 0.014124398712 | 0.02949204 |
| alpha_ROI60_AEC nodestrength | Sum_PC | 0.3496406 | 0.6503594 | 0.5 | 0.5376113 | 0.23967697 | -0.041342519956 | 0.03974900 |
| alpha_ROI61_AEC nodestrength | Blocktapping | 0.2536950 | 0.7463050 | 0.5 | 0.3399347 | 0.84952507 | 0.003412127085 | 0.02643219 |
| alpha_ROI61_AEC nodestrength | Sum_PC | 0.2994198 | 0.7005802 | 0.5 | 0.4273884 | 0.35747293 | -0.027772853216 | 0.03487853 |
| alpha_ROI62_AEC nodestrength | Blocktapping | 0.2587161 | 0.7412839 | 0.5 | 0.3490108 | 0.71710791 | 0.008540121060 | 0.02794026 |
| alpha_ROI62_AEC nodestrength | Sum_PC | 0.2688351 | 0.7311649 | 0.5 | 0.3676805 | 0.54886202 | -0.016064523806 | 0.03150567 |
| alpha_ROI63_AEC nodestrength | Blocktapping | 0.2693360 | 0.7306640 | 0.5 | 0.3686182 | 0.86715376 | 0.003961126198 | 0.02683589 |
| alpha_ROI63_AEC nodestrength | Sum_PC | 0.3476256 | 0.6523744 | 0.5 | 0.5328621 | 0.24502386 | -0.041724467987 | 0.03944135 |
| alpha_ROI64_AEC nodestrength | Blocktapping | 0.2993261 | 0.7006739 | 0.5 | 0.4271975 | 0.78707534 | -0.008563149625 | 0.02794996 |
| alpha_ROI64_AEC nodestrength | Sum_PC | 0.4378219 | 0.5621781 | 0.5 | 0.7787958 | 0.15179281 | -0.067734795454 | 0.05011459 |
| alpha_ROI65_AEC nodestrength | Blocktapping | 0.3068972 | 0.6931028 | 0.5 | 0.4427875 | 0.79998233 | 0.004723309442 | 0.02682710 |
| alpha_ROI65_AEC nodestrength | Sum_PC | 0.4762975 | 0.5237025 | 0.5 | 0.9094810 | 0.10556703 | -0.078523959789 | 0.05386849 |
| alpha_ROI66_AEC nodestrength | Blocktapping | 0.2655827 | 0.7344173 | 0.5 | 0.3616238 | 0.92736957 | 0.000431225621 | 0.02647350 |
| alpha_ROI66_AEC nodestrength | Sum_PC | 0.3283581 | 0.6716419 | 0.5 | 0.4888886 | 0.29329049 | -0.037214678611 | 0.03880466 |
| alpha_ROI67_AEC nodestrength | Blocktapping | 0.3649397 | 0.6350603 | 0.5 | 0.5746536 | 0.42394440 | 0.020793961321 | 0.03147906 |
| alpha_ROI67_AEC nodestrength | Sum_PC | 0.5167229 | 0.4832771 | 0.5 | 1.0692061 | 0.08785451 | -0.083342003678 | 0.05663556 |
| alpha_ROI68_AEC nodestrength | Blocktapping | 0.2975784 | 0.7024216 | 0.5 | 0.4236464 | 0.82889713 | 0.003207235714 | 0.02720877 |
| alpha_ROI68_AEC nodestrength | Sum_PC | 0.4375557 | 0.5624443 | 0.5 | 0.7779540 | 0.13566282 | -0.068423607361 | 0.04994802 |

| DV | Predictor | Sum_PostModProb_Incl | Sum_PostModProb_Excl | Sum_PriorModProb_Incl | BF_Incl | Frequentist_p_value | postMean | postStd |
|---|---|---|---|---|---|---|---|---|
| alpha_ROI69_AECnodestrength | Blocktapping | 0.3336817 | 0.6663183 | 0.5 | 0.5007842 | 0.40604452 | 0.022045238751 | 0.03232382 |
| alpha_ROI69_AECnodestrength | Sum_PC | 0.4261158 | 0.5738842 | 0.5 | 0.7425119 | 0.14354555 | -0.059937509444 | 0.04603337 |
| alpha_ROI7_AECnodestrength | Blocktapping | 0.3377406 | 0.6622594 | 0.5 | 0.5099824 | 0.45910919 | 0.018679821916 | 0.03224450 |
| alpha_ROI7_AECnodestrength | Sum_PC | 0.4490433 | 0.5509567 | 0.5 | 0.8150247 | 0.13539826 | -0.065686879180 | 0.04981418 |
| alpha_ROI70_AECnodestrength | Blocktapping | 0.3395283 | 0.6604717 | 0.5 | 0.5140694 | 0.34515957 | 0.027765949023 | 0.03304082 |
| alpha_ROI70_AECnodestrength | Sum_PC | 0.3620858 | 0.6379142 | 0.5 | 0.5676091 | 0.24574350 | -0.038026052950 | 0.03910358 |
| alpha_ROI71_AECnodestrength | Blocktapping | 0.3088580 | 0.6911420 | 0.5 | 0.4468808 | 0.41556937 | 0.021775545536 | 0.03271439 |
| alpha_ROI71_AECnodestrength | Sum_PC | 0.3215506 | 0.6784494 | 0.5 | 0.4739492 | 0.34120396 | -0.027906053760 | 0.03588842 |
| alpha_ROI72_AECnodestrength | Blocktapping | 0.2754270 | 0.7245730 | 0.5 | 0.3801233 | 0.75842793 | 0.005820439493 | 0.02820147 |
| alpha_ROI72_AECnodestrength | Sum_PC | 0.3527651 | 0.6472349 | 0.5 | 0.5450340 | 0.24120785 | -0.043172515405 | 0.04046328 |
| alpha_ROI73_AECnodestrength | Blocktapping | 0.2669113 | 0.7330887 | 0.5 | 0.3640915 | 0.90004251 | -0.004059725323 | 0.02782319 |
| alpha_ROI73_AECnodestrength | Sum_PC | 0.3330922 | 0.6669078 | 0.5 | 0.4994575 | 0.30467849 | -0.035247369002 | 0.03901834 |
| alpha_ROI74_AECnodestrength | Blocktapping | 0.4208928 | 0.5791072 | 0.5 | 0.7267961 | 0.26053957 | 0.033947431531 | 0.03546637 |
| alpha_ROI74_AECnodestrength | Sum_PC | 0.5677777 | 0.4322223 | 0.5 | 1.3136245 | 0.06711557 | -0.095664910374 | 0.06047196 |
| alpha_ROI75_AECnodestrength | Blocktapping | 0.2745846 | 0.7254154 | 0.5 | 0.3785204 | 0.71524077 | 0.006878937648 | 0.02667128 |
| alpha_ROI75_AECnodestrength | Sum_PC | 0.3896552 | 0.6103448 | 0.5 | 0.6384180 | 0.15816319 | -0.057593173055 | 0.04553631 |
| alpha_ROI76_AECnodestrength | Blocktapping | 0.2554185 | 0.7445815 | 0.5 | 0.3430363 | 0.89828884 | -0.005072752605 | 0.02679762 |
| alpha_ROI76_AECnodestrength | Sum_PC | 0.3077831 | 0.6922169 | 0.5 | 0.4446338 | 0.33241295 | -0.030720078230 | 0.03597848 |
| alpha_ROI77_AECnodestrength | Blocktapping | 0.2844071 | 0.7155929 | 0.5 | 0.3974426 | 0.77689379 | 0.005385520214 | 0.02674740 |
| alpha_ROI77_AECnodestrength | Sum_PC | 0.3945241 | 0.6054759 | 0.5 | 0.6515934 | 0.18049842 | -0.053866342118 | 0.04529949 |
| alpha_ROI78_AECnodestrength | Blocktapping | 0.3175147 | 0.6824853 | 0.5 | 0.4652330 | 0.58088623 | -0.016671358260 | 0.03053561 |
| alpha_ROI78_AECnodestrength | Sum_PC | 0.4151389 | 0.5848611 | 0.5 | 0.7098076 | 0.18138170 | -0.056396451351 | 0.04501920 |
| alpha_ROI79_AECnodestrength | Blocktapping | 0.2639784 | 0.7360216 | 0.5 | 0.3586558 | 0.58647657 | -0.014195021094 | 0.03029495 |
| alpha_ROI79_AECnodestrength | Sum_PC | 0.2478817 | 0.7521183 | 0.5 | 0.3295781 | 0.99781171 | -0.001103358376 | 0.02873944 |
| alpha_ROI8_AECnodestrength | Blocktapping | 0.3555489 | 0.6444511 | 0.5 | 0.5517080 | 0.72073648 | 0.006695961052 | 0.02752507 |
| alpha_ROI8_AECnodestrength | Sum_PC | 0.6189331 | 0.3810669 | 0.5 | 1.6242114 | 0.04566200 | -0.116764820907 | 0.06574489 |
| alpha_ROI80_AECnodestrength | Blocktapping | 0.2655298 | 0.7344702 | 0.5 | 0.3615256 | 0.97984157 | -0.000312704593 | 0.02655480 |
| alpha_ROI80_AECnodestrength | Sum_PC | 0.3381126 | 0.6618874 | 0.5 | 0.5108309 | 0.27085581 | -0.041795943695 | 0.04039033 |
| alpha_ROI81_AECnodestrength | Blocktapping | 0.2792702 | 0.7207298 | 0.5 | 0.3874825 | 0.76319574 | 0.006278564202 | 0.02762725 |

| DV | Predictor | Sum_PostModProb_Incl | Sum_PostModProb_Excl | Sum_PriorModProb_Incl | BF_Incl | Frequentist_p_value | postMean | postStd |
|---|---|---|---|---|---|---|---|---|
| alpha_ROI81_AECnodestrength | Sum_PC | 0.3713540 | 0.6286460 | 0.5 | 0.5907204 | 0.20176628 | -0.048159113603 | 0.04237080 |
| alpha_ROI82_AECnodestrength | Blocktapping | 0.3368932 | 0.6631068 | 0.5 | 0.5080526 | 0.42130503 | 0.022358993225 | 0.03076354 |
| alpha_ROI82_AECnodestrength | Sum_PC | 0.4450655 | 0.5549345 | 0.5 | 0.8020146 | 0.13866887 | -0.063453973792 | 0.04819872 |
| alpha_ROI83_AECnodestrength | Blocktapping | 0.2536383 | 0.7463617 | 0.5 | 0.3398329 | 0.80888420 | 0.006173399908 | 0.02740376 |
| alpha_ROI83_AECnodestrength | Sum_PC | 0.2661650 | 0.7338350 | 0.5 | 0.3627041 | 0.55721248 | 0.017649053405 | 0.03102460 |
| alpha_ROI84_AECnodestrength | Blocktapping | 0.2415851 | 0.7584149 | 0.5 | 0.3185396 | 0.95708282 | 0.000579606474 | 0.02796596 |
| alpha_ROI84_AECnodestrength | Sum_PC | 0.2556791 | 0.7443209 | 0.5 | 0.3435066 | 0.60287429 | -0.014832087268 | 0.03033853 |
| alpha_ROI85_AECnodestrength | Blocktapping | 0.2647609 | 0.7352391 | 0.5 | 0.3601018 | 0.54719943 | -0.017165122644 | 0.03081543 |
| alpha_ROI85_AECnodestrength | Sum_PC | 0.2437841 | 0.7562159 | 0.5 | 0.3223736 | 0.96046561 | 0.000414489046 | 0.02864231 |
| alpha_ROI86_AECnodestrength | Blocktapping | 0.2810337 | 0.7189663 | 0.5 | 0.3908859 | 0.71337230 | 0.007621701295 | 0.02645950 |
| alpha_ROI86_AECnodestrength | Sum_PC | 0.3619007 | 0.6380993 | 0.5 | 0.5671543 | 0.24215016 | -0.042410938950 | 0.04213114 |
| alpha_ROI87_AECnodestrength | Blocktapping | 0.3221089 | 0.6778911 | 0.5 | 0.4751633 | 0.43953342 | 0.020585002261 | 0.03120521 |
| alpha_ROI87_AECnodestrength | Sum_PC | 0.3883035 | 0.6116965 | 0.5 | 0.6347977 | 0.19864045 | -0.047777748095 | 0.04302482 |
| alpha_ROI88_AECnodestrength | Blocktapping | 0.2417317 | 0.7582683 | 0.5 | 0.3187944 | 0.99707975 | 0.001002401252 | 0.02630925 |
| alpha_ROI88_AECnodestrength | Sum_PC | 0.2477284 | 0.7522716 | 0.5 | 0.3293071 | 0.76021447 | -0.008118363471 | 0.02957526 |
| alpha_ROI89_AECnodestrength | Blocktapping | 0.2810915 | 0.7189085 | 0.5 | 0.3909975 | 0.67788770 | 0.009211153372 | 0.02843197 |
| alpha_ROI89_AECnodestrength | Sum_PC | 0.3439503 | 0.6560497 | 0.5 | 0.5242747 | 0.26339291 | -0.039397422869 | 0.03977238 |
| alpha_ROI9_AECnodestrength | Blocktapping | 0.3029860 | 0.6970140 | 0.5 | 0.4346913 | 0.96236209 | -0.002769448773 | 0.02712135 |
| alpha_ROI9_AECnodestrength | Sum_PC | 0.4741821 | 0.5258179 | 0.5 | 0.9017992 | 0.11253102 | -0.076312258690 | 0.05382604 |
| alpha_ROI90_AECnodestrength | Blocktapping | 0.3691977 | 0.6308023 | 0.5 | 0.5852828 | 0.51665265 | 0.015715894953 | 0.02924590 |
| alpha_ROI90_AECnodestrength | Sum_PC | 0.6147378 | 0.3852622 | 0.5 | 1.5956348 | 0.04758747 | -0.117202877015 | 0.06653817 |
| alpha_ROI91_AECnodestrength | Blocktapping | 0.2808252 | 0.7191748 | 0.5 | 0.3904824 | 0.90728085 | 0.001717130467 | 0.02638810 |
| alpha_ROI91_AECnodestrength | Sum_PC | 0.3920699 | 0.6079301 | 0.5 | 0.6449260 | 0.18001313 | -0.053600744072 | 0.04436987 |
| alpha_ROI92_AECnodestrength | Blocktapping | 0.2751934 | 0.7248066 | 0.5 | 0.3796784 | 0.59602648 | -0.015564002438 | 0.02962887 |
| alpha_ROI92_AECnodestrength | Sum_PC | 0.2879580 | 0.7120420 | 0.5 | 0.4044115 | 0.47418229 | -0.022214525403 | 0.03201344 |
| alpha_ROI93_AECnodestrength | Blocktapping | 0.3040565 | 0.6959435 | 0.5 | 0.4368982 | 0.57338684 | 0.012181713117 | 0.02883073 |
| alpha_ROI93_AECnodestrength | Sum_PC | 0.4137818 | 0.5862182 | 0.5 | 0.7058493 | 0.15448775 | -0.057334967506 | 0.04642782 |
| alpha_ROI94_AECnodestrength | Blocktapping | 0.2491658 | 0.7508342 | 0.5 | 0.3318519 | 0.76404216 | -0.009964219024 | 0.02826784 |
| alpha_ROI94_AECnodestrength | Sum_PC | 0.2477464 | 0.7522536 | 0.5 | 0.3293389 | 0.74619960 | -0.008770321361 | 0.02909827 |

| DV | Predictor | Sum_PostModProb_Incl | Sum_PostModProb_Excl | Sum_PriorModProb_Incl | BF_Incl | Frequentist_p_value | postMean | postStd |
|---|---|---|---|---|---|---|---|---|
| alpha_ROI95_AEC nodestrength | Blocktapping | 0.2589261 | 0.7410739 | 0.5 | 0.3493930 | 0.77673611 | -0.009382516173 | 0.02811417 |
| alpha_ROI95_AEC nodestrength | Sum_PC | 0.2892086 | 0.7107914 | 0.5 | 0.4068824 | 0.42901054 | -0.024597192863 | 0.03333933 |
| alpha_ROI96_AEC nodestrength | Blocktapping | 0.2693765 | 0.7306235 | 0.5 | 0.3686940 | 0.53757135 | -0.016836727875 | 0.03102920 |
| alpha_ROI96_AEC nodestrength | Sum_PC | 0.2487249 | 0.7512751 | 0.5 | 0.3310704 | 0.91504403 | -0.005174579824 | 0.02832469 |
| alpha_ROI97_AEC nodestrength | Blocktapping | 0.2471424 | 0.7528576 | 0.5 | 0.3282724 | 0.91786345 | -0.003050999073 | 0.02825909 |
| alpha_ROI97_AEC nodestrength | Sum_PC | 0.2501624 | 0.7498376 | 0.5 | 0.3336220 | 0.76810340 | 0.007384013822 | 0.02956016 |
| alpha_ROI98_AEC nodestrength | Blocktapping | 0.2480179 | 0.7519821 | 0.5 | 0.3298188 | 0.85306327 | -0.005677666111 | 0.02818679 |
| alpha_ROI98_AEC nodestrength | Sum_PC | 0.2408584 | 0.7591416 | 0.5 | 0.3172772 | 0.82025703 | -0.006844003948 | 0.02759410 |
| alpha_ROI99_AEC nodestrength | Blocktapping | 0.2798686 | 0.7201314 | 0.5 | 0.3886355 | 0.63062341 | 0.011133439264 | 0.02950414 |
| alpha_ROI99_AEC nodestrength | Sum_PC | 0.3213236 | 0.6786764 | 0.5 | 0.4734562 | 0.33627094 | -0.031310427303 | 0.03778299 |
| AlphaPow1 | Blocktapping | 0.3684491 | 0.6315509 | 0.5 | 0.5834037 | 0.21352591 | 0.045012026753 | 0.04039110 |
| AlphaPow1 | Sum_PC | 0.2724689 | 0.7275311 | 0.5 | 0.3745117 | 0.88990825 | -0.002231556447 | 0.02765099 |
| AlphaPow10 | Blocktapping | 0.2367687 | 0.7632313 | 0.5 | 0.3102188 | 0.79684294 | 0.006716082532 | 0.02678602 |
| AlphaPow10 | Sum_PC | 0.2327229 | 0.7672771 | 0.5 | 0.3033101 | 0.96730779 | -0.001352851992 | 0.02673081 |
| AlphaPow100 | Blocktapping | 0.4475329 | 0.5524671 | 0.5 | 0.8100627 | 0.14383809 | 0.056320045537 | 0.04523602 |
| AlphaPow100 | Sum_PC | 0.3838726 | 0.6161274 | 0.5 | 0.6230411 | 0.28344122 | -0.033191524602 | 0.03763025 |
| AlphaPow11 | Blocktapping | 0.2968685 | 0.7031315 | 0.5 | 0.4222091 | 0.41286783 | 0.025504635394 | 0.03239214 |
| AlphaPow11 | Sum_PC | 0.2781781 | 0.7218219 | 0.5 | 0.3853834 | 0.57209351 | 0.017636557942 | 0.03071368 |
| AlphaPow12 | Blocktapping | 0.2725911 | 0.7274089 | 0.5 | 0.3747425 | 0.52964088 | 0.018407103971 | 0.02957755 |
| AlphaPow12 | Sum_PC | 0.2648911 | 0.7351089 | 0.5 | 0.3603427 | 0.57946114 | 0.016244437567 | 0.02892517 |
| AlphaPow13 | Blocktapping | 0.3647539 | 0.6352461 | 0.5 | 0.5741932 | 0.21968484 | 0.044290590850 | 0.03929670 |
| AlphaPow13 | Sum_PC | 0.2683084 | 0.7316916 | 0.5 | 0.3666960 | 0.89329127 | 0.004915708949 | 0.02811947 |
| AlphaPow14 | Blocktapping | 0.4097062 | 0.5902938 | 0.5 | 0.6940717 | 0.16363522 | 0.057962603484 | 0.04436814 |
| AlphaPow14 | Sum_PC | 0.2954247 | 0.7045753 | 0.5 | 0.4192948 | 0.65257798 | 0.014026840017 | 0.02932726 |
| AlphaPow15 | Blocktapping | 0.4944876 | 0.5055124 | 0.5 | 0.9781908 | 0.09966086 | 0.079115187765 | 0.05346319 |
| AlphaPow15 | Sum_PC | 0.3146617 | 0.6853383 | 0.5 | 0.4591334 | 0.94360339 | 0.004290485007 | 0.02821488 |
| AlphaPow16 | Blocktapping | 0.2845957 | 0.7154043 | 0.5 | 0.3978110 | 0.42578434 | 0.021253376229 | 0.03185861 |
| AlphaPow16 | Sum_PC | 0.2532374 | 0.7467626 | 0.5 | 0.3391136 | 0.82077389 | -0.004255386085 | 0.02792317 |
| AlphaPow17 | Blocktapping | 0.2912698 | 0.7087302 | 0.5 | 0.4109742 | 0.39891671 | 0.024610093450 | 0.03245416 |
| AlphaPow17 | Sum_PC | 0.2532902 | 0.7467098 | 0.5 | 0.3392083 | 0.97825165 | 0.001706111951 | 0.02765871 |
| AlphaPow18 | Blocktapping | 0.5474156 | 0.4525844 | 0.5 | 1.2095328 | 0.06932596 | 0.092001051937 | 0.05792019 |
| AlphaPow18 | Sum_PC | 0.3238580 | 0.6761420 | 0.5 | 0.4789793 | 0.77646744 | -0.005871702932 | 0.02824321 |
| AlphaPow19 | Blocktapping | 0.3665074 | 0.6334926 | 0.5 | 0.5785505 | 0.22164306 | 0.046363195093 | 0.03890629 |
| AlphaPow19 | Sum_PC | 0.2743677 | 0.7256323 | 0.5 | 0.3781084 | 0.74360447 | 0.009995904929 | 0.02850931 |
| AlphaPow2 | Blocktapping | 0.3264477 | 0.6735523 | 0.5 | 0.4846656 | 0.30134426 | 0.031882201517 | 0.03578547 |
| AlphaPow2 | Sum_PC | 0.2767457 | 0.7232543 | 0.5 | 0.3826395 | 0.69378493 | -0.009497876130 | 0.02929243 |
| AlphaPow20 | Blocktapping | 0.5459717 | 0.4540283 | 0.5 | 1.2025060 | 0.07016853 | 0.091897611131 | 0.05810015 |

| DV | Predictor | Sum_PostModProb_Incl | Sum_PostModProb_Excl | Sum_PriorModProb_Incl | BF_Incl | Frequentist_p_value | postMean | postStd |
|---|---|---|---|---|---|---|---|---|
| AlphaPow20 | Sum_PC | 0.3545065 | 0.6454935 | 0.5 | 0.5492024 | 0.53377355 | -0.017472111609 | 0.03049564 |
| AlphaPow21 | Blocktapping | 0.5512504 | 0.4487496 | 0.5 | 1.2284141 | 0.06510199 | 0.095032824659 | 0.05726298 |
| AlphaPow21 | Sum_PC | 0.3249239 | 0.6750761 | 0.5 | 0.4813145 | 0.85360259 | -0.002606975362 | 0.02821880 |
| AlphaPow22 | Blocktapping | 0.3570333 | 0.6429667 | 0.5 | 0.5552905 | 0.26673853 | 0.037727236053 | 0.03732947 |
| AlphaPow22 | Sum_PC | 0.3153634 | 0.6846366 | 0.5 | 0.4606290 | 0.46279002 | 0.023884853876 | 0.03227435 |
| AlphaPow23 | Blocktapping | 0.4424208 | 0.5575792 | 0.5 | 0.7934672 | 0.14432298 | 0.062768510831 | 0.04791709 |
| AlphaPow23 | Sum_PC | 0.3173192 | 0.6826808 | 0.5 | 0.4648133 | 0.60425348 | 0.016323356425 | 0.03099972 |
| AlphaPow24 | Blocktapping | 0.2745612 | 0.7254388 | 0.5 | 0.3784760 | 0.46430865 | 0.019489471054 | 0.03115725 |
| AlphaPow24 | Sum_PC | 0.2488917 | 0.7511083 | 0.5 | 0.3313659 | 0.84011521 | -0.004291496060 | 0.02824881 |
| AlphaPow25 | Blocktapping | 0.2843262 | 0.7156738 | 0.5 | 0.3972847 | 0.45157716 | 0.021336817823 | 0.03065079 |
| AlphaPow25 | Sum_PC | 0.2666134 | 0.7333866 | 0.5 | 0.3635373 | 0.59478761 | 0.014402250254 | 0.02893650 |
| AlphaPow26 | Blocktapping | 0.2836224 | 0.7163776 | 0.5 | 0.3959119 | 0.48141375 | 0.018565579707 | 0.03044089 |
| AlphaPow26 | Sum_PC | 0.2758146 | 0.7241854 | 0.5 | 0.3808620 | 0.56313430 | 0.017673420075 | 0.03074217 |
| AlphaPow27 | Blocktapping | 0.2322627 | 0.7677373 | 0.5 | 0.3025289 | 0.78153166 | 0.007749397631 | 0.02664231 |
| AlphaPow27 | Sum_PC | 0.2266173 | 0.7733827 | 0.5 | 0.2930209 | 0.95970116 | -0.001537906450 | 0.02725973 |
| AlphaPow28 | Blocktapping | 0.2862679 | 0.7137321 | 0.5 | 0.4010859 | 0.44350446 | 0.022407317217 | 0.03196573 |
| AlphaPow28 | Sum_PC | 0.2619014 | 0.7380986 | 0.5 | 0.3548325 | 0.70546964 | 0.011313685963 | 0.02912833 |
| AlphaPow29 | Blocktapping | 0.4359345 | 0.5640655 | 0.5 | 0.7728439 | 0.13626938 | 0.063883202719 | 0.04590673 |
| AlphaPow29 | Sum_PC | 0.2948795 | 0.7051205 | 0.5 | 0.4181974 | 0.97494003 | 0.001088491568 | 0.02866141 |
| AlphaPow3 | Blocktapping | 0.3315073 | 0.6684927 | 0.5 | 0.4959026 | 0.28400311 | 0.033403271295 | 0.03653372 |
| AlphaPow3 | Sum_PC | 0.2780600 | 0.7219400 | 0.5 | 0.3851567 | 0.69771038 | -0.009022612610 | 0.02945143 |
| AlphaPow30 | Blocktapping | 0.4059505 | 0.5940495 | 0.5 | 0.6833614 | 0.17288442 | 0.055919849334 | 0.04412071 |
| AlphaPow30 | Sum_PC | 0.2891104 | 0.7108896 | 0.5 | 0.4066882 | 0.83278200 | 0.008008615392 | 0.02818777 |
| AlphaPow31 | Blocktapping | 0.2517404 | 0.7482596 | 0.5 | 0.3364346 | 0.59107272 | 0.014995848432 | 0.02929214 |
| AlphaPow31 | Sum_PC | 0.2420649 | 0.7579351 | 0.5 | 0.3193742 | 0.82330432 | 0.005533780351 | 0.02764017 |
| AlphaPow32 | Blocktapping | 0.3060588 | 0.6939412 | 0.5 | 0.4410442 | 0.34748665 | 0.028284653323 | 0.03367114 |
| AlphaPow32 | Sum_PC | 0.2599048 | 0.7400952 | 0.5 | 0.3511776 | 0.81803669 | 0.007061018516 | 0.02770000 |
| AlphaPow33 | Blocktapping | 0.2924035 | 0.7075965 | 0.5 | 0.4132348 | 0.41514676 | 0.022507656886 | 0.03230915 |
| AlphaPow33 | Sum_PC | 0.2573152 | 0.7426848 | 0.5 | 0.3464663 | 0.89880932 | -0.001703125275 | 0.02812437 |
| AlphaPow34 | Blocktapping | 0.4881025 | 0.5118975 | 0.5 | 0.9535159 | 0.10550669 | 0.076126079391 | 0.05249631 |
| AlphaPow34 | Sum_PC | 0.3161386 | 0.6838614 | 0.5 | 0.4622846 | 0.85750563 | 0.006196210498 | 0.02939189 |
| AlphaPow35 | Blocktapping | 0.2427989 | 0.7572011 | 0.5 | 0.3206531 | 0.70365162 | 0.009636949529 | 0.02695044 |
| AlphaPow35 | Sum_PC | 0.2391872 | 0.7608128 | 0.5 | 0.3143838 | 0.73214521 | 0.009099778528 | 0.02742551 |
| AlphaPow36 | Blocktapping | 0.4480506 | 0.5519494 | 0.5 | 0.8117604 | 0.13472096 | 0.060002315117 | 0.04620539 |
| AlphaPow36 | Sum_PC | 0.3612296 | 0.6387704 | 0.5 | 0.5655076 | 0.34627363 | -0.027463670141 | 0.03580868 |
| AlphaPow37 | Blocktapping | 0.4384549 | 0.5615451 | 0.5 | 0.7808008 | 0.13499303 | 0.066321234710 | 0.04907700 |
| AlphaPow37 | Sum_PC | 0.2935881 | 0.7064119 | 0.5 | 0.4156047 | 0.99318543 | 0.001747782200 | 0.02783853 |
| AlphaPow38 | Blocktapping | 0.2613253 | 0.7386747 | 0.5 | 0.3537759 | 0.56980199 | 0.014046678102 | 0.02784896 |
| AlphaPow38 | Sum_PC | 0.2516634 | 0.7483366 | 0.5 | 0.3362971 | 0.73916433 | 0.009950891828 | 0.02844800 |
| AlphaPow39 | Blocktapping | 0.2699735 | 0.7300265 | 0.5 | 0.3698133 | 0.51524211 | 0.016377825730 | 0.02993903 |
| AlphaPow39 | Sum_PC | 0.2509350 | 0.7490650 | 0.5 | 0.3349976 | 0.86481510 | 0.005908484170 | 0.02737334 |

| DV | Predictor | Sum_PostModProb_Incl | Sum_PostModProb_Excl | Sum_PriorModProb_Incl | BF_Incl | Frequentist_p_value | postMean | postStd |
|---|---|---|---|---|---|---|---|---|
| AlphaPow4 | Blocktapping | 0.3899849 | 0.6100151 | 0.5 | 0.6393036 | 0.19151281 | 0.046197368382 | 0.04114211 |
| AlphaPow4 | Sum_PC | 0.3111067 | 0.6888933 | 0.5 | 0.4516036 | 0.50356375 | -0.017914892230 | 0.03075174 |
| AlphaPow40 | Blocktapping | 0.2821100 | 0.7178900 | 0.5 | 0.3929711 | 0.45067855 | 0.019597310288 | 0.03052163 |
| AlphaPow40 | Sum_PC | 0.2546596 | 0.7453404 | 0.5 | 0.3416689 | 0.89295308 | -0.000873060424 | 0.02836802 |
| AlphaPow41 | Blocktapping | 0.3646308 | 0.6353692 | 0.5 | 0.5738881 | 0.17922932 | 0.048466150091 | 0.04027996 |
| AlphaPow41 | Sum_PC | 0.2660622 | 0.7339378 | 0.5 | 0.3625133 | 0.74730270 | 0.009547651740 | 0.02724633 |
| AlphaPow42 | Blocktapping | 0.2673058 | 0.7326942 | 0.5 | 0.3648259 | 0.53557395 | 0.016168531651 | 0.02922150 |
| AlphaPow42 | Sum_PC | 0.2507020 | 0.7492980 | 0.5 | 0.3345825 | 0.75439960 | 0.008336193878 | 0.02760194 |
| AlphaPow43 | Blocktapping | 0.2512765 | 0.7487235 | 0.5 | 0.3356065 | 0.60302946 | 0.013196956535 | 0.02794899 |
| AlphaPow43 | Sum_PC | 0.2393832 | 0.7606168 | 0.5 | 0.3147226 | 0.85168750 | 0.005805873888 | 0.02659933 |
| AlphaPow44 | Blocktapping | 0.2436081 | 0.7563919 | 0.5 | 0.3220660 | 0.65031434 | 0.011659790583 | 0.02734377 |
| AlphaPow44 | Sum_PC | 0.2392274 | 0.7607726 | 0.5 | 0.3144533 | 0.69541933 | -0.009499386605 | 0.02812191 |
| AlphaPow45 | Blocktapping | 0.2114423 | 0.7885577 | 0.5 | 0.2681380 | 0.99005032 | -0.001090001356 | 0.02382459 |
| AlphaPow45 | Sum_PC | 0.2150537 | 0.7849463 | 0.5 | 0.2739725 | 0.78331408 | -0.006326100882 | 0.02552267 |
| AlphaPow46 | Blocktapping | 0.2250774 | 0.7749226 | 0.5 | 0.2904515 | 0.82929420 | 0.005011230828 | 0.02594250 |
| AlphaPow46 | Sum_PC | 0.2239781 | 0.7760219 | 0.5 | 0.2886234 | 0.99265460 | -0.000024104477 | 0.02546658 |
| AlphaPow47 | Blocktapping | 0.3839895 | 0.6160105 | 0.5 | 0.6233489 | 0.19150923 | 0.051054362368 | 0.04126955 |
| AlphaPow47 | Sum_PC | 0.2803244 | 0.7196756 | 0.5 | 0.3895149 | 0.71284871 | 0.011997505226 | 0.02812357 |
| AlphaPow48 | Blocktapping | 0.2848593 | 0.7151407 | 0.5 | 0.3983263 | 0.41402013 | 0.023462336393 | 0.03192911 |
| AlphaPow48 | Sum_PC | 0.2501841 | 0.7498159 | 0.5 | 0.3336606 | 0.74142587 | 0.009180132981 | 0.02830390 |
| AlphaPow49 | Blocktapping | 0.3498039 | 0.6501961 | 0.5 | 0.5379976 | 0.22893232 | 0.040514868577 | 0.03870302 |
| AlphaPow49 | Sum_PC | 0.2778836 | 0.7221164 | 0.5 | 0.3848183 | 0.63132941 | -0.010083525368 | 0.02897780 |
| AlphaPow5 | Blocktapping | 0.3985467 | 0.6014533 | 0.5 | 0.6626394 | 0.18901614 | 0.046634080777 | 0.04088190 |
| AlphaPow5 | Sum_PC | 0.3358091 | 0.6641909 | 0.5 | 0.5055912 | 0.40737591 | -0.022961009108 | 0.03355352 |
| AlphaPow50 | Blocktapping | 0.4489426 | 0.5510574 | 0.5 | 0.8146931 | 0.13298766 | 0.061080920274 | 0.04610512 |
| AlphaPow50 | Sum_PC | 0.3468386 | 0.6531614 | 0.5 | 0.5310152 | 0.42641384 | -0.021873606169 | 0.03330177 |
| AlphaPow51 | Blocktapping | 0.3108730 | 0.6891270 | 0.5 | 0.4511113 | 0.32801204 | 0.031909419467 | 0.03602272 |
| AlphaPow51 | Sum_PC | 0.2533797 | 0.7466203 | 0.5 | 0.3393689 | 0.92384388 | 0.003688881239 | 0.02823078 |
| AlphaPow52 | Blocktapping | 0.3277283 | 0.6722717 | 0.5 | 0.4874939 | 0.30623149 | 0.032629832555 | 0.03673503 |
| AlphaPow52 | Sum_PC | 0.2674654 | 0.7325346 | 0.5 | 0.3651232 | 0.95936540 | 0.003525874123 | 0.02874094 |
| AlphaPow53 | Blocktapping | 0.2551510 | 0.7448490 | 0.5 | 0.3425540 | 0.79690134 | 0.007007077364 | 0.02727043 |
| AlphaPow53 | Sum_PC | 0.2600047 | 0.7399953 | 0.5 | 0.3513600 | 0.63109315 | 0.013344438713 | 0.02963687 |
| AlphaPow54 | Blocktapping | 0.4059433 | 0.5940567 | 0.5 | 0.6833411 | 0.16573809 | 0.053553609285 | 0.04281112 |
| AlphaPow54 | Sum_PC | 0.3001108 | 0.6998892 | 0.5 | 0.4287976 | 0.61387061 | -0.012180983357 | 0.03034471 |
| AlphaPow55 | Blocktapping | 0.4113781 | 0.5886219 | 0.5 | 0.6988836 | 0.16381945 | 0.049862868441 | 0.04351506 |
| AlphaPow55 | Sum_PC | 0.3250170 | 0.6749830 | 0.5 | 0.4815189 | 0.46751918 | -0.019122324664 | 0.03160187 |
| AlphaPow56 | Blocktapping | 0.3874139 | 0.6125861 | 0.5 | 0.6324237 | 0.19528829 | 0.048412105522 | 0.04146651 |
| AlphaPow56 | Sum_PC | 0.2912144 | 0.7087856 | 0.5 | 0.4108639 | 0.60811655 | -0.011135965760 | 0.02879175 |
| AlphaPow57 | Blocktapping | 0.3386021 | 0.6613979 | 0.5 | 0.5119492 | 0.27174304 | 0.036128509454 | 0.03685366 |
| AlphaPow57 | Sum_PC | 0.2724472 | 0.7275528 | 0.5 | 0.3744706 | 0.75422378 | -0.007209916774 | 0.02833746 |
| AlphaPow58 | Blocktapping | 0.5069288 | 0.4930712 | 0.5 | 1.0281046 | 0.10177220 | 0.071177580425 | 0.04979656 |

| DV | Predictor | Sum_PostModProb_Incl | Sum_PostModProb_Excl | Sum_PriorModProb_Incl | BF_Incl | Frequentist_p_value | postMean | postStd |
|---|---|---|---|---|---|---|---|---|
| AlphaPow58 | Sum_PC | 0.4330335 | 0.5669665 | 0.5 | 0.7637726 | 0.21427216 | -0.043680076708 | 0.04050518 |
| AlphaPow59 | Blocktapping | 0.2562343 | 0.7437657 | 0.5 | 0.3445093 | 0.53683872 | 0.014652621184 | 0.02772049 |
| AlphaPow59 | Sum_PC | 0.2363726 | 0.7636274 | 0.5 | 0.3095391 | 0.94086984 | 0.002896482268 | 0.02623547 |
| AlphaPow6 | Blocktapping | 0.3280365 | 0.6719635 | 0.5 | 0.4881762 | 0.29727464 | 0.033734175189 | 0.03540461 |
| AlphaPow6 | Sum_PC | 0.2765243 | 0.7234757 | 0.5 | 0.3822164 | 0.65241452 | -0.010593771726 | 0.02959701 |
| AlphaPow60 | Blocktapping | 0.2836162 | 0.7163838 | 0.5 | 0.3958998 | 0.41469295 | 0.022689119480 | 0.03075476 |
| AlphaPow60 | Sum_PC | 0.2489732 | 0.7510268 | 0.5 | 0.3315105 | 0.80983465 | 0.007465556157 | 0.02750206 |
| AlphaPow61 | Blocktapping | 0.2512399 | 0.7487601 | 0.5 | 0.3355412 | 0.64075172 | 0.011774434608 | 0.02816544 |
| AlphaPow61 | Sum_PC | 0.2453662 | 0.7546338 | 0.5 | 0.3251461 | 0.74185748 | 0.009854103786 | 0.02776537 |
| AlphaPow62 | Blocktapping | 0.2622291 | 0.7377709 | 0.5 | 0.3554342 | 0.48186910 | 0.018264351678 | 0.02918118 |
| AlphaPow62 | Sum_PC | 0.2371050 | 0.7628950 | 0.5 | 0.3107963 | 0.87049684 | -0.002501377508 | 0.02695920 |
| AlphaPow63 | Blocktapping | 0.2949048 | 0.7050952 | 0.5 | 0.4182483 | 0.35359141 | 0.025954422225 | 0.03299288 |
| AlphaPow63 | Sum_PC | 0.2594929 | 0.7405071 | 0.5 | 0.3504260 | 0.66087706 | -0.010490659414 | 0.02816431 |
| AlphaPow64 | Blocktapping | 0.3811049 | 0.6188951 | 0.5 | 0.6157827 | 0.19177208 | 0.045792827672 | 0.04002203 |
| AlphaPow64 | Sum_PC | 0.3130433 | 0.6869567 | 0.5 | 0.4556958 | 0.44092472 | -0.020994290094 | 0.03181640 |
| AlphaPow65 | Blocktapping | 0.4796463 | 0.5203537 | 0.5 | 0.9217697 | 0.11382290 | 0.064371106879 | 0.04738611 |
| AlphaPow65 | Sum_PC | 0.4076664 | 0.5923336 | 0.5 | 0.6882379 | 0.24479263 | -0.039216715308 | 0.04006616 |
| AlphaPow66 | Blocktapping | 0.4878204 | 0.5121796 | 0.5 | 0.9524400 | 0.10132604 | 0.077816739386 | 0.05155313 |
| AlphaPow66 | Sum_PC | 0.3098884 | 0.6901116 | 0.5 | 0.4490411 | 0.91268831 | -0.001207128313 | 0.02740219 |
| AlphaPow67 | Blocktapping | 0.3376646 | 0.6623354 | 0.5 | 0.5098091 | 0.50097372 | 0.021510412917 | 0.03947544 |
| AlphaPow67 | Sum_PC | 0.2299091 | 0.7700909 | 0.5 | 0.2985481 | 0.90439570 | 0.003769835963 | 0.02358572 |
| AlphaPow68 | Blocktapping | 0.3027033 | 0.6972967 | 0.5 | 0.4341097 | 0.34416185 | 0.024640345655 | 0.03210515 |
| AlphaPow68 | Sum_PC | 0.2808066 | 0.7191934 | 0.5 | 0.3904466 | 0.46024650 | -0.018729018559 | 0.03112835 |
| AlphaPow69 | Blocktapping | 0.4387405 | 0.5612595 | 0.5 | 0.7817071 | 0.16438322 | 0.048536340102 | 0.04184366 |
| AlphaPow69 | Sum_PC | 0.4395314 | 0.5604686 | 0.5 | 0.7842213 | 0.16956080 | -0.052148542000 | 0.04458182 |
| AlphaPow7 | Blocktapping | 0.3285183 | 0.6714817 | 0.5 | 0.4892438 | 0.28946073 | 0.032630823700 | 0.03559042 |
| AlphaPow7 | Sum_PC | 0.2653511 | 0.7346489 | 0.5 | 0.3611944 | 0.98849051 | 0.001540857669 | 0.02719703 |
| AlphaPow70 | Blocktapping | 0.3887177 | 0.6112823 | 0.5 | 0.6359054 | 0.20310220 | 0.041650383099 | 0.03881901 |
| AlphaPow70 | Sum_PC | 0.3494383 | 0.6505617 | 0.5 | 0.5371333 | 0.32969426 | -0.027333229495 | 0.03699599 |
| AlphaPow71 | Blocktapping | 0.4522564 | 0.5477436 | 0.5 | 0.8256717 | 0.12763051 | 0.064642667582 | 0.04743979 |
| AlphaPow71 | Sum_PC | 0.3298215 | 0.6701785 | 0.5 | 0.4921399 | 0.50739417 | -0.016087276694 | 0.03100669 |
| AlphaPow72 | Blocktapping | 0.2973199 | 0.7026801 | 0.5 | 0.4231226 | 0.34874868 | 0.026523993894 | 0.03212386 |
| AlphaPow72 | Sum_PC | 0.2517974 | 0.7482026 | 0.5 | 0.3365364 | 0.82774657 | -0.002445494996 | 0.02763374 |
| AlphaPow73 | Blocktapping | 0.3286674 | 0.6713326 | 0.5 | 0.4895747 | 0.28534320 | 0.036164865151 | 0.03759606 |
| AlphaPow73 | Sum_PC | 0.2599116 | 0.7400884 | 0.5 | 0.3511900 | 0.95010683 | 0.003071261368 | 0.02775856 |
| AlphaPow74 | Blocktapping | 0.2657785 | 0.7342215 | 0.5 | 0.3619867 | 0.53136294 | 0.016507466640 | 0.02975270 |
| AlphaPow74 | Sum_PC | 0.2381924 | 0.7618076 | 0.5 | 0.3126674 | 0.98869168 | 0.000665205958 | 0.02695122 |
| AlphaPow75 | Blocktapping | 0.2697420 | 0.7302580 | 0.5 | 0.3693790 | 0.51511738 | 0.014396402946 | 0.02896974 |
| AlphaPow75 | Sum_PC | 0.2674954 | 0.7325046 | 0.5 | 0.3651791 | 0.54574068 | -0.014964461392 | 0.02892356 |
| AlphaPow76 | Blocktapping | 0.2914792 | 0.7085208 | 0.5 | 0.4113911 | 0.43930353 | 0.022920518477 | 0.03158933 |
| AlphaPow76 | Sum_PC | 0.2824409 | 0.7175591 | 0.5 | 0.3936134 | 0.52755691 | 0.018469495214 | 0.03157585 |

| DV | Predictor | Sum_PostModProb_Incl | Sum_PostModProb_Excl | Sum_PriorModProb_Incl | BF_Incl | Frequentist_p_value | postMean | postStd |
|---|---|---|---|---|---|---|---|---|
| AlphaPow77 | Blocktapping | 0.4510294 | 0.5489706 | 0.5 | 0.8215911 | 0.12350863 | 0.065829512017 | 0.04757975 |
| AlphaPow77 | Sum_PC | 0.3177492 | 0.6822508 | 0.5 | 0.4657367 | 0.58277365 | -0.012695023318 | 0.02995161 |
| AlphaPow78 | Blocktapping | 0.3476135 | 0.6523865 | 0.5 | 0.5328338 | 0.25287613 | 0.041385259618 | 0.03804196 |
| AlphaPow78 | Sum_PC | 0.2660682 | 0.7339318 | 0.5 | 0.3625244 | 0.88462430 | 0.005039359469 | 0.02794508 |
| AlphaPow79 | Blocktapping | 0.2796352 | 0.7203648 | 0.5 | 0.3881856 | 0.48952905 | 0.018644751419 | 0.03077751 |
| AlphaPow79 | Sum_PC | 0.2587493 | 0.7412507 | 0.5 | 0.3490712 | 0.69744626 | 0.011324142877 | 0.02961820 |
| AlphaPow8 | Blocktapping | 0.4203106 | 0.5796894 | 0.5 | 0.7250617 | 0.16185892 | 0.053834695408 | 0.04278684 |
| AlphaPow8 | Sum_PC | 0.3270168 | 0.6729832 | 0.5 | 0.4859211 | 0.46642401 | -0.018654934416 | 0.03261517 |
| AlphaPow80 | Blocktapping | 0.2945372 | 0.7054628 | 0.5 | 0.4175091 | 0.38895905 | 0.024173317636 | 0.03214280 |
| AlphaPow80 | Sum_PC | 0.2603502 | 0.7396498 | 0.5 | 0.3519912 | 0.73708395 | 0.010380398150 | 0.02789717 |
| AlphaPow81 | Blocktapping | 0.3013624 | 0.6986376 | 0.5 | 0.4313573 | 0.40826094 | 0.020286327218 | 0.02982569 |
| AlphaPow81 | Sum_PC | 0.3242085 | 0.6757915 | 0.5 | 0.4797464 | 0.31491245 | -0.030865803148 | 0.03541091 |
| AlphaPow82 | Blocktapping | 0.3452558 | 0.6547442 | 0.5 | 0.5273141 | 0.25976977 | 0.038962888098 | 0.03921164 |
| AlphaPow82 | Sum_PC | 0.2712123 | 0.7287877 | 0.5 | 0.3721417 | 0.87801199 | -0.001587418610 | 0.02835307 |
| AlphaPow83 | Blocktapping | 0.2750244 | 0.7249756 | 0.5 | 0.3793567 | 0.42559061 | 0.022167341796 | 0.03068643 |
| AlphaPow83 | Sum_PC | 0.2401208 | 0.7598792 | 0.5 | 0.3159986 | 0.95241953 | 0.001752128979 | 0.02623280 |
| AlphaPow84 | Blocktapping | 0.2562427 | 0.7437573 | 0.5 | 0.3445247 | 0.57949962 | 0.013085051984 | 0.02767623 |
| AlphaPow84 | Sum_PC | 0.2614852 | 0.7385148 | 0.5 | 0.3540689 | 0.54668521 | -0.016111077230 | 0.02993465 |
| AlphaPow85 | Blocktapping | 0.2466871 | 0.7533129 | 0.5 | 0.3274696 | 0.68262140 | 0.009395403660 | 0.02745930 |
| AlphaPow85 | Sum_PC | 0.2434700 | 0.7565300 | 0.5 | 0.3218246 | 0.73448765 | -0.008049851716 | 0.02765405 |
| AlphaPow86 | Blocktapping | 0.3295625 | 0.6704375 | 0.5 | 0.4915632 | 0.25580935 | 0.037901214106 | 0.03680055 |
| AlphaPow86 | Sum_PC | 0.2540666 | 0.7459334 | 0.5 | 0.3406022 | 0.96942853 | 0.000322016681 | 0.02694047 |
| AlphaPow87 | Blocktapping | 0.4588559 | 0.5411441 | 0.5 | 0.8479367 | 0.11957279 | 0.068625157864 | 0.04774918 |
| AlphaPow87 | Sum_PC | 0.3034728 | 0.6965272 | 0.5 | 0.4356941 | 0.79269237 | -0.005506978431 | 0.02881717 |
| AlphaPow88 | Blocktapping | 0.2578523 | 0.7421477 | 0.5 | 0.3474407 | 0.54917993 | 0.016281149237 | 0.02884526 |
| AlphaPow88 | Sum_PC | 0.2454449 | 0.7545551 | 0.5 | 0.3252843 | 0.71160364 | 0.010193692108 | 0.02825125 |
| AlphaPow89 | Blocktapping | 0.4330947 | 0.5669053 | 0.5 | 0.7639630 | 0.16178661 | 0.052961911530 | 0.04427814 |
| AlphaPow89 | Sum_PC | 0.3903835 | 0.6096165 | 0.5 | 0.6403754 | 0.25832492 | -0.038611056324 | 0.04025475 |
| AlphaPow9 | Blocktapping | 0.4156465 | 0.5843535 | 0.5 | 0.7112929 | 0.16965732 | 0.051733199974 | 0.04379842 |
| AlphaPow9 | Sum_PC | 0.3418356 | 0.6581644 | 0.5 | 0.5193771 | 0.39073544 | -0.024422624853 | 0.03388331 |
| AlphaPow90 | Blocktapping | 0.3039313 | 0.6960687 | 0.5 | 0.4366397 | 0.35889031 | 0.026019749540 | 0.03218690 |
| AlphaPow90 | Sum_PC | 0.2820731 | 0.7179269 | 0.5 | 0.3928995 | 0.51588317 | -0.016692128875 | 0.03029067 |
| AlphaPow91 | Blocktapping | 0.2450205 | 0.7549795 | 0.5 | 0.3245392 | 0.73598497 | 0.008013397581 | 0.02735812 |
| AlphaPow91 | Sum_PC | 0.2427755 | 0.7572245 | 0.5 | 0.3206123 | 0.77598684 | 0.008329449713 | 0.02746714 |
| AlphaPow92 | Blocktapping | 0.2715502 | 0.7284498 | 0.5 | 0.3727782 | 0.55416272 | 0.016058296879 | 0.02928391 |
| AlphaPow92 | Sum_PC | 0.2594422 | 0.7405578 | 0.5 | 0.3503335 | 0.73587981 | 0.011130484282 | 0.02753247 |
| AlphaPow93 | Blocktapping | 0.2508539 | 0.7491461 | 0.5 | 0.3348532 | 0.59733087 | 0.012072940404 | 0.02811323 |
| AlphaPow93 | Sum_PC | 0.2371043 | 0.7628957 | 0.5 | 0.3107952 | 0.92954268 | 0.002684214640 | 0.02627983 |
| AlphaPow94 | Blocktapping | 0.2927091 | 0.7072909 | 0.5 | 0.4138455 | 0.41642046 | 0.022717566744 | 0.03185684 |
| AlphaPow94 | Sum_PC | 0.2688217 | 0.7311783 | 0.5 | 0.3676554 | 0.65099101 | 0.012618748361 | 0.02958581 |
| AlphaPow95 | Blocktapping | 0.3064818 | 0.6935182 | 0.5 | 0.4419233 | 0.37350748 | 0.024243058768 | 0.03348355 |

| DV | Predictor | Sum_PostModProb_Incl | Sum_PostModProb_Excl | Sum_PriorModProb_Incl | BF_Incl | Frequentist_p_value | postMean | postStd |
|---|---|---|---|---|---|---|---|---|
| AlphaPow95 | Sum_PC | 0.2747359 | 0.7252641 | 0.5 | 0.3788082 | 0.54492048 | -0.014467451714 | 0.02970088 |
| AlphaPow96 | Blocktapping | 0.2590125 | 0.7409875 | 0.5 | 0.3495503 | 0.49748774 | 0.017874604319 | 0.02913996 |
| AlphaPow96 | Sum_PC | 0.2357373 | 0.7642627 | 0.5 | 0.3084506 | 0.87116058 | -0.002930350273 | 0.02722218 |
| AlphaPow97 | Blocktapping | 0.2297909 | 0.7702091 | 0.5 | 0.2983487 | 0.78237885 | 0.006093805273 | 0.02683410 |
| AlphaPow97 | Sum_PC | 0.2249191 | 0.7750809 | 0.5 | 0.2901879 | 0.93114074 | -0.001405463808 | 0.02644849 |
| AlphaPow98 | Blocktapping | 0.2688738 | 0.7311262 | 0.5 | 0.3677529 | 0.39248732 | 0.023544470389 | 0.03128900 |
| AlphaPow98 | Sum_PC | 0.2288021 | 0.7711979 | 0.5 | 0.2966840 | 0.95966961 | 0.003626951636 | 0.02607367 |
| AlphaPow99 | Blocktapping | 0.4135581 | 0.5864419 | 0.5 | 0.7051988 | 0.16677176 | 0.050695385187 | 0.04230321 |
| AlphaPow99 | Sum_PC | 0.3396920 | 0.6603080 | 0.5 | 0.5144447 | 0.39515159 | -0.023144271526 | 0.03346374 |
| beta_ROI1_AECnodestrength | Blocktapping | 0.2965380 | 0.7034620 | 0.5 | 0.4215408 | 0.35973684 | -0.027471028925 | 0.03333505 |
| beta_ROI1_AECnodestrength | Sum_PC | 0.2619666 | 0.7380334 | 0.5 | 0.3549522 | 0.58240599 | 0.012953153306 | 0.02979379 |
| beta_ROI10_AECnodestrength | Blocktapping | 0.2334720 | 0.7665280 | 0.5 | 0.3045838 | 0.89026243 | 0.005164839728 | 0.02449097 |
| beta_ROI10_AECnodestrength | Sum_PC | 0.3227354 | 0.6772646 | 0.5 | 0.4765279 | 0.22023540 | 0.045062564985 | 0.03798343 |
| beta_ROI100_AECnodestrength | Blocktapping | 0.2304202 | 0.7695798 | 0.5 | 0.2994105 | 0.93729356 | 0.001966081524 | 0.02578482 |
| beta_ROI100_AECnodestrength | Sum_PC | 0.2411450 | 0.7588550 | 0.5 | 0.3177747 | 0.64468777 | 0.012306978911 | 0.02878553 |
| beta_ROI11_AECnodestrength | Blocktapping | 0.2097237 | 0.7902763 | 0.5 | 0.2653802 | 0.82446666 | -0.005549050004 | 0.02550942 |
| beta_ROI11_AECnodestrength | Sum_PC | 0.2064466 | 0.7935534 | 0.5 | 0.2601547 | 0.89497430 | 0.002335535731 | 0.02439070 |
| beta_ROI12_AECnodestrength | Blocktapping | 0.2266089 | 0.7733911 | 0.5 | 0.2930069 | 0.75585696 | -0.005624525242 | 0.02435054 |
| beta_ROI12_AECnodestrength | Sum_PC | 0.2689727 | 0.7310273 | 0.5 | 0.3679380 | 0.34943008 | 0.027106137288 | 0.03258186 |
| beta_ROI13_AECnodestrength | Blocktapping | 0.2204050 | 0.7795950 | 0.5 | 0.2827174 | 0.72758387 | -0.007936986154 | 0.02541662 |
| beta_ROI13_AECnodestrength | Sum_PC | 0.2123296 | 0.7876704 | 0.5 | 0.2695666 | 0.96296839 | -0.000741983838 | 0.02571479 |
| beta_ROI14_AECnodestrength | Blocktapping | 0.2163218 | 0.7836782 | 0.5 | 0.2760340 | 0.86820594 | 0.001977762764 | 0.02248094 |
| beta_ROI14_AECnodestrength | Sum_PC | 0.2769584 | 0.7230416 | 0.5 | 0.3830462 | 0.29306805 | -0.031568368437 | 0.03274899 |
| beta_ROI15_AECnodestrength | Blocktapping | 0.3068427 | 0.6931573 | 0.5 | 0.4426740 | 0.27926057 | 0.034700344665 | 0.03540101 |
| beta_ROI15_AECnodestrength | Sum_PC | 0.2410428 | 0.7589572 | 0.5 | 0.3175973 | 0.93686809 | 0.003293542388 | 0.02587039 |
| beta_ROI16_AECnodestrength | Blocktapping | 0.2208469 | 0.7791531 | 0.5 | 0.2834449 | 0.85341006 | -0.004730047159 | 0.02549456 |
| beta_ROI16_AECnodestrength | Sum_PC | 0.2170497 | 0.7829503 | 0.5 | 0.2772202 | 0.93769323 | 0.002667442914 | 0.02513654 |
| beta_ROI17_AECnodestrength | Blocktapping | 0.2466569 | 0.7533431 | 0.5 | 0.3274164 | 0.50623748 | 0.014488046342 | 0.02778596 |
| beta_ROI17_AECnodestrength | Sum_PC | 0.2332150 | 0.7667850 | 0.5 | 0.3041465 | 0.62740112 | -0.011497099776 | 0.02631664 |
| beta_ROI18_AECnodestrength | Blocktapping | 0.2286048 | 0.7713952 | 0.5 | 0.2963523 | 0.95901925 | -0.002755531084 | 0.02541048 |
| beta_ROI18_AECnodestrength | Sum_PC | 0.2509008 | 0.7490992 | 0.5 | 0.3349367 | 0.50133765 | -0.018841450832 | 0.03062674 |
| beta_ROI19_AECnodestrength | Blocktapping | 0.2406538 | 0.7593462 | 0.5 | 0.3169223 | 0.69419708 | 0.006511969944 | 0.02458457 |

| DV | Predictor | Sum_PostModProb_Incl | Sum_PostModProb_Excl | Sum_PriorModProb_Incl | BF_Incl | Frequentist_p_value | postMean | postStd |
|---|---|---|---|---|---|---|---|---|
| beta_ROI19_AECnodestrength | Sum_PC | 0.3130082 | 0.6869918 | 0.5 | 0.4556213 | 0.23998424 | -0.038663541553 | 0.03700729 |
| beta_ROI2_AECnodestrength | Blocktapping | 0.2880348 | 0.7119652 | 0.5 | 0.4045629 | 0.44424186 | -0.025019605601 | 0.03183088 |
| beta_ROI2_AECnodestrength | Sum_PC | 0.2744208 | 0.7255792 | 0.5 | 0.3782093 | 0.51889363 | -0.019746350190 | 0.03025184 |
| beta_ROI20_AECnodestrength | Blocktapping | 0.2217707 | 0.7782293 | 0.5 | 0.2849682 | 0.97374781 | -0.001261110250 | 0.02530896 |
| beta_ROI20_AECnodestrength | Sum_PC | 0.2285213 | 0.7714787 | 0.5 | 0.2962120 | 0.70193597 | -0.009664158090 | 0.02769318 |
| beta_ROI21_AECnodestrength | Blocktapping | 0.2292332 | 0.7707668 | 0.5 | 0.2974093 | 0.78468664 | 0.003981343415 | 0.02528560 |
| beta_ROI21_AECnodestrength | Sum_PC | 0.2671412 | 0.7328588 | 0.5 | 0.3645194 | 0.37194536 | -0.025818261655 | 0.03143596 |
| beta_ROI22_AECnodestrength | Blocktapping | 0.2131668 | 0.7868332 | 0.5 | 0.2709174 | 0.82005324 | -0.007058684380 | 0.02599936 |
| beta_ROI22_AECnodestrength | Sum_PC | 0.2106126 | 0.7893874 | 0.5 | 0.2668052 | 0.87875752 | 0.002654847021 | 0.02553529 |
| beta_ROI23_AECnodestrength | Blocktapping | 0.2307374 | 0.7692626 | 0.5 | 0.2999462 | 0.63361075 | -0.014639433375 | 0.02710232 |
| beta_ROI23_AECnodestrength | Sum_PC | 0.2191877 | 0.7808123 | 0.5 | 0.2807176 | 0.91578534 | -0.003429153821 | 0.02529125 |
| beta_ROI24_AECnodestrength | Blocktapping | 0.3127629 | 0.6872371 | 0.5 | 0.4551019 | 0.24888187 | 0.038290203263 | 0.03675504 |
| beta_ROI24_AECnodestrength | Sum_PC | 0.2391570 | 0.7608430 | 0.5 | 0.3143316 | 0.86273284 | 0.005878001479 | 0.02569740 |
| beta_ROI25_AECnodestrength | Blocktapping | 0.2623200 | 0.7376800 | 0.5 | 0.3556013 | 0.45714942 | -0.018983938482 | 0.02856729 |
| beta_ROI25_AECnodestrength | Sum_PC | 0.2769917 | 0.7230083 | 0.5 | 0.3831100 | 0.36246526 | 0.024899150711 | 0.03246492 |
| beta_ROI26_AECnodestrength | Blocktapping | 0.2302145 | 0.7697855 | 0.5 | 0.2990632 | 0.67842904 | -0.008617749271 | 0.02659747 |
| beta_ROI26_AECnodestrength | Sum_PC | 0.2549083 | 0.7450917 | 0.5 | 0.3421166 | 0.43138602 | 0.022018130718 | 0.03123640 |
| beta_ROI27_AECnodestrength | Blocktapping | 0.3561939 | 0.6438061 | 0.5 | 0.5532627 | 0.36708273 | 0.027372459166 | 0.03280636 |
| beta_ROI27_AECnodestrength | Sum_PC | 0.4454478 | 0.5545522 | 0.5 | 0.8032566 | 0.15605981 | 0.064422499426 | 0.04893254 |
| beta_ROI28_AECnodestrength | Blocktapping | 0.2434447 | 0.7565553 | 0.5 | 0.3217805 | 0.97317306 | 0.000980801374 | 0.02557693 |
| beta_ROI28_AECnodestrength | Sum_PC | 0.3304879 | 0.6695121 | 0.5 | 0.4936249 | 0.22954506 | 0.043582387403 | 0.03873042 |
| beta_ROI29_AECnodestrength | Blocktapping | 0.2782196 | 0.7217804 | 0.5 | 0.3854629 | 0.40905638 | 0.020066287926 | 0.02923460 |
| beta_ROI29_AECnodestrength | Sum_PC | 0.2821680 | 0.7178320 | 0.5 | 0.3930836 | 0.39527781 | -0.024263069781 | 0.03246034 |
| beta_ROI3_AECnodestrength | Blocktapping | 0.2849290 | 0.7150710 | 0.5 | 0.3984624 | 0.39203135 | -0.026884584363 | 0.03225415 |
| beta_ROI3_AECnodestrength | Sum_PC | 0.2450828 | 0.7549172 | 0.5 | 0.3246486 | 0.82695892 | -0.006636983790 | 0.02771853 |
| beta_ROI30_AECnodestrength | Blocktapping | 0.2205183 | 0.7794817 | 0.5 | 0.2829038 | 0.83847906 | -0.005646829562 | 0.02573218 |
| beta_ROI30_AECnodestrength | Sum_PC | 0.2186146 | 0.7813854 | 0.5 | 0.2797783 | 0.83255916 | 0.004618273670 | 0.02598238 |
| beta_ROI31_AECnodestrength | Blocktapping | 0.2460681 | 0.7539319 | 0.5 | 0.3263797 | 0.95503710 | -0.000131260402 | 0.02648420 |
| beta_ROI31_AECnodestrength | Sum_PC | 0.3140210 | 0.6859790 | 0.5 | 0.4577705 | 0.29122988 | 0.035869695781 | 0.03557398 |

| DV | Predictor | Sum_PostModProb_Incl | Sum_PostModProb_Excl | Sum_PriorModProb_Incl | BF_Incl | Frequentist_p_value | postMean | postStd |
|---|---|---|---|---|---|---|---|---|
| beta_ROI32_AECnodestrength | Blocktapping | 0.2675216 | 0.7324784 | 0.5 | 0.3652280 | 0.45779049 | -0.019889925097 | 0.02978479 |
| beta_ROI32_AECnodestrength | Sum_PC | 0.2565889 | 0.7434111 | 0.5 | 0.3451507 | 0.52472465 | 0.016613418193 | 0.02938241 |
| beta_ROI33_AECnodestrength | Blocktapping | 0.2615484 | 0.7384516 | 0.5 | 0.3541848 | 0.95975418 | 0.001754132352 | 0.02515365 |
| beta_ROI33_AECnodestrength | Sum_PC | 0.3507404 | 0.6492596 | 0.5 | 0.5402159 | 0.22460682 | 0.042695077102 | 0.04069067 |
| beta_ROI34_AECnodestrength | Blocktapping | 0.2281318 | 0.7718682 | 0.5 | 0.2955580 | 0.82501848 | 0.004280845864 | 0.02614333 |
| beta_ROI34_AECnodestrength | Sum_PC | 0.2543866 | 0.7456134 | 0.5 | 0.3411775 | 0.46858915 | -0.020705376219 | 0.03020739 |
| beta_ROI35_AECnodestrength | Blocktapping | 0.2772477 | 0.7227523 | 0.5 | 0.3835999 | 0.58594249 | 0.014435134120 | 0.02788300 |
| beta_ROI35_AECnodestrength | Sum_PC | 0.3646266 | 0.6353734 | 0.5 | 0.5738775 | 0.17946694 | 0.052238229755 | 0.04131770 |
| beta_ROI36_AECnodestrength | Blocktapping | 0.2353550 | 0.7646450 | 0.5 | 0.3077964 | 0.79286316 | -0.009177483112 | 0.02659487 |
| beta_ROI36_AECnodestrength | Sum_PC | 0.2327545 | 0.7672455 | 0.5 | 0.3033638 | 0.68065178 | -0.009880718622 | 0.02794050 |
| beta_ROI37_AECnodestrength | Blocktapping | 0.2160930 | 0.7839070 | 0.5 | 0.2756615 | 0.79071393 | 0.005407150332 | 0.02472503 |
| beta_ROI37_AECnodestrength | Sum_PC | 0.2085845 | 0.7914155 | 0.5 | 0.2635587 | 0.93772129 | -0.001378188126 | 0.02510781 |
| beta_ROI38_AECnodestrength | Blocktapping | 0.2455624 | 0.7544376 | 0.5 | 0.3254906 | 0.92279714 | -0.002033094198 | 0.02441101 |
| beta_ROI38_AECnodestrength | Sum_PC | 0.3386884 | 0.6613116 | 0.5 | 0.5121464 | 0.21554948 | 0.046037337314 | 0.03927343 |
| beta_ROI39_AECnodestrength | Blocktapping | 0.2817498 | 0.7182502 | 0.5 | 0.3922725 | 0.62084750 | 0.012718865054 | 0.02625983 |
| beta_ROI39_AECnodestrength | Sum_PC | 0.4052767 | 0.5947233 | 0.5 | 0.6814543 | 0.16401220 | 0.061200971996 | 0.04554430 |
| beta_ROI4_AECnodestrength | Blocktapping | 0.2610743 | 0.7389257 | 0.5 | 0.3533161 | 0.63762227 | -0.013416464118 | 0.02807009 |
| beta_ROI4_AECnodestrength | Sum_PC | 0.2871353 | 0.7128647 | 0.5 | 0.4027907 | 0.40451548 | -0.027476122423 | 0.03296210 |
| beta_ROI40_AECnodestrength | Blocktapping | 0.2288498 | 0.7711502 | 0.5 | 0.2967642 | 0.89814748 | 0.002323527470 | 0.02475540 |
| beta_ROI40_AECnodestrength | Sum_PC | 0.2381806 | 0.7618194 | 0.5 | 0.3126470 | 0.66474195 | 0.011237713297 | 0.02848271 |
| beta_ROI41_AECnodestrength | Blocktapping | 0.2272704 | 0.7727296 | 0.5 | 0.2941138 | 0.74954724 | 0.007212651496 | 0.02553970 |
| beta_ROI41_AECnodestrength | Sum_PC | 0.2313364 | 0.7686636 | 0.5 | 0.3009593 | 0.70093440 | 0.010042432676 | 0.02649265 |
| beta_ROI42_AECnodestrength | Blocktapping | 0.2614158 | 0.7385842 | 0.5 | 0.3539418 | 0.76473218 | 0.007952663826 | 0.02542744 |
| beta_ROI42_AECnodestrength | Sum_PC | 0.3722771 | 0.6277229 | 0.5 | 0.5930596 | 0.18337001 | 0.055206035577 | 0.04183534 |
| beta_ROI43_AECnodestrength | Blocktapping | 0.3093389 | 0.6906611 | 0.5 | 0.4478882 | 0.36464940 | 0.027110035667 | 0.03109807 |
| beta_ROI43_AECnodestrength | Sum_PC | 0.3633330 | 0.6366670 | 0.5 | 0.5706798 | 0.22877499 | 0.047254781736 | 0.04102644 |
| beta_ROI44_AECnodestrength | Blocktapping | 0.2462219 | 0.7537781 | 0.5 | 0.3266504 | 0.79386313 | -0.005450189615 | 0.02563584 |
| beta_ROI44_AECnodestrength | Sum_PC | 0.3099718 | 0.6900282 | 0.5 | 0.4492161 | 0.28119050 | 0.036130331863 | 0.03671473 |
| beta_ROI45_AECnodestrength | Blocktapping | 0.3279908 | 0.6720092 | 0.5 | 0.4880749 | 0.69190525 | -0.008113822721 | 0.02357553 |

| DV | Predictor | Sum_PostModProb_Incl | Sum_PostModProb_Excl | Sum_PriorModProb_Incl | BF_Incl | Frequentist_p_value | postMean | postStd |
|---|---|---|---|---|---|---|---|---|
| beta_ROI45_AECnodestrength | Sum_PC | 0.6858830 | 0.3141170 | 0.5 | 2.1835267 | 0.02581091 | 0.145796790479 | 0.07113242 |
| beta_ROI46_AECnodestrength | Blocktapping | 0.3472313 | 0.6527687 | 0.5 | 0.5319361 | 0.56296192 | 0.016469038894 | 0.02972319 |
| beta_ROI46_AECnodestrength | Sum_PC | 0.5340526 | 0.4659474 | 0.5 | 1.1461651 | 0.08919255 | 0.088880146180 | 0.05708609 |
| beta_ROI47_AECnodestrength | Blocktapping | 0.2365628 | 0.7634372 | 0.5 | 0.3098654 | 0.67382378 | 0.011518469392 | 0.02630840 |
| beta_ROI47_AECnodestrength | Sum_PC | 0.2556374 | 0.7443626 | 0.5 | 0.3434312 | 0.49930019 | 0.019348334137 | 0.02993939 |
| beta_ROI48_AECnodestrength | Blocktapping | 0.2395941 | 0.7604059 | 0.5 | 0.3150871 | 0.73445046 | 0.008153771393 | 0.02677331 |
| beta_ROI48_AECnodestrength | Sum_PC | 0.2558902 | 0.7441098 | 0.5 | 0.3438876 | 0.51238832 | 0.019345540860 | 0.03085557 |
| beta_ROI49_AECnodestrength | Blocktapping | 0.2718906 | 0.7281094 | 0.5 | 0.3734200 | 0.44782290 | -0.023133653316 | 0.03021158 |
| beta_ROI49_AECnodestrength | Sum_PC | 0.2459885 | 0.7540115 | 0.5 | 0.3262397 | 0.68139040 | -0.011494946665 | 0.02805690 |
| beta_ROI5_AECnodestrength | Blocktapping | 0.2749618 | 0.7250382 | 0.5 | 0.3792376 | 0.51760299 | -0.019333546262 | 0.03089803 |
| beta_ROI5_AECnodestrength | Sum_PC | 0.2743070 | 0.7256930 | 0.5 | 0.3779932 | 0.47617593 | -0.022318023824 | 0.03094151 |
| beta_ROI50_AECnodestrength | Blocktapping | 0.2192038 | 0.7807962 | 0.5 | 0.2807439 | 0.92579515 | 0.000435438402 | 0.02497664 |
| beta_ROI50_AECnodestrength | Sum_PC | 0.2214290 | 0.7785710 | 0.5 | 0.2844045 | 0.86092355 | -0.004387067133 | 0.02719653 |
| beta_ROI51_AECnodestrength | Blocktapping | 0.2579576 | 0.7420424 | 0.5 | 0.3476319 | 0.55288339 | -0.015852579560 | 0.02927104 |
| beta_ROI51_AECnodestrength | Sum_PC | 0.2388543 | 0.7611457 | 0.5 | 0.3138088 | 0.84048469 | 0.004034366817 | 0.02835433 |
| beta_ROI52_AECnodestrength | Blocktapping | 0.2639641 | 0.7360359 | 0.5 | 0.3586295 | 0.53681259 | -0.016906939747 | 0.02945639 |
| beta_ROI52_AECnodestrength | Sum_PC | 0.2469289 | 0.7530711 | 0.5 | 0.3278959 | 0.76076897 | 0.007951966303 | 0.02831938 |
| beta_ROI53_AECnodestrength | Blocktapping | 0.2481538 | 0.7518462 | 0.5 | 0.3300593 | 0.93023401 | -0.003307279272 | 0.02685569 |
| beta_ROI53_AECnodestrength | Sum_PC | 0.2801374 | 0.7198626 | 0.5 | 0.3891541 | 0.45043735 | 0.021321974823 | 0.03353320 |
| beta_ROI54_AECnodestrength | Blocktapping | 0.2139117 | 0.7860883 | 0.5 | 0.2721217 | 0.90932509 | -0.003287396474 | 0.02415545 |
| beta_ROI54_AECnodestrength | Sum_PC | 0.2269110 | 0.7730890 | 0.5 | 0.2935122 | 0.68104083 | 0.010721868861 | 0.02808313 |
| beta_ROI55_AECnodestrength | Blocktapping | 0.2698788 | 0.7301212 | 0.5 | 0.3696356 | 0.39966345 | -0.025855878345 | 0.03064237 |
| beta_ROI55_AECnodestrength | Sum_PC | 0.2301303 | 0.7698697 | 0.5 | 0.2989210 | 0.81850021 | 0.004387589715 | 0.02577421 |
| beta_ROI56_AECnodestrength | Blocktapping | 0.2770048 | 0.7229952 | 0.5 | 0.3831351 | 0.39705129 | -0.025491685028 | 0.03116228 |
| beta_ROI56_AECnodestrength | Sum_PC | 0.2342146 | 0.7657854 | 0.5 | 0.3058489 | 0.90270977 | 0.001438140069 | 0.02612792 |
| beta_ROI57_AECnodestrength | Blocktapping | 0.2364917 | 0.7635083 | 0.5 | 0.3097435 | 0.73960060 | -0.008777319076 | 0.02677040 |
| beta_ROI57_AECnodestrength | Sum_PC | 0.2421707 | 0.7578293 | 0.5 | 0.3195584 | 0.62221090 | -0.013486492081 | 0.02888946 |
| beta_ROI58_AECnodestrength | Blocktapping | 0.2724540 | 0.7275460 | 0.5 | 0.3744835 | 0.46817521 | -0.022471322882 | 0.03108351 |
| beta_ROI58_AECnodestrength | Sum_PC | 0.2503974 | 0.7496026 | 0.5 | 0.3340402 | 0.59013527 | -0.014659780853 | 0.02814130 |

| DV | Predictor | Sum_PostModProb_Incl | Sum_PostModProb_Excl | Sum_PriorModProb_Incl | BF_Incl | Frequentist_p_value | postMean | postStd |
|---|---|---|---|---|---|---|---|---|
| beta_ROI59_AECnodestrength | Blocktapping | 0.2577754 | 0.7422246 | 0.5 | 0.3473010 | 0.61101312 | 0.012281490857 | 0.02737735 |
| beta_ROI59_AECnodestrength | Sum_PC | 0.2938417 | 0.7061583 | 0.5 | 0.4161130 | 0.37445373 | 0.028828730985 | 0.03363157 |
| beta_ROI6_AECnodestrength | Blocktapping | 0.2847677 | 0.7152323 | 0.5 | 0.3981471 | 0.40619531 | -0.027487629131 | 0.03226855 |
| beta_ROI6_AECnodestrength | Sum_PC | 0.2536183 | 0.7463817 | 0.5 | 0.3397970 | 0.58478267 | -0.014697347859 | 0.02779918 |
| beta_ROI60_AECnodestrength | Blocktapping | 0.2717465 | 0.7282535 | 0.5 | 0.3731482 | 0.57933673 | 0.014549929265 | 0.02708437 |
| beta_ROI60_AECnodestrength | Sum_PC | 0.3365575 | 0.6634425 | 0.5 | 0.5072896 | 0.26198004 | 0.040873540384 | 0.03976741 |
| beta_ROI61_AECnodestrength | Blocktapping | 0.2625645 | 0.7374355 | 0.5 | 0.3560509 | 0.64749788 | 0.011432074013 | 0.02755649 |
| beta_ROI61_AECnodestrength | Sum_PC | 0.3186314 | 0.6813686 | 0.5 | 0.4676344 | 0.30425911 | 0.035130197569 | 0.03724352 |
| beta_ROI62_AECnodestrength | Blocktapping | 0.2852894 | 0.7147106 | 0.5 | 0.3991676 | 0.58584201 | 0.014635545221 | 0.02881817 |
| beta_ROI62_AECnodestrength | Sum_PC | 0.3483670 | 0.6516330 | 0.5 | 0.5346061 | 0.26782322 | 0.040605858815 | 0.03857863 |
| beta_ROI63_AECnodestrength | Blocktapping | 0.3225555 | 0.6774445 | 0.5 | 0.4761357 | 0.45535641 | 0.020805259235 | 0.03033030 |
| beta_ROI63_AECnodestrength | Sum_PC | 0.4077329 | 0.5922671 | 0.5 | 0.6884275 | 0.19115635 | 0.052902082749 | 0.04335804 |
| beta_ROI64_AECnodestrength | Blocktapping | 0.2787108 | 0.7212892 | 0.5 | 0.3864064 | 0.54512022 | 0.015044332983 | 0.02906639 |
| beta_ROI64_AECnodestrength | Sum_PC | 0.3031069 | 0.6968931 | 0.5 | 0.4349403 | 0.38817669 | 0.028099676204 | 0.03320495 |
| beta_ROI65_AECnodestrength | Blocktapping | 0.2829094 | 0.7170906 | 0.5 | 0.3945240 | 0.37111090 | 0.025741899294 | 0.03128095 |
| beta_ROI65_AECnodestrength | Sum_PC | 0.2432886 | 0.7567114 | 0.5 | 0.3215077 | 0.81195342 | 0.007052019702 | 0.02655032 |
| beta_ROI66_AECnodestrength | Blocktapping | 0.3668747 | 0.6331253 | 0.5 | 0.5794661 | 0.20427796 | 0.045836313697 | 0.03864910 |
| beta_ROI66_AECnodestrength | Sum_PC | 0.2879171 | 0.7120829 | 0.5 | 0.4043309 | 0.57740841 | 0.016008570832 | 0.02937495 |
| beta_ROI67_AECnodestrength | Blocktapping | 0.2505930 | 0.7494070 | 0.5 | 0.3343884 | 0.77180345 | -0.006335471993 | 0.02734411 |
| beta_ROI67_AECnodestrength | Sum_PC | 0.2613483 | 0.7386517 | 0.5 | 0.3538180 | 0.50683147 | 0.019601784830 | 0.03090611 |
| beta_ROI68_AECnodestrength | Blocktapping | 0.4177342 | 0.5822658 | 0.5 | 0.7174287 | 0.18835496 | 0.047486629605 | 0.04218400 |
| beta_ROI68_AECnodestrength | Sum_PC | 0.3821474 | 0.6178526 | 0.5 | 0.6185091 | 0.28869063 | 0.037537740895 | 0.03667801 |
| beta_ROI69_AECnodestrength | Blocktapping | 0.2863853 | 0.7136147 | 0.5 | 0.4013165 | 0.37552160 | 0.025554830278 | 0.03222630 |
| beta_ROI69_AECnodestrength | Sum_PC | 0.2446522 | 0.7553478 | 0.5 | 0.3238935 | 0.94009326 | 0.003680387428 | 0.02765147 |
| beta_ROI7_AECnodestrength | Blocktapping | 0.2323221 | 0.7676779 | 0.5 | 0.3026297 | 0.79906779 | 0.004118668381 | 0.02648220 |
| beta_ROI7_AECnodestrength | Sum_PC | 0.2516962 | 0.7483038 | 0.5 | 0.3363557 | 0.53732862 | -0.015776055322 | 0.03040116 |
| beta_ROI70_AECnodestrength | Blocktapping | 0.2128253 | 0.7871747 | 0.5 | 0.2703660 | 0.82101764 | 0.004744614872 | 0.02310152 |
| beta_ROI70_AECnodestrength | Sum_PC | 0.2973133 | 0.7026867 | 0.5 | 0.4231093 | 0.77396631 | 0.011273917244 | 0.03575412 |
| beta_ROI71_AECnodestrength | Blocktapping | 0.2661354 | 0.7338646 | 0.5 | 0.3626492 | 0.48695739 | 0.018503821306 | 0.02996587 |

| DV | Predictor | Sum_PostModProb_Incl | Sum_PostModProb_Excl | Sum_PriorModProb_Incl | BF_Incl | Frequentist_p_value | postMean | postStd |
|---|---|---|---|---|---|---|---|---|
| beta_ROI71_AECnodestrength | Sum_PC | 0.2496260 | 0.7503740 | 0.5 | 0.3326688 | 0.71286564 | 0.010593702484 | 0.02797158 |
| beta_ROI72_AECnodestrength | Blocktapping | 0.3202711 | 0.6797289 | 0.5 | 0.4711748 | 0.69253337 | 0.010142633068 | 0.02675274 |
| beta_ROI72_AECnodestrength | Sum_PC | 0.5440579 | 0.4559421 | 0.5 | 1.1932609 | 0.07282652 | 0.097315735319 | 0.05989710 |
| beta_ROI73_AECnodestrength | Blocktapping | 0.2870575 | 0.7129425 | 0.5 | 0.4026377 | 0.59932742 | 0.013347583882 | 0.02766125 |
| beta_ROI73_AECnodestrength | Sum_PC | 0.3927604 | 0.6072396 | 0.5 | 0.6467965 | 0.17584929 | 0.054828955576 | 0.04288689 |
| beta_ROI74_AECnodestrength | Blocktapping | 0.2317924 | 0.7682076 | 0.5 | 0.3017315 | 0.89411436 | 0.003606381926 | 0.02455324 |
| beta_ROI74_AECnodestrength | Sum_PC | 0.2616205 | 0.7383795 | 0.5 | 0.3543170 | 0.44244250 | 0.020583918043 | 0.03159034 |
| beta_ROI75_AECnodestrength | Blocktapping | 0.2386431 | 0.7613569 | 0.5 | 0.3134445 | 0.83302027 | 0.005126048529 | 0.02448936 |
| beta_ROI75_AECnodestrength | Sum_PC | 0.2933858 | 0.7066142 | 0.5 | 0.4151994 | 0.32261345 | 0.031727531127 | 0.03278663 |
| beta_ROI76_AECnodestrength | Blocktapping | 0.2617312 | 0.7382688 | 0.5 | 0.3545202 | 0.98901730 | 0.000229887591 | 0.02525008 |
| beta_ROI76_AECnodestrength | Sum_PC | 0.3879878 | 0.6120122 | 0.5 | 0.6339544 | 0.16345730 | 0.056541148487 | 0.04658547 |
| beta_ROI77_AECnodestrength | Blocktapping | 0.2444839 | 0.7555161 | 0.5 | 0.3235985 | 0.61937397 | 0.012625386343 | 0.02661216 |
| beta_ROI77_AECnodestrength | Sum_PC | 0.2477807 | 0.7522193 | 0.5 | 0.3293996 | 0.64495269 | 0.013546825225 | 0.02932316 |
| beta_ROI78_AECnodestrength | Blocktapping | 0.2313378 | 0.7686622 | 0.5 | 0.3009616 | 0.95314611 | 0.001009226809 | 0.02530563 |
| beta_ROI78_AECnodestrength | Sum_PC | 0.2702192 | 0.7297808 | 0.5 | 0.3702745 | 0.39841783 | 0.025981298557 | 0.03362347 |
| beta_ROI79_AECnodestrength | Blocktapping | 0.2517702 | 0.7482298 | 0.5 | 0.3364878 | 0.93050974 | 0.002939606586 | 0.02637002 |
| beta_ROI79_AECnodestrength | Sum_PC | 0.3187429 | 0.6812571 | 0.5 | 0.4678747 | 0.30206641 | 0.036367509124 | 0.03736190 |
| beta_ROI8_AECnodestrength | Blocktapping | 0.2986529 | 0.7013471 | 0.5 | 0.4258276 | 0.99970427 | -0.002573192962 | 0.02541055 |
| beta_ROI8_AECnodestrength | Sum_PC | 0.5092447 | 0.4907553 | 0.5 | 1.0376755 | 0.08240060 | -0.090886381043 | 0.05535808 |
| beta_ROI80_AECnodestrength | Blocktapping | 0.2458433 | 0.7541567 | 0.5 | 0.3259843 | 0.93536799 | 0.003697049988 | 0.02546818 |
| beta_ROI80_AECnodestrength | Sum_PC | 0.3021546 | 0.6978454 | 0.5 | 0.4329822 | 0.33074832 | 0.033756822451 | 0.03573841 |
| beta_ROI81_AECnodestrength | Blocktapping | 0.2785425 | 0.7214575 | 0.5 | 0.3860831 | 0.45921618 | 0.019093745483 | 0.02972310 |
| beta_ROI81_AECnodestrength | Sum_PC | 0.2615257 | 0.7384743 | 0.5 | 0.3541433 | 0.67545632 | 0.011667534863 | 0.02870471 |
| beta_ROI82_AECnodestrength | Blocktapping | 0.2287453 | 0.7712547 | 0.5 | 0.2965885 | 0.96426396 | 0.001353527025 | 0.02591737 |
| beta_ROI82_AECnodestrength | Sum_PC | 0.2326408 | 0.7673592 | 0.5 | 0.3031706 | 0.74474063 | 0.008137294772 | 0.02828510 |
| beta_ROI83_AECnodestrength | Blocktapping | 0.4338470 | 0.5661530 | 0.5 | 0.7663070 | 0.32336707 | 0.032226962923 | 0.03451128 |
| beta_ROI83_AECnodestrength | Sum_PC | 0.6635563 | 0.3364437 | 0.5 | 1.9722653 | 0.04695266 | 0.127004127957 | 0.06596075 |
| beta_ROI84_AECnodestrength | Blocktapping | 0.2931481 | 0.7068519 | 0.5 | 0.4147235 | 0.90906615 | -0.001621169317 | 0.02612606 |
| beta_ROI84_AECnodestrength | Sum_PC | 0.4622327 | 0.5377673 | 0.5 | 0.8595403 | 0.10982133 | 0.076366424522 | 0.05173147 |

| DV | Predictor | Sum_PostModProb_Incl | Sum_PostModProb_Excl | Sum_PriorModProb_Incl | BF_Incl | Frequentist_p_value | postMean | postStd |
|---|---|---|---|---|---|---|---|---|
| beta_ROI85_AECnodestrength | Blocktapping | 0.2574312 | 0.7425688 | 0.5 | 0.3466766 | 0.85172008 | 0.004199766040 | 0.02712472 |
| beta_ROI85_AECnodestrength | Sum_PC | 0.3257838 | 0.6742162 | 0.5 | 0.4832038 | 0.27927015 | 0.038544150954 | 0.03786992 |
| beta_ROI86_AECnodestrength | Blocktapping | 0.3636568 | 0.6363432 | 0.5 | 0.5714790 | 0.46071388 | 0.019491662206 | 0.02979481 |
| beta_ROI86_AECnodestrength | Sum_PC | 0.5851897 | 0.4148103 | 0.5 | 1.4107403 | 0.06301519 | 0.109417475362 | 0.06054754 |
| beta_ROI87_AECnodestrength | Blocktapping | 0.2150046 | 0.7849954 | 0.5 | 0.2738927 | 0.82825830 | 0.004273307526 | 0.02520507 |
| beta_ROI87_AECnodestrength | Sum_PC | 0.2152094 | 0.7847906 | 0.5 | 0.2742252 | 0.83568010 | 0.005174593116 | 0.02656729 |
| beta_ROI88_AECnodestrength | Blocktapping | 0.3048270 | 0.6951730 | 0.5 | 0.4384908 | 0.89227199 | -0.002343241483 | 0.02574134 |
| beta_ROI88_AECnodestrength | Sum_PC | 0.5141431 | 0.4858569 | 0.5 | 1.0582192 | 0.08013606 | 0.090218838963 | 0.05748504 |
| beta_ROI89_AECnodestrength | Blocktapping | 0.2211834 | 0.7788166 | 0.5 | 0.2839994 | 0.96519538 | 0.000984692393 | 0.02535721 |
| beta_ROI89_AECnodestrength | Sum_PC | 0.2278879 | 0.7721121 | 0.5 | 0.2951486 | 0.72334043 | 0.009066156986 | 0.02689431 |
| beta_ROI9_AECnodestrength | Blocktapping | 0.2591818 | 0.7408182 | 0.5 | 0.3498588 | 0.54538806 | -0.017954544422 | 0.02900015 |
| beta_ROI9_AECnodestrength | Sum_PC | 0.2515618 | 0.7484382 | 0.5 | 0.3361157 | 0.55792032 | -0.016175904240 | 0.02865727 |
| beta_ROI90_AECnodestrength | Blocktapping | 0.2229851 | 0.7770149 | 0.5 | 0.2869766 | 0.71073075 | 0.007715003496 | 0.02592314 |
| beta_ROI90_AECnodestrength | Sum_PC | 0.2147037 | 0.7852963 | 0.5 | 0.2734047 | 0.86929218 | 0.004505543022 | 0.02575977 |
| beta_ROI91_AECnodestrength | Blocktapping | 0.2559868 | 0.7440132 | 0.5 | 0.3440622 | 0.55024216 | 0.017155781067 | 0.02865929 |
| beta_ROI91_AECnodestrength | Sum_PC | 0.2381313 | 0.7618687 | 0.5 | 0.3125621 | 0.90748262 | 0.002913799670 | 0.02743947 |
| beta_ROI92_AECnodestrength | Blocktapping | 0.2774230 | 0.7225770 | 0.5 | 0.3839355 | 0.67743637 | 0.011594935250 | 0.02754524 |
| beta_ROI92_AECnodestrength | Sum_PC | 0.3574507 | 0.6425493 | 0.5 | 0.5563008 | 0.23951582 | 0.048020797630 | 0.04164272 |
| beta_ROI93_AECnodestrength | Blocktapping | 0.2359621 | 0.7640379 | 0.5 | 0.3088356 | 0.65241228 | 0.010125962282 | 0.02684684 |
| beta_ROI93_AECnodestrength | Sum_PC | 0.2308657 | 0.7691343 | 0.5 | 0.3001630 | 0.72911033 | 0.009858954899 | 0.02780766 |
| beta_ROI94_AECnodestrength | Blocktapping | 0.2830758 | 0.7169242 | 0.5 | 0.3948476 | 0.90954675 | 0.004094402512 | 0.02616168 |
| beta_ROI94_AECnodestrength | Sum_PC | 0.4349528 | 0.5650472 | 0.5 | 0.7697636 | 0.13127354 | 0.071702572232 | 0.04995719 |
| beta_ROI95_AECnodestrength | Blocktapping | 0.2712637 | 0.7287363 | 0.5 | 0.3722384 | 0.89133471 | 0.003820993495 | 0.02535267 |
| beta_ROI95_AECnodestrength | Sum_PC | 0.3936616 | 0.6063384 | 0.5 | 0.6492440 | 0.17733982 | 0.057507911488 | 0.04563022 |
| beta_ROI96_AECnodestrength | Blocktapping | 0.2574694 | 0.7425306 | 0.5 | 0.3467459 | 0.77858624 | -0.006036974224 | 0.02644029 |
| beta_ROI96_AECnodestrength | Sum_PC | 0.3272363 | 0.6727637 | 0.5 | 0.4864060 | 0.26196136 | 0.039823406821 | 0.03854465 |
| beta_ROI97_AECnodestrength | Blocktapping | 0.3067468 | 0.6932532 | 0.5 | 0.4424743 | 0.64996755 | 0.012706221551 | 0.02878264 |
| beta_ROI97_AECnodestrength | Sum_PC | 0.4310577 | 0.5689423 | 0.5 | 0.7576475 | 0.16229115 | 0.064618637644 | 0.04858108 |
| beta_ROI98_AECnodestrength | Blocktapping | 0.3448704 | 0.6551296 | 0.5 | 0.5264154 | 0.45415568 | 0.021543707603 | 0.03029536 |

| DV | Predictor | Sum_PostModProb_Incl | Sum_PostModProb_Excl | Sum_PriorModProb_Incl | BF_Incl | Frequentist_p_value | postMean | postStd |
|---|---|---|---|---|---|---|---|---|
| beta_ROI98_AECnodestrength | Sum_PC | 0.4940560 | 0.5059440 | 0.5 | 0.9765032 | 0.10907143 | 0.083127155767 | 0.05344778 |
| beta_ROI99_AECnodestrength | Blocktapping | 0.2275299 | 0.7724701 | 0.5 | 0.2945485 | 0.66604234 | -0.010864230984 | 0.02679018 |
| beta_ROI99_AECnodestrength | Sum_PC | 0.2178382 | 0.7821618 | 0.5 | 0.2785078 | 0.98367523 | -0.001239675752 | 0.02667701 |
| BetaPow1 | Blocktapping | 0.2758544 | 0.7241456 | 0.5 | 0.3809377 | 0.69548204 | 0.010733588642 | 0.02689686 |
| BetaPow1 | Sum_PC | 0.3091912 | 0.6908088 | 0.5 | 0.4475785 | 0.39496195 | 0.027184292633 | 0.03405002 |
| BetaPow10 | Blocktapping | 0.2856270 | 0.7143730 | 0.5 | 0.3998290 | 0.54349141 | 0.015921035762 | 0.02940134 |
| BetaPow10 | Sum_PC | 0.3079686 | 0.6920314 | 0.5 | 0.4450211 | 0.44738853 | 0.024940165398 | 0.03337307 |
| BetaPow100 | Blocktapping | 0.3178123 | 0.6821877 | 0.5 | 0.4658723 | 0.39881051 | 0.025094964154 | 0.03335346 |
| BetaPow100 | Sum_PC | 0.2709441 | 0.7290559 | 0.5 | 0.3716370 | 0.63586937 | 0.012788504016 | 0.02756165 |
| BetaPow11 | Blocktapping | 0.3139279 | 0.6860721 | 0.5 | 0.4575727 | 0.48455278 | 0.019811500351 | 0.03121505 |
| BetaPow11 | Sum_PC | 0.3483246 | 0.6516754 | 0.5 | 0.5345062 | 0.31655522 | 0.033967419570 | 0.03701283 |
| BetaPow12 | Blocktapping | 0.2767620 | 0.7232380 | 0.5 | 0.3826708 | 0.60814321 | 0.013019063553 | 0.02796391 |
| BetaPow12 | Sum_PC | 0.2933633 | 0.7066367 | 0.5 | 0.4151543 | 0.48072331 | 0.021487177290 | 0.03111386 |
| BetaPow13 | Blocktapping | 0.3199321 | 0.6800679 | 0.5 | 0.4704414 | 0.44230192 | 0.025619988592 | 0.03410210 |
| BetaPow13 | Sum_PC | 0.2517449 | 0.7482551 | 0.5 | 0.3364427 | 0.72350402 | 0.008921776224 | 0.02623965 |
| BetaPow14 | Blocktapping | 0.4982447 | 0.5017553 | 0.5 | 0.9930035 | 0.11005136 | 0.072395139716 | 0.04844472 |
| BetaPow14 | Sum_PC | 0.3654047 | 0.6345953 | 0.5 | 0.5758075 | 0.44581935 | 0.025093674546 | 0.03274244 |
| BetaPow15 | Blocktapping | 0.4609708 | 0.5390292 | 0.5 | 0.8551869 | 0.13983998 | 0.062609713187 | 0.04747788 |
| BetaPow15 | Sum_PC | 0.3641342 | 0.6358658 | 0.5 | 0.5726588 | 0.41663981 | 0.026002508198 | 0.03441074 |
| BetaPow16 | Blocktapping | 0.2721813 | 0.7278187 | 0.5 | 0.3739685 | 0.59874034 | 0.013994885008 | 0.02744140 |
| BetaPow16 | Sum_PC | 0.2788781 | 0.7211219 | 0.5 | 0.3867281 | 0.58137785 | 0.016289623004 | 0.02959647 |
| BetaPow17 | Blocktapping | 0.3224510 | 0.6775490 | 0.5 | 0.4759081 | 0.31244985 | 0.029962998133 | 0.03498747 |
| BetaPow17 | Sum_PC | 0.2688221 | 0.7311779 | 0.5 | 0.3676563 | 0.80495837 | 0.008060889027 | 0.02775382 |
| BetaPow18 | Blocktapping | 0.3781763 | 0.6218237 | 0.5 | 0.6081729 | 0.20446753 | 0.043476816521 | 0.03953677 |
| BetaPow18 | Sum_PC | 0.2855430 | 0.7144570 | 0.5 | 0.3996644 | 0.84188168 | 0.006265623230 | 0.02801814 |
| BetaPow19 | Blocktapping | 0.5017167 | 0.4982833 | 0.5 | 1.0068905 | 0.10690261 | 0.072500769598 | 0.05066894 |
| BetaPow19 | Sum_PC | 0.3559451 | 0.6440549 | 0.5 | 0.5526628 | 0.52028260 | 0.019694862479 | 0.03130816 |
| BetaPow2 | Blocktapping | 0.2814750 | 0.7185250 | 0.5 | 0.3917400 | 0.51612924 | 0.016848969013 | 0.02880429 |
| BetaPow2 | Sum_PC | 0.2748656 | 0.7251344 | 0.5 | 0.3790547 | 0.60557081 | 0.015369865120 | 0.02904047 |
| BetaPow20 | Blocktapping | 0.3937080 | 0.6062920 | 0.5 | 0.6493704 | 0.18157595 | 0.049356884211 | 0.04170134 |
| BetaPow20 | Sum_PC | 0.2873008 | 0.7126992 | 0.5 | 0.4031165 | 0.83023911 | 0.007297297342 | 0.02795388 |
| BetaPow21 | Blocktapping | 0.4596264 | 0.5403736 | 0.5 | 0.8505715 | 0.11968259 | 0.066100833039 | 0.04781723 |
| BetaPow21 | Sum_PC | 0.3110422 | 0.6889578 | 0.5 | 0.4514677 | 0.77603923 | 0.009596753248 | 0.02832726 |
| BetaPow22 | Blocktapping | 0.2822043 | 0.7177957 | 0.5 | 0.3931541 | 0.60093130 | 0.014911702033 | 0.02960178 |
| BetaPow22 | Sum_PC | 0.3008813 | 0.6991187 | 0.5 | 0.4303723 | 0.38390064 | 0.027467887097 | 0.03304589 |
| BetaPow23 | Blocktapping | 0.4209270 | 0.5790730 | 0.5 | 0.7268981 | 0.18693964 | 0.051894909084 | 0.04422156 |
| BetaPow23 | Sum_PC | 0.3527100 | 0.6472900 | 0.5 | 0.5449027 | 0.40449462 | 0.027659514335 | 0.03441663 |
| BetaPow24 | Blocktapping | 0.3362789 | 0.6637211 | 0.5 | 0.5066570 | 0.29300881 | 0.033796687719 | 0.03610001 |
| BetaPow24 | Sum_PC | 0.2719267 | 0.7280733 | 0.5 | 0.3734882 | 0.86585583 | 0.005567255957 | 0.02710721 |
| BetaPow25 | Blocktapping | 0.2686290 | 0.7313710 | 0.5 | 0.3672951 | 0.76522990 | 0.008355635606 | 0.02692595 |

| DV | Predictor | Sum_PostModProb_Incl | Sum_PostModProb_Excl | Sum_PriorModProb_Incl | BF_Incl | Frequentist_p_value | postMean | postStd |
|---|---|---|---|---|---|---|---|---|
| BetaPow25 | Sum_PC | 0.3160369 | 0.6839631 | 0.5 | 0.4620673 | 0.35796571 | 0.029288746722 | 0.03543715 |
| BetaPow26 | Blocktapping | 0.2697668 | 0.7302332 | 0.5 | 0.3694255 | 0.91033573 | 0.002877885243 | 0.02545241 |
| BetaPow26 | Sum_PC | 0.3336696 | 0.6663304 | 0.5 | 0.5007569 | 0.30340530 | 0.035266680829 | 0.03812426 |
| BetaPow27 | Blocktapping | 0.2089908 | 0.7910092 | 0.5 | 0.2642079 | 0.86723948 | -0.002386002354 | 0.02166193 |
| BetaPow27 | Sum_PC | 0.3376803 | 0.6623197 | 0.5 | 0.5098450 | 0.56571671 | 0.022555662902 | 0.04053906 |
| BetaPow28 | Blocktapping | 0.2578994 | 0.7421006 | 0.5 | 0.3475262 | 0.69903053 | 0.009689646246 | 0.02730350 |
| BetaPow28 | Sum_PC | 0.2860183 | 0.7139817 | 0.5 | 0.4005961 | 0.43963051 | 0.025726061495 | 0.03294769 |
| BetaPow29 | Blocktapping | 0.4187060 | 0.5812940 | 0.5 | 0.7202997 | 0.19658193 | 0.047750130113 | 0.04146417 |
| BetaPow29 | Sum_PC | 0.3697091 | 0.6302909 | 0.5 | 0.5865691 | 0.35768792 | 0.031771633101 | 0.03532661 |
| BetaPow3 | Blocktapping | 0.2779000 | 0.7221000 | 0.5 | 0.3848497 | 0.61433786 | 0.012715583898 | 0.02831004 |
| BetaPow3 | Sum_PC | 0.3034332 | 0.6965668 | 0.5 | 0.4356126 | 0.43111689 | 0.025132962785 | 0.03241202 |
| BetaPow30 | Blocktapping | 0.4228462 | 0.5771538 | 0.5 | 0.7326405 | 0.17658674 | 0.055018289062 | 0.04304222 |
| BetaPow30 | Sum_PC | 0.3439633 | 0.6560367 | 0.5 | 0.5243049 | 0.44056732 | 0.024511916501 | 0.03295229 |
| BetaPow31 | Blocktapping | 0.2619887 | 0.7380113 | 0.5 | 0.3549929 | 0.71655574 | -0.008629038515 | 0.02747409 |
| BetaPow31 | Sum_PC | 0.2820503 | 0.7179497 | 0.5 | 0.3928553 | 0.45787651 | 0.020517193853 | 0.03172748 |
| BetaPow32 | Blocktapping | 0.2582379 | 0.7417621 | 0.5 | 0.3481411 | 0.86590805 | 0.005174086711 | 0.02601554 |
| BetaPow32 | Sum_PC | 0.2955727 | 0.7044273 | 0.5 | 0.4195929 | 0.41747007 | 0.025287876560 | 0.03313232 |
| BetaPow33 | Blocktapping | 0.2645426 | 0.7354574 | 0.5 | 0.3596981 | 0.70494166 | 0.009002428026 | 0.02722995 |
| BetaPow33 | Sum_PC | 0.2770306 | 0.7229694 | 0.5 | 0.3831845 | 0.56036832 | 0.015990158266 | 0.03001585 |
| BetaPow34 | Blocktapping | 0.4968587 | 0.5031413 | 0.5 | 0.9875133 | 0.10663363 | 0.071219362000 | 0.04876852 |
| BetaPow34 | Sum_PC | 0.3581309 | 0.6418691 | 0.5 | 0.5579501 | 0.49665376 | 0.022060265618 | 0.03102678 |
| BetaPow35 | Blocktapping | 0.2515163 | 0.7484837 | 0.5 | 0.3360345 | 0.83285968 | -0.003768585034 | 0.02539394 |
| BetaPow35 | Sum_PC | 0.3013339 | 0.6986661 | 0.5 | 0.4312990 | 0.33711880 | 0.031533859681 | 0.03516841 |
| BetaPow36 | Blocktapping | 0.3972366 | 0.6027634 | 0.5 | 0.6590259 | 0.17718251 | 0.050566759639 | 0.04093112 |
| BetaPow36 | Sum_PC | 0.2858931 | 0.7141069 | 0.5 | 0.4003505 | 0.84694345 | 0.006230315874 | 0.02728163 |
| BetaPow37 | Blocktapping | 0.3423068 | 0.6576932 | 0.5 | 0.5204659 | 0.38661624 | 0.025517409889 | 0.03385222 |
| BetaPow37 | Sum_PC | 0.3789655 | 0.6210345 | 0.5 | 0.6102164 | 0.26305087 | 0.042316967803 | 0.04085489 |
| BetaPow38 | Blocktapping | 0.2596245 | 0.7403755 | 0.5 | 0.3506659 | 0.77664785 | 0.007369042766 | 0.02708399 |
| BetaPow38 | Sum_PC | 0.2883222 | 0.7116778 | 0.5 | 0.4051302 | 0.45159996 | 0.022037556918 | 0.03083410 |
| BetaPow39 | Blocktapping | 0.2775777 | 0.7224223 | 0.5 | 0.3842319 | 0.55376878 | 0.015093212659 | 0.02809009 |
| BetaPow39 | Sum_PC | 0.2824810 | 0.7175190 | 0.5 | 0.3936913 | 0.55235165 | 0.017989948559 | 0.03035834 |
| BetaPow4 | Blocktapping | 0.4185443 | 0.5814557 | 0.5 | 0.7198214 | 0.14211161 | 0.055382452815 | 0.04365361 |
| BetaPow4 | Sum_PC | 0.2884284 | 0.7115716 | 0.5 | 0.4053399 | 0.98693663 | 0.002406592196 | 0.02651530 |
| BetaPow40 | Blocktapping | 0.3239608 | 0.6760392 | 0.5 | 0.4792043 | 0.32921414 | 0.029708429590 | 0.03354124 |
| BetaPow40 | Sum_PC | 0.2826105 | 0.7173895 | 0.5 | 0.3939429 | 0.66854079 | 0.012599549418 | 0.02961618 |
| BetaPow41 | Blocktapping | 0.4968475 | 0.5031525 | 0.5 | 0.9874692 | 0.10993347 | 0.069667978037 | 0.04806667 |
| BetaPow41 | Sum_PC | 0.3673298 | 0.6326702 | 0.5 | 0.5806023 | 0.46173770 | 0.024128235860 | 0.03149664 |
| BetaPow42 | Blocktapping | 0.2556822 | 0.7443178 | 0.5 | 0.3435121 | 0.88954609 | -0.002920093285 | 0.02530299 |
| BetaPow42 | Sum_PC | 0.2947990 | 0.7052010 | 0.5 | 0.4180355 | 0.39827180 | 0.024401165431 | 0.03316893 |
| BetaPow43 | Blocktapping | 0.2376759 | 0.7623241 | 0.5 | 0.3117780 | 0.88646562 | -0.003144295004 | 0.02539805 |
| BetaPow43 | Sum_PC | 0.2544442 | 0.7455558 | 0.5 | 0.3412812 | 0.58901595 | 0.014376148534 | 0.02912430 |

| DV | Predictor | Sum_PostModProb_Incl | Sum_PostModProb_Excl | Sum_PriorModProb_Incl | BF_Incl | Frequentist_p_value | postMean | postStd |
|---|---|---|---|---|---|---|---|---|
| BetaPow44 | Blocktapping | 0.2321639 | 0.7678361 | 0.5 | 0.3023612 | 0.84156560 | -0.006224300561 | 0.02591923 |
| BetaPow44 | Sum_PC | 0.2316293 | 0.7683707 | 0.5 | 0.3014551 | 0.88124698 | 0.003423995666 | 0.02726299 |
| BetaPow45 | Blocktapping | 0.2563417 | 0.7436583 | 0.5 | 0.3447036 | 0.59251924 | -0.015252968628 | 0.02787824 |
| BetaPow45 | Sum_PC | 0.2422769 | 0.7577231 | 0.5 | 0.3197434 | 0.76561274 | -0.008158551549 | 0.02694357 |
| BetaPow46 | Blocktapping | 0.2219895 | 0.7780105 | 0.5 | 0.2853297 | 0.97357860 | -0.000910748412 | 0.02556432 |
| BetaPow46 | Sum_PC | 0.2233206 | 0.7766794 | 0.5 | 0.2875325 | 0.83828651 | 0.005121909039 | 0.02698574 |
| BetaPow47 | Blocktapping | 0.2873616 | 0.7126384 | 0.5 | 0.4032362 | 0.38370274 | 0.025662836256 | 0.03142814 |
| BetaPow47 | Sum_PC | 0.2594047 | 0.7405953 | 0.5 | 0.3502651 | 0.61669612 | 0.013590422951 | 0.02849604 |
| BetaPow48 | Blocktapping | 0.2809747 | 0.7190253 | 0.5 | 0.3907717 | 0.48840461 | 0.019211330963 | 0.02910281 |
| BetaPow48 | Sum_PC | 0.2738122 | 0.7261878 | 0.5 | 0.3770542 | 0.53600330 | 0.018086591794 | 0.03056116 |
| BetaPow49 | Blocktapping | 0.3073277 | 0.6926723 | 0.5 | 0.4436841 | 0.44132375 | 0.021321438114 | 0.03080343 |
| BetaPow49 | Sum_PC | 0.3146547 | 0.6853453 | 0.5 | 0.4591185 | 0.41729107 | 0.025416988829 | 0.03315951 |
| BetaPow5 | Blocktapping | 0.3251415 | 0.6748585 | 0.5 | 0.4817920 | 0.29923988 | 0.031724440421 | 0.03292571 |
| BetaPow5 | Sum_PC | 0.2652240 | 0.7347760 | 0.5 | 0.3609590 | 0.92279609 | 0.003368925244 | 0.02650511 |
| BetaPow50 | Blocktapping | 0.3482715 | 0.6517285 | 0.5 | 0.5343814 | 0.29016415 | 0.033406053708 | 0.03579371 |
| BetaPow50 | Sum_PC | 0.2954591 | 0.7045409 | 0.5 | 0.4193639 | 0.58108370 | 0.017237477628 | 0.02955497 |
| BetaPow51 | Blocktapping | 0.2599637 | 0.7400363 | 0.5 | 0.3512851 | 0.89838958 | 0.004357554076 | 0.02619250 |
| BetaPow51 | Sum_PC | 0.3030305 | 0.6969695 | 0.5 | 0.4347831 | 0.38760105 | 0.026894828172 | 0.03432509 |
| BetaPow52 | Blocktapping | 0.2656574 | 0.7343426 | 0.5 | 0.3617623 | 0.67594256 | 0.010965499204 | 0.02732173 |
| BetaPow52 | Sum_PC | 0.2856997 | 0.7143003 | 0.5 | 0.3999714 | 0.52821603 | 0.019058586942 | 0.03070405 |
| BetaPow53 | Blocktapping | 0.2696769 | 0.7303231 | 0.5 | 0.3692570 | 0.92772457 | 0.002141182394 | 0.02551451 |
| BetaPow53 | Sum_PC | 0.3276066 | 0.6723934 | 0.5 | 0.4872245 | 0.32278717 | 0.032140012631 | 0.03424061 |
| BetaPow54 | Blocktapping | 0.3247997 | 0.6752003 | 0.5 | 0.4810420 | 0.30058725 | 0.030126314016 | 0.03334121 |
| BetaPow54 | Sum_PC | 0.2624961 | 0.7375039 | 0.5 | 0.3559250 | 0.95761534 | 0.003097355466 | 0.02584055 |
| BetaPow55 | Blocktapping | 0.2839324 | 0.7160676 | 0.5 | 0.3965161 | 0.44344365 | 0.020590425724 | 0.03012981 |
| BetaPow55 | Sum_PC | 0.2624933 | 0.7375067 | 0.5 | 0.3559199 | 0.72074435 | 0.010433311267 | 0.02801758 |
| BetaPow56 | Blocktapping | 0.2753899 | 0.7246101 | 0.5 | 0.3800525 | 0.59901550 | 0.014433185954 | 0.02814666 |
| BetaPow56 | Sum_PC | 0.2770343 | 0.7229657 | 0.5 | 0.3831915 | 0.56337460 | 0.016107599239 | 0.03032076 |
| BetaPow57 | Blocktapping | 0.3202997 | 0.6797003 | 0.5 | 0.4712366 | 0.31997595 | 0.027398258736 | 0.03284152 |
| BetaPow57 | Sum_PC | 0.2662465 | 0.7337535 | 0.5 | 0.3628556 | 0.84767416 | 0.006886255756 | 0.02710791 |
| BetaPow58 | Blocktapping | 0.3476715 | 0.6523285 | 0.5 | 0.5329699 | 0.22593725 | 0.038452692130 | 0.03614281 |
| BetaPow58 | Sum_PC | 0.2672175 | 0.7327825 | 0.5 | 0.3646614 | 0.83224376 | -0.003089011552 | 0.02610436 |
| BetaPow59 | Blocktapping | 0.2574498 | 0.7425502 | 0.5 | 0.3467102 | 0.98619221 | -0.000086583475 | 0.02529622 |
| BetaPow59 | Sum_PC | 0.3065248 | 0.6934752 | 0.5 | 0.4420126 | 0.36726046 | 0.027770176548 | 0.03365297 |
| BetaPow6 | Blocktapping | 0.3094846 | 0.6905154 | 0.5 | 0.4481935 | 0.42056752 | 0.024038985729 | 0.03131907 |
| BetaPow6 | Sum_PC | 0.3052930 | 0.6947070 | 0.5 | 0.4394558 | 0.46241309 | 0.022622544503 | 0.03165217 |
| BetaPow60 | Blocktapping | 0.2706967 | 0.7293033 | 0.5 | 0.3711717 | 0.86102538 | 0.005321246371 | 0.02707647 |
| BetaPow60 | Sum_PC | 0.3375658 | 0.6624342 | 0.5 | 0.5095839 | 0.28929808 | 0.037976320167 | 0.03690026 |
| BetaPow61 | Blocktapping | 0.2534985 | 0.7465015 | 0.5 | 0.3395820 | 0.74681261 | -0.007985241583 | 0.02646647 |
| BetaPow61 | Sum_PC | 0.2605469 | 0.7394531 | 0.5 | 0.3523507 | 0.60901715 | 0.013419468072 | 0.02862966 |
| BetaPow62 | Blocktapping | 0.2327612 | 0.7672388 | 0.5 | 0.3033752 | 0.92989665 | -0.003479727986 | 0.02483936 |

| DV | Predictor | Sum_PostModProb_Incl | Sum_PostModProb_Excl | Sum_PriorModProb_Incl | BF_Incl | Frequentist_p_value | postMean | postStd |
|---|---|---|---|---|---|---|---|---|
| BetaPow62 | Sum_PC | 0.2306586 | 0.7693414 | 0.5 | 0.2998131 | 0.98152144 | 0.000324329716 | 0.02644548 |
| BetaPow63 | Blocktapping | 0.2616420 | 0.7383580 | 0.5 | 0.3543566 | 0.51726253 | 0.017087946862 | 0.02823842 |
| BetaPow63 | Sum_PC | 0.2495937 | 0.7504063 | 0.5 | 0.3326115 | 0.69597321 | 0.011298035295 | 0.02756261 |
| BetaPow64 | Blocktapping | 0.3569292 | 0.6430708 | 0.5 | 0.5550387 | 0.22254961 | 0.040773245840 | 0.03789448 |
| BetaPow64 | Sum_PC | 0.2735856 | 0.7264144 | 0.5 | 0.3766247 | 0.81978967 | 0.007609273163 | 0.02683121 |
| BetaPow65 | Blocktapping | 0.3578577 | 0.6421423 | 0.5 | 0.5572873 | 0.22601542 | 0.040869710503 | 0.03684602 |
| BetaPow65 | Sum_PC | 0.2730865 | 0.7269135 | 0.5 | 0.3756794 | 0.77230233 | 0.009346090063 | 0.02671247 |
| BetaPow66 | Blocktapping | 0.4624537 | 0.5375463 | 0.5 | 0.8603049 | 0.17002835 | 0.058804290161 | 0.04730253 |
| BetaPow66 | Sum_PC | 0.3453970 | 0.6546030 | 0.5 | 0.5276434 | 0.38325102 | 0.027433513782 | 0.03203921 |
| BetaPow67 | Blocktapping | 0.2859739 | 0.7140261 | 0.5 | 0.4005089 | 0.57800979 | 0.014079031693 | 0.02843018 |
| BetaPow67 | Sum_PC | 0.3164047 | 0.6835953 | 0.5 | 0.4628538 | 0.37533269 | 0.029198949945 | 0.03409540 |
| BetaPow68 | Blocktapping | 0.2937043 | 0.7062957 | 0.5 | 0.4158375 | 0.38314538 | 0.024033755998 | 0.03208484 |
| BetaPow68 | Sum_PC | 0.2502044 | 0.7497956 | 0.5 | 0.3336968 | 0.84506024 | 0.006061891153 | 0.02580650 |
| BetaPow69 | Blocktapping | 0.2796089 | 0.7203911 | 0.5 | 0.3881348 | 0.46018052 | 0.019036933393 | 0.02961317 |
| BetaPow69 | Sum_PC | 0.2525552 | 0.7474448 | 0.5 | 0.3378915 | 0.84643035 | 0.005355539846 | 0.02645115 |
| BetaPow7 | Blocktapping | 0.3680798 | 0.6319202 | 0.5 | 0.5824784 | 0.22529473 | 0.040671874279 | 0.03728035 |
| BetaPow7 | Sum_PC | 0.2890641 | 0.7109359 | 0.5 | 0.4065967 | 0.74518051 | 0.009571756160 | 0.02765281 |
| BetaPow70 | Blocktapping | 0.2571208 | 0.7428792 | 0.5 | 0.3461139 | 0.69926614 | 0.008450016970 | 0.02648258 |
| BetaPow70 | Sum_PC | 0.2559443 | 0.7440557 | 0.5 | 0.3439854 | 0.73846071 | 0.009489780032 | 0.02820749 |
| BetaPow71 | Blocktapping | 0.3214983 | 0.6785017 | 0.5 | 0.4738356 | 0.32075623 | 0.029975083771 | 0.03422279 |
| BetaPow71 | Sum_PC | 0.2800024 | 0.7199976 | 0.5 | 0.3888935 | 0.64270904 | 0.013825038713 | 0.02917031 |
| BetaPow72 | Blocktapping | 0.2365685 | 0.7634315 | 0.5 | 0.3098752 | 0.95730032 | -0.000985773880 | 0.02550439 |
| BetaPow72 | Sum_PC | 0.2529421 | 0.7470579 | 0.5 | 0.3385844 | 0.57923890 | 0.014920175661 | 0.02792100 |
| BetaPow73 | Blocktapping | 0.2887828 | 0.7112172 | 0.5 | 0.4060403 | 0.45616093 | 0.020989032980 | 0.03093850 |
| BetaPow73 | Sum_PC | 0.2981024 | 0.7018976 | 0.5 | 0.4247092 | 0.44305352 | 0.024618267205 | 0.03295266 |
| BetaPow74 | Blocktapping | 0.2933796 | 0.7066204 | 0.5 | 0.4151870 | 0.69351617 | 0.009728191091 | 0.02704356 |
| BetaPow74 | Sum_PC | 0.3718959 | 0.6281041 | 0.5 | 0.5920929 | 0.23978738 | 0.045397298015 | 0.03913758 |
| BetaPow75 | Blocktapping | 0.2827759 | 0.7172241 | 0.5 | 0.3942644 | 0.74768275 | 0.007913523274 | 0.02832406 |
| BetaPow75 | Sum_PC | 0.2892850 | 0.7107150 | 0.5 | 0.4070338 | 0.61155229 | 0.016672385045 | 0.03003826 |
| BetaPow76 | Blocktapping | 0.2630247 | 0.7369753 | 0.5 | 0.3568976 | 0.88791875 | -0.002551735706 | 0.02748765 |
| BetaPow76 | Sum_PC | 0.3207948 | 0.6792052 | 0.5 | 0.4723092 | 0.33165495 | 0.031851983160 | 0.03652235 |
| BetaPow77 | Blocktapping | 0.3380017 | 0.6619983 | 0.5 | 0.5105778 | 0.34119828 | 0.031650384205 | 0.03443086 |
| BetaPow77 | Sum_PC | 0.3294238 | 0.6705762 | 0.5 | 0.4912547 | 0.39988302 | 0.028359761871 | 0.03402549 |
| BetaPow78 | Blocktapping | 0.3376303 | 0.6623697 | 0.5 | 0.5097308 | 0.29983242 | 0.032937662509 | 0.03560762 |
| BetaPow78 | Sum_PC | 0.3121761 | 0.6878239 | 0.5 | 0.4538604 | 0.45420012 | 0.024305562636 | 0.03329021 |
| BetaPow79 | Blocktapping | 0.2460882 | 0.7539118 | 0.5 | 0.3264152 | 0.99756933 | -0.000238379195 | 0.02636971 |
| BetaPow79 | Sum_PC | 0.2707335 | 0.7292665 | 0.5 | 0.3712408 | 0.50855621 | 0.020062772860 | 0.03145690 |
| BetaPow8 | Blocktapping | 0.3838896 | 0.6161104 | 0.5 | 0.6230858 | 0.18712838 | 0.045750138907 | 0.04073402 |
| BetaPow8 | Sum_PC | 0.2812959 | 0.7187041 | 0.5 | 0.3913932 | 0.94271011 | 0.003331178140 | 0.02741391 |
| BetaPow80 | Blocktapping | 0.2627705 | 0.7372295 | 0.5 | 0.3564298 | 0.96554926 | 0.000337246449 | 0.02671819 |
| BetaPow80 | Sum_PC | 0.3103376 | 0.6896624 | 0.5 | 0.4499848 | 0.35360209 | 0.029739642684 | 0.03640753 |

| DV | Predictor | Sum_PostModProb_Incl | Sum_PostModProb_Excl | Sum_PriorModProb_Incl | BF_Incl | Frequentist_p_value | postMean | postStd |
|---|---|---|---|---|---|---|---|---|
| BetaPow81 | Blocktapping | 0.2794900 | 0.7205100 | 0.5 | 0.3879058 | 0.44764066 | 0.018510039622 | 0.02951093 |
| BetaPow81 | Sum_PC | 0.2464510 | 0.7535490 | 0.5 | 0.3270536 | 0.99472124 | 0.000334114805 | 0.02644896 |
| BetaPow82 | Blocktapping | 0.2650848 | 0.7349152 | 0.5 | 0.3607012 | 0.71333141 | 0.008150892298 | 0.02715000 |
| BetaPow82 | Sum_PC | 0.2883254 | 0.7116746 | 0.5 | 0.4051365 | 0.47913959 | 0.021156944181 | 0.03153935 |
| BetaPow83 | Blocktapping | 0.2325900 | 0.7674100 | 0.5 | 0.3030844 | 0.98156111 | -0.001013456748 | 0.02652418 |
| BetaPow83 | Sum_PC | 0.2358018 | 0.7641982 | 0.5 | 0.3085610 | 0.83098947 | -0.005344022043 | 0.02785462 |
| BetaPow84 | Blocktapping | 0.2359998 | 0.7640002 | 0.5 | 0.3089003 | 0.88547623 | -0.003822173406 | 0.02680379 |
| BetaPow84 | Sum_PC | 0.2332886 | 0.7667114 | 0.5 | 0.3042717 | 0.93404538 | -0.001167479658 | 0.02778563 |
| BetaPow85 | Blocktapping | 0.2307189 | 0.7692811 | 0.5 | 0.2999149 | 0.93880168 | -0.002495505762 | 0.02624791 |
| BetaPow85 | Sum_PC | 0.2289623 | 0.7710377 | 0.5 | 0.2969534 | 0.83491885 | -0.005866264520 | 0.02653205 |
| BetaPow86 | Blocktapping | 0.2307024 | 0.7692976 | 0.5 | 0.2998870 | 0.77829422 | 0.006011610271 | 0.02558120 |
| BetaPow86 | Sum_PC | 0.2344486 | 0.7655514 | 0.5 | 0.3062479 | 0.74239854 | 0.007434594273 | 0.02681401 |
| BetaPow87 | Blocktapping | 0.3222765 | 0.6777235 | 0.5 | 0.4755280 | 0.44226901 | 0.022408890417 | 0.03137034 |
| BetaPow87 | Sum_PC | 0.3583325 | 0.6416675 | 0.5 | 0.5584396 | 0.30218240 | 0.037911881983 | 0.03910631 |
| BetaPow88 | Blocktapping | 0.2378824 | 0.7621176 | 0.5 | 0.3121335 | 0.96849579 | 0.001742020513 | 0.02535854 |
| BetaPow88 | Sum_PC | 0.2593613 | 0.7406387 | 0.5 | 0.3501860 | 0.52638577 | 0.016793639523 | 0.03036961 |
| BetaPow89 | Blocktapping | 0.3208969 | 0.6791031 | 0.5 | 0.4725305 | 0.32120167 | 0.028099510480 | 0.03420718 |
| BetaPow89 | Sum_PC | 0.2707677 | 0.7292323 | 0.5 | 0.3713051 | 0.78373865 | 0.009485493628 | 0.02746038 |
| BetaPow9 | Blocktapping | 0.3898621 | 0.6101379 | 0.5 | 0.6389737 | 0.17572953 | 0.048325622413 | 0.04006889 |
| BetaPow9 | Sum_PC | 0.2838003 | 0.7161997 | 0.5 | 0.3962586 | 0.81757786 | 0.007250458493 | 0.02630087 |
| BetaPow90 | Blocktapping | 0.2796397 | 0.7203603 | 0.5 | 0.3881942 | 0.59003378 | 0.013997792960 | 0.02713699 |
| BetaPow90 | Sum_PC | 0.2956266 | 0.7043734 | 0.5 | 0.4197016 | 0.46867301 | 0.021540949530 | 0.03055535 |
| BetaPow91 | Blocktapping | 0.2664631 | 0.7335369 | 0.5 | 0.3632580 | 0.99377345 | 0.001352456309 | 0.02518674 |
| BetaPow91 | Sum_PC | 0.3280275 | 0.6719725 | 0.5 | 0.4881561 | 0.30987759 | 0.033145843694 | 0.03561044 |
| BetaPow92 | Blocktapping | 0.2693467 | 0.7306533 | 0.5 | 0.3686382 | 0.86628869 | -0.003363110074 | 0.02825609 |
| BetaPow92 | Sum_PC | 0.2947729 | 0.7052271 | 0.5 | 0.4179829 | 0.38472335 | 0.024625891001 | 0.03291273 |
| BetaPow93 | Blocktapping | 0.2669072 | 0.7330928 | 0.5 | 0.3640838 | 0.89142436 | 0.004071578084 | 0.02541241 |
| BetaPow93 | Sum_PC | 0.3365039 | 0.6634961 | 0.5 | 0.5071679 | 0.27619844 | 0.035938280372 | 0.03765875 |
| BetaPow94 | Blocktapping | 0.3333567 | 0.6666433 | 0.5 | 0.5000525 | 0.88046790 | -0.005538279386 | 0.03716562 |
| BetaPow94 | Sum_PC | 0.2599965 | 0.7400035 | 0.5 | 0.3513450 | 0.44348026 | 0.019042265446 | 0.02676693 |
| BetaPow95 | Blocktapping | 0.2475977 | 0.7524023 | 0.5 | 0.3290763 | 0.79445673 | -0.006484076740 | 0.02648122 |
| BetaPow95 | Sum_PC | 0.2495666 | 0.7504334 | 0.5 | 0.3325632 | 0.72457633 | 0.008131252313 | 0.02794715 |
| BetaPow96 | Blocktapping | 0.2252308 | 0.7747692 | 0.5 | 0.2907069 | 0.99216883 | -0.000513142843 | 0.02493083 |
| BetaPow96 | Sum_PC | 0.2249583 | 0.7750417 | 0.5 | 0.2902532 | 0.91206832 | 0.002144274411 | 0.02643397 |
| BetaPow97 | Blocktapping | 0.2259884 | 0.7740116 | 0.5 | 0.2919703 | 0.80226323 | -0.006018374854 | 0.02498037 |
| BetaPow97 | Sum_PC | 0.2220878 | 0.7779122 | 0.5 | 0.2854921 | 0.87387699 | -0.003826377573 | 0.02664904 |
| BetaPow98 | Blocktapping | 0.2264632 | 0.7735368 | 0.5 | 0.2927634 | 0.82907041 | 0.004476728388 | 0.02591335 |
| BetaPow98 | Sum_PC | 0.2265522 | 0.7734478 | 0.5 | 0.2929120 | 0.84745585 | 0.005608205301 | 0.02714155 |
| BetaPow99 | Blocktapping | 0.2763842 | 0.7236158 | 0.5 | 0.3819488 | 0.53923728 | 0.015658390316 | 0.02850647 |
| BetaPow99 | Sum_PC | 0.2722607 | 0.7277393 | 0.5 | 0.3741185 | 0.60254110 | 0.015067782017 | 0.02988712 |

| DV | Predictor | Sum_PostModProb_Incl | Sum_PostModProb_Excl | Sum_PriorModProb_Incl | BF_Incl | Frequentist_p_value | postMean | postStd |
|---|---|---|---|---|---|---|---|---|
| gamma_ROI1_AECnodestrength | Blocktapping | 0.2380525 | 0.7619475 | 0.5 | 0.3124264 | 0.99558678 | 0.001465912483 | 0.02657461 |
| gamma_ROI1_AECnodestrength | Sum_PC | 0.2437359 | 0.7562641 | 0.5 | 0.3222894 | 0.79101517 | 0.005643497178 | 0.02940600 |
| gamma_ROI10_AECnodestrength | Blocktapping | 0.2680774 | 0.7319226 | 0.5 | 0.3662647 | 0.68610485 | 0.008676974646 | 0.02887621 |
| gamma_ROI10_AECnodestrength | Sum_PC | 0.3035445 | 0.6964555 | 0.5 | 0.4358420 | 0.36369579 | -0.028566532145 | 0.03585913 |
| gamma_ROI100_AECnodestrength | Blocktapping | 0.2742104 | 0.7257896 | 0.5 | 0.3778098 | 0.62215212 | -0.014757316524 | 0.03037743 |
| gamma_ROI100_AECnodestrength | Sum_PC | 0.2844405 | 0.7155595 | 0.5 | 0.3975078 | 0.46116433 | -0.024083851859 | 0.03277389 |
| gamma_ROI11_AECnodestrength | Blocktapping | 0.2391092 | 0.7608908 | 0.5 | 0.3142491 | 0.91085263 | -0.003635210135 | 0.02722203 |
| gamma_ROI11_AECnodestrength | Sum_PC | 0.2501261 | 0.7498739 | 0.5 | 0.3335575 | 0.64658742 | -0.012194099704 | 0.02938852 |
| gamma_ROI12_AECnodestrength | Blocktapping | 0.2570056 | 0.7429944 | 0.5 | 0.3459052 | 0.62670000 | 0.011949224742 | 0.02797573 |
| gamma_ROI12_AECnodestrength | Sum_PC | 0.2579141 | 0.7420859 | 0.5 | 0.3475529 | 0.59785955 | -0.012746619612 | 0.02980478 |
| gamma_ROI13_AECnodestrength | Blocktapping | 0.2407285 | 0.7592715 | 0.5 | 0.3170520 | 0.97408585 | -0.001588979114 | 0.02615758 |
| gamma_ROI13_AECnodestrength | Sum_PC | 0.2697761 | 0.7302239 | 0.5 | 0.3694429 | 0.46205814 | -0.020672673534 | 0.03264494 |
| gamma_ROI14_AECnodestrength | Blocktapping | 0.2757581 | 0.7242419 | 0.5 | 0.3807541 | 0.58554571 | -0.016682780680 | 0.02943857 |
| gamma_ROI14_AECnodestrength | Sum_PC | 0.2924987 | 0.7075013 | 0.5 | 0.4134249 | 0.42882199 | -0.025529854231 | 0.03246210 |
| gamma_ROI15_AECnodestrength | Blocktapping | 0.2987677 | 0.7012323 | 0.5 | 0.4260610 | 0.58400068 | -0.017316281509 | 0.02773382 |
| gamma_ROI15_AECnodestrength | Sum_PC | 0.3957494 | 0.6042506 | 0.5 | 0.6549426 | 0.18400516 | -0.054356607727 | 0.04371529 |
| gamma_ROI16_AECnodestrength | Blocktapping | 0.2270747 | 0.7729253 | 0.5 | 0.2937860 | 0.87850491 | 0.002523667731 | 0.02426970 |
| gamma_ROI16_AECnodestrength | Sum_PC | 0.2569269 | 0.7430731 | 0.5 | 0.3457626 | 0.43866834 | -0.021437764902 | 0.03170356 |
| gamma_ROI17_AECnodestrength | Blocktapping | 0.2609622 | 0.7390378 | 0.5 | 0.3531107 | 0.65759491 | -0.011388583468 | 0.02811070 |
| gamma_ROI17_AECnodestrength | Sum_PC | 0.2770517 | 0.7229483 | 0.5 | 0.3832248 | 0.47601368 | -0.022577261208 | 0.03257831 |
| gamma_ROI18_AECnodestrength | Blocktapping | 0.2642224 | 0.7357776 | 0.5 | 0.3591063 | 0.65970109 | -0.013028554886 | 0.02884500 |
| gamma_ROI18_AECnodestrength | Sum_PC | 0.2893404 | 0.7106596 | 0.5 | 0.4071434 | 0.44306170 | -0.023414851413 | 0.03417537 |
| gamma_ROI19_AECnodestrength | Blocktapping | 0.3749507 | 0.6250493 | 0.5 | 0.5998738 | 0.36534422 | -0.030166399758 | 0.03396163 |
| gamma_ROI19_AECnodestrength | Sum_PC | 0.4674811 | 0.5325189 | 0.5 | 0.8778675 | 0.13975221 | -0.069870283933 | 0.04988131 |
| gamma_ROI2_AECnodestrength | Blocktapping | 0.2446009 | 0.7553991 | 0.5 | 0.3238036 | 0.67026957 | -0.011756630687 | 0.02756296 |
| gamma_ROI2_AECnodestrength | Sum_PC | 0.2323430 | 0.7676570 | 0.5 | 0.3026652 | 0.92820308 | -0.002703037876 | 0.02631535 |
| gamma_ROI20_AECnodestrength | Blocktapping | 0.4450658 | 0.5549342 | 0.5 | 0.8020154 | 0.41540111 | -0.025129750304 | 0.03317816 |
| gamma_ROI20_AECnodestrength | Sum_PC | 0.7390526 | 0.2609474 | 0.5 | 2.8321905 | 0.02788283 | -0.158476030046 | 0.07842667 |
| gamma_ROI21_AECnodestrength | Blocktapping | 0.3226813 | 0.6773187 | 0.5 | 0.4764098 | 0.96058471 | -0.003232757296 | 0.02586403 |

| DV | Predictor | Sum_PostModProb_Incl | Sum_PostModProb_Excl | Sum_PriorModProb_Incl | BF_Incl | Frequentist_p_value | postMean | postStd |
|---|---|---|---|---|---|---|---|---|
| gamma_ROI21_AECnodestrength | Sum_PC | 0.5536844 | 0.4463156 | 0.5 | 1.2405669 | 0.06639753 | -0.101730710062 | 0.06127218 |
| gamma_ROI22_AECnodestrength | Blocktapping | 0.2479923 | 0.7520077 | 0.5 | 0.3297735 | 0.98765190 | 0.001767666897 | 0.02627198 |
| gamma_ROI22_AECnodestrength | Sum_PC | 0.2777948 | 0.7222052 | 0.5 | 0.3846480 | 0.50281980 | 0.020173425983 | 0.03286554 |
| gamma_ROI23_AECnodestrength | Blocktapping | 0.2386826 | 0.7613174 | 0.5 | 0.3135126 | 0.75821218 | 0.004459321313 | 0.02564358 |
| gamma_ROI23_AECnodestrength | Sum_PC | 0.2639207 | 0.7360793 | 0.5 | 0.3585493 | 0.39144104 | -0.023960493982 | 0.03227313 |
| gamma_ROI24_AECnodestrength | Blocktapping | 0.3882369 | 0.6117631 | 0.5 | 0.6346197 | 0.38896040 | -0.027153842161 | 0.03402343 |
| gamma_ROI24_AECnodestrength | Sum_PC | 0.5206217 | 0.4793783 | 0.5 | 1.0860351 | 0.11109258 | -0.083822122071 | 0.05521046 |
| gamma_ROI25_AECnodestrength | Blocktapping | 0.2498067 | 0.7501933 | 0.5 | 0.3329897 | 0.87798340 | -0.003867068341 | 0.02738266 |
| gamma_ROI25_AECnodestrength | Sum_PC | 0.2674936 | 0.7325064 | 0.5 | 0.3651758 | 0.54100191 | 0.017795812260 | 0.03203418 |
| gamma_ROI26_AECnodestrength | Blocktapping | 0.2789113 | 0.7210887 | 0.5 | 0.3867920 | 0.80005845 | -0.003680875977 | 0.02706395 |
| gamma_ROI26_AECnodestrength | Sum_PC | 0.3688674 | 0.6311326 | 0.5 | 0.5844532 | 0.21054295 | 0.046384966265 | 0.04228689 |
| gamma_ROI27_AECnodestrength | Blocktapping | 0.3524707 | 0.6475293 | 0.5 | 0.5443317 | 0.31736011 | -0.029423189980 | 0.03506741 |
| gamma_ROI27_AECnodestrength | Sum_PC | 0.3929249 | 0.6070751 | 0.5 | 0.6472428 | 0.20237466 | 0.047883450933 | 0.04319933 |
| gamma_ROI28_AECnodestrength | Blocktapping | 0.3007734 | 0.6992266 | 0.5 | 0.4301515 | 0.41237893 | -0.026465048744 | 0.03416205 |
| gamma_ROI28_AECnodestrength | Sum_PC | 0.2594400 | 0.7405600 | 0.5 | 0.3503294 | 0.87894225 | -0.005449270425 | 0.02920701 |
| gamma_ROI29_AECnodestrength | Blocktapping | 0.2867240 | 0.7132760 | 0.5 | 0.4019818 | 0.63366740 | -0.015342306996 | 0.02886723 |
| gamma_ROI29_AECnodestrength | Sum_PC | 0.3574637 | 0.6425363 | 0.5 | 0.5563324 | 0.23411284 | -0.045459914898 | 0.04079525 |
| gamma_ROI3_AECnodestrength | Blocktapping | 0.2709265 | 0.7290735 | 0.5 | 0.3716038 | 0.46699515 | -0.019628334053 | 0.03033317 |
| gamma_ROI3_AECnodestrength | Sum_PC | 0.2432562 | 0.7567438 | 0.5 | 0.3214512 | 0.79571876 | 0.005576683843 | 0.02714989 |
| gamma_ROI30_AECnodestrength | Blocktapping | 0.2937494 | 0.7062506 | 0.5 | 0.4159280 | 0.58016334 | -0.016698312257 | 0.03034238 |
| gamma_ROI30_AECnodestrength | Sum_PC | 0.3246624 | 0.6753376 | 0.5 | 0.4807409 | 0.34580060 | -0.033081035343 | 0.03686933 |
| gamma_ROI31_AECnodestrength | Blocktapping | 0.2811184 | 0.7188816 | 0.5 | 0.3910496 | 0.57488604 | -0.013584731204 | 0.02843238 |
| gamma_ROI31_AECnodestrength | Sum_PC | 0.3312662 | 0.6687338 | 0.5 | 0.4953633 | 0.28094259 | 0.037069209399 | 0.03803874 |
| gamma_ROI32_AECnodestrength | Blocktapping | 0.2668259 | 0.7331741 | 0.5 | 0.3639326 | 0.57991512 | -0.013384569532 | 0.02962750 |
| gamma_ROI32_AECnodestrength | Sum_PC | 0.2647566 | 0.7352434 | 0.5 | 0.3600938 | 0.62058979 | 0.012263451742 | 0.03043166 |
| gamma_ROI33_AECnodestrength | Blocktapping | 0.2413125 | 0.7586875 | 0.5 | 0.3180657 | 0.91651539 | -0.002818500685 | 0.02700572 |
| gamma_ROI33_AECnodestrength | Sum_PC | 0.2511147 | 0.7488853 | 0.5 | 0.3353179 | 0.64409664 | -0.012584090852 | 0.02991355 |
| gamma_ROI34_AECnodestrength | Blocktapping | 0.2885278 | 0.7114722 | 0.5 | 0.4055363 | 0.93430267 | -0.003738067007 | 0.02537379 |
| gamma_ROI34_AECnodestrength | Sum_PC | 0.4600515 | 0.5399485 | 0.5 | 0.8520284 | 0.11069244 | -0.076336801380 | 0.05190402 |

| DV | Predictor | Sum_PostModProb_Incl | Sum_PostModProb_Excl | Sum_PriorModProb_Incl | BF_Incl | Frequentist_p_value | postMean | postStd |
|---|---|---|---|---|---|---|---|---|
| gamma_ROI35_AECnodestrength | Blocktapping | 0.3195033 | 0.6804967 | 0.5 | 0.4695149 | 0.98838665 | 0.001165829682 | 0.02650561 |
| gamma_ROI35_AECnodestrength | Sum_PC | 0.5255372 | 0.4744628 | 0.5 | 1.1076467 | 0.08201736 | 0.090900789489 | 0.05883135 |
| gamma_ROI36_AECnodestrength | Blocktapping | 0.2792126 | 0.7207874 | 0.5 | 0.3873717 | 0.48447928 | -0.019297865264 | 0.03165849 |
| gamma_ROI36_AECnodestrength | Sum_PC | 0.2622407 | 0.7377593 | 0.5 | 0.3554557 | 0.72936667 | -0.010611160063 | 0.02943794 |
| gamma_ROI37_AECnodestrength | Blocktapping | 0.2625722 | 0.7374278 | 0.5 | 0.3560649 | 0.66588134 | -0.013086398484 | 0.02682948 |
| gamma_ROI37_AECnodestrength | Sum_PC | 0.3030174 | 0.6969826 | 0.5 | 0.4347561 | 0.35540005 | -0.030202575945 | 0.03471538 |
| gamma_ROI38_AECnodestrength | Blocktapping | 0.2565945 | 0.7434055 | 0.5 | 0.3451609 | 0.67219592 | -0.011384826464 | 0.02867077 |
| gamma_ROI38_AECnodestrength | Sum_PC | 0.2481275 | 0.7518725 | 0.5 | 0.3300127 | 0.82375243 | -0.005807303320 | 0.02893679 |
| gamma_ROI39_AECnodestrength | Blocktapping | 0.2209822 | 0.7790178 | 0.5 | 0.2836677 | 0.80949620 | 0.003935273493 | 0.02444233 |
| gamma_ROI39_AECnodestrength | Sum_PC | 0.2592999 | 0.7407001 | 0.5 | 0.3500740 | 0.36761824 | -0.025128041063 | 0.03077364 |
| gamma_ROI4_AECnodestrength | Blocktapping | 0.2520049 | 0.7479951 | 0.5 | 0.3369072 | 0.84639181 | -0.005942586886 | 0.02808323 |
| gamma_ROI4_AECnodestrength | Sum_PC | 0.2788443 | 0.7211557 | 0.5 | 0.3866631 | 0.46764246 | -0.021270519400 | 0.03210078 |
| gamma_ROI40_AECnodestrength | Blocktapping | 0.2543904 | 0.7456096 | 0.5 | 0.3411845 | 0.84863677 | 0.003750915243 | 0.02663826 |
| gamma_ROI40_AECnodestrength | Sum_PC | 0.3122822 | 0.6877178 | 0.5 | 0.4540848 | 0.30314727 | -0.034558736585 | 0.03656847 |
| gamma_ROI41_AECnodestrength | Blocktapping | 0.3271126 | 0.6728874 | 0.5 | 0.4861328 | 0.89112925 | 0.002458406443 | 0.02644639 |
| gamma_ROI41_AECnodestrength | Sum_PC | 0.5976403 | 0.4023597 | 0.5 | 1.4853386 | 0.05050111 | -0.114017423801 | 0.06829144 |
| gamma_ROI42_AECnodestrength | Blocktapping | 0.3218097 | 0.6781903 | 0.5 | 0.4745124 | 0.49280481 | -0.016928969185 | 0.03032588 |
| gamma_ROI42_AECnodestrength | Sum_PC | 0.4027616 | 0.5972384 | 0.5 | 0.6743733 | 0.18265953 | 0.052383159638 | 0.04505402 |
| gamma_ROI43_AECnodestrength | Blocktapping | 0.3185077 | 0.6814923 | 0.5 | 0.4673681 | 0.44547674 | -0.020920043162 | 0.03256016 |
| gamma_ROI43_AECnodestrength | Sum_PC | 0.3627048 | 0.6372952 | 0.5 | 0.5691315 | 0.25094009 | 0.040426511374 | 0.03957841 |
| gamma_ROI44_AECnodestrength | Blocktapping | 0.3524255 | 0.6475745 | 0.5 | 0.5442238 | 0.93051566 | 0.004470089777 | 0.02729640 |
| gamma_ROI44_AECnodestrength | Sum_PC | 0.6390124 | 0.3609876 | 0.5 | 1.7701785 | 0.04253361 | 0.125829035800 | 0.06995494 |
| gamma_ROI45_AECnodestrength | Blocktapping | 0.3213424 | 0.6786576 | 0.5 | 0.4734971 | 0.72421844 | -0.007320318673 | 0.02692831 |
| gamma_ROI45_AECnodestrength | Sum_PC | 0.5203410 | 0.4796590 | 0.5 | 1.0848144 | 0.08107766 | 0.091186042996 | 0.05851524 |
| gamma_ROI46_AECnodestrength | Blocktapping | 0.3971015 | 0.6028985 | 0.5 | 0.6586539 | 0.20087451 | -0.050715783830 | 0.04326427 |
| gamma_ROI46_AECnodestrength | Sum_PC | 0.3017013 | 0.6982987 | 0.5 | 0.4320519 | 0.69438398 | -0.013078610127 | 0.03125207 |
| gamma_ROI47_AECnodestrength | Blocktapping | 0.2793454 | 0.7206546 | 0.5 | 0.3876273 | 0.57334844 | -0.013080552645 | 0.02959410 |
| gamma_ROI47_AECnodestrength | Sum_PC | 0.2981714 | 0.7018286 | 0.5 | 0.4248493 | 0.42941013 | 0.023157525429 | 0.03402354 |
| gamma_ROI48_AECnodestrength | Blocktapping | 0.3070724 | 0.6929276 | 0.5 | 0.4431521 | 0.39614018 | -0.027554768791 | 0.03418134 |

| DV | Predictor | Sum_PostModProb_Incl | Sum_PostModProb_Excl | Sum_PriorModProb_Incl | BF_Incl | Frequentist_p_value | postMean | postStd |
|---|---|---|---|---|---|---|---|---|
| gamma_ROI48_AECnodestrength | Sum_PC | 0.2628620 | 0.7371380 | 0.5 | 0.3565981 | 0.86217932 | -0.006281632203 | 0.02844791 |
| gamma_ROI49_AECnodestrength | Blocktapping | 0.2664243 | 0.7335757 | 0.5 | 0.3631859 | 0.54749376 | -0.017035946561 | 0.03054846 |
| gamma_ROI49_AECnodestrength | Sum_PC | 0.2525775 | 0.7474225 | 0.5 | 0.3379313 | 0.70098590 | -0.010689203444 | 0.02893031 |
| gamma_ROI5_AECnodestrength | Blocktapping | 0.5226280 | 0.4773720 | 0.5 | 1.0948026 | 0.08635686 | -0.089187260068 | 0.05649373 |
| gamma_ROI5_AECnodestrength | Sum_PC | 0.3219181 | 0.6780819 | 0.5 | 0.4747480 | 0.96488524 | -0.002575242991 | 0.02854763 |
| gamma_ROI50_AECnodestrength | Blocktapping | 0.2904998 | 0.7095002 | 0.5 | 0.4094428 | 0.51524065 | -0.018340563378 | 0.03088017 |
| gamma_ROI50_AECnodestrength | Sum_PC | 0.3007282 | 0.6992718 | 0.5 | 0.4300591 | 0.44019594 | -0.025046282805 | 0.03336469 |
| gamma_ROI51_AECnodestrength | Blocktapping | 0.2558387 | 0.7441613 | 0.5 | 0.3437948 | 0.69223368 | 0.010167922459 | 0.02806366 |
| gamma_ROI51_AECnodestrength | Sum_PC | 0.2603152 | 0.7396848 | 0.5 | 0.3519272 | 0.64596436 | -0.011679818069 | 0.03007955 |
| gamma_ROI52_AECnodestrength | Blocktapping | 0.2426519 | 0.7573481 | 0.5 | 0.3203969 | 0.84999251 | -0.005813127739 | 0.02757464 |
| gamma_ROI52_AECnodestrength | Sum_PC | 0.2405535 | 0.7594465 | 0.5 | 0.3167484 | 0.87662610 | -0.004462830246 | 0.02799717 |
| gamma_ROI53_AECnodestrength | Blocktapping | 0.2776084 | 0.7223916 | 0.5 | 0.3842907 | 0.52993041 | 0.017887319477 | 0.03162402 |
| gamma_ROI53_AECnodestrength | Sum_PC | 0.2687205 | 0.7312795 | 0.5 | 0.3674661 | 0.61212601 | 0.015120074254 | 0.02975516 |
| gamma_ROI54_AECnodestrength | Blocktapping | 0.3352804 | 0.6647196 | 0.5 | 0.5043937 | 0.27772040 | -0.036577224297 | 0.03691556 |
| gamma_ROI54_AECnodestrength | Sum_PC | 0.2675144 | 0.7324856 | 0.5 | 0.3652146 | 0.80928905 | 0.004237672366 | 0.02915243 |
| gamma_ROI55_AECnodestrength | Blocktapping | 0.5022618 | 0.4977382 | 0.5 | 1.0090882 | 0.09762829 | -0.081479442967 | 0.05217496 |
| gamma_ROI55_AECnodestrength | Sum_PC | 0.3197872 | 0.6802128 | 0.5 | 0.4701281 | 0.77733996 | 0.006115937701 | 0.02834964 |
| gamma_ROI56_AECnodestrength | Blocktapping | 0.2429951 | 0.7570049 | 0.5 | 0.3209954 | 0.77701219 | -0.006799977677 | 0.02790583 |
| gamma_ROI56_AECnodestrength | Sum_PC | 0.2379074 | 0.7620926 | 0.5 | 0.3121765 | 0.94142147 | -0.002527763013 | 0.02781599 |
| gamma_ROI57_AECnodestrength | Blocktapping | 0.2808933 | 0.7191067 | 0.5 | 0.3906141 | 0.38600622 | -0.025060976971 | 0.03250357 |
| gamma_ROI57_AECnodestrength | Sum_PC | 0.2416807 | 0.7583193 | 0.5 | 0.3187058 | 0.78524210 | 0.006283570184 | 0.02796808 |
| gamma_ROI58_AECnodestrength | Blocktapping | 0.4460211 | 0.5539789 | 0.5 | 0.8051228 | 0.13622351 | -0.065478295069 | 0.04904200 |
| gamma_ROI58_AECnodestrength | Sum_PC | 0.2985612 | 0.7014388 | 0.5 | 0.4256412 | 0.77559715 | 0.005753784928 | 0.02891139 |
| gamma_ROI59_AECnodestrength | Blocktapping | 0.2950959 | 0.7049041 | 0.5 | 0.4186326 | 0.63443678 | -0.014117340680 | 0.02868924 |
| gamma_ROI59_AECnodestrength | Sum_PC | 0.3763616 | 0.6236384 | 0.5 | 0.6034934 | 0.22359248 | -0.047832582140 | 0.04365403 |
| gamma_ROI6_AECnodestrength | Blocktapping | 0.3249449 | 0.6750551 | 0.5 | 0.4813606 | 0.30544483 | -0.034167753759 | 0.03708213 |
| gamma_ROI6_AECnodestrength | Sum_PC | 0.2603157 | 0.7396843 | 0.5 | 0.3519282 | 0.93381604 | -0.004445482523 | 0.02816624 |
| gamma_ROI60_AECnodestrength | Blocktapping | 0.3058120 | 0.6941880 | 0.5 | 0.4405321 | 0.45599967 | -0.023317463058 | 0.03270740 |
| gamma_ROI60_AECnodestrength | Sum_PC | 0.3121943 | 0.6878057 | 0.5 | 0.4538990 | 0.40769960 | -0.028328034360 | 0.03581256 |

| DV | Predictor | Sum_PostModProb_Incl | Sum_PostModProb_Excl | Sum_PriorModProb_Incl | BF_Incl | Frequentist_p_value | postMean | postStd |
|---|---|---|---|---|---|---|---|---|
| gamma_ROI61_AECnodestrength | Blocktapping | 0.2523999 | 0.7476001 | 0.5 | 0.3376135 | 0.85446899 | -0.005418233428 | 0.02723966 |
| gamma_ROI61_AECnodestrength | Sum_PC | 0.2874579 | 0.7125421 | 0.5 | 0.4034258 | 0.42080267 | -0.025457732426 | 0.03343932 |
| gamma_ROI62_AECnodestrength | Blocktapping | 0.3125361 | 0.6874639 | 0.5 | 0.4546219 | 0.35972153 | -0.030832649094 | 0.03423179 |
| gamma_ROI62_AECnodestrength | Sum_PC | 0.2689046 | 0.7310954 | 0.5 | 0.3678106 | 0.72069903 | -0.010712847110 | 0.02927374 |
| gamma_ROI63_AECnodestrength | Blocktapping | 0.2877404 | 0.7122596 | 0.5 | 0.4039824 | 0.69297663 | -0.012950283476 | 0.02832419 |
| gamma_ROI63_AECnodestrength | Sum_PC | 0.3705349 | 0.6294651 | 0.5 | 0.5886504 | 0.22916167 | -0.046745226388 | 0.04180862 |
| gamma_ROI64_AECnodestrength | Blocktapping | 0.4023849 | 0.5976151 | 0.5 | 0.6733178 | 0.29252826 | -0.037216667579 | 0.03761143 |
| gamma_ROI64_AECnodestrength | Sum_PC | 0.4902431 | 0.5097569 | 0.5 | 0.9617195 | 0.12766952 | -0.074315684508 | 0.05043365 |
| gamma_ROI65_AECnodestrength | Blocktapping | 0.3777496 | 0.6222504 | 0.5 | 0.6070702 | 0.47745085 | -0.021375472819 | 0.02937653 |
| gamma_ROI65_AECnodestrength | Sum_PC | 0.6347709 | 0.3652291 | 0.5 | 1.7380071 | 0.05138169 | -0.122211785458 | 0.06723265 |
| gamma_ROI66_AECnodestrength | Blocktapping | 0.3204655 | 0.6795345 | 0.5 | 0.4715957 | 0.55105640 | -0.018832457288 | 0.02964678 |
| gamma_ROI66_AECnodestrength | Sum_PC | 0.4481666 | 0.5518334 | 0.5 | 0.8121412 | 0.13266077 | -0.070745768141 | 0.04825348 |
| gamma_ROI67_AECnodestrength | Blocktapping | 0.2316618 | 0.7683382 | 0.5 | 0.3015102 | 0.78999747 | 0.006474765850 | 0.02624713 |
| gamma_ROI67_AECnodestrength | Sum_PC | 0.2275696 | 0.7724304 | 0.5 | 0.2946151 | 0.88068970 | 0.003755436998 | 0.02665222 |
| gamma_ROI68_AECnodestrength | Blocktapping | 0.5415207 | 0.4584793 | 0.5 | 1.1811233 | 0.12498515 | -0.072655442944 | 0.05079873 |
| gamma_ROI68_AECnodestrength | Sum_PC | 0.5186605 | 0.4813395 | 0.5 | 1.0775356 | 0.14630638 | -0.067755361375 | 0.04970953 |
| gamma_ROI69_AECnodestrength | Blocktapping | 0.4236278 | 0.5763722 | 0.5 | 0.7349901 | 0.30024591 | -0.037685557049 | 0.03773266 |
| gamma_ROI69_AECnodestrength | Sum_PC | 0.5196928 | 0.4803072 | 0.5 | 1.0820008 | 0.10697991 | -0.079472426226 | 0.05487028 |
| gamma_ROI7_AECnodestrength | Blocktapping | 0.3221212 | 0.6778788 | 0.5 | 0.4751899 | 0.95764352 | -0.003711113487 | 0.02653358 |
| gamma_ROI7_AECnodestrength | Sum_PC | 0.5581596 | 0.4418404 | 0.5 | 1.2632608 | 0.06502525 | -0.105068407869 | 0.06347838 |
| gamma_ROI70_AECnodestrength | Blocktapping | 0.3128590 | 0.6871410 | 0.5 | 0.4553054 | 0.52754952 | -0.018727670722 | 0.02956645 |
| gamma_ROI70_AECnodestrength | Sum_PC | 0.4044357 | 0.5955643 | 0.5 | 0.6790797 | 0.18059491 | -0.056196056235 | 0.04456661 |
| gamma_ROI71_AECnodestrength | Blocktapping | 0.5013798 | 0.4986202 | 0.5 | 1.0055345 | 0.22878660 | -0.047563187572 | 0.04046643 |
| gamma_ROI71_AECnodestrength | Sum_PC | 0.6737285 | 0.3262715 | 0.5 | 2.0649320 | 0.05041091 | -0.123442115149 | 0.06662887 |
| gamma_ROI72_AECnodestrength | Blocktapping | 0.3455005 | 0.6544995 | 0.5 | 0.5278850 | 0.33367369 | -0.033770203155 | 0.03581646 |
| gamma_ROI72_AECnodestrength | Sum_PC | 0.3288357 | 0.6711643 | 0.5 | 0.4899480 | 0.39169152 | -0.028883971322 | 0.03444943 |
| gamma_ROI73_AECnodestrength | Blocktapping | 0.2769119 | 0.7230881 | 0.5 | 0.3829573 | 0.51655650 | -0.017804867963 | 0.02989043 |
| gamma_ROI73_AECnodestrength | Sum_PC | 0.2720122 | 0.7279878 | 0.5 | 0.3736494 | 0.56636726 | -0.017446208323 | 0.03149684 |
| gamma_ROI74_AECnodestrength | Blocktapping | 0.2455929 | 0.7544071 | 0.5 | 0.3255443 | 0.96782240 | -0.001253601650 | 0.02717168 |

| DV | Predictor | Sum_PostModProb_Incl | Sum_PostModProb_Excl | Sum_PriorModProb_Incl | BF_Incl | Frequentist_p_value | postMean | postStd |
|---|---|---|---|---|---|---|---|---|
| gamma_ROI74_AECnodestrength | Sum_PC | 0.2797980 | 0.7202020 | 0.5 | 0.3884993 | 0.43866682 | -0.022181163826 | 0.03242566 |
| gamma_ROI75_AECnodestrength | Blocktapping | 0.3453795 | 0.6546205 | 0.5 | 0.5276025 | 0.28898473 | -0.036474215689 | 0.03726643 |
| gamma_ROI75_AECnodestrength | Sum_PC | 0.2913972 | 0.7086028 | 0.5 | 0.4112279 | 0.57958932 | -0.017805948258 | 0.03087921 |
| gamma_ROI76_AECnodestrength | Blocktapping | 0.2485575 | 0.7514425 | 0.5 | 0.3307739 | 0.91296329 | 0.002018570721 | 0.02820744 |
| gamma_ROI76_AECnodestrength | Sum_PC | 0.2358595 | 0.7641405 | 0.5 | 0.3086599 | 0.84615363 | -0.004889582225 | 0.02790010 |
| gamma_ROI77_AECnodestrength | Blocktapping | 0.3142684 | 0.6857316 | 0.5 | 0.4582966 | 0.61222065 | -0.016808420730 | 0.02879249 |
| gamma_ROI77_AECnodestrength | Sum_PC | 0.4120782 | 0.5879218 | 0.5 | 0.7009064 | 0.18100062 | -0.056704578778 | 0.04542269 |
| gamma_ROI78_AECnodestrength | Blocktapping | 0.3093712 | 0.6906288 | 0.5 | 0.4479558 | 0.42577683 | -0.025669111973 | 0.03479773 |
| gamma_ROI78_AECnodestrength | Sum_PC | 0.2768789 | 0.7231211 | 0.5 | 0.3828942 | 0.56546782 | -0.016292514332 | 0.02975715 |
| gamma_ROI79_AECnodestrength | Blocktapping | 0.2702540 | 0.7297460 | 0.5 | 0.3703397 | 0.98205876 | 0.000220621034 | 0.02706163 |
| gamma_ROI79_AECnodestrength | Sum_PC | 0.3554075 | 0.6445925 | 0.5 | 0.5513678 | 0.23537526 | 0.043642421405 | 0.04236469 |
| gamma_ROI8_AECnodestrength | Blocktapping | 0.2496636 | 0.7503364 | 0.5 | 0.3327355 | 0.62043295 | -0.013387393178 | 0.02889114 |
| gamma_ROI8_AECnodestrength | Sum_PC | 0.2386215 | 0.7613785 | 0.5 | 0.3134073 | 0.79050138 | -0.008321756443 | 0.02778876 |
| gamma_ROI80_AECnodestrength | Blocktapping | 0.2523642 | 0.7476358 | 0.5 | 0.3375496 | 0.89006506 | 0.002624919195 | 0.02699547 |
| gamma_ROI80_AECnodestrength | Sum_PC | 0.2820066 | 0.7179934 | 0.5 | 0.3927705 | 0.45635887 | -0.023259187090 | 0.03196389 |
| gamma_ROI81_AECnodestrength | Blocktapping | 0.6956505 | 0.3043495 | 0.5 | 2.2856965 | 0.06880778 | -0.102156726607 | 0.06148286 |
| gamma_ROI81_AECnodestrength | Sum_PC | 0.7283645 | 0.2716355 | 0.5 | 2.6814043 | 0.05007036 | -0.122974308113 | 0.06899206 |
| gamma_ROI82_AECnodestrength | Blocktapping | 0.2623198 | 0.7376802 | 0.5 | 0.3556010 | 0.76964763 | -0.009328276819 | 0.02686813 |
| gamma_ROI82_AECnodestrength | Sum_PC | 0.3248420 | 0.6751580 | 0.5 | 0.4811348 | 0.29171361 | -0.036645562140 | 0.03777308 |
| gamma_ROI83_AECnodestrength | Blocktapping | 0.3144231 | 0.6855769 | 0.5 | 0.4586256 | 0.51468445 | 0.021188208438 | 0.03153100 |
| gamma_ROI83_AECnodestrength | Sum_PC | 0.3680669 | 0.6319331 | 0.5 | 0.5824460 | 0.26553228 | 0.042621516460 | 0.04061543 |
| gamma_ROI84_AECnodestrength | Blocktapping | 0.2391890 | 0.7608110 | 0.5 | 0.3143870 | 0.98073113 | 0.000474584856 | 0.02679277 |
| gamma_ROI84_AECnodestrength | Sum_PC | 0.2522280 | 0.7477720 | 0.5 | 0.3373060 | 0.63391120 | 0.012651799777 | 0.03040412 |
| gamma_ROI85_AECnodestrength | Blocktapping | 0.2254209 | 0.7745791 | 0.5 | 0.2910238 | 0.73400903 | -0.007643398200 | 0.02692570 |
| gamma_ROI85_AECnodestrength | Sum_PC | 0.2225472 | 0.7774528 | 0.5 | 0.2862518 | 0.79649035 | 0.007829928668 | 0.02681937 |
| gamma_ROI86_AECnodestrength | Blocktapping | 0.3121823 | 0.6878177 | 0.5 | 0.4538737 | 0.43399015 | -0.023274337929 | 0.03140456 |
| gamma_ROI86_AECnodestrength | Sum_PC | 0.3263548 | 0.6736452 | 0.5 | 0.4844611 | 0.35805347 | -0.031246239050 | 0.03637370 |
| gamma_ROI87_AECnodestrength | Blocktapping | 0.2646850 | 0.7353150 | 0.5 | 0.3599614 | 0.69082812 | -0.011652265860 | 0.02832631 |
| gamma_ROI87_AECnodestrength | Sum_PC | 0.2961466 | 0.7038534 | 0.5 | 0.4207504 | 0.39003792 | -0.027868780168 | 0.03376046 |

| DV | Predictor | Sum_PostModProb_Incl | Sum_PostModProb_Excl | Sum_PriorModProb_Incl | BF_Incl | Frequentist_p_value | postMean | postStd |
|---|---|---|---|---|---|---|---|---|
| gamma_ROI88_AECnodestrength | Blocktapping | 0.2859593 | 0.7140407 | 0.5 | 0.4004804 | 0.45345194 | -0.021796500904 | 0.03316233 |
| gamma_ROI88_AECnodestrength | Sum_PC | 0.2491369 | 0.7508631 | 0.5 | 0.3318007 | 0.94693883 | 0.000786378601 | 0.02807719 |
| gamma_ROI89_AECnodestrength | Blocktapping | 0.3248302 | 0.6751698 | 0.5 | 0.4811089 | 0.40448322 | -0.028011366289 | 0.03414794 |
| gamma_ROI89_AECnodestrength | Sum_PC | 0.3316090 | 0.6683910 | 0.5 | 0.4961302 | 0.35882566 | -0.031806327173 | 0.03723993 |
| gamma_ROI9_AECnodestrength | Blocktapping | 0.3314068 | 0.6685932 | 0.5 | 0.4956778 | 0.28847313 | -0.034629695453 | 0.03661532 |
| gamma_ROI9_AECnodestrength | Sum_PC | 0.2771219 | 0.7228781 | 0.5 | 0.3833591 | 0.96186220 | -0.000810979343 | 0.03072463 |
| gamma_ROI90_AECnodestrength | Blocktapping | 0.3053676 | 0.6946324 | 0.5 | 0.4396105 | 0.42189231 | -0.024912542870 | 0.03254738 |
| gamma_ROI90_AECnodestrength | Sum_PC | 0.3099736 | 0.6900264 | 0.5 | 0.4492199 | 0.37805782 | -0.029385839175 | 0.03426247 |
| gamma_ROI91_AECnodestrength | Blocktapping | 0.2649146 | 0.7350854 | 0.5 | 0.3603861 | 0.71346666 | -0.010149528766 | 0.02733018 |
| gamma_ROI91_AECnodestrength | Sum_PC | 0.3262880 | 0.6737120 | 0.5 | 0.4843138 | 0.28934667 | -0.036732837674 | 0.03789247 |
| gamma_ROI92_AECnodestrength | Blocktapping | 0.2632529 | 0.7367471 | 0.5 | 0.3573179 | 0.98481883 | -0.001098149391 | 0.02867402 |
| gamma_ROI92_AECnodestrength | Sum_PC | 0.2950499 | 0.7049501 | 0.5 | 0.4185400 | 0.40482018 | -0.024803151027 | 0.03482552 |
| gamma_ROI93_AECnodestrength | Blocktapping | 0.2669102 | 0.7330898 | 0.5 | 0.3640894 | 0.66680398 | -0.012503001776 | 0.02809358 |
| gamma_ROI93_AECnodestrength | Sum_PC | 0.3090349 | 0.6909651 | 0.5 | 0.4472512 | 0.34783837 | -0.031617001767 | 0.03510106 |
| gamma_ROI94_AECnodestrength | Blocktapping | 0.2442584 | 0.7557416 | 0.5 | 0.3232036 | 0.94901039 | 0.000222335178 | 0.02732010 |
| gamma_ROI94_AECnodestrength | Sum_PC | 0.2421256 | 0.7578744 | 0.5 | 0.3194798 | 0.94950853 | 0.002956395725 | 0.02935621 |
| gamma_ROI95_AECnodestrength | Blocktapping | 0.2400599 | 0.7599401 | 0.5 | 0.3158932 | 0.86106485 | -0.006362501649 | 0.02721726 |
| gamma_ROI95_AECnodestrength | Sum_PC | 0.2395133 | 0.7604867 | 0.5 | 0.3149475 | 0.93335456 | -0.002695428636 | 0.02787836 |
| gamma_ROI96_AECnodestrength | Blocktapping | 0.3507151 | 0.6492849 | 0.5 | 0.5401559 | 0.82818537 | -0.004882804228 | 0.02744990 |
| gamma_ROI96_AECnodestrength | Sum_PC | 0.6307777 | 0.3692223 | 0.5 | 1.7083954 | 0.04290581 | 0.127817520971 | 0.06658091 |
| gamma_ROI97_AECnodestrength | Blocktapping | 0.2306801 | 0.7693199 | 0.5 | 0.2998493 | 0.88297106 | -0.003666662372 | 0.02593147 |
| gamma_ROI97_AECnodestrength | Sum_PC | 0.2299555 | 0.7700445 | 0.5 | 0.2986263 | 0.95218269 | -0.002493189234 | 0.02749089 |
| gamma_ROI98_AECnodestrength | Blocktapping | 0.3513532 | 0.6486468 | 0.5 | 0.5416711 | 0.35439829 | -0.032963165385 | 0.03638534 |
| gamma_ROI98_AECnodestrength | Sum_PC | 0.3608835 | 0.6391165 | 0.5 | 0.5646600 | 0.30241764 | -0.039431044283 | 0.03828132 |
| gamma_ROI99_AECnodestrength | Blocktapping | 0.2650481 | 0.7349519 | 0.5 | 0.3606332 | 0.54217147 | -0.015807555230 | 0.02952447 |
| gamma_ROI99_AECnodestrength | Sum_PC | 0.2448018 | 0.7551982 | 0.5 | 0.3241557 | 0.82872766 | -0.006705917780 | 0.02781696 |
| GammaPow1 | Blocktapping | 0.2706975 | 0.7293025 | 0.5 | 0.3711732 | 0.45929021 | -0.020445864315 | 0.02859108 |
| GammaPow1 | Sum_PC | 0.2457837 | 0.7542163 | 0.5 | 0.3258796 | 0.91416653 | 0.002123339719 | 0.02715859 |
| GammaPow10 | Blocktapping | 0.3270001 | 0.6729999 | 0.5 | 0.4858843 | 0.33893817 | -0.031912360231 | 0.03406028 |
| GammaPow10 | Sum_PC | 0.3031853 | 0.6968147 | 0.5 | 0.4351018 | 0.43954424 | -0.024211227353 | 0.03128745 |

| DV | Predictor | Sum_PostModProb_Incl | Sum_PostModProb_Excl | Sum_PriorModProb_Incl | BF_Incl | Frequentist_p_value | postMean | postStd |
|---|---|---|---|---|---|---|---|---|
| GammaPow100 | Blocktapping | 0.2780633 | 0.7219367 | 0.5 | 0.3851630 | 0.41084209 | -0.023314598370 | 0.02988305 |
| GammaPow100 | Sum_PC | 0.2387946 | 0.7612054 | 0.5 | 0.3137059 | 0.98438152 | -0.000551821408 | 0.02495038 |
| GammaPow11 | Blocktapping | 0.2947218 | 0.7052782 | 0.5 | 0.4178802 | 0.38134693 | -0.027933562895 | 0.03145941 |
| GammaPow11 | Sum_PC | 0.2620186 | 0.7379814 | 0.5 | 0.3550476 | 0.61200792 | -0.013359083608 | 0.02863464 |
| GammaPow12 | Blocktapping | 0.3457235 | 0.6542765 | 0.5 | 0.5284059 | 0.29073836 | -0.036436642947 | 0.03558142 |
| GammaPow12 | Sum_PC | 0.3216739 | 0.6783261 | 0.5 | 0.4742173 | 0.38421135 | -0.027950888358 | 0.03290728 |
| GammaPow13 | Blocktapping | 0.2980772 | 0.7019228 | 0.5 | 0.4246582 | 0.48985967 | -0.021229748415 | 0.03177045 |
| GammaPow13 | Sum_PC | 0.3269677 | 0.6730323 | 0.5 | 0.4858127 | 0.31953782 | -0.033623066087 | 0.03519219 |
| GammaPow14 | Blocktapping | 0.2491167 | 0.7508833 | 0.5 | 0.3317649 | 0.81477177 | 0.003198715066 | 0.02566658 |
| GammaPow14 | Sum_PC | 0.2913132 | 0.7086868 | 0.5 | 0.4110607 | 0.37180140 | -0.027546712396 | 0.03324204 |
| GammaPow15 | Blocktapping | 0.2662004 | 0.7337996 | 0.5 | 0.3627698 | 0.67696251 | -0.011625053250 | 0.02777779 |
| GammaPow15 | Sum_PC | 0.2823393 | 0.7176607 | 0.5 | 0.3934161 | 0.49322128 | -0.020019372085 | 0.03141245 |
| GammaPow16 | Blocktapping | 0.2407851 | 0.7592149 | 0.5 | 0.3171501 | 0.71489718 | -0.010144066256 | 0.02656514 |
| GammaPow16 | Sum_PC | 0.2336512 | 0.7663488 | 0.5 | 0.3048888 | 0.80060725 | -0.005414444226 | 0.02564699 |
| GammaPow17 | Blocktapping | 0.2491276 | 0.7508724 | 0.5 | 0.3317842 | 0.72685529 | -0.010695398336 | 0.02472975 |
| GammaPow17 | Sum_PC | 0.3261126 | 0.6738874 | 0.5 | 0.4839274 | 0.37757289 | -0.029916782714 | 0.03662776 |
| GammaPow18 | Blocktapping | 0.2303317 | 0.7696683 | 0.5 | 0.2992610 | 0.72231630 | -0.009437007017 | 0.02448117 |
| GammaPow18 | Sum_PC | 0.2852161 | 0.7147839 | 0.5 | 0.3990242 | 0.66331411 | -0.014457171285 | 0.03296030 |
| GammaPow19 | Blocktapping | 0.2446215 | 0.7553785 | 0.5 | 0.3238396 | 0.91524584 | -0.003905950428 | 0.02636802 |
| GammaPow19 | Sum_PC | 0.2760101 | 0.7239899 | 0.5 | 0.3812348 | 0.45050254 | -0.021936083958 | 0.03234885 |
| GammaPow2 | Blocktapping | 0.2448118 | 0.7551882 | 0.5 | 0.3241732 | 0.58018408 | -0.013535004711 | 0.02619117 |
| GammaPow2 | Sum_PC | 0.2325032 | 0.7674968 | 0.5 | 0.3029370 | 0.79457832 | 0.005619406159 | 0.02633265 |
| GammaPow20 | Blocktapping | 0.2569956 | 0.7430044 | 0.5 | 0.3458871 | 0.76012068 | -0.008748498573 | 0.02643516 |
| GammaPow20 | Sum_PC | 0.2816792 | 0.7183208 | 0.5 | 0.3921356 | 0.45761016 | -0.020927981936 | 0.03106632 |
| GammaPow21 | Blocktapping | 0.2612756 | 0.7387244 | 0.5 | 0.3536848 | 0.89436345 | -0.004024690271 | 0.02723554 |
| GammaPow21 | Sum_PC | 0.2933945 | 0.7066055 | 0.5 | 0.4152169 | 0.39454775 | -0.025768667635 | 0.03237342 |
| GammaPow22 | Blocktapping | 0.2888691 | 0.7111309 | 0.5 | 0.4062108 | 0.43983058 | -0.023188278944 | 0.03118964 |
| GammaPow22 | Sum_PC | 0.2797222 | 0.7202778 | 0.5 | 0.3883532 | 0.47817291 | -0.022000521008 | 0.03122904 |
| GammaPow23 | Blocktapping | 0.2422625 | 0.7577375 | 0.5 | 0.3197183 | 0.93624773 | -0.002851841310 | 0.02547811 |
| GammaPow23 | Sum_PC | 0.2682503 | 0.7317497 | 0.5 | 0.3665875 | 0.48031151 | -0.019702218803 | 0.03018019 |
| GammaPow24 | Blocktapping | 0.2520188 | 0.7479812 | 0.5 | 0.3369321 | 0.83385080 | -0.006107093984 | 0.02605678 |
| GammaPow24 | Sum_PC | 0.2968462 | 0.7031538 | 0.5 | 0.4221639 | 0.36511339 | -0.027528621871 | 0.03434406 |
| GammaPow25 | Blocktapping | 0.3620096 | 0.6379904 | 0.5 | 0.5674217 | 0.25650134 | -0.039695828970 | 0.03725199 |
| GammaPow25 | Sum_PC | 0.3150491 | 0.6849509 | 0.5 | 0.4599587 | 0.43935458 | -0.024235547612 | 0.03219686 |
| GammaPow26 | Blocktapping | 0.3796952 | 0.6203048 | 0.5 | 0.6121106 | 0.24814023 | -0.043361539392 | 0.03878796 |
| GammaPow26 | Sum_PC | 0.3221195 | 0.6778805 | 0.5 | 0.4751862 | 0.40726321 | -0.025928796465 | 0.03150806 |
| GammaPow27 | Blocktapping | 0.2640260 | 0.7359740 | 0.5 | 0.3587436 | 0.56125330 | -0.016653810593 | 0.02765514 |
| GammaPow27 | Sum_PC | 0.2884612 | 0.7115388 | 0.5 | 0.4054048 | 0.40289232 | -0.026537220703 | 0.03314065 |
| GammaPow28 | Blocktapping | 0.2565073 | 0.7434927 | 0.5 | 0.3450031 | 0.69298432 | -0.011207881062 | 0.02771154 |
| GammaPow28 | Sum_PC | 0.2768813 | 0.7231187 | 0.5 | 0.3828988 | 0.46191566 | -0.022454337415 | 0.03149940 |
| GammaPow29 | Blocktapping | 0.2569785 | 0.7430215 | 0.5 | 0.3458562 | 0.61766477 | -0.014141788694 | 0.02732735 |

| DV | Predictor | Sum_PostModProb_Incl | Sum_PostModProb_Excl | Sum_PriorModProb_Incl | BF_Incl | Frequentist_p_value | postMean | postStd |
|---|---|---|---|---|---|---|---|---|
| GammaPow29 | Sum_PC | 0.2571654 | 0.7428346 | 0.5 | 0.3461948 | 0.61933951 | -0.014441197789 | 0.02887258 |
| GammaPow3 | Blocktapping | 0.2620773 | 0.7379227 | 0.5 | 0.3551555 | 0.47778100 | -0.018258885842 | 0.02768616 |
| GammaPow3 | Sum_PC | 0.2385083 | 0.7614917 | 0.5 | 0.3132120 | 0.75833975 | 0.007093314974 | 0.02600694 |
| GammaPow30 | Blocktapping | 0.2622414 | 0.7377586 | 0.5 | 0.3554569 | 0.72260702 | -0.009655281804 | 0.02690651 |
| GammaPow30 | Sum_PC | 0.2926691 | 0.7073309 | 0.5 | 0.4137654 | 0.46585600 | -0.020560059038 | 0.03156358 |
| GammaPow31 | Blocktapping | 0.3630620 | 0.6369380 | 0.5 | 0.5700116 | 0.24359276 | -0.045191220698 | 0.03764736 |
| GammaPow31 | Sum_PC | 0.3005871 | 0.6994129 | 0.5 | 0.4297705 | 0.52212001 | -0.019640171704 | 0.03044961 |
| GammaPow32 | Blocktapping | 0.3324822 | 0.6675178 | 0.5 | 0.4980875 | 0.30788764 | -0.033930604692 | 0.03553278 |
| GammaPow32 | Sum_PC | 0.2829603 | 0.7170397 | 0.5 | 0.3946228 | 0.58223755 | -0.016139539726 | 0.02887669 |
| GammaPow33 | Blocktapping | 0.2865451 | 0.7134549 | 0.5 | 0.4016302 | 0.44713495 | -0.021927246418 | 0.03003977 |
| GammaPow33 | Sum_PC | 0.2719325 | 0.7280675 | 0.5 | 0.3734990 | 0.55748047 | -0.015121660670 | 0.02906086 |
| GammaPow34 | Blocktapping | 0.2559215 | 0.7440785 | 0.5 | 0.3439442 | 0.80886031 | -0.007312399435 | 0.02746345 |
| GammaPow34 | Sum_PC | 0.2833534 | 0.7166466 | 0.5 | 0.3953880 | 0.45684631 | -0.021012236884 | 0.03267159 |
| GammaPow35 | Blocktapping | 0.3301803 | 0.6698197 | 0.5 | 0.4929392 | 0.31344067 | -0.033691487012 | 0.03376389 |
| GammaPow35 | Sum_PC | 0.2971454 | 0.7028546 | 0.5 | 0.4227693 | 0.44705774 | -0.023244352347 | 0.03123637 |
| GammaPow36 | Blocktapping | 0.2528788 | 0.7471212 | 0.5 | 0.3384708 | 0.53452000 | -0.016094305467 | 0.02785595 |
| GammaPow36 | Sum_PC | 0.2328845 | 0.7671155 | 0.5 | 0.3035847 | 0.97844767 | -0.001384124852 | 0.02479070 |
| GammaPow37 | Blocktapping | 0.2655393 | 0.7344607 | 0.5 | 0.3615433 | 0.49657779 | -0.018658698319 | 0.02843985 |
| GammaPow37 | Sum_PC | 0.2385744 | 0.7614256 | 0.5 | 0.3133259 | 0.82116298 | -0.006307599852 | 0.02646784 |
| GammaPow38 | Blocktapping | 0.3529693 | 0.6470307 | 0.5 | 0.5455217 | 0.30289291 | -0.036240919672 | 0.03545217 |
| GammaPow38 | Sum_PC | 0.3240700 | 0.6759300 | 0.5 | 0.4794431 | 0.38838394 | -0.026934092826 | 0.03363264 |
| GammaPow39 | Blocktapping | 0.3034214 | 0.6965786 | 0.5 | 0.4355882 | 0.41244460 | -0.026525688052 | 0.03153589 |
| GammaPow39 | Sum_PC | 0.2980917 | 0.7019083 | 0.5 | 0.4246875 | 0.42349742 | -0.023835850546 | 0.03121706 |
| GammaPow4 | Blocktapping | 0.2368908 | 0.7631092 | 0.5 | 0.3104285 | 0.91696468 | -0.003673336992 | 0.02439067 |
| GammaPow4 | Sum_PC | 0.2357722 | 0.7642278 | 0.5 | 0.3085103 | 0.95675074 | -0.001978637586 | 0.02654457 |
| GammaPow40 | Blocktapping | 0.2463002 | 0.7536998 | 0.5 | 0.3267882 | 0.92667689 | -0.003074468618 | 0.02603547 |
| GammaPow40 | Sum_PC | 0.2769955 | 0.7230045 | 0.5 | 0.3831173 | 0.45169669 | -0.021390257403 | 0.03107090 |
| GammaPow41 | Blocktapping | 0.2504974 | 0.7495026 | 0.5 | 0.3342181 | 0.83543892 | 0.004486828093 | 0.02622511 |
| GammaPow41 | Sum_PC | 0.2734692 | 0.7265308 | 0.5 | 0.3764041 | 0.48904986 | -0.018814505014 | 0.03123131 |
| GammaPow42 | Blocktapping | 0.4010595 | 0.5989405 | 0.5 | 0.6696149 | 0.21311802 | -0.048121133400 | 0.03967435 |
| GammaPow42 | Sum_PC | 0.3486193 | 0.6513807 | 0.5 | 0.5352006 | 0.36813725 | -0.029055533844 | 0.03490031 |
| GammaPow43 | Blocktapping | 0.3943481 | 0.6056519 | 0.5 | 0.6511134 | 0.23845412 | -0.044421101369 | 0.03832251 |
| GammaPow43 | Sum_PC | 0.3762368 | 0.6237632 | 0.5 | 0.6031724 | 0.28468800 | -0.039290779413 | 0.03802984 |
| GammaPow44 | Blocktapping | 0.3579142 | 0.6420858 | 0.5 | 0.5574242 | 0.29423264 | -0.036311024649 | 0.03578582 |
| GammaPow44 | Sum_PC | 0.3304226 | 0.6695774 | 0.5 | 0.4934792 | 0.37357898 | -0.031377258852 | 0.03619582 |
| GammaPow45 | Blocktapping | 0.3852270 | 0.6147730 | 0.5 | 0.6266168 | 0.31835237 | -0.033181940685 | 0.03459958 |
| GammaPow45 | Sum_PC | 0.4637243 | 0.5362757 | 0.5 | 0.8647125 | 0.14314480 | -0.063390890409 | 0.04564928 |
| GammaPow46 | Blocktapping | 0.2850428 | 0.7149572 | 0.5 | 0.3986852 | 0.64423888 | -0.014233079106 | 0.02833989 |
| GammaPow46 | Sum_PC | 0.3506526 | 0.6493474 | 0.5 | 0.5400078 | 0.25274955 | -0.040685204602 | 0.03855796 |
| GammaPow47 | Blocktapping | 0.2348129 | 0.7651871 | 0.5 | 0.3068699 | 0.77615304 | -0.007768123787 | 0.02527108 |
| GammaPow47 | Sum_PC | 0.2586545 | 0.7413455 | 0.5 | 0.3488987 | 0.50297577 | -0.019316427486 | 0.02987744 |

| DV | Predictor | Sum_PostModProb_Incl | Sum_PostModProb_Excl | Sum_PriorModProb_Incl | BF_Incl | Frequentist_p_value | postMean | postStd |
|---|---|---|---|---|---|---|---|---|
| GammaPow48 | Blocktapping | 0.2391607 | 0.7608393 | 0.5 | 0.3143380 | 0.88429675 | -0.005396878993 | 0.02609788 |
| GammaPow48 | Sum_PC | 0.2754644 | 0.7245356 | 0.5 | 0.3801944 | 0.47300345 | -0.022096570732 | 0.03227033 |
| GammaPow49 | Blocktapping | 0.2716395 | 0.7283605 | 0.5 | 0.3729464 | 0.44645677 | -0.019764692313 | 0.02840398 |
| GammaPow49 | Sum_PC | 0.2369870 | 0.7630130 | 0.5 | 0.3105937 | 0.83552157 | 0.002798624150 | 0.02554935 |
| GammaPow5 | Blocktapping | 0.2461901 | 0.7538099 | 0.5 | 0.3265944 | 0.53981903 | -0.014945128350 | 0.02619068 |
| GammaPow5 | Sum_PC | 0.2246039 | 0.7753961 | 0.5 | 0.2896634 | 0.90262647 | 0.001705703468 | 0.02432957 |
| GammaPow50 | Blocktapping | 0.2601862 | 0.7398138 | 0.5 | 0.3516915 | 0.49384195 | -0.017787748907 | 0.02787791 |
| GammaPow50 | Sum_PC | 0.2330115 | 0.7669885 | 0.5 | 0.3038005 | 0.92592038 | -0.002473933648 | 0.02513129 |
| GammaPow51 | Blocktapping | 0.2816203 | 0.7183797 | 0.5 | 0.3920215 | 0.43241486 | -0.020821515445 | 0.02961766 |
| GammaPow51 | Sum_PC | 0.2468322 | 0.7531678 | 0.5 | 0.3277255 | 0.83603853 | 0.004800198056 | 0.02680150 |
| GammaPow52 | Blocktapping | 0.2795254 | 0.7204746 | 0.5 | 0.3879740 | 0.39749760 | -0.024413314389 | 0.02896513 |
| GammaPow52 | Sum_PC | 0.2363959 | 0.7636041 | 0.5 | 0.3095792 | 0.87778358 | 0.003024399345 | 0.02454241 |
| GammaPow53 | Blocktapping | 0.2479442 | 0.7520558 | 0.5 | 0.3296885 | 0.56216552 | -0.014522282138 | 0.02625458 |
| GammaPow53 | Sum_PC | 0.2260036 | 0.7739964 | 0.5 | 0.2919957 | 0.92081111 | 0.002505706671 | 0.02405610 |
| GammaPow54 | Blocktapping | 0.2457509 | 0.7542491 | 0.5 | 0.3258219 | 0.54183683 | -0.015094191644 | 0.02593039 |
| GammaPow54 | Sum_PC | 0.2264951 | 0.7735049 | 0.5 | 0.2928166 | 0.85521388 | -0.004253252321 | 0.02409911 |
| GammaPow55 | Blocktapping | 0.3052531 | 0.6947469 | 0.5 | 0.4393732 | 0.30682762 | -0.032168808994 | 0.03261508 |
| GammaPow55 | Sum_PC | 0.2400268 | 0.7599732 | 0.5 | 0.3158359 | 0.93799238 | 0.000468351555 | 0.02388309 |
| GammaPow56 | Blocktapping | 0.3002132 | 0.6997868 | 0.5 | 0.4290067 | 0.32430801 | -0.028345296120 | 0.03064264 |
| GammaPow56 | Sum_PC | 0.2443466 | 0.7556534 | 0.5 | 0.3233580 | 0.84776191 | 0.003048047804 | 0.02495203 |
| GammaPow57 | Blocktapping | 0.2563812 | 0.7436188 | 0.5 | 0.3447751 | 0.48854204 | -0.018174758883 | 0.02667576 |
| GammaPow57 | Sum_PC | 0.2268076 | 0.7731924 | 0.5 | 0.2933392 | 0.94868198 | -0.002767293141 | 0.02287587 |
| GammaPow58 | Blocktapping | 0.2745817 | 0.7254183 | 0.5 | 0.3785151 | 0.39984070 | -0.023446510018 | 0.02923627 |
| GammaPow58 | Sum_PC | 0.2320228 | 0.7679772 | 0.5 | 0.3021220 | 0.91781857 | -0.002829770659 | 0.02441018 |
| GammaPow59 | Blocktapping | 0.2579910 | 0.7420090 | 0.5 | 0.3476926 | 0.60021044 | -0.013996375687 | 0.02722016 |
| GammaPow59 | Sum_PC | 0.2501619 | 0.7498381 | 0.5 | 0.3336212 | 0.73563163 | -0.010617069561 | 0.02787120 |
| GammaPow6 | Blocktapping | 0.2721648 | 0.7278352 | 0.5 | 0.3739375 | 0.44228178 | -0.020618228076 | 0.02886084 |
| GammaPow6 | Sum_PC | 0.2384404 | 0.7615596 | 0.5 | 0.3130949 | 0.80720374 | 0.004439413416 | 0.02491038 |
| GammaPow60 | Blocktapping | 0.2691606 | 0.7308394 | 0.5 | 0.3682897 | 0.47458671 | -0.019731798643 | 0.02808303 |
| GammaPow60 | Sum_PC | 0.2429688 | 0.7570312 | 0.5 | 0.3209496 | 0.89804947 | -0.004018551766 | 0.02668085 |
| GammaPow61 | Blocktapping | 0.2742622 | 0.7257378 | 0.5 | 0.3779081 | 0.58870903 | -0.015832415884 | 0.02906373 |
| GammaPow61 | Sum_PC | 0.2816665 | 0.7183335 | 0.5 | 0.3921111 | 0.47724341 | -0.023084941871 | 0.03181465 |
| GammaPow62 | Blocktapping | 0.2774647 | 0.7225353 | 0.5 | 0.3840154 | 0.66451704 | -0.011883926375 | 0.02728140 |
| GammaPow62 | Sum_PC | 0.3206414 | 0.6793586 | 0.5 | 0.4719766 | 0.34358989 | -0.031147767377 | 0.03612607 |
| GammaPow63 | Blocktapping | 0.2644371 | 0.7355629 | 0.5 | 0.3595030 | 0.76053188 | -0.009017092585 | 0.02673833 |
| GammaPow63 | Sum_PC | 0.3091510 | 0.6908490 | 0.5 | 0.4474944 | 0.34762554 | -0.029453845777 | 0.03399219 |
| GammaPow64 | Blocktapping | 0.2645996 | 0.7354004 | 0.5 | 0.3598034 | 0.65415018 | -0.013218988140 | 0.02716482 |
| GammaPow64 | Sum_PC | 0.2872054 | 0.7127946 | 0.5 | 0.4029287 | 0.42997240 | -0.023776842490 | 0.03104828 |
| GammaPow65 | Blocktapping | 0.2958777 | 0.7041223 | 0.5 | 0.4202079 | 0.44179608 | -0.022857146130 | 0.03004135 |
| GammaPow65 | Sum_PC | 0.2746190 | 0.7253810 | 0.5 | 0.3785859 | 0.60252600 | -0.013581390377 | 0.02851991 |
| GammaPow66 | Blocktapping | 0.2741689 | 0.7258311 | 0.5 | 0.3777309 | 0.59032804 | -0.015629701335 | 0.02896365 |

| DV | Predictor | Sum_PostModProb_Incl | Sum_PostModProb_Excl | Sum_PriorModProb_Incl | BF_Incl | Frequentist_p_value | postMean | postStd |
|---|---|---|---|---|---|---|---|---|
| GammaPow66 | Sum_PC | 0.2788210 | 0.7211790 | 0.5 | 0.3866184 | 0.50521177 | -0.019181023332 | 0.02995672 |
| GammaPow67 | Blocktapping | 0.2640957 | 0.7359043 | 0.5 | 0.3588723 | 0.50229768 | -0.017532156664 | 0.02708109 |
| GammaPow67 | Sum_PC | 0.2367224 | 0.7632776 | 0.5 | 0.3101394 | 0.91797994 | -0.002509869456 | 0.02582819 |
| GammaPow68 | Blocktapping | 0.2631437 | 0.7368563 | 0.5 | 0.3571167 | 0.62424102 | -0.012468799729 | 0.02656143 |
| GammaPow68 | Sum_PC | 0.2782683 | 0.7217317 | 0.5 | 0.3855565 | 0.49101861 | -0.018940308313 | 0.03102434 |
| GammaPow69 | Blocktapping | 0.3068263 | 0.6931737 | 0.5 | 0.4426399 | 0.38039628 | -0.026224582484 | 0.03131123 |
| GammaPow69 | Sum_PC | 0.2700333 | 0.7299667 | 0.5 | 0.3699256 | 0.62142334 | -0.014286854234 | 0.02777555 |
| GammaPow7 | Blocktapping | 0.2371999 | 0.7628001 | 0.5 | 0.3109595 | 0.91016996 | -0.003195280364 | 0.02574486 |
| GammaPow7 | Sum_PC | 0.2425961 | 0.7574039 | 0.5 | 0.3202995 | 0.70682186 | -0.009199654076 | 0.02756326 |
| GammaPow70 | Blocktapping | 0.2907400 | 0.7092600 | 0.5 | 0.4099203 | 0.45485872 | -0.020948134792 | 0.02914604 |
| GammaPow70 | Sum_PC | 0.2610584 | 0.7389416 | 0.5 | 0.3532869 | 0.72231766 | -0.009481904337 | 0.02640981 |
| GammaPow71 | Blocktapping | 0.2785591 | 0.7214409 | 0.5 | 0.3861150 | 0.66945058 | -0.011884389149 | 0.02743493 |
| GammaPow71 | Sum_PC | 0.2998907 | 0.7001093 | 0.5 | 0.4283485 | 0.41359802 | -0.023786451193 | 0.03171788 |
| GammaPow72 | Blocktapping | 0.2757508 | 0.7242492 | 0.5 | 0.3807402 | 0.57619023 | -0.016043750315 | 0.02878786 |
| GammaPow72 | Sum_PC | 0.2878251 | 0.7121749 | 0.5 | 0.4041494 | 0.45777849 | -0.023751856149 | 0.03266804 |
| GammaPow73 | Blocktapping | 0.2584572 | 0.7415428 | 0.5 | 0.3485398 | 0.62292280 | -0.012438431575 | 0.02825679 |
| GammaPow73 | Sum_PC | 0.2659888 | 0.7340112 | 0.5 | 0.3623771 | 0.52736950 | -0.018461992919 | 0.02924587 |
| GammaPow74 | Blocktapping | 0.2431037 | 0.7568963 | 0.5 | 0.3211850 | 0.66412196 | -0.011382990172 | 0.02585467 |
| GammaPow74 | Sum_PC | 0.2326635 | 0.7673365 | 0.5 | 0.3032093 | 0.87173822 | -0.003606558113 | 0.02565120 |
| GammaPow75 | Blocktapping | 0.2489893 | 0.7510107 | 0.5 | 0.3315390 | 0.66491038 | -0.011407445983 | 0.02660065 |
| GammaPow75 | Sum_PC | 0.2524754 | 0.7475246 | 0.5 | 0.3377486 | 0.62899792 | -0.013517006404 | 0.02804278 |
| GammaPow76 | Blocktapping | 0.2887106 | 0.7112894 | 0.5 | 0.4058976 | 0.44459906 | -0.023384629813 | 0.03046471 |
| GammaPow76 | Sum_PC | 0.2576339 | 0.7423661 | 0.5 | 0.3470442 | 0.75887272 | -0.008819984926 | 0.02901804 |
| GammaPow77 | Blocktapping | 0.2813959 | 0.7186041 | 0.5 | 0.3915869 | 0.47454350 | -0.021581351734 | 0.03101937 |
| GammaPow77 | Sum_PC | 0.2489735 | 0.7510265 | 0.5 | 0.3315110 | 0.75552371 | -0.007787375434 | 0.02631445 |
| GammaPow78 | Blocktapping | 0.2571768 | 0.7428232 | 0.5 | 0.3462154 | 0.68036587 | -0.011482979224 | 0.02760081 |
| GammaPow78 | Sum_PC | 0.2718555 | 0.7281445 | 0.5 | 0.3733538 | 0.51079599 | -0.018952968479 | 0.03014131 |
| GammaPow79 | Blocktapping | 0.3367720 | 0.6632280 | 0.5 | 0.5077771 | 0.29320963 | -0.036000205576 | 0.03669155 |
| GammaPow79 | Sum_PC | 0.2875324 | 0.7124676 | 0.5 | 0.4035726 | 0.61067057 | -0.015163752861 | 0.03063931 |
| GammaPow8 | Blocktapping | 0.2347902 | 0.7652098 | 0.5 | 0.3068311 | 0.92254106 | -0.002854271413 | 0.02587337 |
| GammaPow8 | Sum_PC | 0.2332932 | 0.7667068 | 0.5 | 0.3042795 | 0.98677512 | 0.000002728649 | 0.02543927 |
| GammaPow80 | Blocktapping | 0.2923630 | 0.7076370 | 0.5 | 0.4131540 | 0.41584695 | -0.024706400125 | 0.03205110 |
| GammaPow80 | Sum_PC | 0.2557245 | 0.7442755 | 0.5 | 0.3435884 | 0.84650802 | -0.005354730395 | 0.02702853 |
| GammaPow81 | Blocktapping | 0.2791955 | 0.7208045 | 0.5 | 0.3873387 | 0.48415378 | -0.019526281775 | 0.02864533 |
| GammaPow81 | Sum_PC | 0.2629090 | 0.7370910 | 0.5 | 0.3566846 | 0.63800040 | -0.012513834842 | 0.02742660 |
| GammaPow82 | Blocktapping | 0.3267864 | 0.6732136 | 0.5 | 0.4854127 | 0.33987082 | -0.030009372285 | 0.03220105 |
| GammaPow82 | Sum_PC | 0.2785046 | 0.7214954 | 0.5 | 0.3860101 | 0.66625335 | -0.011771627692 | 0.02716091 |
| GammaPow83 | Blocktapping | 0.3766646 | 0.6233354 | 0.5 | 0.6042729 | 0.29341443 | -0.038468995960 | 0.03592510 |
| GammaPow83 | Sum_PC | 0.4003744 | 0.5996256 | 0.5 | 0.6677074 | 0.22125628 | -0.046715636200 | 0.04049065 |
| GammaPow84 | Blocktapping | 0.3513022 | 0.6486978 | 0.5 | 0.5415498 | 0.37780051 | -0.030149295513 | 0.03434720 |
| GammaPow84 | Sum_PC | 0.3943110 | 0.6056890 | 0.5 | 0.6510123 | 0.23880709 | -0.046066940350 | 0.04093649 |

| DV | Predictor | Sum_PostModProb_Incl | Sum_PostModProb_Excl | Sum_PriorModProb_Incl | BF_Incl | Frequentist_p_value | postMean | postStd |
|---|---|---|---|---|---|---|---|---|
| GammaPow85 | Blocktapping | 0.3979454 | 0.6020546 | 0.5 | 0.6609791 | 0.37533376 | -0.028535962537 | 0.03301530 |
| GammaPow85 | Sum_PC | 0.5580179 | 0.4419821 | 0.5 | 1.2625352 | 0.08988470 | -0.090853616483 | 0.05957113 |
| GammaPow86 | Blocktapping | 0.2732651 | 0.7267349 | 0.5 | 0.3760176 | 0.72422414 | -0.009672973007 | 0.02764443 |
| GammaPow86 | Sum_PC | 0.3298117 | 0.6701883 | 0.5 | 0.4921180 | 0.29275089 | -0.036361081414 | 0.03704885 |
| GammaPow87 | Blocktapping | 0.2718194 | 0.7281806 | 0.5 | 0.3732856 | 0.45797996 | -0.020155273624 | 0.02935973 |
| GammaPow87 | Sum_PC | 0.2597979 | 0.7402021 | 0.5 | 0.3509824 | 0.89569715 | -0.004072016772 | 0.02827265 |
| GammaPow88 | Blocktapping | 0.2833324 | 0.7166676 | 0.5 | 0.3953471 | 0.54682656 | -0.017472673020 | 0.02970723 |
| GammaPow88 | Sum_PC | 0.2998004 | 0.7001996 | 0.5 | 0.4281642 | 0.40437089 | -0.027337653567 | 0.03377338 |
| GammaPow89 | Blocktapping | 0.2682792 | 0.7317208 | 0.5 | 0.3666414 | 0.49620717 | -0.018003409453 | 0.02865865 |
| GammaPow89 | Sum_PC | 0.2402458 | 0.7597542 | 0.5 | 0.3162151 | 0.86355030 | -0.006088466020 | 0.02578881 |
| GammaPow9 | Blocktapping | 0.2596544 | 0.7403456 | 0.5 | 0.3507206 | 0.50516327 | -0.017585079226 | 0.02822277 |
| GammaPow9 | Sum_PC | 0.2312368 | 0.7687632 | 0.5 | 0.3007906 | 0.94219049 | -0.000327863778 | 0.02426368 |
| GammaPow90 | Blocktapping | 0.2943592 | 0.7056408 | 0.5 | 0.4171517 | 0.40388534 | -0.023999664513 | 0.02932667 |
| GammaPow90 | Sum_PC | 0.2536592 | 0.7463408 | 0.5 | 0.3398704 | 0.86153673 | -0.005670203978 | 0.02517034 |
| GammaPow91 | Blocktapping | 0.2409650 | 0.7590350 | 0.5 | 0.3174624 | 0.73048871 | -0.010219790738 | 0.02613356 |
| GammaPow91 | Sum_PC | 0.2379036 | 0.7620964 | 0.5 | 0.3121700 | 0.79679356 | -0.007197014287 | 0.02694237 |
| GammaPow92 | Blocktapping | 0.2805454 | 0.7194546 | 0.5 | 0.3899417 | 0.50366274 | -0.020022788948 | 0.03025259 |
| GammaPow92 | Sum_PC | 0.2635021 | 0.7364979 | 0.5 | 0.3577772 | 0.65588002 | -0.012798306781 | 0.02848861 |
| GammaPow93 | Blocktapping | 0.2403122 | 0.7596878 | 0.5 | 0.3163301 | 0.69499453 | -0.009928641076 | 0.02594672 |
| GammaPow93 | Sum_PC | 0.2326510 | 0.7673490 | 0.5 | 0.3031880 | 0.83416977 | -0.005501982290 | 0.02669414 |
| GammaPow94 | Blocktapping | 0.3504735 | 0.6495265 | 0.5 | 0.5395832 | 0.27436629 | -0.038271649779 | 0.03761926 |
| GammaPow94 | Sum_PC | 0.2818531 | 0.7181469 | 0.5 | 0.3924727 | 0.66797396 | -0.013387216981 | 0.02985527 |
| GammaPow95 | Blocktapping | 0.3127275 | 0.6872725 | 0.5 | 0.4550270 | 0.39203335 | -0.026833937848 | 0.03361255 |
| GammaPow95 | Sum_PC | 0.2989150 | 0.7010850 | 0.5 | 0.4263605 | 0.49960050 | -0.021298800455 | 0.03234578 |
| GammaPow96 | Blocktapping | 0.3480173 | 0.6519827 | 0.5 | 0.5337830 | 0.32266126 | -0.033610120356 | 0.03572528 |
| GammaPow96 | Sum_PC | 0.3419803 | 0.6580197 | 0.5 | 0.5197114 | 0.35102873 | -0.031916932352 | 0.03632381 |
| GammaPow97 | Blocktapping | 0.3790110 | 0.6209890 | 0.5 | 0.6103344 | 0.42040207 | -0.026559219885 | 0.03245952 |
| GammaPow97 | Sum_PC | 0.5256491 | 0.4743509 | 0.5 | 1.1081438 | 0.09744880 | -0.084051000099 | 0.05515230 |
| GammaPow98 | Blocktapping | 0.2765700 | 0.7234300 | 0.5 | 0.3823037 | 0.71328913 | -0.011416090765 | 0.02737835 |
| GammaPow98 | Sum_PC | 0.3483505 | 0.6516495 | 0.5 | 0.5345672 | 0.24833568 | -0.041607819007 | 0.03864771 |
| GammaPow99 | Blocktapping | 0.2997536 | 0.7002464 | 0.5 | 0.4280688 | 0.32948064 | -0.028063389035 | 0.03068150 |
| GammaPow99 | Sum_PC | 0.2442937 | 0.7557063 | 0.5 | 0.3232655 | 0.86193390 | 0.002355741757 | 0.02437477 |
| global_absPower_alpha | Blocktapping | 0.4063531 | 0.5936469 | 0.5 | 0.6845031 | 0.16537010 | 0.055749896607 | 0.04207510 |
| global_absPower_alpha | Sum_PC | 0.2939308 | 0.7060692 | 0.5 | 0.4162918 | 0.68177129 | -0.009303838587 | 0.02864948 |
| global_absPower_beta | Blocktapping | 0.3125265 | 0.6874735 | 0.5 | 0.4546015 | 0.29673315 | 0.029519807283 | 0.03271381 |
| global_absPower_beta | Sum_PC | 0.2651262 | 0.7348738 | 0.5 | 0.3607780 | 0.66432199 | -0.009402428258 | 0.02686345 |
| global_absPower_gamma | Blocktapping | 0.3485910 | 0.6514090 | 0.5 | 0.5351338 | 0.67356035 | -0.011566607352 | 0.02455640 |
| global_absPower_gamma | Sum_PC | 0.6670966 | 0.3329034 | 0.5 | 2.0038740 | 0.03570576 | -0.128141529323 | 0.06769666 |

| DV | Predictor | Sum_PostModProb_Incl | Sum_PostModProb_Excl | Sum_PriorModProb_Incl | BF_Incl | Frequentist_p_value | postMean | postStd |
|---|---|---|---|---|---|---|---|---|
| global_absPower_theta | Blocktapping | 0.2421800 | 0.7578200 | 0.5 | 0.3195746 | 0.64232703 | 0.009522933429 | 0.02635213 |
| global_absPower_theta | Sum_PC | 0.2538762 | 0.7461238 | 0.5 | 0.3402601 | 0.49524453 | -0.017947216517 | 0.03008180 |
| global_PAF_Cog | Blocktapping | 0.1873100 | 0.8126900 | 0.5 | 0.2304814 | 0.74026766 | 0.006606869742 | 0.02224685 |
| global_PAF_Cog | Sum_PC | 0.1852047 | 0.8147953 | 0.5 | 0.2273022 | 0.88664916 | -0.001461900828 | 0.02217618 |
| PAF1 | Blocktapping | 0.2227519 | 0.7772481 | 0.5 | 0.2865905 | 0.54254941 | -0.013964774931 | 0.02442135 |
| PAF1 | Sum_PC | 0.2376416 | 0.7623584 | 0.5 | 0.3117190 | 0.41677892 | 0.019842462781 | 0.02801963 |
| PAF10 | Blocktapping | 0.2531841 | 0.7468159 | 0.5 | 0.3390181 | 0.98828723 | 0.001015143694 | 0.02488111 |
| PAF10 | Sum_PC | 0.3718427 | 0.6281573 | 0.5 | 0.5919580 | 0.16887411 | 0.055170894519 | 0.04388152 |
| PAF100 | Blocktapping | 0.2427413 | 0.7572587 | 0.5 | 0.3205527 | 0.42476100 | 0.021345624838 | 0.02755184 |
| PAF100 | Sum_PC | 0.2198533 | 0.7801467 | 0.5 | 0.2818102 | 0.63059424 | 0.012670238237 | 0.02491592 |
| PAF11 | Blocktapping | 0.2428764 | 0.7571236 | 0.5 | 0.3207883 | 0.92972039 | 0.002462980113 | 0.02559605 |
| PAF11 | Sum_PC | 0.3194046 | 0.6805954 | 0.5 | 0.4693016 | 0.25248735 | 0.041032503893 | 0.03732064 |
| PAF12 | Blocktapping | 0.2685812 | 0.7314188 | 0.5 | 0.3672058 | 0.72463732 | -0.006546479740 | 0.02506827 |
| PAF12 | Sum_PC | 0.3946501 | 0.6053499 | 0.5 | 0.6519373 | 0.13727364 | 0.062376596330 | 0.04558062 |
| PAF13 | Blocktapping | 0.2539565 | 0.7460435 | 0.5 | 0.3404044 | 0.89790485 | -0.002457247605 | 0.02698823 |
| PAF13 | Sum_PC | 0.3208431 | 0.6791569 | 0.5 | 0.4724138 | 0.28322934 | 0.036886137722 | 0.03769988 |
| PAF14 | Blocktapping | 0.2562641 | 0.7437359 | 0.5 | 0.3445633 | 0.64884188 | 0.012497963037 | 0.02748950 |
| PAF14 | Sum_PC | 0.2647243 | 0.7352757 | 0.5 | 0.3600341 | 0.53769070 | 0.019199778909 | 0.03138877 |
| PAF15 | Blocktapping | 0.2535558 | 0.7464442 | 0.5 | 0.3396849 | 0.46050427 | 0.019533086366 | 0.02859509 |
| PAF15 | Sum_PC | 0.2271478 | 0.7728522 | 0.5 | 0.2939084 | 0.97568970 | 0.001276512431 | 0.02590461 |
| PAF16 | Blocktapping | 0.2311185 | 0.7688815 | 0.5 | 0.3005905 | 0.47051152 | -0.015897009155 | 0.02528311 |
| PAF16 | Sum_PC | 0.2329191 | 0.7670809 | 0.5 | 0.3036435 | 0.45204374 | 0.017944217615 | 0.02710353 |
| PAF17 | Blocktapping | 0.2261451 | 0.7738549 | 0.5 | 0.2922318 | 0.95970784 | 0.001194422337 | 0.02548487 |
| PAF17 | Sum_PC | 0.2325611 | 0.7674389 | 0.5 | 0.3030354 | 0.72069552 | 0.009485815979 | 0.02772914 |
| PAF18 | Blocktapping | 0.2315727 | 0.7684273 | 0.5 | 0.3013593 | 0.52813245 | 0.016268665749 | 0.02698143 |
| PAF18 | Sum_PC | 0.2116663 | 0.7883337 | 0.5 | 0.2684984 | 0.83900726 | 0.006440241433 | 0.02396702 |
| PAF19 | Blocktapping | 0.2681096 | 0.7318904 | 0.5 | 0.3663247 | 0.65360616 | 0.012304866840 | 0.02709787 |
| PAF19 | Sum_PC | 0.2813942 | 0.7186058 | 0.5 | 0.3915835 | 0.51092638 | 0.019275411954 | 0.03066803 |
| PAF2 | Blocktapping | 0.2000599 | 0.7999401 | 0.5 | 0.2500936 | 0.67613103 | -0.009811747646 | 0.02307509 |
| PAF2 | Sum_PC | 0.1997606 | 0.8002394 | 0.5 | 0.2496260 | 0.65830011 | 0.009442118198 | 0.02360427 |
| PAF20 | Blocktapping | 0.2581342 | 0.7418658 | 0.5 | 0.3479527 | 0.37291289 | 0.023494034626 | 0.02868617 |
| PAF20 | Sum_PC | 0.2175104 | 0.7824896 | 0.5 | 0.2779723 | 0.89528686 | 0.003770690480 | 0.02445258 |
| PAF21 | Blocktapping | 0.2305005 | 0.7694995 | 0.5 | 0.2995460 | 0.66272143 | 0.009551116974 | 0.02660779 |
| PAF21 | Sum_PC | 0.2203355 | 0.7796645 | 0.5 | 0.2826030 | 0.90090128 | -0.002790651004 | 0.02644238 |
| PAF22 | Blocktapping | 0.2600254 | 0.7399746 | 0.5 | 0.3513978 | 0.85595512 | -0.002931144854 | 0.02708861 |
| PAF22 | Sum_PC | 0.3083342 | 0.6916658 | 0.5 | 0.4457850 | 0.31658826 | 0.031295093554 | 0.03673119 |
| PAF23 | Blocktapping | 0.2466530 | 0.7533470 | 0.5 | 0.3274095 | 0.74985908 | 0.007349971695 | 0.02838398 |
| PAF23 | Sum_PC | 0.2440318 | 0.7559682 | 0.5 | 0.3228070 | 0.92291364 | 0.003306470084 | 0.02849376 |
| PAF24 | Blocktapping | 0.2187299 | 0.7812701 | 0.5 | 0.2799671 | 0.86417241 | 0.003510867523 | 0.02563097 |
| PAF24 | Sum_PC | 0.2193629 | 0.7806371 | 0.5 | 0.2810049 | 0.82655734 | 0.005055832641 | 0.02636588 |

| DV | Predictor | Sum_PostModProb_Incl | Sum_PostModProb_Excl | Sum_PriorModProb_Incl | BF_Incl | Frequentist_p_value | postMean | postStd |
|---|---|---|---|---|---|---|---|---|
| PAF25 | Blocktapping | 0.2870850 | 0.7129150 | 0.5 | 0.4026918 | 0.54726613 | -0.014191225659 | 0.02659776 |
| PAF25 | Sum_PC | 0.4132904 | 0.5867096 | 0.5 | 0.7044208 | 0.13367313 | 0.062459109389 | 0.04653857 |
| PAF26 | Blocktapping | 0.3027607 | 0.6972393 | 0.5 | 0.4342279 | 0.54524626 | -0.014385605857 | 0.02858924 |
| PAF26 | Sum_PC | 0.4220817 | 0.5779183 | 0.5 | 0.7303483 | 0.13172102 | 0.064136475200 | 0.04777696 |
| PAF27 | Blocktapping | 0.2638970 | 0.7361030 | 0.5 | 0.3585055 | 0.57756109 | -0.012591747082 | 0.02883411 |
| PAF27 | Sum_PC | 0.2755160 | 0.7244840 | 0.5 | 0.3802927 | 0.47842582 | 0.019342969692 | 0.03217033 |
| PAF28 | Blocktapping | 0.2614748 | 0.7385252 | 0.5 | 0.3540500 | 0.68680310 | 0.011285353684 | 0.02908896 |
| PAF28 | Sum_PC | 0.2705848 | 0.7294152 | 0.5 | 0.3709613 | 0.61170331 | 0.014037370561 | 0.03130121 |
| PAF29 | Blocktapping | 0.2636685 | 0.7363315 | 0.5 | 0.3580840 | 0.43656075 | 0.020663950731 | 0.02995077 |
| PAF29 | Sum_PC | 0.2395108 | 0.7604892 | 0.5 | 0.3149431 | 0.68950921 | 0.011203477882 | 0.02705995 |
| PAF3 | Blocktapping | 0.2067515 | 0.7932485 | 0.5 | 0.2606390 | 0.64654941 | -0.009395517035 | 0.02277581 |
| PAF3 | Sum_PC | 0.2197264 | 0.7802736 | 0.5 | 0.2816018 | 0.48157945 | 0.016651430949 | 0.02565728 |
| PAF30 | Blocktapping | 0.2846103 | 0.7153897 | 0.5 | 0.3978395 | 0.46393187 | 0.021574833935 | 0.03131595 |
| PAF30 | Sum_PC | 0.2544963 | 0.7455037 | 0.5 | 0.3413749 | 0.93424663 | 0.003621685717 | 0.02862881 |
| PAF31 | Blocktapping | 0.2977431 | 0.7022569 | 0.5 | 0.4239804 | 0.43348252 | -0.021092020904 | 0.02898493 |
| PAF31 | Sum_PC | 0.3667774 | 0.6332226 | 0.5 | 0.5792236 | 0.18193905 | 0.050589971622 | 0.04108524 |
| PAF32 | Blocktapping | 0.2811529 | 0.7188471 | 0.5 | 0.3911163 | 0.34742515 | -0.023964773730 | 0.02994811 |
| PAF32 | Sum_PC | 0.2857382 | 0.7142618 | 0.5 | 0.4000469 | 0.32869881 | 0.027790723533 | 0.03155578 |
| PAF33 | Blocktapping | 0.2465352 | 0.7534648 | 0.5 | 0.3272021 | 0.46334724 | -0.016316453026 | 0.02601118 |
| PAF33 | Sum_PC | 0.2817538 | 0.7182462 | 0.5 | 0.3922802 | 0.27203500 | 0.032787491768 | 0.03234242 |
| PAF34 | Blocktapping | 0.2772176 | 0.7227824 | 0.5 | 0.3835422 | 0.51658459 | 0.017931369480 | 0.02940359 |
| PAF34 | Sum_PC | 0.3005634 | 0.6994366 | 0.5 | 0.4297222 | 0.37228494 | 0.028005211239 | 0.03339596 |
| PAF35 | Blocktapping | 0.2971352 | 0.7028648 | 0.5 | 0.4227488 | 0.46942122 | -0.017335468663 | 0.02992134 |
| PAF35 | Sum_PC | 0.3460576 | 0.6539424 | 0.5 | 0.5291867 | 0.24945489 | 0.039789409231 | 0.03898201 |
| PAF36 | Blocktapping | 0.1979923 | 0.8020077 | 0.5 | 0.2468708 | 0.98939478 | -0.000377531319 | 0.02239565 |
| PAF36 | Sum_PC | 0.1991614 | 0.8008386 | 0.5 | 0.2486910 | 0.81495065 | 0.005476114414 | 0.02270087 |
| PAF37 | Blocktapping | 0.2641994 | 0.7358006 | 0.5 | 0.3590638 | 0.51804737 | 0.016995433175 | 0.02856584 |
| PAF37 | Sum_PC | 0.2642700 | 0.7357300 | 0.5 | 0.3591942 | 0.47355298 | 0.021018991927 | 0.02986043 |
| PAF38 | Blocktapping | 0.2458370 | 0.7541630 | 0.5 | 0.3259733 | 0.63600877 | -0.010203629299 | 0.02528391 |
| PAF38 | Sum_PC | 0.3106459 | 0.6893541 | 0.5 | 0.4506333 | 0.25748936 | 0.036368942574 | 0.03601795 |
| PAF39 | Blocktapping | 0.2198012 | 0.7801988 | 0.5 | 0.2817245 | 0.82881842 | -0.004206217833 | 0.02257516 |
| PAF39 | Sum_PC | 0.2905483 | 0.7094517 | 0.5 | 0.4095393 | 0.25872404 | 0.038176350360 | 0.03438306 |
| PAF4 | Blocktapping | 0.2038662 | 0.7961338 | 0.5 | 0.2560702 | 0.62046823 | -0.011049465615 | 0.02294353 |
| PAF4 | Sum_PC | 0.1933497 | 0.8066503 | 0.5 | 0.2396946 | 0.78220939 | 0.005313069299 | 0.02309809 |
| PAF40 | Blocktapping | 0.2073527 | 0.7926473 | 0.5 | 0.2615951 | 0.73940677 | -0.007234694044 | 0.02307899 |
| PAF40 | Sum_PC | 0.2278233 | 0.7721767 | 0.5 | 0.2950404 | 0.48345342 | 0.017212093236 | 0.02781715 |
| PAF41 | Blocktapping | 0.2008143 | 0.7991857 | 0.5 | 0.2512737 | 0.93163353 | -0.001938765841 | 0.02260954 |
| PAF41 | Sum_PC | 0.2215404 | 0.7784596 | 0.5 | 0.2845882 | 0.50595325 | 0.016139424946 | 0.02698868 |
| PAF42 | Blocktapping | 0.3309780 | 0.6690220 | 0.5 | 0.4947191 | 0.35197214 | -0.025284932963 | 0.03221280 |
| PAF42 | Sum_PC | 0.3958345 | 0.6041655 | 0.5 | 0.6551755 | 0.16725763 | 0.054250685048 | 0.04391860 |
| PAF43 | Blocktapping | 0.2880386 | 0.7119614 | 0.5 | 0.4045705 | 0.38858602 | -0.024084579561 | 0.02990687 |

| DV | Predictor | Sum_PostModProb_Incl | Sum_PostModProb_Excl | Sum_PriorModProb_Incl | BF_Incl | Frequentist_p_value | postMean | postStd |
|---|---|---|---|---|---|---|---|---|
| PAF43 | Sum_PC | 0.3067863 | 0.6932137 | 0.5 | 0.4425566 | 0.30484068 | 0.031751104043 | 0.03493496 |
| PAF44 | Blocktapping | 0.3040947 | 0.6959053 | 0.5 | 0.4369772 | 0.49330585 | -0.016429306944 | 0.02951329 |
| PAF44 | Sum_PC | 0.3774038 | 0.6225962 | 0.5 | 0.6061776 | 0.20427225 | 0.048424246152 | 0.04294383 |
| PAF45 | Blocktapping | 0.2480067 | 0.7519933 | 0.5 | 0.3297991 | 0.70292798 | -0.010255223976 | 0.02764223 |
| PAF45 | Sum_PC | 0.2483922 | 0.7516078 | 0.5 | 0.3304812 | 0.69762787 | 0.008939978762 | 0.02865861 |
| PAF46 | Blocktapping | 0.2458374 | 0.7541626 | 0.5 | 0.3259740 | 0.89534423 | 0.004801672278 | 0.02621120 |
| PAF46 | Sum_PC | 0.2654003 | 0.7345997 | 0.5 | 0.3612856 | 0.54526022 | 0.017004145313 | 0.03143803 |
| PAF47 | Blocktapping | 0.2455364 | 0.7544636 | 0.5 | 0.3254450 | 0.83685448 | -0.005359508652 | 0.02820654 |
| PAF47 | Sum_PC | 0.2465187 | 0.7534813 | 0.5 | 0.3271730 | 0.76003575 | 0.007915078699 | 0.02915331 |
| PAF48 | Blocktapping | 0.2521397 | 0.7478603 | 0.5 | 0.3371482 | 0.71076018 | 0.010580985812 | 0.02798564 |
| PAF48 | Sum_PC | 0.2451837 | 0.7548163 | 0.5 | 0.3248256 | 0.87133742 | 0.005848307654 | 0.02929852 |
| PAF49 | Blocktapping | 0.1948457 | 0.8051543 | 0.5 | 0.2419980 | 0.97147043 | -0.000046074436 | 0.02229301 |
| PAF49 | Sum_PC | 0.2137614 | 0.7862386 | 0.5 | 0.2718786 | 0.53073248 | 0.014073762185 | 0.02673834 |
| PAF5 | Blocktapping | 0.1992169 | 0.8007831 | 0.5 | 0.2487776 | 0.62286832 | -0.011258826677 | 0.02351057 |
| PAF5 | Sum_PC | 0.1938098 | 0.8061902 | 0.5 | 0.2404021 | 0.64896983 | 0.010050220469 | 0.02250957 |
| PAF50 | Blocktapping | 0.2206099 | 0.7793901 | 0.5 | 0.2830546 | 0.56752591 | 0.013806485589 | 0.02538652 |
| PAF50 | Sum_PC | 0.2172564 | 0.7827436 | 0.5 | 0.2775576 | 0.68593421 | 0.010950090886 | 0.02516760 |
| PAF51 | Blocktapping | 0.2066670 | 0.7933330 | 0.5 | 0.2605047 | 0.58059414 | -0.011780373508 | 0.02287410 |
| PAF51 | Sum_PC | 0.2276741 | 0.7723259 | 0.5 | 0.2947901 | 0.38959348 | 0.021428111700 | 0.02782796 |
| PAF52 | Blocktapping | 0.1966949 | 0.8033051 | 0.5 | 0.2448570 | 0.63470284 | -0.010728401992 | 0.02278230 |
| PAF52 | Sum_PC | 0.1884695 | 0.8115305 | 0.5 | 0.2322396 | 0.71968419 | 0.006842314518 | 0.02177220 |
| PAF53 | Blocktapping | 0.1823284 | 0.8176716 | 0.5 | 0.2229849 | 0.95603341 | -0.002825835712 | 0.02234365 |
| PAF53 | Sum_PC | 0.1734226 | 0.8265774 | 0.5 | 0.2098080 | 0.89705540 | -0.002168689196 | 0.02179641 |
| PAF54 | Blocktapping | 0.2032350 | 0.7967650 | 0.5 | 0.2550752 | 0.63171317 | -0.011003494856 | 0.02429297 |
| PAF54 | Sum_PC | 0.1940108 | 0.8059892 | 0.5 | 0.2407114 | 0.73164859 | 0.007014052136 | 0.02255058 |
| PAF55 | Blocktapping | 0.2100466 | 0.7899534 | 0.5 | 0.2658974 | 0.52814407 | -0.013871166391 | 0.02403995 |
| PAF55 | Sum_PC | 0.2090823 | 0.7909177 | 0.5 | 0.2643541 | 0.51991315 | 0.013715496327 | 0.02478572 |
| PAF56 | Blocktapping | 0.1992126 | 0.8007874 | 0.5 | 0.2487709 | 0.71373016 | -0.007331319591 | 0.02245639 |
| PAF56 | Sum_PC | 0.2182704 | 0.7817296 | 0.5 | 0.2792147 | 0.45920718 | 0.017835160318 | 0.02631962 |
| PAF57 | Blocktapping | 0.1811303 | 0.8188697 | 0.5 | 0.2211955 | 0.81629079 | -0.005034051922 | 0.02159372 |
| PAF57 | Sum_PC | 0.1797330 | 0.8202670 | 0.5 | 0.2191153 | 0.84373542 | 0.004339513790 | 0.02168520 |
| PAF58 | Blocktapping | 0.2025802 | 0.7974198 | 0.5 | 0.2540445 | 0.53514784 | -0.013500676848 | 0.02290855 |
| PAF58 | Sum_PC | 0.1966392 | 0.8033608 | 0.5 | 0.2447707 | 0.58345681 | 0.011411196678 | 0.02278256 |
| PAF59 | Blocktapping | 0.2170413 | 0.7829587 | 0.5 | 0.2772066 | 0.63907776 | -0.010256493175 | 0.02453756 |
| PAF59 | Sum_PC | 0.2367262 | 0.7632738 | 0.5 | 0.3101458 | 0.43744705 | 0.019968289431 | 0.02925424 |
| PAF6 | Blocktapping | 0.2050748 | 0.7949252 | 0.5 | 0.2579801 | 0.72988265 | -0.006517533663 | 0.02276701 |
| PAF6 | Sum_PC | 0.2287663 | 0.7712337 | 0.5 | 0.2966238 | 0.42880313 | 0.020306288775 | 0.02734865 |
| PAF60 | Blocktapping | 0.2239172 | 0.7760828 | 0.5 | 0.2885223 | 0.88364130 | -0.003379914740 | 0.02331555 |
| PAF60 | Sum_PC | 0.2779734 | 0.7220266 | 0.5 | 0.3849905 | 0.32882838 | 0.031117296656 | 0.03276824 |
| PAF61 | Blocktapping | 0.2441164 | 0.7558836 | 0.5 | 0.3229551 | 0.50785630 | -0.016216881898 | 0.02810461 |
| PAF61 | Sum_PC | 0.2402247 | 0.7597753 | 0.5 | 0.3161786 | 0.52955528 | 0.014750652995 | 0.02898788 |

| DV | Predictor | Sum_PostModProb_Incl | Sum_PostModProb_Excl | Sum_PriorModProb_Incl | BF_Incl | Frequentist_p_value | postMean | postStd |
|---|---|---|---|---|---|---|---|---|
| PAF62 | Blocktapping | 0.2206693 | 0.7793307 | 0.5 | 0.2831522 | 0.81419821 | -0.005291176564 | 0.02502996 |
| PAF62 | Sum_PC | 0.2275769 | 0.7724231 | 0.5 | 0.2946273 | 0.64720145 | 0.009310239636 | 0.02780873 |
| PAF63 | Blocktapping | 0.2438705 | 0.7561295 | 0.5 | 0.3225248 | 0.95871220 | 0.001144197417 | 0.02776044 |
| PAF63 | Sum_PC | 0.2638770 | 0.7361230 | 0.5 | 0.3584686 | 0.42536884 | 0.023760271794 | 0.03043403 |
| PAF64 | Blocktapping | 0.2347360 | 0.7652640 | 0.5 | 0.3067386 | 0.73623814 | 0.008966655835 | 0.02548742 |
| PAF64 | Sum_PC | 0.2578593 | 0.7421407 | 0.5 | 0.3474534 | 0.46906465 | 0.021642169257 | 0.03021804 |
| PAF65 | Blocktapping | 0.2378484 | 0.7621516 | 0.5 | 0.3120749 | 0.59977840 | 0.012071586819 | 0.02567696 |
| PAF65 | Sum_PC | 0.2468535 | 0.7531465 | 0.5 | 0.3277630 | 0.52702216 | 0.016884061140 | 0.02988713 |
| PAF66 | Blocktapping | 0.2598569 | 0.7401431 | 0.5 | 0.3510901 | 0.40615807 | 0.021595391390 | 0.02929148 |
| PAF66 | Sum_PC | 0.2241108 | 0.7758892 | 0.5 | 0.2888438 | 0.98353193 | 0.001374547958 | 0.02495530 |
| PAF67 | Blocktapping | 0.1782244 | 0.8217756 | 0.5 | 0.2168772 | 0.95844081 | -0.001556553587 | 0.02110863 |
| PAF67 | Sum_PC | 0.1780930 | 0.8219070 | 0.5 | 0.2166826 | 0.90056296 | -0.002425783848 | 0.02223279 |
| PAF68 | Blocktapping | 0.2271575 | 0.7728425 | 0.5 | 0.2939247 | 0.91016404 | -0.001961264008 | 0.02426088 |
| PAF68 | Sum_PC | 0.2791764 | 0.7208236 | 0.5 | 0.3873020 | 0.33655527 | 0.029534624590 | 0.03357944 |
| PAF69 | Blocktapping | 0.2232477 | 0.7767523 | 0.5 | 0.2874117 | 0.87500094 | 0.003342336377 | 0.02474571 |
| PAF69 | Sum_PC | 0.2567749 | 0.7432251 | 0.5 | 0.3454874 | 0.43657448 | 0.022625193653 | 0.03029533 |
| PAF7 | Blocktapping | 0.1965447 | 0.8034553 | 0.5 | 0.2446243 | 0.66046882 | -0.009440284085 | 0.02306292 |
| PAF7 | Sum_PC | 0.1895142 | 0.8104858 | 0.5 | 0.2338280 | 0.81019390 | 0.004302156390 | 0.02229547 |
| PAF70 | Blocktapping | 0.2064308 | 0.7935692 | 0.5 | 0.2601295 | 0.65611401 | 0.009550915270 | 0.02407355 |
| PAF70 | Sum_PC | 0.2006649 | 0.7993351 | 0.5 | 0.2510397 | 0.83457047 | 0.003987348608 | 0.02376376 |
| PAF71 | Blocktapping | 0.2350255 | 0.7649745 | 0.5 | 0.3072331 | 0.50329471 | 0.016521456767 | 0.02654787 |
| PAF71 | Sum_PC | 0.2128858 | 0.7871142 | 0.5 | 0.2704637 | 0.95609358 | 0.002018452007 | 0.02505007 |
| PAF72 | Blocktapping | 0.2607168 | 0.7392832 | 0.5 | 0.3526616 | 0.75009197 | -0.006251437874 | 0.02646599 |
| PAF72 | Sum_PC | 0.3299576 | 0.6700424 | 0.5 | 0.4924428 | 0.25709500 | 0.038105689980 | 0.03691827 |
| PAF73 | Blocktapping | 0.2581494 | 0.7418506 | 0.5 | 0.3479802 | 0.62661006 | 0.013073770840 | 0.02837805 |
| PAF73 | Sum_PC | 0.2577935 | 0.7422065 | 0.5 | 0.3473339 | 0.65648865 | 0.012632701095 | 0.02957740 |
| PAF74 | Blocktapping | 0.1898388 | 0.8101612 | 0.5 | 0.2343223 | 0.92001599 | -0.002112725357 | 0.02136164 |
| PAF74 | Sum_PC | 0.2051528 | 0.7948472 | 0.5 | 0.2581035 | 0.58906669 | 0.012792070709 | 0.02474550 |
| PAF75 | Blocktapping | 0.2379085 | 0.7620915 | 0.5 | 0.3121784 | 0.66665780 | -0.010436308740 | 0.02727045 |
| PAF75 | Sum_PC | 0.2555745 | 0.7444255 | 0.5 | 0.3433177 | 0.38932256 | 0.024105298832 | 0.03021211 |
| PAF76 | Blocktapping | 0.2882321 | 0.7117679 | 0.5 | 0.4049523 | 0.47044494 | -0.017209299503 | 0.02671509 |
| PAF76 | Sum_PC | 0.3853556 | 0.6146444 | 0.5 | 0.6269569 | 0.15340486 | 0.056944973902 | 0.04243279 |
| PAF77 | Blocktapping | 0.2784629 | 0.7215371 | 0.5 | 0.3859301 | 0.38450254 | 0.024889196313 | 0.03198593 |
| PAF77 | Sum_PC | 0.2517886 | 0.7482114 | 0.5 | 0.3365207 | 0.59546774 | 0.014971899571 | 0.02765323 |
| PAF78 | Blocktapping | 0.2815515 | 0.7184485 | 0.5 | 0.3918882 | 0.48751981 | 0.018999400171 | 0.03125637 |
| PAF78 | Sum_PC | 0.2514875 | 0.7485125 | 0.5 | 0.3359831 | 0.86875391 | 0.004581591135 | 0.02840371 |
| PAF79 | Blocktapping | 0.3129219 | 0.6870781 | 0.5 | 0.4554386 | 0.36287092 | -0.026245780238 | 0.03247832 |
| PAF79 | Sum_PC | 0.3329170 | 0.6670830 | 0.5 | 0.4990638 | 0.27287177 | 0.035591880642 | 0.03753467 |
| PAF8 | Blocktapping | 0.1899877 | 0.8100123 | 0.5 | 0.2345491 | 0.94987751 | -0.000588838106 | 0.02183281 |
| PAF8 | Sum_PC | 0.2171789 | 0.7828211 | 0.5 | 0.2774311 | 0.48039246 | 0.016658723316 | 0.02643264 |
| PAF80 | Blocktapping | 0.2786984 | 0.7213016 | 0.5 | 0.3863826 | 0.41247424 | -0.018765758375 | 0.02676835 |

| DV | Predictor | Sum_PostModProb_Incl | Sum_PostModProb_Excl | Sum_PriorModProb_Incl | BF_Incl | Frequentist_p_value | postMean | postStd |
|---|---|---|---|---|---|---|---|---|
| PAF80 | Sum_PC | 0.3751361 | 0.6248639 | 0.5 | 0.6003484 | 0.14015528 | 0.057809580808 | 0.04232985 |
| PAF81 | Blocktapping | 0.2431069 | 0.7568931 | 0.5 | 0.3211906 | 0.80851523 | -0.005702972830 | 0.02408082 |
| PAF81 | Sum_PC | 0.3183104 | 0.6816896 | 0.5 | 0.4669432 | 0.24842471 | 0.037575723964 | 0.03636231 |
| PAF82 | Blocktapping | 0.2048513 | 0.7951487 | 0.5 | 0.2576264 | 0.84909180 | -0.004502811614 | 0.02330147 |
| PAF82 | Sum_PC | 0.2153949 | 0.7846051 | 0.5 | 0.2745265 | 0.61436592 | 0.012280082237 | 0.02519184 |
| PAF83 | Blocktapping | 0.2683477 | 0.7316523 | 0.5 | 0.3667694 | 0.55922109 | -0.014496868393 | 0.02911119 |
| PAF83 | Sum_PC | 0.2805343 | 0.7194657 | 0.5 | 0.3899203 | 0.45360824 | 0.021112649400 | 0.03300760 |
| PAF84 | Blocktapping | 0.2897814 | 0.7102186 | 0.5 | 0.4080171 | 0.43185819 | -0.019452335801 | 0.03128927 |
| PAF84 | Sum_PC | 0.2990464 | 0.7009536 | 0.5 | 0.4266280 | 0.36419559 | 0.026447294975 | 0.03431868 |
| PAF85 | Blocktapping | 0.2724501 | 0.7275499 | 0.5 | 0.3744762 | 0.50109882 | -0.018144514223 | 0.03049475 |
| PAF85 | Sum_PC | 0.2599430 | 0.7400570 | 0.5 | 0.3512473 | 0.60684716 | 0.013961104580 | 0.02950153 |
| PAF86 | Blocktapping | 0.2514141 | 0.7485859 | 0.5 | 0.3358520 | 0.98467910 | 0.000326055212 | 0.02639313 |
| PAF86 | Sum_PC | 0.2915737 | 0.7084263 | 0.5 | 0.4115795 | 0.42388618 | 0.025083895168 | 0.03406181 |
| PAF87 | Blocktapping | 0.2578955 | 0.7421045 | 0.5 | 0.3475191 | 0.46269176 | 0.019403313574 | 0.02870370 |
| PAF87 | Sum_PC | 0.2638799 | 0.7361201 | 0.5 | 0.3584739 | 0.43098169 | 0.023247786158 | 0.03011383 |
| PAF88 | Blocktapping | 0.2436696 | 0.7563304 | 0.5 | 0.3221734 | 0.90569290 | -0.003073673057 | 0.02653427 |
| PAF88 | Sum_PC | 0.2643743 | 0.7356257 | 0.5 | 0.3593869 | 0.53708202 | 0.018126254520 | 0.03125592 |
| PAF89 | Blocktapping | 0.2029606 | 0.7970394 | 0.5 | 0.2546431 | 0.58374067 | 0.011604308758 | 0.02410393 |
| PAF89 | Sum_PC | 0.1920931 | 0.8079069 | 0.5 | 0.2377664 | 0.80232895 | 0.005899714693 | 0.02260101 |
| PAF9 | Blocktapping | 0.2002203 | 0.7997797 | 0.5 | 0.2503444 | 0.66775129 | -0.009024378160 | 0.02197696 |
| PAF9 | Sum_PC | 0.2330725 | 0.7669275 | 0.5 | 0.3039043 | 0.46451587 | 0.018485730446 | 0.02743837 |
| PAF90 | Blocktapping | 0.2161809 | 0.7838191 | 0.5 | 0.2758047 | 0.78046861 | -0.005757457867 | 0.02324319 |
| PAF90 | Sum_PC | 0.2501295 | 0.7498705 | 0.5 | 0.3335636 | 0.39890386 | 0.023060727662 | 0.03004958 |
| PAF91 | Blocktapping | 0.2010023 | 0.7989977 | 0.5 | 0.2515681 | 0.62825283 | -0.009996448482 | 0.02352728 |
| PAF91 | Sum_PC | 0.2044899 | 0.7955101 | 0.5 | 0.2570551 | 0.55674437 | 0.012656499378 | 0.02396009 |
| PAF92 | Blocktapping | 0.2781743 | 0.7218257 | 0.5 | 0.3853760 | 0.43410990 | -0.017659122324 | 0.02719357 |
| PAF92 | Sum_PC | 0.3607502 | 0.6392498 | 0.5 | 0.5643337 | 0.16476707 | 0.051572531303 | 0.04236389 |
| PAF93 | Blocktapping | 0.1985968 | 0.8014032 | 0.5 | 0.2478113 | 0.67686731 | -0.009604143904 | 0.02378561 |
| PAF93 | Sum_PC | 0.2032775 | 0.7967225 | 0.5 | 0.2551422 | 0.57012701 | 0.012992175211 | 0.02534227 |
| PAF94 | Blocktapping | 0.3275992 | 0.6724008 | 0.5 | 0.4872083 | 0.30878066 | -0.029454032731 | 0.03088666 |
| PAF94 | Sum_PC | 0.3858316 | 0.6141684 | 0.5 | 0.6282179 | 0.16273734 | 0.051126442460 | 0.04187820 |
| PAF95 | Blocktapping | 0.3332702 | 0.6667298 | 0.5 | 0.4998580 | 0.33991356 | -0.026661937021 | 0.03173707 |
| PAF95 | Sum_PC | 0.4218125 | 0.5781875 | 0.5 | 0.7295426 | 0.13006604 | 0.063702953164 | 0.04511734 |
| PAF96 | Blocktapping | 0.2895604 | 0.7104396 | 0.5 | 0.4075792 | 0.44648866 | -0.019687030639 | 0.03007311 |
| PAF96 | Sum_PC | 0.3232444 | 0.6767556 | 0.5 | 0.4776384 | 0.28482468 | 0.033837651432 | 0.03709454 |
| PAF97 | Blocktapping | 0.2503786 | 0.7496214 | 0.5 | 0.3340067 | 0.67782115 | -0.008837250488 | 0.02648290 |
| PAF97 | Sum_PC | 0.2885039 | 0.7114961 | 0.5 | 0.4054890 | 0.51312597 | 0.018009260721 | 0.03393446 |
| PAF98 | Blocktapping | 0.2451592 | 0.7548408 | 0.5 | 0.3247826 | 0.97417955 | -0.000524002966 | 0.02731159 |
| PAF98 | Sum_PC | 0.2507417 | 0.7492583 | 0.5 | 0.3346532 | 0.70802868 | 0.010596132861 | 0.02980958 |
| PAF99 | Blocktapping | 0.1919401 | 0.8080599 | 0.5 | 0.2375321 | 0.95951588 | -0.000732799275 | 0.02138221 |
| PAF99 | Sum_PC | 0.2188415 | 0.7811585 | 0.5 | 0.2801499 | 0.48268049 | 0.018090892185 | 0.02635704 |

| DV | Predictor | Sum_PostModProb_Incl | Sum_PostModProb_Excl | Sum_PriorModProb_Incl | BF_Incl | Frequentist_p_value | postMean | postStd |
|---|---|---|---|---|---|---|---|---|
| theta_ROI1_AECnodestrength | Blocktapping | 0.2798147 | 0.7201853 | 0.5 | 0.3885316 | 0.53417171 | -0.017763939338 | 0.03185615 |
| theta_ROI1_AECnodestrength | Sum_PC | 0.2419252 | 0.7580748 | 0.5 | 0.3191310 | 0.92317200 | 0.001934109313 | 0.02690396 |
| theta_ROI10_AECnodestrength | Blocktapping | 0.2413839 | 0.7586161 | 0.5 | 0.3181898 | 0.76652045 | -0.007661640326 | 0.02744824 |
| theta_ROI10_AECnodestrength | Sum_PC | 0.2356734 | 0.7643266 | 0.5 | 0.3083413 | 0.82461052 | 0.005915703007 | 0.02876128 |
| theta_ROI100_AECnodestrength | Blocktapping | 0.3385883 | 0.6614117 | 0.5 | 0.5119177 | 0.28846677 | -0.036187505114 | 0.03586777 |
| theta_ROI100_AECnodestrength | Sum_PC | 0.2783246 | 0.7216754 | 0.5 | 0.3856645 | 0.65994052 | 0.010276983394 | 0.02983603 |
| theta_ROI11_AECnodestrength | Blocktapping | 0.2561153 | 0.7438847 | 0.5 | 0.3442943 | 0.57034828 | -0.014944683279 | 0.02811292 |
| theta_ROI11_AECnodestrength | Sum_PC | 0.2379706 | 0.7620294 | 0.5 | 0.3122853 | 0.85322909 | 0.003481152833 | 0.02763763 |
| theta_ROI12_AECnodestrength | Blocktapping | 0.2371051 | 0.7628949 | 0.5 | 0.3107965 | 0.73546886 | -0.008104820416 | 0.02601865 |
| theta_ROI12_AECnodestrength | Sum_PC | 0.2380984 | 0.7619016 | 0.5 | 0.3125054 | 0.70400451 | 0.009731142567 | 0.02821172 |
| theta_ROI13_AECnodestrength | Blocktapping | 0.2315594 | 0.7684406 | 0.5 | 0.3013367 | 0.82993726 | -0.004073963088 | 0.02693227 |
| theta_ROI13_AECnodestrength | Sum_PC | 0.2362256 | 0.7637744 | 0.5 | 0.3092872 | 0.70226521 | 0.009116941657 | 0.02876022 |
| theta_ROI14_AECnodestrength | Blocktapping | 0.3489807 | 0.6510193 | 0.5 | 0.5360527 | 0.24365264 | -0.037724640038 | 0.03813747 |
| theta_ROI14_AECnodestrength | Sum_PC | 0.2984111 | 0.7015889 | 0.5 | 0.4253360 | 0.43776915 | 0.020762877290 | 0.03146300 |
| theta_ROI15_AECnodestrength | Blocktapping | 0.3766079 | 0.6233921 | 0.5 | 0.6041268 | 0.21109006 | -0.046675319137 | 0.04001066 |
| theta_ROI15_AECnodestrength | Sum_PC | 0.3137125 | 0.6862875 | 0.5 | 0.4571152 | 0.43080613 | 0.022860721326 | 0.03177325 |
| theta_ROI16_AECnodestrength | Blocktapping | 0.2844238 | 0.7155762 | 0.5 | 0.3974752 | 0.45230241 | -0.021827665683 | 0.03252344 |
| theta_ROI16_AECnodestrength | Sum_PC | 0.2497797 | 0.7502203 | 0.5 | 0.3329419 | 0.96277724 | 0.001950362937 | 0.02788730 |
| theta_ROI17_AECnodestrength | Blocktapping | 0.2605283 | 0.7394717 | 0.5 | 0.3523168 | 0.57702822 | -0.015390215067 | 0.02879666 |
| theta_ROI17_AECnodestrength | Sum_PC | 0.2556330 | 0.7443670 | 0.5 | 0.3434233 | 0.58570047 | -0.014681262771 | 0.02973732 |
| theta_ROI18_AECnodestrength | Blocktapping | 0.3147779 | 0.6852221 | 0.5 | 0.4593808 | 0.34157114 | -0.030420234608 | 0.03365174 |
| theta_ROI18_AECnodestrength | Sum_PC | 0.2728733 | 0.7271267 | 0.5 | 0.3752762 | 0.61144143 | 0.012938513645 | 0.02976651 |
| theta_ROI19_AECnodestrength | Blocktapping | 0.3335231 | 0.6664769 | 0.5 | 0.5004272 | 0.27687479 | -0.034561328214 | 0.03484778 |
| theta_ROI19_AECnodestrength | Sum_PC | 0.2894858 | 0.7105142 | 0.5 | 0.4074314 | 0.49300873 | 0.017027852070 | 0.03079936 |
| theta_ROI2_AECnodestrength | Blocktapping | 0.2951294 | 0.7048706 | 0.5 | 0.4187001 | 0.39721430 | -0.026284298382 | 0.03354605 |
| theta_ROI2_AECnodestrength | Sum_PC | 0.2512931 | 0.7487069 | 0.5 | 0.3356361 | 0.99429824 | -0.001426738471 | 0.02793408 |
| theta_ROI20_AECnodestrength | Blocktapping | 0.3115309 | 0.6884691 | 0.5 | 0.4524981 | 0.37272900 | -0.026587767046 | 0.03277188 |
| theta_ROI20_AECnodestrength | Sum_PC | 0.2918879 | 0.7081121 | 0.5 | 0.4122058 | 0.47672563 | 0.018564744206 | 0.03214686 |
| theta_ROI21_AECnodestrength | Blocktapping | 0.3166666 | 0.6833334 | 0.5 | 0.4634144 | 0.34312623 | -0.028528327739 | 0.03358785 |

| DV | Predictor | Sum_PostModProb_Incl | Sum_PostModProb_Excl | Sum_PriorModProb_Incl | BF_Incl | Frequentist_p_value | postMean | postStd |
|---|---|---|---|---|---|---|---|---|
| theta_ROI21_AECnodestrength | Sum_PC | 0.2995267 | 0.7004733 | 0.5 | 0.4276061 | 0.43114159 | 0.021246810662 | 0.03220896 |
| theta_ROI22_AECnodestrength | Blocktapping | 0.2338362 | 0.7661638 | 0.5 | 0.3052038 | 0.85844885 | -0.004632650008 | 0.02492808 |
| theta_ROI22_AECnodestrength | Sum_PC | 0.2609057 | 0.7390943 | 0.5 | 0.3530074 | 0.44338336 | 0.020853746465 | 0.03005568 |
| theta_ROI23_AECnodestrength | Blocktapping | 0.3085560 | 0.6914440 | 0.5 | 0.4462487 | 0.43341907 | -0.021348121982 | 0.03142462 |
| theta_ROI23_AECnodestrength | Sum_PC | 0.3549872 | 0.6450128 | 0.5 | 0.5503567 | 0.23557570 | 0.042085468391 | 0.03899677 |
| theta_ROI24_AECnodestrength | Blocktapping | 0.2408605 | 0.7591395 | 0.5 | 0.3172809 | 0.69600123 | -0.010111814142 | 0.02770313 |
| theta_ROI24_AECnodestrength | Sum_PC | 0.2347516 | 0.7652484 | 0.5 | 0.3067653 | 0.87684114 | -0.004775666682 | 0.02773361 |
| theta_ROI25_AECnodestrength | Blocktapping | 0.2095147 | 0.7904853 | 0.5 | 0.2650456 | 0.62464744 | -0.011791847980 | 0.02374860 |
| theta_ROI25_AECnodestrength | Sum_PC | 0.2035243 | 0.7964757 | 0.5 | 0.2555310 | 0.73799289 | 0.006843275417 | 0.02445672 |
| theta_ROI26_AECnodestrength | Blocktapping | 0.2373544 | 0.7626456 | 0.5 | 0.3112250 | 0.86081003 | -0.004361183312 | 0.02599589 |
| theta_ROI26_AECnodestrength | Sum_PC | 0.2504859 | 0.7495141 | 0.5 | 0.3341977 | 0.59052018 | 0.014460493918 | 0.02980765 |
| theta_ROI27_AECnodestrength | Blocktapping | 0.2886019 | 0.7113981 | 0.5 | 0.4056827 | 0.45338901 | -0.019324626201 | 0.02972773 |
| theta_ROI27_AECnodestrength | Sum_PC | 0.3047930 | 0.6952070 | 0.5 | 0.4384204 | 0.35399698 | 0.027942372851 | 0.03514534 |
| theta_ROI28_AECnodestrength | Blocktapping | 0.2888797 | 0.7111203 | 0.5 | 0.4062318 | 0.40189909 | -0.023665314964 | 0.03026244 |
| theta_ROI28_AECnodestrength | Sum_PC | 0.2928050 | 0.7071950 | 0.5 | 0.4140372 | 0.37030361 | 0.025573995395 | 0.03237328 |
| theta_ROI29_AECnodestrength | Blocktapping | 0.3087369 | 0.6912631 | 0.5 | 0.4466272 | 0.37270468 | -0.027587835438 | 0.03334061 |
| theta_ROI29_AECnodestrength | Sum_PC | 0.2603100 | 0.7396900 | 0.5 | 0.3519177 | 0.68298593 | 0.008677955442 | 0.02934586 |
| theta_ROI3_AECnodestrength | Blocktapping | 0.2830391 | 0.7169609 | 0.5 | 0.3947761 | 0.44076475 | -0.022486841765 | 0.03208941 |
| theta_ROI3_AECnodestrength | Sum_PC | 0.2530211 | 0.7469789 | 0.5 | 0.3387260 | 0.74488221 | 0.007171330306 | 0.02842633 |
| theta_ROI30_AECnodestrength | Blocktapping | 0.3058838 | 0.6941162 | 0.5 | 0.4406810 | 0.43533591 | -0.021824941877 | 0.03103498 |
| theta_ROI30_AECnodestrength | Sum_PC | 0.3237172 | 0.6762828 | 0.5 | 0.4786714 | 0.31820781 | 0.031819206850 | 0.03558363 |
| theta_ROI31_AECnodestrength | Blocktapping | 0.2396335 | 0.7603665 | 0.5 | 0.3151553 | 0.72985113 | -0.007201231578 | 0.02522546 |
| theta_ROI31_AECnodestrength | Sum_PC | 0.2627879 | 0.7372121 | 0.5 | 0.3564618 | 0.45336122 | 0.021514094132 | 0.03120913 |
| theta_ROI32_AECnodestrength | Blocktapping | 0.2400224 | 0.7599776 | 0.5 | 0.3158283 | 0.77824981 | -0.006456909668 | 0.02804710 |
| theta_ROI32_AECnodestrength | Sum_PC | 0.2341450 | 0.7658550 | 0.5 | 0.3057303 | 0.99192875 | -0.000494482582 | 0.02765472 |
| theta_ROI33_AECnodestrength | Blocktapping | 0.2532556 | 0.7467444 | 0.5 | 0.3391462 | 0.70476861 | -0.009199144075 | 0.02862293 |
| theta_ROI33_AECnodestrength | Sum_PC | 0.2490396 | 0.7509604 | 0.5 | 0.3316281 | 0.73666196 | 0.009138714386 | 0.02863149 |
| theta_ROI34_AECnodestrength | Blocktapping | 0.3628412 | 0.6371588 | 0.5 | 0.5694674 | 0.23347257 | -0.044261572788 | 0.04005229 |
| theta_ROI34_AECnodestrength | Sum_PC | 0.2768533 | 0.7231467 | 0.5 | 0.3828452 | 0.66649388 | 0.009843938193 | 0.02840668 |

| DV | Predictor | Sum_PostModProb_Incl | Sum_PostModProb_Excl | Sum_PriorModProb_Incl | BF_Incl | Frequentist_p_value | postMean | postStd |
|---|---|---|---|---|---|---|---|---|
| theta_ROI35_AECnodestrength | Blocktapping | 0.2540793 | 0.7459207 | 0.5 | 0.3406251 | 0.81200852 | -0.005050622971 | 0.02692430 |
| theta_ROI35_AECnodestrength | Sum_PC | 0.2929520 | 0.7070480 | 0.5 | 0.4143311 | 0.39479868 | 0.026466994342 | 0.03424253 |
| theta_ROI36_AECnodestrength | Blocktapping | 0.2994936 | 0.7005064 | 0.5 | 0.4275386 | 0.37808501 | -0.024687413802 | 0.03301158 |
| theta_ROI36_AECnodestrength | Sum_PC | 0.2668521 | 0.7331479 | 0.5 | 0.3639813 | 0.61906930 | 0.011697670178 | 0.02925067 |
| theta_ROI37_AECnodestrength | Blocktapping | 0.2667965 | 0.7332035 | 0.5 | 0.3638778 | 0.54159567 | -0.017339550079 | 0.02946556 |
| theta_ROI37_AECnodestrength | Sum_PC | 0.2573549 | 0.7426451 | 0.5 | 0.3465382 | 0.63003909 | 0.011774951568 | 0.02925939 |
| theta_ROI38_AECnodestrength | Blocktapping | 0.2453110 | 0.7546890 | 0.5 | 0.3250491 | 0.78318117 | -0.007864783525 | 0.02753170 |
| theta_ROI38_AECnodestrength | Sum_PC | 0.2492660 | 0.7507340 | 0.5 | 0.3320296 | 0.66350437 | 0.010361147609 | 0.02935474 |
| theta_ROI39_AECnodestrength | Blocktapping | 0.2463214 | 0.7536786 | 0.5 | 0.3268256 | 0.78378002 | -0.007990979248 | 0.02754287 |
| theta_ROI39_AECnodestrength | Sum_PC | 0.2433204 | 0.7566796 | 0.5 | 0.3215633 | 0.75325455 | 0.007703256630 | 0.02858516 |
| theta_ROI4_AECnodestrength | Blocktapping | 0.2723295 | 0.7276705 | 0.5 | 0.3742484 | 0.52875935 | -0.016406374961 | 0.03078049 |
| theta_ROI4_AECnodestrength | Sum_PC | 0.2601275 | 0.7398725 | 0.5 | 0.3515843 | 0.68038628 | 0.009895737208 | 0.03055599 |
| theta_ROI40_AECnodestrength | Blocktapping | 0.2475098 | 0.7524902 | 0.5 | 0.3289210 | 0.72091152 | -0.010339833355 | 0.02878898 |
| theta_ROI40_AECnodestrength | Sum_PC | 0.2388236 | 0.7611764 | 0.5 | 0.3137560 | 0.90052647 | -0.003529812167 | 0.02807095 |
| theta_ROI41_AECnodestrength | Blocktapping | 0.3128919 | 0.6871081 | 0.5 | 0.4553750 | 0.33620684 | -0.032329525513 | 0.03564529 |
| theta_ROI41_AECnodestrength | Sum_PC | 0.2594969 | 0.7405031 | 0.5 | 0.3504332 | 0.97386686 | -0.001658180959 | 0.02797866 |
| theta_ROI42_AECnodestrength | Blocktapping | 0.2592474 | 0.7407526 | 0.5 | 0.3499783 | 0.66654123 | -0.012201691637 | 0.03046887 |
| theta_ROI42_AECnodestrength | Sum_PC | 0.2284636 | 0.7715364 | 0.5 | 0.2961152 | 0.86483156 | 0.004143575866 | 0.02636025 |
| theta_ROI43_AECnodestrength | Blocktapping | 0.2387574 | 0.7612426 | 0.5 | 0.3136417 | 0.80426564 | -0.006777049022 | 0.02735930 |
| theta_ROI43_AECnodestrength | Sum_PC | 0.2385712 | 0.7614288 | 0.5 | 0.3133204 | 0.75023043 | 0.007544209440 | 0.02838929 |
| theta_ROI44_AECnodestrength | Blocktapping | 0.2850077 | 0.7149923 | 0.5 | 0.3986165 | 0.47096624 | -0.017388098930 | 0.02932174 |
| theta_ROI44_AECnodestrength | Sum_PC | 0.3046701 | 0.6953299 | 0.5 | 0.4381662 | 0.34571076 | 0.030348997210 | 0.03388648 |
| theta_ROI45_AECnodestrength | Blocktapping | 0.2608478 | 0.7391522 | 0.5 | 0.3529014 | 0.83040953 | -0.003818049158 | 0.02738605 |
| theta_ROI45_AECnodestrength | Sum_PC | 0.2927431 | 0.7072569 | 0.5 | 0.4139133 | 0.41457291 | 0.024342728466 | 0.03363628 |
| theta_ROI46_AECnodestrength | Blocktapping | 0.3491689 | 0.6508311 | 0.5 | 0.5364970 | 0.23042943 | -0.040737656610 | 0.03740430 |
| theta_ROI46_AECnodestrength | Sum_PC | 0.2956263 | 0.7043737 | 0.5 | 0.4197009 | 0.42298745 | 0.021390017277 | 0.03063604 |
| theta_ROI47_AECnodestrength | Blocktapping | 0.3276436 | 0.6723564 | 0.5 | 0.4873064 | 0.43445395 | -0.020606538780 | 0.03151993 |
| theta_ROI47_AECnodestrength | Sum_PC | 0.4229447 | 0.5770553 | 0.5 | 0.7329363 | 0.14631548 | 0.059358578083 | 0.04713096 |
| theta_ROI48_AECnodestrength | Blocktapping | 0.3113113 | 0.6886887 | 0.5 | 0.4520349 | 0.40972258 | -0.025312821414 | 0.03408029 |

| DV | Predictor | Sum_PostModProb_Incl | Sum_PostModProb_Excl | Sum_PriorModProb_Incl | BF_Incl | Frequentist_p_value | postMean | postStd |
|---|---|---|---|---|---|---|---|---|
| theta_ROI48_AECnodestrength | Sum_PC | 0.2805519 | 0.7194481 | 0.5 | 0.3899543 | 0.36066546 | 0.025720700652 | 0.03029790 |
| theta_ROI49_AECnodestrength | Blocktapping | 0.2977609 | 0.7022391 | 0.5 | 0.4240164 | 0.39514158 | -0.025132559937 | 0.03244861 |
| theta_ROI49_AECnodestrength | Sum_PC | 0.2582006 | 0.7417994 | 0.5 | 0.3480733 | 0.73325509 | 0.007671540732 | 0.02827608 |
| theta_ROI5_AECnodestrength | Blocktapping | 0.2837092 | 0.7162908 | 0.5 | 0.3960811 | 0.42147932 | -0.023039192607 | 0.03272989 |
| theta_ROI5_AECnodestrength | Sum_PC | 0.2543274 | 0.7456726 | 0.5 | 0.3410711 | 0.72856364 | 0.008058787191 | 0.02808227 |
| theta_ROI50_AECnodestrength | Blocktapping | 0.3046422 | 0.6953578 | 0.5 | 0.4381086 | 0.38008738 | -0.027004480535 | 0.03374023 |
| theta_ROI50_AECnodestrength | Sum_PC | 0.2577980 | 0.7422020 | 0.5 | 0.3473422 | 0.91496657 | 0.002368052100 | 0.02843486 |
| theta_ROI51_AECnodestrength | Blocktapping | 0.2906744 | 0.7093256 | 0.5 | 0.4097898 | 0.43960456 | -0.021934889866 | 0.03243184 |
| theta_ROI51_AECnodestrength | Sum_PC | 0.2670540 | 0.7329460 | 0.5 | 0.3643570 | 0.65182000 | 0.011186882682 | 0.03014923 |
| theta_ROI52_AECnodestrength | Blocktapping | 0.3382819 | 0.6617181 | 0.5 | 0.5112176 | 0.29388628 | -0.036866139864 | 0.03762966 |
| theta_ROI52_AECnodestrength | Sum_PC | 0.2657479 | 0.7342521 | 0.5 | 0.3619300 | 0.99745314 | -0.000712299444 | 0.02929548 |
| theta_ROI53_AECnodestrength | Blocktapping | 0.2687788 | 0.7312212 | 0.5 | 0.3675753 | 0.54739574 | -0.015713947848 | 0.03018724 |
| theta_ROI53_AECnodestrength | Sum_PC | 0.2464784 | 0.7535216 | 0.5 | 0.3271020 | 0.89984042 | 0.002376277502 | 0.02736550 |
| theta_ROI54_AECnodestrength | Blocktapping | 0.3373861 | 0.6626139 | 0.5 | 0.5091744 | 0.29532133 | -0.034705828297 | 0.03538364 |
| theta_ROI54_AECnodestrength | Sum_PC | 0.2925695 | 0.7074305 | 0.5 | 0.4135665 | 0.52976635 | 0.017073722660 | 0.03045940 |
| theta_ROI55_AECnodestrength | Blocktapping | 0.4039720 | 0.5960280 | 0.5 | 0.6777736 | 0.16852706 | -0.054855092071 | 0.04474643 |
| theta_ROI55_AECnodestrength | Sum_PC | 0.2829695 | 0.7170305 | 0.5 | 0.3946408 | 0.75704438 | 0.006448402435 | 0.02829969 |
| theta_ROI56_AECnodestrength | Blocktapping | 0.3513952 | 0.6486048 | 0.5 | 0.5417709 | 0.25979406 | -0.040846287545 | 0.03886194 |
| theta_ROI56_AECnodestrength | Sum_PC | 0.2741343 | 0.7258657 | 0.5 | 0.3776654 | 0.84727944 | 0.001850098306 | 0.02858452 |
| theta_ROI57_AECnodestrength | Blocktapping | 0.3675495 | 0.6324505 | 0.5 | 0.5811514 | 0.22805877 | -0.043375806799 | 0.04038093 |
| theta_ROI57_AECnodestrength | Sum_PC | 0.2770082 | 0.7229918 | 0.5 | 0.3831415 | 0.93530445 | 0.000392497876 | 0.02818799 |
| theta_ROI58_AECnodestrength | Blocktapping | 0.3932300 | 0.6067700 | 0.5 | 0.6480708 | 0.18191223 | -0.051873825767 | 0.04327319 |
| theta_ROI58_AECnodestrength | Sum_PC | 0.2889681 | 0.7110319 | 0.5 | 0.4064067 | 0.65410294 | 0.010091435327 | 0.02935732 |
| theta_ROI59_AECnodestrength | Blocktapping | 0.2497738 | 0.7502262 | 0.5 | 0.3329314 | 0.94962041 | 0.001333497379 | 0.02711770 |
| theta_ROI59_AECnodestrength | Sum_PC | 0.2703144 | 0.7296856 | 0.5 | 0.3704532 | 0.52986602 | 0.018748433574 | 0.03187887 |
| theta_ROI6_AECnodestrength | Blocktapping | 0.3035644 | 0.6964356 | 0.5 | 0.4358830 | 0.35493810 | -0.027890549691 | 0.03338714 |
| theta_ROI6_AECnodestrength | Sum_PC | 0.2574238 | 0.7425762 | 0.5 | 0.3466632 | 0.70488016 | 0.007315280298 | 0.02888985 |
| theta_ROI60_AECnodestrength | Blocktapping | 0.2446011 | 0.7553989 | 0.5 | 0.3238039 | 0.75434071 | -0.009087870168 | 0.02760537 |
| theta_ROI60_AECnodestrength | Sum_PC | 0.2395403 | 0.7604597 | 0.5 | 0.3149941 | 0.82027019 | 0.005583773588 | 0.02782384 |

| DV | Predictor | Sum_PostModProb_Incl | Sum_PostModProb_Excl | Sum_PriorModProb_Incl | BF_Incl | Frequentist_p_value | postMean | postStd |
|---|---|---|---|---|---|---|---|---|
| theta_ROI61_AECnodestrength | Blocktapping | 0.2457875 | 0.7542125 | 0.5 | 0.3258862 | 0.77009146 | 0.008035764347 | 0.02770395 |
| theta_ROI61_AECnodestrength | Sum_PC | 0.2667222 | 0.7332778 | 0.5 | 0.3637396 | 0.51728565 | 0.018477352713 | 0.03143069 |
| theta_ROI62_AECnodestrength | Blocktapping | 0.2514685 | 0.7485315 | 0.5 | 0.3359492 | 0.84443498 | 0.004751420179 | 0.02600586 |
| theta_ROI62_AECnodestrength | Sum_PC | 0.3162096 | 0.6837904 | 0.5 | 0.4624365 | 0.29334866 | 0.035463494494 | 0.03752388 |
| theta_ROI63_AECnodestrength | Blocktapping | 0.2547446 | 0.7452554 | 0.5 | 0.3418219 | 0.56739709 | -0.013109694504 | 0.02799843 |
| theta_ROI63_AECnodestrength | Sum_PC | 0.2673906 | 0.7326094 | 0.5 | 0.3649839 | 0.46107048 | 0.020255233660 | 0.03059165 |
| theta_ROI64_AECnodestrength | Blocktapping | 0.3963117 | 0.6036883 | 0.5 | 0.6564840 | 0.17441670 | -0.052040092079 | 0.04263910 |
| theta_ROI64_AECnodestrength | Sum_PC | 0.3021326 | 0.6978674 | 0.5 | 0.4329369 | 0.53722184 | 0.015253591943 | 0.03016894 |
| theta_ROI65_AECnodestrength | Blocktapping | 0.4140369 | 0.5859631 | 0.5 | 0.7065922 | 0.16455976 | -0.052879113348 | 0.04279073 |
| theta_ROI65_AECnodestrength | Sum_PC | 0.3447305 | 0.6552695 | 0.5 | 0.5260897 | 0.34899391 | 0.026367116936 | 0.03420816 |
| theta_ROI66_AECnodestrength | Blocktapping | 0.4263339 | 0.5736661 | 0.5 | 0.7431743 | 0.15586859 | -0.058002573792 | 0.04644702 |
| theta_ROI66_AECnodestrength | Sum_PC | 0.3164273 | 0.6835727 | 0.5 | 0.4629022 | 0.50024133 | 0.017251537600 | 0.03077417 |
| theta_ROI67_AECnodestrength | Blocktapping | 0.3161211 | 0.6838789 | 0.5 | 0.4622473 | 0.34610207 | -0.030188185175 | 0.03600913 |
| theta_ROI67_AECnodestrength | Sum_PC | 0.2615100 | 0.7384900 | 0.5 | 0.3541144 | 0.83923605 | 0.005670847621 | 0.02800757 |
| theta_ROI68_AECnodestrength | Blocktapping | 0.2594774 | 0.7405226 | 0.5 | 0.3503977 | 0.55295790 | -0.015368732689 | 0.02977198 |
| theta_ROI68_AECnodestrength | Sum_PC | 0.2541680 | 0.7458320 | 0.5 | 0.3407845 | 0.60550403 | 0.012736809686 | 0.03018227 |
| theta_ROI69_AECnodestrength | Blocktapping | 0.4945609 | 0.5054391 | 0.5 | 0.9784776 | 0.11100506 | -0.065256841986 | 0.04831363 |
| theta_ROI69_AECnodestrength | Sum_PC | 0.4224213 | 0.5775787 | 0.5 | 0.7313659 | 0.22344336 | 0.042077713243 | 0.04074160 |
| theta_ROI7_AECnodestrength | Blocktapping | 0.3284934 | 0.6715066 | 0.5 | 0.4891886 | 0.30181403 | -0.035477553347 | 0.03757709 |
| theta_ROI7_AECnodestrength | Sum_PC | 0.2608913 | 0.7391087 | 0.5 | 0.3529810 | 0.99968939 | -0.000470544994 | 0.02676959 |
| theta_ROI70_AECnodestrength | Blocktapping | 0.4161672 | 0.5838328 | 0.5 | 0.7128191 | 0.17856578 | -0.049524943278 | 0.04216453 |
| theta_ROI70_AECnodestrength | Sum_PC | 0.3710233 | 0.6289767 | 0.5 | 0.5898840 | 0.29188108 | 0.033766970714 | 0.03780188 |
| theta_ROI71_AECnodestrength | Blocktapping | 0.4268746 | 0.5731254 | 0.5 | 0.7448189 | 0.16325810 | -0.054348618386 | 0.04312123 |
| theta_ROI71_AECnodestrength | Sum_PC | 0.3654874 | 0.6345126 | 0.5 | 0.5760129 | 0.30250527 | 0.030654245505 | 0.03566741 |
| theta_ROI72_AECnodestrength | Blocktapping | 0.2690075 | 0.7309925 | 0.5 | 0.3680031 | 0.57957153 | -0.013347083651 | 0.02839910 |
| theta_ROI72_AECnodestrength | Sum_PC | 0.3197952 | 0.6802048 | 0.5 | 0.4701454 | 0.28626882 | 0.035597638441 | 0.03601392 |
| theta_ROI73_AECnodestrength | Blocktapping | 0.2888326 | 0.7111674 | 0.5 | 0.4061387 | 0.46141040 | -0.020194972626 | 0.03064994 |
| theta_ROI73_AECnodestrength | Sum_PC | 0.2932066 | 0.7067934 | 0.5 | 0.4148407 | 0.42069113 | 0.022644514968 | 0.03249805 |
| theta_ROI74_AECnodestrength | Blocktapping | 0.2500601 | 0.7499399 | 0.5 | 0.3334401 | 0.73049070 | -0.009863743828 | 0.02742689 |

| DV | Predictor | Sum_PostModProb_Incl | Sum_PostModProb_Excl | Sum_PriorModProb_Incl | BF_Incl | Frequentist_p_value | postMean | postStd |
|---|---|---|---|---|---|---|---|---|
| theta_ROI74_AEC nodestrength | Sum_PC | 0.2511993 | 0.7488007 | 0.5 | 0.3354688 | 0.68585334 | 0.009113729405 | 0.02890960 |
| theta_ROI75_AEC nodestrength | Blocktapping | 0.2672745 | 0.7327255 | 0.5 | 0.3647675 | 0.66023750 | -0.011264792379 | 0.02883608 |
| theta_ROI75_AEC nodestrength | Sum_PC | 0.2709076 | 0.7290924 | 0.5 | 0.3715682 | 0.52752887 | 0.017372498068 | 0.03006964 |
| theta_ROI76_AEC nodestrength | Blocktapping | 0.2460640 | 0.7539360 | 0.5 | 0.3263726 | 0.65566973 | -0.010543495771 | 0.02801798 |
| theta_ROI76_AEC nodestrength | Sum_PC | 0.2486117 | 0.7513883 | 0.5 | 0.3308698 | 0.60296331 | 0.012421322485 | 0.02980346 |
| theta_ROI77_AEC nodestrength | Blocktapping | 0.3591363 | 0.6408637 | 0.5 | 0.5603942 | 0.23448347 | -0.040074979324 | 0.03989897 |
| theta_ROI77_AEC nodestrength | Sum_PC | 0.2812117 | 0.7187883 | 0.5 | 0.3912303 | 0.67211324 | 0.009113916226 | 0.02828067 |
| theta_ROI78_AEC nodestrength | Blocktapping | 0.3066114 | 0.6933886 | 0.5 | 0.4421928 | 0.36187342 | -0.027304921037 | 0.03344938 |
| theta_ROI78_AEC nodestrength | Sum_PC | 0.2909506 | 0.7090494 | 0.5 | 0.4103390 | 0.44561924 | 0.019942680408 | 0.03234275 |
| theta_ROI79_AEC nodestrength | Blocktapping | 0.4370251 | 0.5629749 | 0.5 | 0.7762783 | 0.13408698 | -0.061491078530 | 0.04652313 |
| theta_ROI79_AEC nodestrength | Sum_PC | 0.3132078 | 0.6867922 | 0.5 | 0.4560445 | 0.51412217 | 0.016458294175 | 0.03029393 |
| theta_ROI8_AECn odestrength | Blocktapping | 0.3789184 | 0.6210816 | 0.5 | 0.6100945 | 0.19922789 | -0.047152588994 | 0.04179202 |
| theta_ROI8_AECn odestrength | Sum_PC | 0.2918325 | 0.7081675 | 0.5 | 0.4120953 | 0.64768922 | 0.011776330232 | 0.03026418 |
| theta_ROI80_AEC nodestrength | Blocktapping | 0.2726711 | 0.7273289 | 0.5 | 0.3748938 | 0.51365304 | -0.016919795301 | 0.03044485 |
| theta_ROI80_AEC nodestrength | Sum_PC | 0.2578324 | 0.7421676 | 0.5 | 0.3474046 | 0.66117442 | 0.009965296580 | 0.02964453 |
| theta_ROI81_AEC nodestrength | Blocktapping | 0.2958514 | 0.7041486 | 0.5 | 0.4201548 | 0.29339990 | -0.031643900957 | 0.03230936 |
| theta_ROI81_AEC nodestrength | Sum_PC | 0.2571386 | 0.7428614 | 0.5 | 0.3461462 | 0.49821508 | 0.015870073837 | 0.02864708 |
| theta_ROI82_AEC nodestrength | Blocktapping | 0.3887142 | 0.6112858 | 0.5 | 0.6358960 | 0.19601039 | -0.049683407409 | 0.04263150 |
| theta_ROI82_AEC nodestrength | Sum_PC | 0.2952422 | 0.7047578 | 0.5 | 0.4189272 | 0.62992167 | 0.012142860551 | 0.02991463 |
| theta_ROI83_AEC nodestrength | Blocktapping | 0.3300539 | 0.6699461 | 0.5 | 0.4926575 | 0.37793029 | -0.024120186659 | 0.03248093 |
| theta_ROI83_AEC nodestrength | Sum_PC | 0.3807463 | 0.6192537 | 0.5 | 0.6148470 | 0.20769962 | 0.044983519653 | 0.04116115 |
| theta_ROI84_AEC nodestrength | Blocktapping | 0.3429981 | 0.6570019 | 0.5 | 0.5220656 | 0.38549489 | -0.024516395991 | 0.03110904 |
| theta_ROI84_AEC nodestrength | Sum_PC | 0.4499970 | 0.5500030 | 0.5 | 0.8181720 | 0.11720217 | 0.066519586776 | 0.04830783 |
| theta_ROI85_AEC nodestrength | Blocktapping | 0.3526948 | 0.6473052 | 0.5 | 0.5448663 | 0.23189346 | -0.040635572563 | 0.03684191 |
| theta_ROI85_AEC nodestrength | Sum_PC | 0.3051284 | 0.6948716 | 0.5 | 0.4391147 | 0.40310503 | 0.023200521980 | 0.03243507 |
| theta_ROI86_AEC nodestrength | Blocktapping | 0.2705408 | 0.7294592 | 0.5 | 0.3708787 | 0.47959566 | -0.018331694813 | 0.02905131 |
| theta_ROI86_AEC nodestrength | Sum_PC | 0.2712788 | 0.7287212 | 0.5 | 0.3722669 | 0.46389325 | 0.020168361694 | 0.03069379 |
| theta_ROI87_AEC nodestrength | Blocktapping | 0.3009353 | 0.6990647 | 0.5 | 0.4304828 | 0.38260269 | -0.025895138304 | 0.03388321 |
| theta_ROI87_AEC nodestrength | Sum_PC | 0.2631379 | 0.7368621 | 0.5 | 0.3571061 | 0.70279962 | 0.008198651627 | 0.02959097 |

| DV | Predictor | Sum_PostModProb_Incl | Sum_PostModProb_Excl | Sum_PriorModProb_Incl | BF_Incl | Frequentist_p_value | postMean | postStd |
|---|---|---|---|---|---|---|---|---|
| theta_ROI88_AECnodestrength | Blocktapping | 0.2869827 | 0.7130173 | 0.5 | 0.4024906 | 0.43020373 | -0.021062146562 | 0.03053766 |
| theta_ROI88_AECnodestrength | Sum_PC | 0.2930605 | 0.7069395 | 0.5 | 0.4145483 | 0.38329605 | 0.025010401508 | 0.03225937 |
| theta_ROI89_AECnodestrength | Blocktapping | 0.3036138 | 0.6963862 | 0.5 | 0.4359847 | 0.37916922 | -0.025627501227 | 0.03351595 |
| theta_ROI89_AECnodestrength | Sum_PC | 0.2706194 | 0.7293806 | 0.5 | 0.3710262 | 0.61557101 | 0.012734715751 | 0.03033742 |
| theta_ROI9_AECnodestrength | Blocktapping | 0.4324825 | 0.5675175 | 0.5 | 0.7620601 | 0.14037479 | -0.061930188770 | 0.04653952 |
| theta_ROI9_AECnodestrength | Sum_PC | 0.3061962 | 0.6938038 | 0.5 | 0.4413297 | 0.55823936 | 0.014930415024 | 0.03029307 |
| theta_ROI90_AECnodestrength | Blocktapping | 0.2827778 | 0.7172222 | 0.5 | 0.3942680 | 0.50608327 | -0.018018082657 | 0.03078360 |
| theta_ROI90_AECnodestrength | Sum_PC | 0.2770424 | 0.7229576 | 0.5 | 0.3832070 | 0.53599478 | 0.016629146613 | 0.03090293 |
| theta_ROI91_AECnodestrength | Blocktapping | 0.2477464 | 0.7522536 | 0.5 | 0.3293390 | 0.98188857 | 0.000494275524 | 0.02708320 |
| theta_ROI91_AECnodestrength | Sum_PC | 0.2702585 | 0.7297415 | 0.5 | 0.3703482 | 0.55503739 | 0.018065045238 | 0.03207397 |
| theta_ROI92_AECnodestrength | Blocktapping | 0.2460132 | 0.7539868 | 0.5 | 0.3262832 | 0.98080296 | -0.001416094377 | 0.02703915 |
| theta_ROI92_AECnodestrength | Sum_PC | 0.2665761 | 0.7334239 | 0.5 | 0.3634681 | 0.51735090 | 0.019174672915 | 0.03132691 |
| theta_ROI93_AECnodestrength | Blocktapping | 0.2525567 | 0.7474433 | 0.5 | 0.3378942 | 0.86272281 | 0.004310495897 | 0.02707474 |
| theta_ROI93_AECnodestrength | Sum_PC | 0.2694710 | 0.7305290 | 0.5 | 0.3688710 | 0.57391844 | 0.015720458618 | 0.03173622 |
| theta_ROI94_AECnodestrength | Blocktapping | 0.3786923 | 0.6213077 | 0.5 | 0.6095085 | 0.19303629 | -0.047374632716 | 0.04182718 |
| theta_ROI94_AECnodestrength | Sum_PC | 0.2781557 | 0.7218443 | 0.5 | 0.3853403 | 0.69885478 | 0.007531553950 | 0.02765661 |
| theta_ROI95_AECnodestrength | Blocktapping | 0.2928059 | 0.7071941 | 0.5 | 0.4140389 | 0.46885198 | -0.018804720671 | 0.02958113 |
| theta_ROI95_AECnodestrength | Sum_PC | 0.3456639 | 0.6543361 | 0.5 | 0.5282665 | 0.23697242 | 0.041068405147 | 0.03761892 |
| theta_ROI96_AECnodestrength | Blocktapping | 0.4610706 | 0.5389294 | 0.5 | 0.8555305 | 0.12356531 | -0.065146506511 | 0.04928980 |
| theta_ROI96_AECnodestrength | Sum_PC | 0.3615775 | 0.6384225 | 0.5 | 0.5663607 | 0.33944220 | 0.028108264763 | 0.03351265 |
| theta_ROI97_AECnodestrength | Blocktapping | 0.3223579 | 0.6776421 | 0.5 | 0.4757052 | 0.29179103 | -0.032002496206 | 0.03461973 |
| theta_ROI97_AECnodestrength | Sum_PC | 0.2897007 | 0.7102993 | 0.5 | 0.4078573 | 0.42369341 | 0.021514715639 | 0.03043609 |
| theta_ROI98_AECnodestrength | Blocktapping | 0.2731367 | 0.7268633 | 0.5 | 0.3757745 | 0.42454102 | -0.022253941550 | 0.03138987 |
| theta_ROI98_AECnodestrength | Sum_PC | 0.2469149 | 0.7530851 | 0.5 | 0.3278712 | 0.66420822 | 0.010472230010 | 0.02876494 |
| theta_ROI99_AECnodestrength | Blocktapping | 0.3044452 | 0.6955548 | 0.5 | 0.4377013 | 0.40140477 | -0.026583545121 | 0.03436798 |
| theta_ROI99_AECnodestrength | Sum_PC | 0.2676163 | 0.7323837 | 0.5 | 0.3654045 | 0.75382230 | 0.007208129146 | 0.02975154 |
| ThetaPow1 | Blocktapping | 0.2756534 | 0.7243466 | 0.5 | 0.3805545 | 0.42375041 | 0.023399304316 | 0.03084400 |
| ThetaPow1 | Sum_PC | 0.2404342 | 0.7595658 | 0.5 | 0.3165417 | 0.90690802 | 0.003579589764 | 0.02709935 |
| ThetaPow10 | Blocktapping | 0.2470867 | 0.7529133 | 0.5 | 0.3281742 | 0.64143119 | 0.010037636434 | 0.02826976 |
| ThetaPow10 | Sum_PC | 0.2324563 | 0.7675437 | 0.5 | 0.3028574 | 0.94999841 | -0.001140568957 | 0.02652968 |

| DV | Predictor | Sum_PostModProb_Incl | Sum_PostModProb_Excl | Sum_PriorModProb_Incl | BF_Incl | Frequentist_p_value | postMean | postStd |
|---|---|---|---|---|---|---|---|---|
| ThetaPow100 | Blocktapping | 0.2621373 | 0.7378627 | 0.5 | 0.3552657 | 0.56715166 | 0.014572100377 | 0.02909316 |
| ThetaPow100 | Sum_PC | 0.2519349 | 0.7480651 | 0.5 | 0.3367820 | 0.66312737 | -0.010489503960 | 0.02886577 |
| ThetaPow11 | Blocktapping | 0.2554820 | 0.7445180 | 0.5 | 0.3431510 | 0.59835408 | 0.013901943807 | 0.02871832 |
| ThetaPow11 | Sum_PC | 0.2425476 | 0.7574524 | 0.5 | 0.3202150 | 0.76579368 | 0.007601766117 | 0.02819556 |
| ThetaPow12 | Blocktapping | 0.2597758 | 0.7402242 | 0.5 | 0.3509421 | 0.56822981 | 0.014344511911 | 0.02804202 |
| ThetaPow12 | Sum_PC | 0.2437529 | 0.7562471 | 0.5 | 0.3223192 | 0.80688549 | -0.005024620560 | 0.02816933 |
| ThetaPow13 | Blocktapping | 0.2669583 | 0.7330417 | 0.5 | 0.3641789 | 0.51660482 | 0.017559868943 | 0.02913919 |
| ThetaPow13 | Sum_PC | 0.2508437 | 0.7491563 | 0.5 | 0.3348350 | 0.73092315 | -0.007370818184 | 0.02869751 |
| ThetaPow14 | Blocktapping | 0.2596231 | 0.7403769 | 0.5 | 0.3506635 | 0.64627065 | 0.012173360208 | 0.02937522 |
| ThetaPow14 | Sum_PC | 0.2582263 | 0.7417737 | 0.5 | 0.3481200 | 0.64625211 | 0.013537560957 | 0.03026080 |
| ThetaPow15 | Blocktapping | 0.2500439 | 0.7499561 | 0.5 | 0.3334113 | 0.82269784 | 0.005981382492 | 0.02772684 |
| ThetaPow15 | Sum_PC | 0.2637308 | 0.7362692 | 0.5 | 0.3581989 | 0.59033328 | 0.015538880509 | 0.03039264 |
| ThetaPow16 | Blocktapping | 0.2852629 | 0.7147371 | 0.5 | 0.3991158 | 0.38625994 | 0.024326265303 | 0.03259952 |
| ThetaPow16 | Sum_PC | 0.2445964 | 0.7554036 | 0.5 | 0.3237957 | 0.94221738 | -0.000673602666 | 0.02741813 |
| ThetaPow17 | Blocktapping | 0.2471184 | 0.7528816 | 0.5 | 0.3282301 | 0.76660052 | 0.006364211193 | 0.02807897 |
| ThetaPow17 | Sum_PC | 0.2451598 | 0.7548402 | 0.5 | 0.3247836 | 0.75474880 | -0.007413980649 | 0.02857057 |
| ThetaPow18 | Blocktapping | 0.3302118 | 0.6697882 | 0.5 | 0.4930092 | 0.29404493 | 0.033675045102 | 0.03628709 |
| ThetaPow18 | Sum_PC | 0.2705950 | 0.7294050 | 0.5 | 0.3709804 | 0.76303600 | -0.006097927591 | 0.02888144 |
| ThetaPow19 | Blocktapping | 0.2537983 | 0.7462017 | 0.5 | 0.3401203 | 0.65902206 | 0.011843109192 | 0.02845605 |
| ThetaPow19 | Sum_PC | 0.2481149 | 0.7518851 | 0.5 | 0.3299905 | 0.77699727 | 0.007895457038 | 0.02901274 |
| ThetaPow2 | Blocktapping | 0.3090158 | 0.6909842 | 0.5 | 0.4472111 | 0.31850008 | 0.030453950587 | 0.03381199 |
| ThetaPow2 | Sum_PC | 0.2542177 | 0.7457823 | 0.5 | 0.3408738 | 0.84706146 | -0.004429152354 | 0.02798878 |
| ThetaPow20 | Blocktapping | 0.2696101 | 0.7303899 | 0.5 | 0.3691318 | 0.51536282 | 0.018621499475 | 0.02916618 |
| ThetaPow20 | Sum_PC | 0.2483376 | 0.7516624 | 0.5 | 0.3303845 | 0.79693425 | -0.005581449166 | 0.02845014 |
| ThetaPow21 | Blocktapping | 0.2647564 | 0.7352436 | 0.5 | 0.3600934 | 0.52730810 | 0.016336385336 | 0.02917598 |
| ThetaPow21 | Sum_PC | 0.2445978 | 0.7554022 | 0.5 | 0.3237982 | 0.88493948 | 0.004865966104 | 0.02893483 |
| ThetaPow22 | Blocktapping | 0.2581954 | 0.7418046 | 0.5 | 0.3480639 | 0.56549912 | 0.015241861805 | 0.02916105 |
| ThetaPow22 | Sum_PC | 0.2427475 | 0.7572525 | 0.5 | 0.3205635 | 0.82327422 | 0.007277065728 | 0.02754566 |
| ThetaPow23 | Blocktapping | 0.2645903 | 0.7354097 | 0.5 | 0.3597862 | 0.61704449 | 0.014062381598 | 0.02857733 |
| ThetaPow23 | Sum_PC | 0.2676188 | 0.7323812 | 0.5 | 0.3654092 | 0.59584463 | 0.015262255379 | 0.03053004 |
| ThetaPow24 | Blocktapping | 0.2450783 | 0.7549217 | 0.5 | 0.3246407 | 0.74556265 | 0.007962782109 | 0.02784567 |
| ThetaPow24 | Sum_PC | 0.2455419 | 0.7544581 | 0.5 | 0.3254546 | 0.73779215 | -0.009054174682 | 0.02982837 |
| ThetaPow25 | Blocktapping | 0.2641977 | 0.7358023 | 0.5 | 0.3590607 | 0.46149410 | 0.020143451703 | 0.03031335 |
| ThetaPow25 | Sum_PC | 0.2324396 | 0.7675604 | 0.5 | 0.3028290 | 0.98339886 | 0.001266475991 | 0.02649206 |
| ThetaPow26 | Blocktapping | 0.2472457 | 0.7527543 | 0.5 | 0.3284547 | 0.64486627 | 0.012739028882 | 0.02809408 |
| ThetaPow26 | Sum_PC | 0.2369667 | 0.7630333 | 0.5 | 0.3105587 | 0.96024871 | -0.000171886411 | 0.02718817 |
| ThetaPow27 | Blocktapping | 0.2387374 | 0.7612626 | 0.5 | 0.3136071 | 0.85841195 | -0.004357214419 | 0.02624191 |
| ThetaPow27 | Sum_PC | 0.2384866 | 0.7615134 | 0.5 | 0.3131745 | 0.97963829 | -0.000518334901 | 0.02858231 |
| ThetaPow28 | Blocktapping | 0.2367392 | 0.7632608 | 0.5 | 0.3101681 | 0.93794986 | 0.002184668456 | 0.02600584 |
| ThetaPow28 | Sum_PC | 0.2475884 | 0.7524116 | 0.5 | 0.3290598 | 0.67219067 | 0.012164830688 | 0.02844881 |
| ThetaPow29 | Blocktapping | 0.2597475 | 0.7402525 | 0.5 | 0.3508905 | 0.58932650 | 0.014621143400 | 0.02871073 |

| DV | Predictor | Sum_PostModProb_Incl | Sum_PostModProb_Excl | Sum_PriorModProb_Incl | BF_Incl | Frequentist_p_value | postMean | postStd |
|---|---|---|---|---|---|---|---|---|
| ThetaPow29 | Sum_PC | 0.2464944 | 0.7535056 | 0.5 | 0.3271302 | 0.83666930 | 0.006609836099 | 0.02870209 |
| ThetaPow3 | Blocktapping | 0.2860377 | 0.7139623 | 0.5 | 0.4006341 | 0.37250959 | 0.026184775941 | 0.03169349 |
| ThetaPow3 | Sum_PC | 0.2429013 | 0.7570987 | 0.5 | 0.3208317 | 0.88792089 | -0.002545578675 | 0.02780683 |
| ThetaPow30 | Blocktapping | 0.2662211 | 0.7337789 | 0.5 | 0.3628084 | 0.56866134 | 0.014986886168 | 0.02895863 |
| ThetaPow30 | Sum_PC | 0.2572979 | 0.7427021 | 0.5 | 0.3464348 | 0.71579605 | 0.010193533081 | 0.03024433 |
| ThetaPow31 | Blocktapping | 0.2383434 | 0.7616566 | 0.5 | 0.3129276 | 0.99223067 | -0.001062264934 | 0.02648983 |
| ThetaPow31 | Sum_PC | 0.2392659 | 0.7607341 | 0.5 | 0.3145197 | 0.82169038 | -0.004763041788 | 0.02748253 |
| ThetaPow32 | Blocktapping | 0.2770264 | 0.7229736 | 0.5 | 0.3831763 | 0.44238620 | 0.021283098478 | 0.03271090 |
| ThetaPow32 | Sum_PC | 0.2454427 | 0.7545573 | 0.5 | 0.3252804 | 0.90540601 | -0.002184052848 | 0.02771138 |
| ThetaPow33 | Blocktapping | 0.2770058 | 0.7229942 | 0.5 | 0.3831369 | 0.45543735 | 0.020758966008 | 0.03154467 |
| ThetaPow33 | Sum_PC | 0.2428457 | 0.7571543 | 0.5 | 0.3207348 | 0.86366051 | -0.003192516665 | 0.02663312 |
| ThetaPow34 | Blocktapping | 0.2911450 | 0.7088550 | 0.5 | 0.4107258 | 0.42495865 | 0.022670814951 | 0.03303311 |
| ThetaPow34 | Sum_PC | 0.2557470 | 0.7442530 | 0.5 | 0.3436290 | 0.80902868 | 0.007983374368 | 0.02822520 |
| ThetaPow35 | Blocktapping | 0.2339811 | 0.7660189 | 0.5 | 0.3054508 | 0.95917733 | -0.001286913494 | 0.02700297 |
| ThetaPow35 | Sum_PC | 0.2351938 | 0.7648062 | 0.5 | 0.3075207 | 0.82016576 | 0.006106956252 | 0.02787231 |
| ThetaPow36 | Blocktapping | 0.3054719 | 0.6945281 | 0.5 | 0.4398265 | 0.33626426 | 0.027829989786 | 0.03414663 |
| ThetaPow36 | Sum_PC | 0.2621265 | 0.7378735 | 0.5 | 0.3552459 | 0.66804233 | -0.009773698563 | 0.02843776 |
| ThetaPow37 | Blocktapping | 0.2547618 | 0.7452382 | 0.5 | 0.3418528 | 0.64234216 | 0.013490596451 | 0.02890936 |
| ThetaPow37 | Sum_PC | 0.2485591 | 0.7514409 | 0.5 | 0.3307767 | 0.79227905 | 0.006955276208 | 0.02932143 |
| ThetaPow38 | Blocktapping | 0.2830996 | 0.7169004 | 0.5 | 0.3948940 | 0.41907830 | 0.022749239487 | 0.03159717 |
| ThetaPow38 | Sum_PC | 0.2476083 | 0.7523917 | 0.5 | 0.3290950 | 0.81739575 | -0.003771578858 | 0.02724687 |
| ThetaPow39 | Blocktapping | 0.2472835 | 0.7527165 | 0.5 | 0.3285214 | 0.60866064 | 0.013088839821 | 0.02821711 |
| ThetaPow39 | Sum_PC | 0.2359169 | 0.7640831 | 0.5 | 0.3087583 | 0.89375681 | -0.001585554214 | 0.02751209 |
| ThetaPow4 | Blocktapping | 0.3952271 | 0.6047729 | 0.5 | 0.6535132 | 0.16751770 | 0.053486202184 | 0.04184645 |
| ThetaPow4 | Sum_PC | 0.2800441 | 0.7199559 | 0.5 | 0.3889739 | 0.86043081 | -0.002704384964 | 0.02741048 |
| ThetaPow40 | Blocktapping | 0.2726274 | 0.7273726 | 0.5 | 0.3748113 | 0.47744971 | 0.019011637965 | 0.03006964 |
| ThetaPow40 | Sum_PC | 0.2446830 | 0.7553170 | 0.5 | 0.3239474 | 0.84992681 | -0.003305732860 | 0.02713693 |
| ThetaPow41 | Blocktapping | 0.2937228 | 0.7062772 | 0.5 | 0.4158746 | 0.38536657 | 0.025350403101 | 0.03297858 |
| ThetaPow41 | Sum_PC | 0.2505238 | 0.7494762 | 0.5 | 0.3342652 | 0.84415711 | 0.006399164158 | 0.02853600 |
| ThetaPow42 | Blocktapping | 0.2617477 | 0.7382523 | 0.5 | 0.3545504 | 0.65412465 | 0.011472683313 | 0.02970128 |
| ThetaPow42 | Sum_PC | 0.2436466 | 0.7563534 | 0.5 | 0.3221333 | 0.71085553 | -0.008669651910 | 0.02781503 |
| ThetaPow43 | Blocktapping | 0.2455167 | 0.7544833 | 0.5 | 0.3254104 | 0.69129238 | 0.008317605281 | 0.02624171 |
| ThetaPow43 | Sum_PC | 0.2437086 | 0.7562914 | 0.5 | 0.3222416 | 0.73705777 | -0.008782251913 | 0.02756502 |
| ThetaPow44 | Blocktapping | 0.2415747 | 0.7584253 | 0.5 | 0.3185214 | 0.84819618 | 0.002559706935 | 0.02590282 |
| ThetaPow44 | Sum_PC | 0.2768547 | 0.7231453 | 0.5 | 0.3828480 | 0.44599120 | -0.020966835519 | 0.03287570 |
| ThetaPow45 | Blocktapping | 0.2264438 | 0.7735562 | 0.5 | 0.2927308 | 0.83022798 | 0.002997980967 | 0.02559130 |
| ThetaPow45 | Sum_PC | 0.2382883 | 0.7617117 | 0.5 | 0.3128326 | 0.65246116 | -0.010436573393 | 0.02869027 |
| ThetaPow46 | Blocktapping | 0.2427592 | 0.7572408 | 0.5 | 0.3205839 | 0.72093682 | -0.008173503736 | 0.02689948 |
| ThetaPow46 | Sum_PC | 0.2373350 | 0.7626650 | 0.5 | 0.3111917 | 0.83883253 | -0.005944409439 | 0.02719931 |
| ThetaPow47 | Blocktapping | 0.2534813 | 0.7465187 | 0.5 | 0.3395511 | 0.62370621 | 0.012855409924 | 0.02917771 |
| ThetaPow47 | Sum_PC | 0.2464259 | 0.7535741 | 0.5 | 0.3270096 | 0.77318755 | 0.008931553164 | 0.02873657 |

| DV | Predictor | Sum_PostModProb_Incl | Sum_PostModProb_Excl | Sum_PriorModProb_Incl | BF_Incl | Frequentist_p_value | postMean | postStd |
|---|---|---|---|---|---|---|---|---|
| ThetaPow48 | Blocktapping | 0.2398974 | 0.7601026 | 0.5 | 0.3156118 | 0.87143566 | 0.003739056044 | 0.02725287 |
| ThetaPow48 | Sum_PC | 0.2440257 | 0.7559743 | 0.5 | 0.3227962 | 0.77767906 | 0.007207971910 | 0.02928282 |
| ThetaPow49 | Blocktapping | 0.2833622 | 0.7166378 | 0.5 | 0.3954051 | 0.40618798 | 0.023335377453 | 0.03279683 |
| ThetaPow49 | Sum_PC | 0.2441044 | 0.7558956 | 0.5 | 0.3229341 | 0.82692841 | -0.003853351407 | 0.02750671 |
| ThetaPow5 | Blocktapping | 0.3630599 | 0.6369401 | 0.5 | 0.5700063 | 0.18884518 | 0.045416344845 | 0.04013640 |
| ThetaPow5 | Sum_PC | 0.2618585 | 0.7381415 | 0.5 | 0.3547538 | 0.84749530 | -0.003835032399 | 0.02664361 |
| ThetaPow50 | Blocktapping | 0.2830455 | 0.7169545 | 0.5 | 0.3947887 | 0.44623951 | 0.020947926837 | 0.03211268 |
| ThetaPow50 | Sum_PC | 0.2525501 | 0.7474499 | 0.5 | 0.3378823 | 0.75997099 | -0.006554474581 | 0.02855883 |
| ThetaPow51 | Blocktapping | 0.2279457 | 0.7720543 | 0.5 | 0.2952457 | 0.96541270 | -0.000028780985 | 0.02589661 |
| ThetaPow51 | Sum_PC | 0.2264917 | 0.7735083 | 0.5 | 0.2928109 | 0.90422100 | 0.003302372600 | 0.02794407 |
| ThetaPow52 | Blocktapping | 0.2395586 | 0.7604414 | 0.5 | 0.3150257 | 0.68768229 | 0.010442011864 | 0.02688604 |
| ThetaPow52 | Sum_PC | 0.2351018 | 0.7648982 | 0.5 | 0.3073635 | 0.94341381 | 0.003268426095 | 0.02701207 |
| ThetaPow53 | Blocktapping | 0.2437817 | 0.7562183 | 0.5 | 0.3223695 | 0.86735687 | -0.003849766878 | 0.02719963 |
| ThetaPow53 | Sum_PC | 0.2582582 | 0.7417418 | 0.5 | 0.3481780 | 0.56213410 | 0.016026069475 | 0.03030340 |
| ThetaPow54 | Blocktapping | 0.3104414 | 0.6895586 | 0.5 | 0.4502031 | 0.31209305 | 0.029171052192 | 0.03417904 |
| ThetaPow54 | Sum_PC | 0.2550016 | 0.7449984 | 0.5 | 0.3422847 | 0.84064970 | -0.003535179389 | 0.02681314 |
| ThetaPow55 | Blocktapping | 0.2857540 | 0.7142460 | 0.5 | 0.4000778 | 0.38855220 | 0.024416001260 | 0.03173542 |
| ThetaPow55 | Sum_PC | 0.2526116 | 0.7473884 | 0.5 | 0.3379925 | 0.77713575 | -0.006769750074 | 0.02853966 |
| ThetaPow56 | Blocktapping | 0.2456527 | 0.7543473 | 0.5 | 0.3256494 | 0.62008275 | 0.011906646994 | 0.02817165 |
| ThetaPow56 | Sum_PC | 0.2363279 | 0.7636721 | 0.5 | 0.3094625 | 0.81880557 | -0.006319631068 | 0.02763241 |
| ThetaPow57 | Blocktapping | 0.2582634 | 0.7417366 | 0.5 | 0.3481875 | 0.54808738 | 0.014463783152 | 0.02832828 |
| ThetaPow57 | Sum_PC | 0.2438051 | 0.7561949 | 0.5 | 0.3224104 | 0.85368793 | -0.004546780778 | 0.02798262 |
| ThetaPow58 | Blocktapping | 0.3449052 | 0.6550948 | 0.5 | 0.5264965 | 0.23514217 | 0.038339608963 | 0.03656660 |
| ThetaPow58 | Sum_PC | 0.2728483 | 0.7271517 | 0.5 | 0.3752288 | 0.67733774 | -0.009519054952 | 0.02855321 |
| ThetaPow59 | Blocktapping | 0.2368455 | 0.7631545 | 0.5 | 0.3103506 | 0.89006777 | -0.004832292424 | 0.02589155 |
| ThetaPow59 | Sum_PC | 0.2496823 | 0.7503177 | 0.5 | 0.3327688 | 0.61239710 | -0.014344943782 | 0.02928732 |
| ThetaPow6 | Blocktapping | 0.2912925 | 0.7087075 | 0.5 | 0.4110193 | 0.36399297 | 0.025758357705 | 0.03282567 |
| ThetaPow6 | Sum_PC | 0.2473413 | 0.7526587 | 0.5 | 0.3286235 | 0.86151508 | -0.003006223312 | 0.02710935 |
| ThetaPow60 | Blocktapping | 0.2302859 | 0.7697141 | 0.5 | 0.2991837 | 0.91942549 | -0.004922537340 | 0.02696795 |
| ThetaPow60 | Sum_PC | 0.2496180 | 0.7503820 | 0.5 | 0.3326545 | 0.80919568 | -0.007081726032 | 0.02900825 |
| ThetaPow61 | Blocktapping | 0.2488558 | 0.7511442 | 0.5 | 0.3313023 | 0.91915413 | -0.004035389592 | 0.02618541 |
| ThetaPow61 | Sum_PC | 0.2828574 | 0.7171426 | 0.5 | 0.3944228 | 0.44709300 | -0.022775292499 | 0.03333084 |
| ThetaPow62 | Blocktapping | 0.2476521 | 0.7523479 | 0.5 | 0.3291723 | 0.85861489 | 0.003365399899 | 0.02647307 |
| ThetaPow62 | Sum_PC | 0.2817095 | 0.7182905 | 0.5 | 0.3921944 | 0.41629774 | -0.023336369398 | 0.03271448 |
| ThetaPow63 | Blocktapping | 0.2348823 | 0.7651177 | 0.5 | 0.3069885 | 0.88176460 | 0.002101329607 | 0.02597511 |
| ThetaPow63 | Sum_PC | 0.2459249 | 0.7540751 | 0.5 | 0.3261278 | 0.62982804 | -0.012734565610 | 0.02897765 |
| ThetaPow64 | Blocktapping | 0.2235516 | 0.7764484 | 0.5 | 0.2879155 | 0.82971509 | 0.003833466935 | 0.02409251 |
| ThetaPow64 | Sum_PC | 0.2923201 | 0.7076799 | 0.5 | 0.4130682 | 0.62155072 | -0.015476905880 | 0.03412820 |
| ThetaPow65 | Blocktapping | 0.2389116 | 0.7610884 | 0.5 | 0.3139079 | 0.81956451 | 0.004778104754 | 0.02542929 |
| ThetaPow65 | Sum_PC | 0.2622630 | 0.7377370 | 0.5 | 0.3554966 | 0.60697364 | -0.014056274994 | 0.03153261 |
| ThetaPow66 | Blocktapping | 0.2428814 | 0.7571186 | 0.5 | 0.3207970 | 0.86515663 | 0.003995468145 | 0.02664249 |

| DV | Predictor | Sum_PostModProb_Incl | Sum_PostModProb_Excl | Sum_PriorModProb_Incl | BF_Incl | Frequentist_p_value | postMean | postStd |
|---|---|---|---|---|---|---|---|---|
| ThetaPow66 | Sum_PC | 0.2492736 | 0.7507264 | 0.5 | 0.3320432 | 0.68082570 | 0.012186286885 | 0.02994478 |
| ThetaPow67 | Blocktapping | 0.2311068 | 0.7688932 | 0.5 | 0.3005707 | 0.93383880 | -0.001659512331 | 0.02512135 |
| ThetaPow67 | Sum_PC | 0.2408611 | 0.7591389 | 0.5 | 0.3172820 | 0.69015919 | 0.011018870677 | 0.02839185 |
| ThetaPow68 | Blocktapping | 0.2389452 | 0.7610548 | 0.5 | 0.3139658 | 0.94661669 | -0.000059074593 | 0.02652345 |
| ThetaPow68 | Sum_PC | 0.2660843 | 0.7339157 | 0.5 | 0.3625544 | 0.49160864 | -0.018184768927 | 0.03134101 |
| ThetaPow69 | Blocktapping | 0.2447631 | 0.7552369 | 0.5 | 0.3240879 | 0.94165351 | -0.004097565783 | 0.02643620 |
| ThetaPow69 | Sum_PC | 0.2662616 | 0.7337384 | 0.5 | 0.3628835 | 0.51773360 | -0.019732771736 | 0.03051575 |
| ThetaPow7 | Blocktapping | 0.2993151 | 0.7006849 | 0.5 | 0.4271750 | 0.34049213 | 0.029633357512 | 0.03335899 |
| ThetaPow7 | Sum_PC | 0.2528631 | 0.7471369 | 0.5 | 0.3384428 | 0.90751395 | -0.002921367067 | 0.02804615 |
| ThetaPow70 | Blocktapping | 0.2470413 | 0.7529587 | 0.5 | 0.3280941 | 0.81784011 | 0.005779345438 | 0.02689051 |
| ThetaPow70 | Sum_PC | 0.2613064 | 0.7386936 | 0.5 | 0.3537413 | 0.61379845 | -0.012509646319 | 0.03041863 |
| ThetaPow71 | Blocktapping | 0.2399407 | 0.7600593 | 0.5 | 0.3156868 | 0.82227850 | 0.004634102102 | 0.02785567 |
| ThetaPow71 | Sum_PC | 0.2409573 | 0.7590427 | 0.5 | 0.3174490 | 0.83185784 | -0.005252649137 | 0.02848423 |
| ThetaPow72 | Blocktapping | 0.2426567 | 0.7573433 | 0.5 | 0.3204052 | 0.76669433 | 0.006970204419 | 0.02716539 |
| ThetaPow72 | Sum_PC | 0.2405159 | 0.7594841 | 0.5 | 0.3166833 | 0.77730428 | -0.006958129622 | 0.02825993 |
| ThetaPow73 | Blocktapping | 0.2379556 | 0.7620444 | 0.5 | 0.3122596 | 0.92239044 | 0.002659712129 | 0.02734635 |
| ThetaPow73 | Sum_PC | 0.2354764 | 0.7645236 | 0.5 | 0.3080041 | 0.87189076 | 0.003702545430 | 0.02783953 |
| ThetaPow74 | Blocktapping | 0.2306467 | 0.7693533 | 0.5 | 0.2997929 | 0.92201680 | -0.001650336054 | 0.02620690 |
| ThetaPow74 | Sum_PC | 0.2313489 | 0.7686511 | 0.5 | 0.3009803 | 0.84709988 | 0.004399476413 | 0.02772374 |
| ThetaPow75 | Blocktapping | 0.2405361 | 0.7594639 | 0.5 | 0.3167182 | 0.93064680 | -0.004090375908 | 0.02641254 |
| ThetaPow75 | Sum_PC | 0.2722709 | 0.7277291 | 0.5 | 0.3741378 | 0.44522024 | -0.023055733626 | 0.03186724 |
| ThetaPow76 | Blocktapping | 0.2329957 | 0.7670043 | 0.5 | 0.3037737 | 0.95117738 | 0.002171097281 | 0.02619814 |
| ThetaPow76 | Sum_PC | 0.2359092 | 0.7640908 | 0.5 | 0.3087449 | 0.79627153 | -0.006842797405 | 0.02808137 |
| ThetaPow77 | Blocktapping | 0.2424527 | 0.7575473 | 0.5 | 0.3200496 | 0.75025129 | 0.007789400588 | 0.02791371 |
| ThetaPow77 | Sum_PC | 0.2377469 | 0.7622531 | 0.5 | 0.3119002 | 0.94161441 | -0.001989220987 | 0.02703104 |
| ThetaPow78 | Blocktapping | 0.2497310 | 0.7502690 | 0.5 | 0.3328553 | 0.68616501 | 0.010929278681 | 0.02755999 |
| ThetaPow78 | Sum_PC | 0.2517847 | 0.7482153 | 0.5 | 0.3365136 | 0.72831407 | 0.009777511716 | 0.02931702 |
| ThetaPow79 | Blocktapping | 0.2357710 | 0.7642290 | 0.5 | 0.3085083 | 0.92216488 | 0.002468778933 | 0.02719149 |
| ThetaPow79 | Sum_PC | 0.2350104 | 0.7649896 | 0.5 | 0.3072074 | 0.92777156 | -0.002097376611 | 0.02784312 |
| ThetaPow8 | Blocktapping | 0.3402785 | 0.6597215 | 0.5 | 0.5157911 | 0.23365063 | 0.040132934275 | 0.03712582 |
| ThetaPow8 | Sum_PC | 0.2609242 | 0.7390758 | 0.5 | 0.3530412 | 0.81209676 | -0.004322060583 | 0.02738248 |
| ThetaPow80 | Blocktapping | 0.2299295 | 0.7700705 | 0.5 | 0.2985824 | 0.94233674 | -0.002384996083 | 0.02677772 |
| ThetaPow80 | Sum_PC | 0.2322078 | 0.7677922 | 0.5 | 0.3024357 | 0.79299989 | -0.006273177560 | 0.02733567 |
| ThetaPow81 | Blocktapping | 0.2555802 | 0.7444198 | 0.5 | 0.3433280 | 0.80359408 | -0.007717464859 | 0.02718446 |
| ThetaPow81 | Sum_PC | 0.2696034 | 0.7303966 | 0.5 | 0.3691193 | 0.54523290 | -0.017734096099 | 0.03095208 |
| ThetaPow82 | Blocktapping | 0.2390519 | 0.7609481 | 0.5 | 0.3141500 | 0.95384382 | -0.001540271179 | 0.02617456 |
| ThetaPow82 | Sum_PC | 0.2396115 | 0.7603885 | 0.5 | 0.3151173 | 0.89469392 | -0.003324642930 | 0.02783926 |
| ThetaPow83 | Blocktapping | 0.2336681 | 0.7663319 | 0.5 | 0.3049175 | 0.93310118 | 0.002471372833 | 0.02645287 |
| ThetaPow83 | Sum_PC | 0.2347202 | 0.7652798 | 0.5 | 0.3067117 | 0.85308467 | -0.004141347942 | 0.02820178 |
| ThetaPow84 | Blocktapping | 0.2512266 | 0.7487734 | 0.5 | 0.3355176 | 0.96780969 | -0.002417423145 | 0.02706252 |
| ThetaPow84 | Sum_PC | 0.2900798 | 0.7099202 | 0.5 | 0.4086090 | 0.40786816 | -0.024191648780 | 0.03495611 |

| DV | Predictor | Sum_PostModProb_Incl | Sum_PostModProb_Excl | Sum_PriorModProb_Incl | BF_Incl | Frequentist_p_value | postMean | postStd |
|---|---|---|---|---|---|---|---|---|
| ThetaPow85 | Blocktapping | 0.2452801 | 0.7547199 | 0.5 | 0.3249949 | 0.92106837 | -0.003241767428 | 0.02791435 |
| ThetaPow85 | Sum_PC | 0.2558280 | 0.7441720 | 0.5 | 0.3437754 | 0.66167522 | -0.011243525092 | 0.03137513 |
| ThetaPow86 | Blocktapping | 0.2559038 | 0.7440962 | 0.5 | 0.3439123 | 0.60900669 | 0.012682463910 | 0.02843743 |
| ThetaPow86 | Sum_PC | 0.2438492 | 0.7561508 | 0.5 | 0.3224875 | 0.90004364 | -0.002144419116 | 0.02851079 |
| ThetaPow87 | Blocktapping | 0.2462716 | 0.7537284 | 0.5 | 0.3267379 | 0.70053233 | 0.009740910242 | 0.02731361 |
| ThetaPow87 | Sum_PC | 0.2399751 | 0.7600249 | 0.5 | 0.3157464 | 0.91031290 | 0.004059696638 | 0.02876405 |
| ThetaPow88 | Blocktapping | 0.2422318 | 0.7577682 | 0.5 | 0.3196648 | 0.90422713 | -0.001820025114 | 0.02724487 |
| ThetaPow88 | Sum_PC | 0.2441955 | 0.7558045 | 0.5 | 0.3230936 | 0.76624389 | 0.008997300153 | 0.02903419 |
| ThetaPow89 | Blocktapping | 0.2667349 | 0.7332651 | 0.5 | 0.3637632 | 0.55297603 | 0.015634827619 | 0.02998075 |
| ThetaPow89 | Sum_PC | 0.2656601 | 0.7343399 | 0.5 | 0.3617672 | 0.59766930 | -0.012488981878 | 0.03049399 |
| ThetaPow9 | Blocktapping | 0.3476540 | 0.6523460 | 0.5 | 0.5329287 | 0.23011740 | 0.041154029633 | 0.03939861 |
| ThetaPow9 | Sum_PC | 0.2635383 | 0.7364617 | 0.5 | 0.3578438 | 0.81221633 | -0.003552848799 | 0.02743841 |
| ThetaPow90 | Blocktapping | 0.2329745 | 0.7670255 | 0.5 | 0.3037376 | 0.88627132 | -0.004153144887 | 0.02718189 |
| ThetaPow90 | Sum_PC | 0.2306557 | 0.7693443 | 0.5 | 0.2998082 | 0.99009786 | -0.001210289722 | 0.02719288 |
| ThetaPow91 | Blocktapping | 0.2275593 | 0.7724407 | 0.5 | 0.2945978 | 0.86827168 | -0.003775068028 | 0.02610109 |
| ThetaPow91 | Sum_PC | 0.2273804 | 0.7726196 | 0.5 | 0.2942980 | 0.89787908 | 0.003110804671 | 0.02701304 |
| ThetaPow92 | Blocktapping | 0.2384689 | 0.7615311 | 0.5 | 0.3131440 | 0.91033436 | -0.002846848271 | 0.02676245 |
| ThetaPow92 | Sum_PC | 0.2561999 | 0.7438001 | 0.5 | 0.3444472 | 0.56689856 | -0.015330081774 | 0.03004730 |
| ThetaPow93 | Blocktapping | 0.2267973 | 0.7732027 | 0.5 | 0.2933220 | 0.90454280 | -0.004642007359 | 0.02534073 |
| ThetaPow93 | Sum_PC | 0.2241925 | 0.7758075 | 0.5 | 0.2889795 | 0.98241095 | -0.000776917223 | 0.02593748 |
| ThetaPow94 | Blocktapping | 0.2372736 | 0.7627264 | 0.5 | 0.3110861 | 0.90051282 | 0.002784077571 | 0.02587141 |
| ThetaPow94 | Sum_PC | 0.2385415 | 0.7614585 | 0.5 | 0.3132692 | 0.86770687 | -0.004297008323 | 0.02736883 |
| ThetaPow95 | Blocktapping | 0.2402685 | 0.7597315 | 0.5 | 0.3162545 | 0.94271376 | 0.002233929046 | 0.02561150 |
| ThetaPow95 | Sum_PC | 0.2549084 | 0.7450916 | 0.5 | 0.3421168 | 0.56384299 | -0.014486121772 | 0.03020079 |
| ThetaPow96 | Blocktapping | 0.2440281 | 0.7559719 | 0.5 | 0.3228005 | 0.85878878 | 0.001889896065 | 0.02660218 |
| ThetaPow96 | Sum_PC | 0.2598494 | 0.7401506 | 0.5 | 0.3510765 | 0.56356512 | -0.016140986102 | 0.03096389 |
| ThetaPow97 | Blocktapping | 0.2442881 | 0.7557119 | 0.5 | 0.3232557 | 0.76830666 | -0.008997711079 | 0.02738602 |
| ThetaPow97 | Sum_PC | 0.2456844 | 0.7543156 | 0.5 | 0.3257051 | 0.78402435 | -0.008104030200 | 0.02887513 |
| ThetaPow98 | Blocktapping | 0.2353954 | 0.7646046 | 0.5 | 0.3078655 | 0.92078413 | 0.001091350682 | 0.02714172 |
| ThetaPow98 | Sum_PC | 0.2357357 | 0.7642643 | 0.5 | 0.3084479 | 0.99292837 | -0.001348911300 | 0.02764397 |
| ThetaPow99 | Blocktapping | 0.2600639 | 0.7399361 | 0.5 | 0.3514680 | 0.55500029 | 0.013956812442 | 0.02897476 |
| ThetaPow99 | Sum_PC | 0.2479701 | 0.7520299 | 0.5 | 0.3297344 | 0.70395735 | -0.010382857677 | 0.02832175 |
